# Leveraging the global genomic epidemiology of carbapenemase-producing *Klebsiella pneumoniae* to inform infection prevention in Tunisian hospitals

**DOI:** 10.1101/2025.09.22.25332449

**Authors:** Basma Menif, Jay N. Worley, Noura Ben Mansour, Faouzia Mahjoubi, Adnene Hammami, Lynn Bry

**Affiliations:** Laboratory of Microbiology, Habib Bourguiba University Hospital, Sfax University, Sfax, Tunisia; School of Medicine of Sfax, Sfax University, Sfax, Tunisia; Massachusetts Host-Microbiome Center, Department of Pathology, Brigham and Women’s Hospital, Harvard Medical School, Boston, Massachusetts, USA; Harvard Medical School, Boston, Massachusetts, USA; National Center for Biotechnology Information, National Library of Medicine, National Institutes of Health, Bethesda, Maryland, USA; Clinical Microbiology Laboratory, Department of Pathology, Brigham and Women’s Hospital, Harvard Medical School, Boston, Massachusetts, USA

## Abstract

Habib Bourguiba Hospital in Sfax, Tunisia, a 500-bed regional medical center, experienced profound increases in carbapenemase-producing *Klebsiella pneumoniae* (CPK) infections over 2009-2022. To implement improved control measures, we leveraged clinical microbiologic results on 1013 unique CPK, followed by targeted genomic analyses to conduct high-resolution clustering of strains and evaluate replicons mobilizing carbapenemases and other forms of resistance. Analyses resolved transmission chains, reservoirs, and informed approaches to facilitate improved infection control and prevention.

Hospital-acquired CPK increased >10-fold over 2009-2022, from 0·95 to 9·59 cases per 10000 patient days. OXA-48-like enzymes occurred most frequently (730 strains, 56·7%), followed by NDMs (281, 43·1%), or both (207, 26·9%). Genomic analyses revealed 23 genomic sequence types (STs), with dynamic shifts among clusters within ST101, ST147, and ST383 (70% of isolates). In 2009, ST383 carrying *bla*_OXA-204_ in IncC arose first, followed by ST101 carrying *bla*_OXA-48_ on IncL/M in 2011, and ST147 carrying *bla*_NDM-1_ on IncFIB (pQIL)/IncFII multi-replicon plasmids in 2014. Since 2019, emerging epidemic strains of ST383 carrying *bla*_NDM-5_ and *bla*_OXA-48_ on hybrid IncFIB/IncHI1B virulence plasmids became dominant, highlighting the capacity of these lineages to disseminate and sustain nosocomial outbreaks.

Clustering with 80,252 *K. pneumoniae* genomes within the internationally available FDA GenomeTrakr and NCBI Pathogen Detection tools placed Tunisian strains within the broader global context, of which 82% STs clustered with Mediterranean and global clusters, while 18% remained unique to the hospital. Findings identified local through international reservoirs of strain entry into the hospital and supported clinical microbiologic approaches for patient surveillance and rapid assessments to identify putative outbreaks.

## Introduction

Carbapenem resistance is a major global public health problem.^1,2^ The World Health Organization (WHO) prioritized carbapenemase-producing *Klebsiella pneumoniae* (CPK) as a top multidrug-resistant pathogen for 2024.^3^ Prevalent high-risk clones (HiRC) include clonal groups (CGs) 258 (ST258, ST1512, ST11), CG15 (ST15, ST14), ST101, and ST307,^4^ which cause 72% of reported hospital outbreaks.^5^ These clusters share antimicrobial resistance profiles but differ from hypervirulent *K. pneumoniae* strains (HvKp) associated with community-acquired invasive infections, though some Asian ST11 and ST23 HvKP strains have demonstrated carbapenem resistance.^6,7^ These dynamics led to WHO’s 2024 guidance to implement measures for CPK surveillance to inform local prevention measures.^8^

An ongoing *K. pneumoniae* surveillance program at Habib Bourguiba Hospital (HBH) in Sfax, Tunisia optimizes use of Clinical Microbiologic testing with pulse field gel electrophoresis (PFGE) to select strains for whole-genome analyses, followed by genomic-epidemiologic investigation to identify putative transmission chains, reservoirs, and inform the implementation of improved prevention programs. The program’s prior successes include identifying and supporting containment of an outbreak of VIM-4-producing *K. pneumoniae* in 2005,^9^ and confirming entry of the first OXA-48-like producing *K. pneumoniae* in 2009.^10^ Since then, OXA-48- and NDM-producing CPK have become endemic in Tunisian hospitals.^11–14^

Despite these achievements, the genetic epidemiology of nosocomial CPKs in Tunisia remains limited and hampers efforts to identify reservoirs to inform prevention efforts.^11,13,14^ The Laboratory of Antibiotic-Resistance in Tunisia (LART) ,^15^ showed that Tunisian rates for CPK infections have risen dramatically from 0.6% of infections in 2010 to 22·4% in 2019, and 18·5% in 2022, which is comparable to resistance rates seen across Africa and the Middle East (5·7-26·9%).^16^ This increase requires active surveillance to inform effective strategies in CPK prevention.

Over 14 years, from 2009-2022, we evaluated CPK dynamics in HBH. Our analyses identified time-dependent emergence of genomic clones and antibiotic resistance (AMR)-transmitting replicons, with recent and repeated emergence of global ST383 strains. Our findings demonstrate internal and external reservoirs of strains impacting patient infections within HBH. The findings informed our infection prevention and control efforts and brought clarity to the active KPC crisis affecting a region with limited genomic resources. In particular, our genomic analyses identified an emerging global epidemic strain of ST383 driving international dissemination of carbapenemase and virulence-carrying plasmids, emphasizing the need to expand genomic surveillance efforts in low- and middle-income countries.

## Results

### Demographics and dynamics of patient-infecting CPKs

We evaluated 1013 unique patient CPK strains identified at HBH from 2009 to 2022. Among these strains, 115 (11·3%) originated from outpatients, 114 (11·4%) from inpatients within 48 hours of admission, and 748 (77·4%) in inpatients after 48 hours of admission, giving an overall incidence of 4.365 hospital-acquired CPK per 10,000 hospital days (table 1). Notably, CPK rates increased >10-fold from 0·950 to 9·590 cases per 10000 patient-days from 2009-2022 (p<0·001, Fig. 1).

**Fig. 1:**
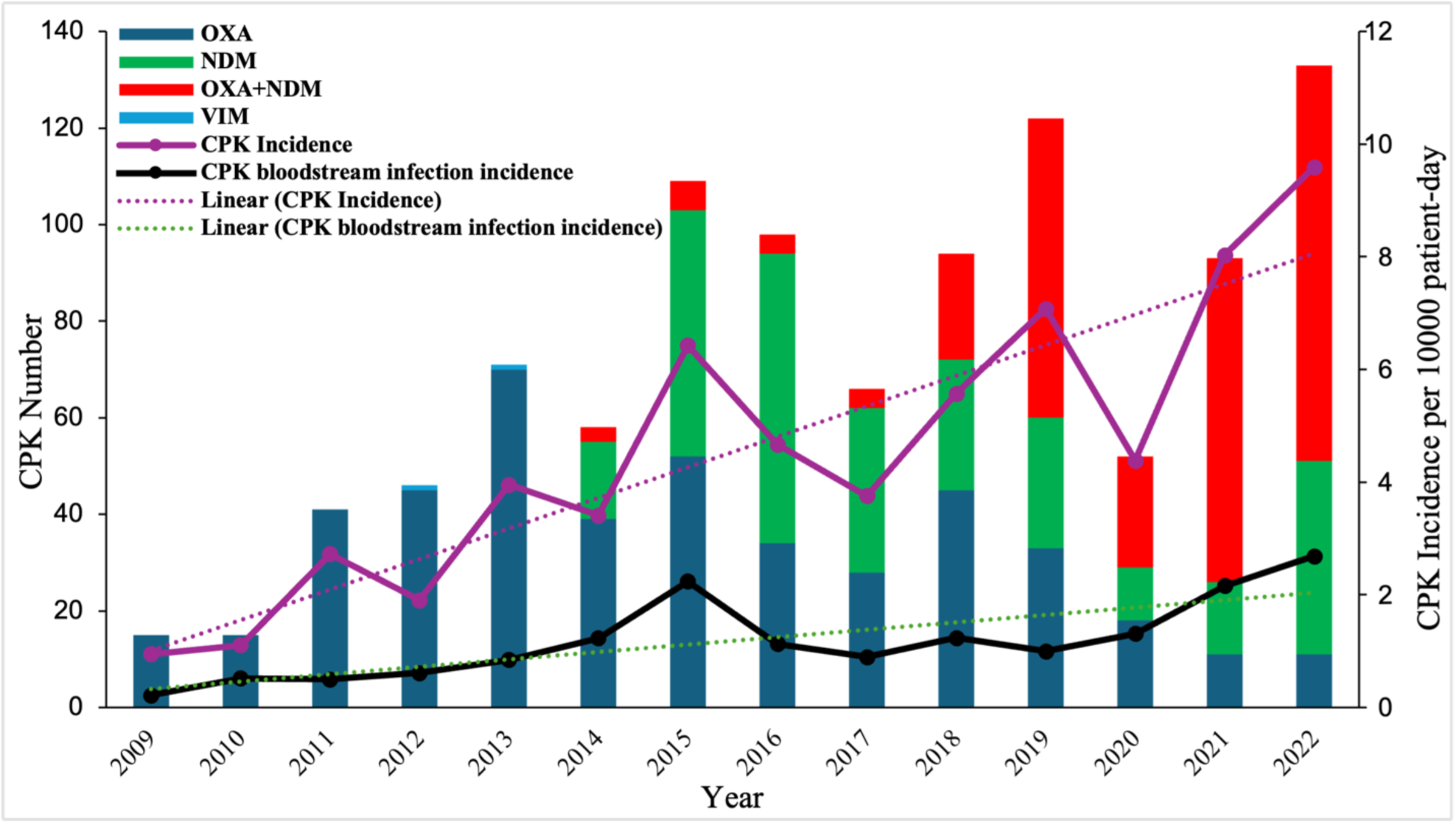
Annual incidence and carbapenemase profiles of CPK isolates in HBH, 2009–2022. X axis shows years; Y axis on the right shows the number of CPK isolated by year, and on the left shows the incidence of CPK per 10,000 patient-day. Sample key indicates the carbapenemase types and the linear regression lines representing the trend in CPK incidence

**Table 1:** Demographics of CPK-infected patients.

| **Demographic Information** | **Data** |
| --- | --- |
| **Gender (% of cases)** |  |
| Male | 658 (64.9) |
| Female | 355 (35.1) |
| **Age: Years (Standard Deviation)** | 54.21 (18.3) |
| **Hospital ward: (% of cases)** |  |
| Intensive care | 504 (49.8) |
| Medical | 38 (3.8) |
| Surgical | 423 (41.8) |
| Emergency room | 48 (4.7) |
| **Isolate site (% of cases)** |  |
| Urine | 380 (37.5) |
| Blood | 220 (21.7) |
| Sputum | 152 (15.0) |
| Skin/soft tissue | 198 (19.5) |
| Other* | 63 (6.2) |
| **Origin (% of cases)** |  |
| Hospital-acquired ($\geq 48$h after admission) | 784 (77.4) |
| Community-acquired ($< 48$h after admission) | 114 (11.3) |
| Outpatient | 115 (11.4) |

Demographic data of CPK isolates from patients seen at Habib Bourguiba Hospital in Sfax, Tunisia, over 2009-2022. Numbers represent patients; percentages are in parentheses.
\*: catheter, drain, biopsy, material, and genital specimens

CPK infections occurred most frequently in medical ICU patients (45·8%), with an average rate of 37·5 infections per 10,000 patient-days (range of 4·7 to 64·5 infections) 2009-2022 (p<0·001). Urinary tract infections (n=380, 37·5%) were most common, followed by sepsis (220, 21·7%), wound infections (198, 19·5%), and pneumonias (152, 15·0%) (table 1). Clinical PCR testing for carbapenemase carriage identified OXA-48-like enzymes (730, 56·7%) and NDMs (281, 43·1%) as the most prevalent, with 26·9% of CPKs encoding both (n=273). Only two isolates expressed VIM enzymes, and no isolates carried KPCs.^17^ OXA-48 producers predominated until 2014, while NDM-carrying strains predominated from 2017 (Fig. 1).

Antimicrobial susceptibility testing found 859 isolates (84·8%) to be resistant to imipenem (MIC >2 mg/L) and 846 (83·5%) to meropenem (MIC > 2 mg/L). OXA-48-like producers remained susceptible to ceftazidime/avibactam, in contrast to NDM producers which were resistant. While all CPK isolates remained susceptible to aztreonam-avibactam, they co-harbored high levels of resistance to ciprofloxacin (98·1%), gentamicin (89·5%), and amikacin (54·6%). For aminoglycoside resistance, PCR-based testing detected the methylase-encoding genes *armA*, *rmtF*, *rmtC*, *rmtB*, and *rmtE* in 219 (21·6%), 46 (4·5%), 23 (2·3%), 5 (0·5%), and 3 (0·3%) isolates, respectively. In addition, 57·3% of isolates were resistant to tigecycline (MIC > 0·5 mg/L), and 212 (20·9%) to colistin. The colistin-resistant strains were negative for *mcr* genes.

Given concerns for the spread of hypervirulent CPK, string-test and PCR evaluation of putative *K. pneumoniae* virulence loci *rmpA/rmpA2* and aerobactin (*iuc)* were performed. These studies identified 265 (26·2%) convergent CPK strains that harbored r*mpA/rmpA2* and/or aerobactin. However, only four isolates were string-test positive. The string-test positive isolates carried the hv-CRKP hypervirulence locus: ST23-KL1 in 2013 (n=1), ST86-KL2 in 2010 (n=1), and ST218-KL57 in 2010 (n=2). The remaining 261 convergent strains lacked typical hypervirulent capsular types and were string-test negative. These convergent CPK strains emerged in 2018 and rapidly expanded, from an initial prevalence of 11% in 2018 to 72% in 2021 and 63·9% in 2022.

### CPK isolate typing

Given limited resources for strain genomic analyses, we subjected 220 CPK sepsis isolates to PFGE and selected 95 representative strains per their PFGE and AMR profiles, per year, for genomic analyses. The sepsis isolates exhibited the same resistance and virulence profiles as those from other sites, indicating they are likely representative of the whole set (Supplementary Table S1). While PFGE identified 2 to 12 pulsotypes per year, a single pulsotype accounted for >30% of sepsis isolates each year (Supplementary Fig.S1, Table S2). This similarity reduced the number of isolates needed to build a representative set of the most prevalent strains.

Genomic analyses revealed 23 multi-locus sequence types (STs), of which 9 STs included >2 strains (39·1%) and 14 STs (60·9%) 1-2 strains. Three STs accounted for 70% of sequenced CPK sepsis isolates: ST101 (n=73, [33·2%]), ST383 (n=41 [18·6%]), and ST147 (n=41 [18·6%]). Additional clusters ST11 (n=9), ST15 (n=9), ST13 (n=5), ST307 (n=5), and ST2096 (n=5) occurred with less frequency and for shorter durations than the major three.

Analyses identified shifting dynamics and evolving CPK strains over time, with new STs and carbapenemase genes replacing prior strains within HBH (Fig. 2, Supplementary Fig. S2). ST101-OXA-48 predominated over 2011-2018. In contrast, ST383, which caused the first OXA-204 CPK epidemic in 2009-2010, was reintroduced in 2019 and became the predominant MLST while now encoding the OXA-48, NDM-5, and ArmA enzymes. ST147 has driven the dissemination of NDM-1 from 2014 and was prevalent through the end of the study period.

**Fig. 2:**
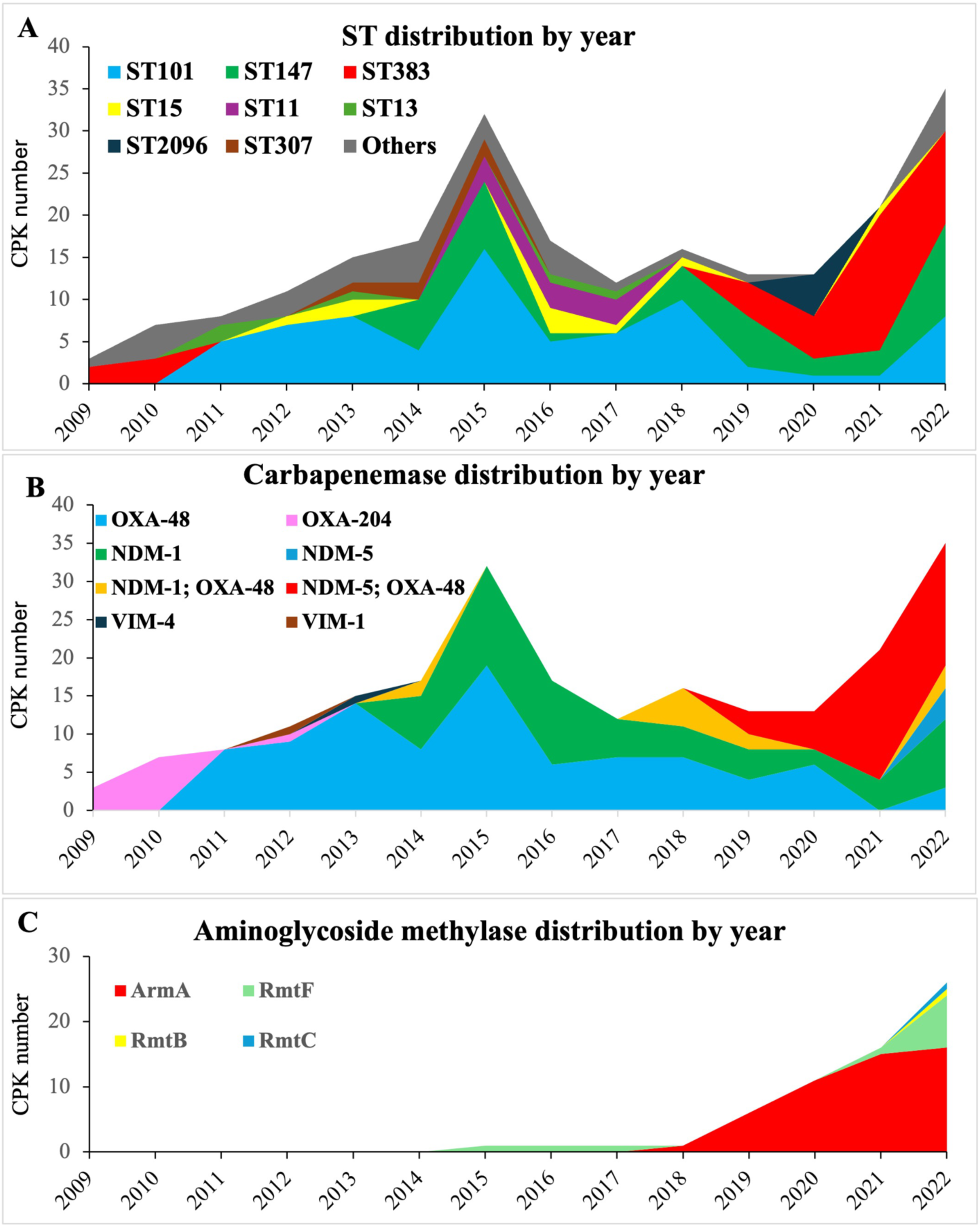
Temporal distribution of HBH’s sepsis CPK isolates from 2009–2022. X axis shows years; Y axis shows the number of CPK isolated by year. Sample key shows the carbapenemase types. (A) Temporal distribution of clonal types; (B) Temporal distribution of carbapenemase types (C) Temporal distribution of methylase types associated with aminoglycoside resistance

OXA-48 (n=144 [52·7%]) occurred most frequently among sepsis CPK isolates, followed by NDM-1 (n = 71 [26%]), NDM-5 (n = 45 [16·5%]), OXA-204 (n = 11 [4%]), VIM-1 (n = 1), and VIM4 (n = 1). In addition, 53 (24%) isolates harbored >1 carbapenemase, with 41 carrying *bla*_OXA-48_ and *bla*_NDM-5_, and 12 carrying *bla*_OXA-48_ and *bla*_NDM-1_.

Among the sepsis CPKs, we identified 61 (27·7%) convergent isolates among ST383 (n=33), ST147 (n=14), ST2096 (n=5) and ST101 (n=3). From 2019, the prevalence of *rmpA*-positive convergent strains rose in ST383 (33/35; 94·3%) compared to other STs (24/47; 51·1%). Nevertheless, CPK sepsis mortality over 2015-2022 did not differ relative to the presence or absence of these virulence markers (34/51, 67%) (51/93, 55%) (p = 0·2289).

### Genomic clustering identifies local to global strain relationships

NCBI’s Pathogen Detection Resource assigned 68% (65 of 95 strains) of HBH’s CPKs to 26 international genomic SNP clusters, 3 of which were previously reported in Tunisia: ST101 and ST11 seen clinically, and ST147 identified in seafood. Five new clusters contained only strains from this study and 30 CPK strains from HBH (31·6%) did not cluster with other NCBI-deposited *K. pneumoniae*.

Within HBH, 15 clusters involved 2-8 patients, suggesting potential internal transmission chains for at least 58% (n= 55) of sequenced isolates. Among the 55 strains in ST101, ST147, and ST383, 45 (81%) clustered with strains from 22 countries, particularly Mediterranean and other European countries, as well as the US, China, India, the Middle East, and Australia (Fig. 3).

**Fig. 3:**
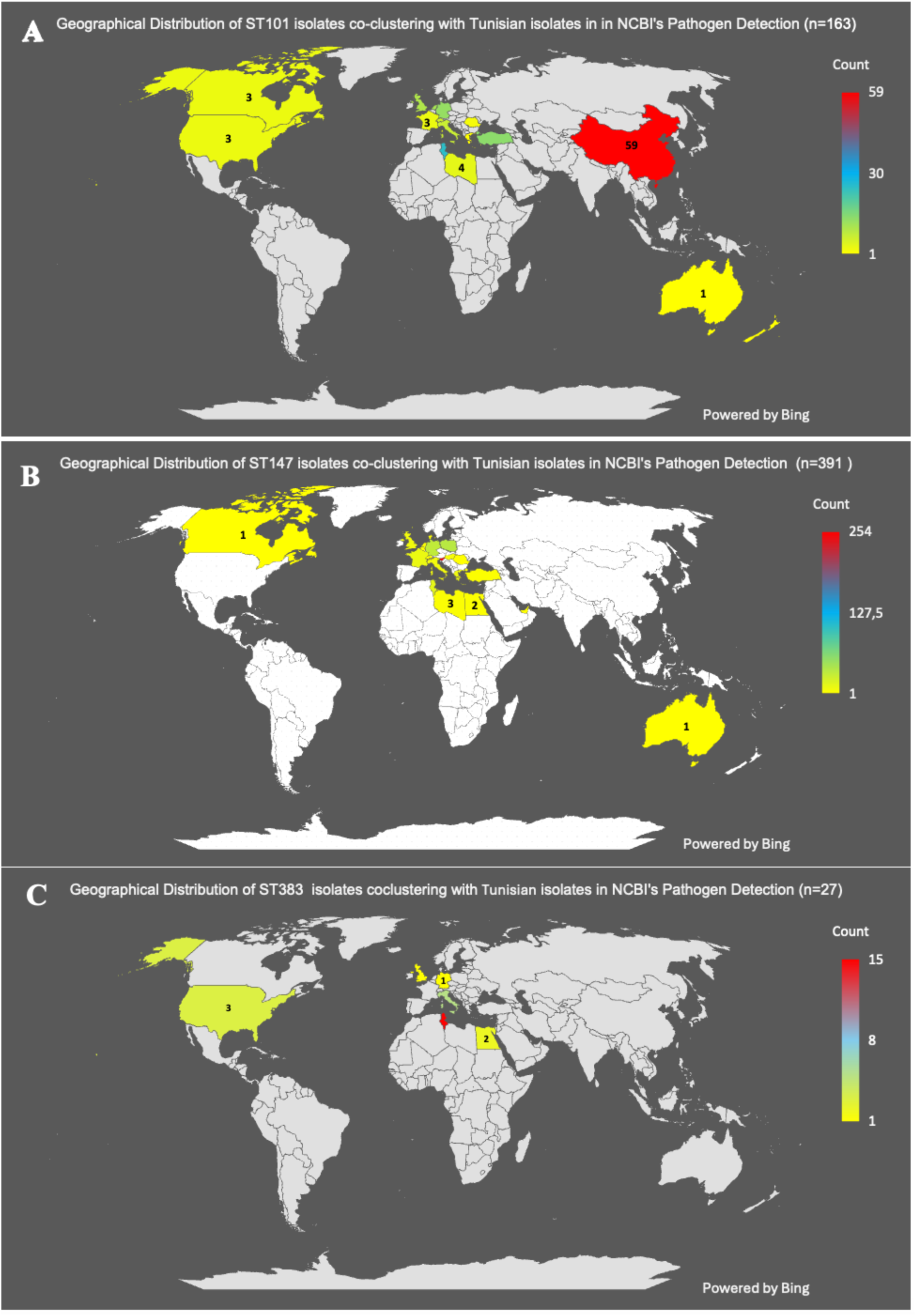
Geographic distribution of dominant STs seen in HBH (A) ST101, (B) ST383, and (C) ST147. Color key shows the number of strains per country, from NCBI Pathogen Detection, that co-cluster with corresponding Tunisian clones. Genomic data accessed on June 18, 2025.

To validate the relationships shown by NCBI’s clustering, wgMLST on 80,252 of the available 86,290 assemblies were used to identify the 100 most closely related genome assemblies. Phylogenetic estimation of the reference gene SNPs validated NCBI’s SNP-based clustering and phylogenetic relationships revealed among SNP clusters (Supplementary data 1; interactive results at https://itol.embl.de/shared/pUTb8g8Zw3eX ).

### Comparative genomics of HBH’s dominant ST383, ST147 and ST101 clusters

GalaxyTrakr analyses identified considerable diversity among ST101 (range 2–293 SNPs, mean 150 [SD 72]), ST147 (range 9–274, mean 166 [SD 76]), and ST383 strains (range 2–251, mean 119 [SD 78]).

HBH’s first ST101 isolate in 2011 belonged to NCBI SNP cluster PDS000041735.6, which includes contemporaneous isolates from Germany, the UK, China, and the US, suggesting pre-existing global prevalence of this lineage. Within HBH, we detected four putative transmission events among ST101 strains (dotted outlined boxes, Fig. 4A). Transmitted strains shared the same AMR profile and carbapenemase-transmitting-plasmids (Fig. 4B), further indicating persistence of this clone within HBH over 2011-2022. However, strains within SNP clusters PDS000199425.2 and PDS000041735.6 (Fig. 4C) were more closely related to strains from other countries, and not other HBH ST101 isolates, suggesting reintroduction of ST101 sub-lineages into HBH many times during this period.

**Fig. 4:**
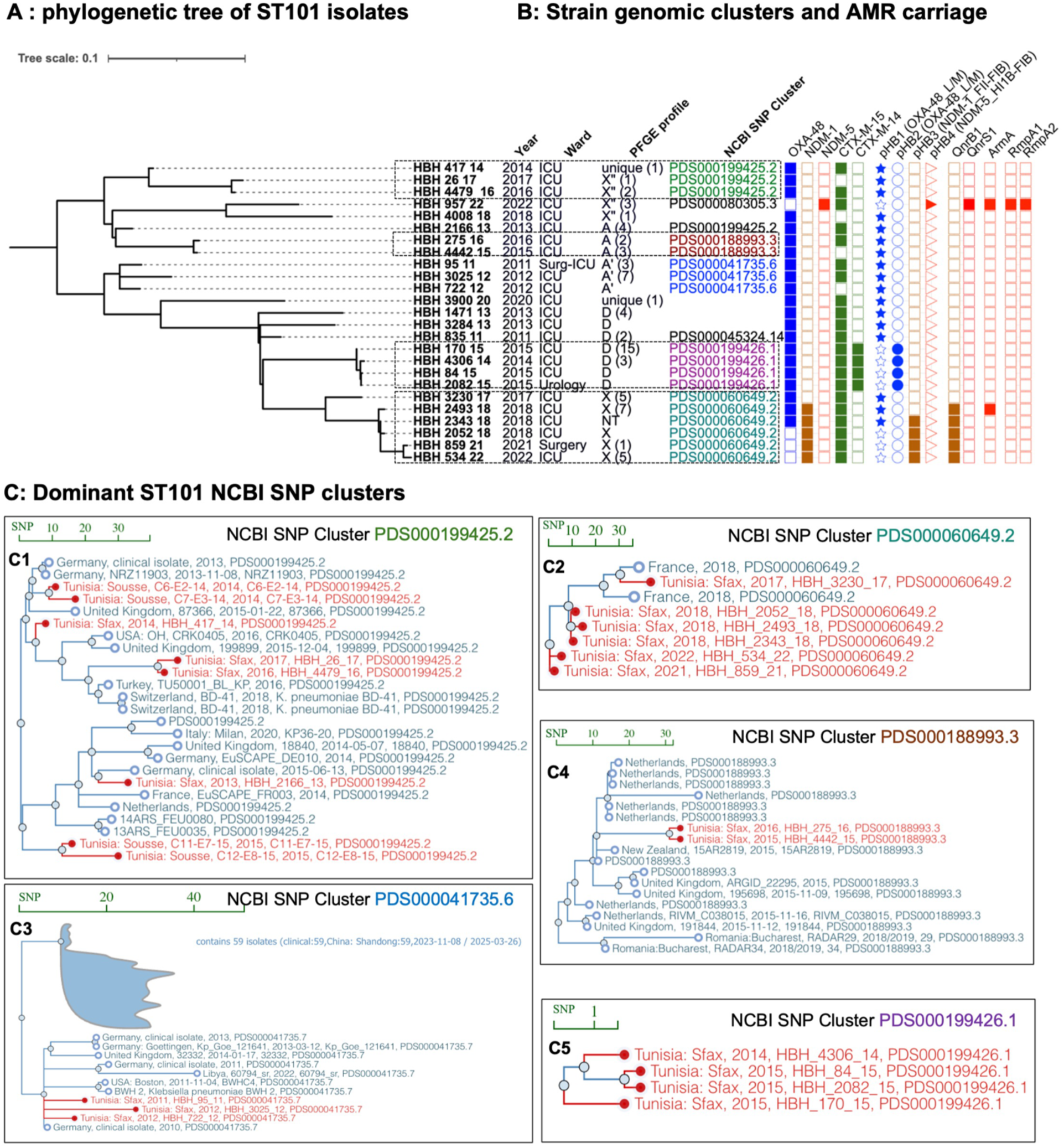
International context of ST101 genomic clusters seen in HBH. **A:** phylogenetic tree of ST101 strains seen at HBH over 2009-2022 **B**: Isolate metadata: isolation year and ward, NCBI SNP cluster ID, PFGE profile, resistance genes, carbapenemase-producing plasmids. **C:** NCBI SNP cluster analyses reveal that closely related strains from different countries occur within the same NCBI SNP cluster, suggesting sporadic introduction into HBH (C1, C2), whereas other strains are specific to the setting (C3-C5). Bar in each graph indicates the SNPs distance, the branches highlighted in bold indicate the study strains or the Tunisian strains from other settings. The NCBI SNP clusters are indicated in the same colors as in the phylogeny tree.

Similar patterns occurred among the ST147 strains, where *bla*_NDM-1_-containing isolates in NCBI cluster PDS000045135.25 appeared in HBH in 2014. From 2014-2022, this cluster, with distances of 9-25 SNPs among strains, encompassed 110 isolates from 15 countries. Genomic analyses of the 22 ST147 strains showed carriage of *bla*_NDM-1_ on [pHB3] (pQIL, IncFII-FIB), suggesting persistence and transmission of this strain-plasmid complex within HBH (transparent boxes, Fig. 5A, Fig. 5B). While comparable PFGE and AMR profiles suggested internal transmission, NCBI Pathogen Detection’s analysis indicated that these highly related isolates were part of regional or global cryptic outbreaks in the late 2010’s, suggesting likely repeated and sporadic introductions into HBH (Fig. 3B-5C, Supplementary data2 Tree10-29). After 2019, we identified phylogenetically distinct ST147 harboring *bla*_NDM-1_ in the FIB [pHB5] plasmid as well as carriage of additional virulence plasmids.

**Fig. 5:**
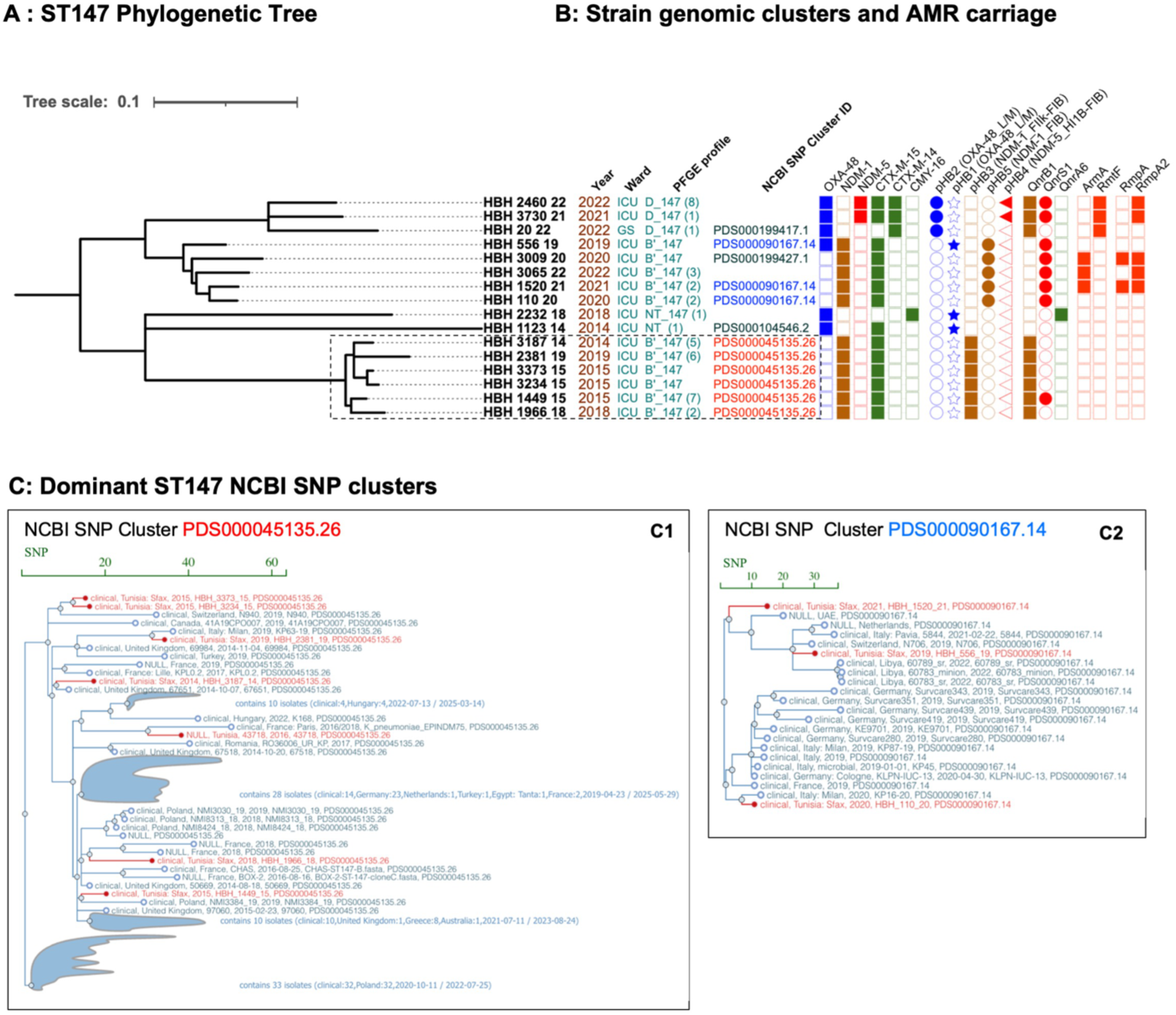
International context of ST147 genomic clusters seen in HBH. **A:** phylogenetic tree of ST147 strains seen at HBH over 2009-2022 **B:** Isolate features including ward, PFGE profile, NCBI genomic cluster ID (if present), resistance genes, and mobilizing plasmids. **C:** NCBI SNP cluster analyses reveal that many other closely related strains from different countries fell within the same SNP cluster, supporting their repetitive introduction to the setting (C1, C2). Bar in each graph indicates the SNPs distance, the branches highlighted in bold indicate the study strains. The NCBI SNP clusters are indicated in the same colors as in the phylogeny tree.

Analyses also identified new and recent clusters of ST383 in HBH that co-clustered with isolates seen globally. Within these clusters, genomic analyses identified carriage of *bla*_NDM-5_ in the hybrid plasmid [pHB4] IncFIB(pNDM-Mar)-IncHI1B(pNDM-MAR). These ST383 isolates from 2009-2010 were separated by 8-13 SNPs from strains occurring in HBH’s first *bla*_OXA-204_ outbreak in 2009-2010, suggesting a common ancestor strain carrying different replicons and resistance genes.^10^ In contrast, the isolates from 2019-2022 formed a separate cluster showing 2-125 SNPs difference among strains (NCBI clusters PDS0000885592.11 and PDS000098809.4). These later isolates shared the same PFGE and AMR profiles, and *bla*_NDM-5_ hybrid plasmid: IncFIB(pNDM-Mar)-IncHI1B(pNDM-MAR). Isolates from these SNP clusters originated from Italy, Egypt, Germany, the UK, the Netherlands, and the US, suggesting global dissemination of this strain (Fig. 6C). Within HBH, genomic-epidemiologic analyses identified HBH_3269-21 and HBH_1879-22 as being part of patient transmission chains (Fig. 6A, Supplementary data 1, Tree 27, 42, 43). The combined hospital-focused and placement of HBH strains within an international context resolved nosocomial transmission (Fig. 6A, dotted line boxes) from putative sporadic reintroduction from outside reservoirs (Fig. 6C).

**Fig. 6:**
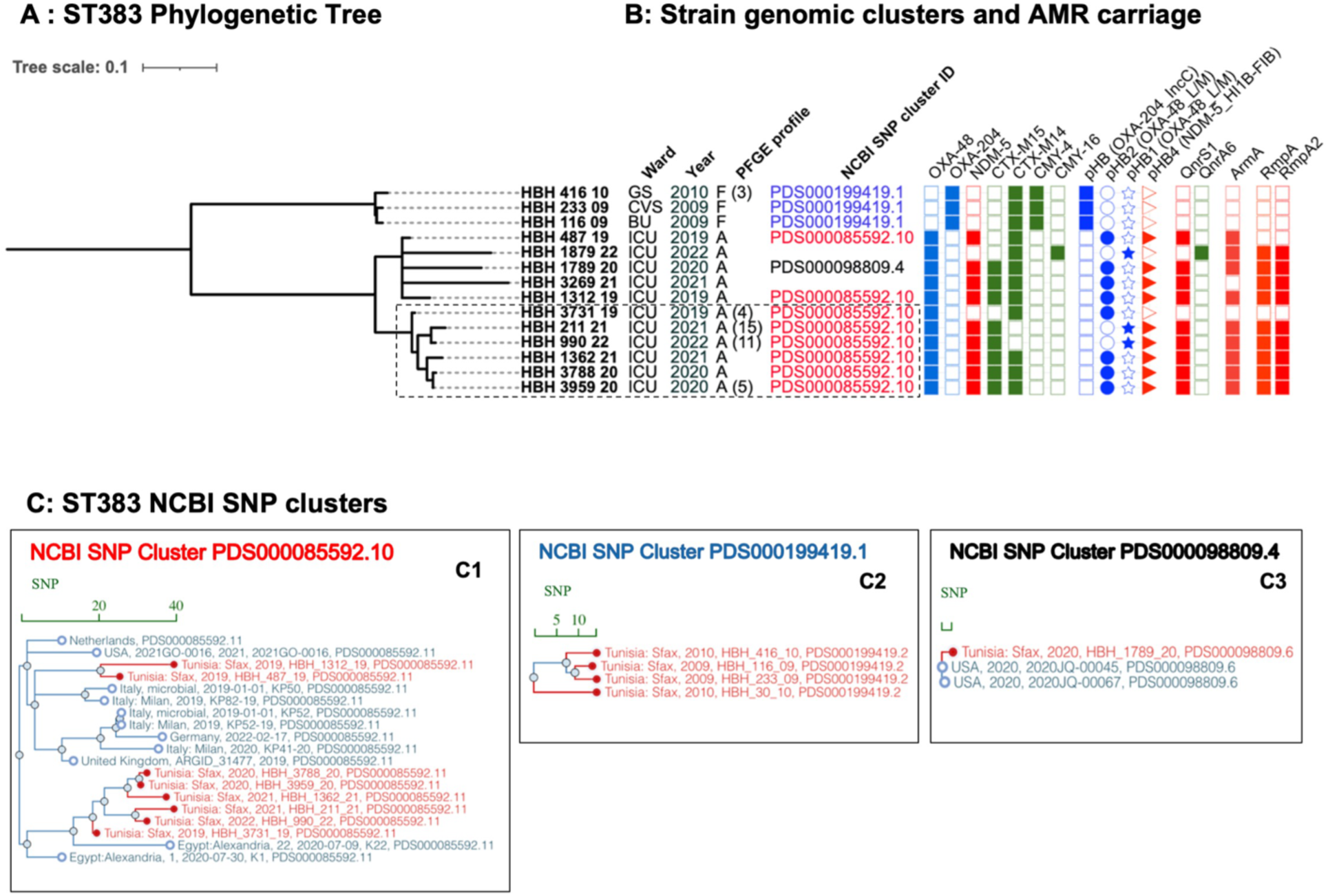
International context of ST383 genomic clusters seen in HBH. **A:** phylogenetic tree of ST383 strains seen at HBH over 2009-2022 **B:** Isolate features including ward, PFGE profile, NCBI genomic cluster ID (if present), resistance genes, and mobilizing plasmids. **C:** SNP cluster analyses reveal that many other closely related strains from different countries fell within the same SNP cluster, supporting their repetitive introduction to the setting, C1. Some clusters are specific to the setting, C2. Bar in each graph indicates the SNPs distance, the branches highlighted in bold indicate the study strains. The NCBI SNP clusters are indicated in the same colors as in the phylogeny tree.

### *K. pneumoniae* ST383’s phylogeny reveals recent origins contributing to four global outbreak events

*K. pneumoniae* ST383’s dominance at HBH (Fig. 2A) occurred in the context of limited global reports of these strains (Fig. 3C). Nonetheless, we leveraged the combined HBH and international ST383 in NCBI Pathogen Detection to evaluate relationships among our recent patient isolates. The refined phylogeny of ST383 strains revealed a single monophyletic population of 258 members, composed of carbapenemase producers identified after 2010 (monophyletic clonal group 383 (CG383)). Analyses identified four well-defined clades (Fig. 7, labeled A to D), which include a major descendant subclade D1, of clade D (Fig. 7, Supplementary data 1, Tree 43). CG383 extends beyond ST383, including three ST6188 in subclade D1, two ST4853 isolates in clade D, and 1 ST5410 isolate in clade C (Supplementary data 1, Tree 43). While the earliest OXA-204 Tunisian ST383 (2009-2010) belonged to clade B (61 isolates), this clade mostly contained 2011-2018 VIM-producing outbreak strains reported in Greece, UK, Netherlands, and Romania. In contrast, strains in more recent clades C (71 strains) and D (115 strains) acquired OXA-48 and NDM-5-hybrid plasmids (Fig. 7, Supplementary data 1, Tree 43). These latter Tunisian strains clustered within clade D1, with 67 strains first isolated in 2015-2016 in Egypt and Qatar.

**Fig. 7:**
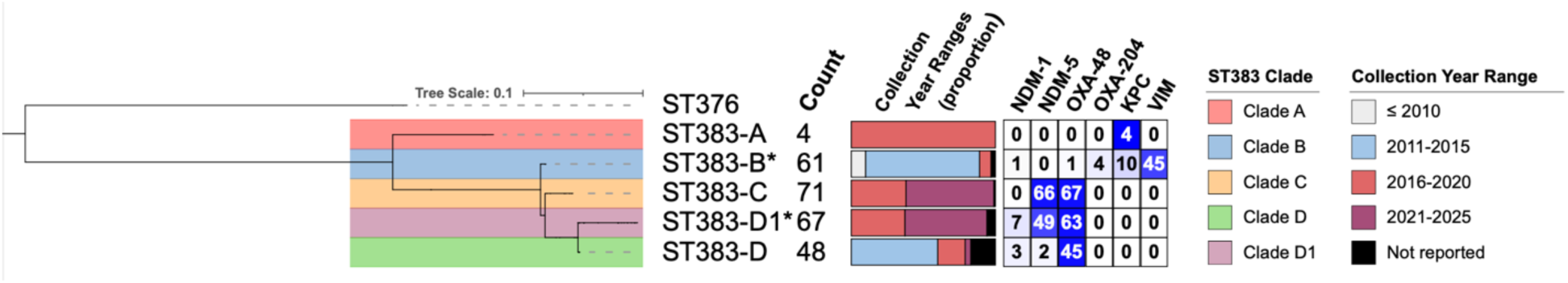
Global phylogenetic analysis of ST383/CG383 *K. pneumoniae*. Details about the clades, collection year, and the content of carbapenemases are shown. *: clades encompassing the HBH strains

Our longitudinal analyses of HBH’s ST383 CPK provide key bookends for the emergence of this concerning lineage, indicating origins around 2007 followed by repeated global dissemination. Recent isolates of CG383, predominantly ST383 in clades C and D1, demonstrate a high prevalence of OXA48 and NDM-5 producers with global reach (Supplementary Table S3).

### AMR transmission by sepsis CPK isolates

Genomic analyses identified 22 plasmid replicons among the 95 sepsis CPKs. Isolates carried a mean of 4·6 replicons per strain (median=4; range of 1-8 replicons). Among the dominant STs (Supplementary Fig. S3), ST147, associated with FIB/pKPHS1 replicons, commonly carried NDM-1 on FIB/pQil-FII.K replicons, while ST101 isolates, associated with repB1701, FIA(HI1), Col440I replicons, carried OXA-48 on IncL/M replicons. In contrast, ST383 isolates most commonly harbored NDM-5 on IncFIB(pNDM-Mar)-IncHI1B(pNDM-MAR) multireplicons.

Conjugation studies from sepsis isolates into the naive recipient strain J53 demonstrated host range of the carbapenemase-transmitting plasmids, including 38 *bla_OXA-48_* -IncL/M conjugative plasmids (38/53 OXA-48 CPKs, 71·7%), 18 *bla*_NDM-1_-IncFII_k_ conjugative plasmids, and one *bla*_NDM-1_-IncA/C conjugative plasmid (19/35 NDM-1 CPKs, 54·2%). Only one *bla*_NDM-5_-IncFIB-IncHI1B plasmid was transferred from an ST383 isolate (Supplementary Table S4-S5).

Bioinformatic reconstructions of the carbapenemase-carrying plasmids (Supplementary Table S4-S5) identified two major IncL/M plasmids among sepsis isolates, referred to as pHB1 (mob cluster AA018/AH562, OXA-48, CP018717.1), and pHB2 (AA002/AH529, OXA-48 + CTX-M-14, CP019078.1), with >96% coverage and >99% identity (Supplementary Fig. S4-5, Table S4, Supplementary data 2 Tables S6, S7) and worldwide (Supplementary data 1).

We identified four replicons associated with *bla*_NDM-1_: IncFII_K2_+FIB_(pQil)_ (23), IncFIB_(pQil)_ (5), IncFIB_K_+FII_K1_(2), and IncA/C (2) (Supplementary Fig. S6-9, Table S5). The *bla*_NDM-1_ pHB3 (FII_K2_+FIB_(pQil)_ , AA018/AH562) plasmid had a backbone showing >90% coverage and >99% identity with p1203/15 (FII_K2_+FIB_(pQil)_, MW363916), a plasmid reported in a Polish ST147 outbreak that originated from Tunisia ^18^ (Supplementary Fig. S6). This pHB3 plasmid occurred in six ST147 isolates from HBH as well as in six other STs seen in HBH since 2014, highlighting its high transmissibility among STs and patient populations. Moreover, pHB3 was the predominant plasmid encoding *bla*_NDM-1_ among the closest NCBI strains (Supplementary data 1). In contrast, the non-mobilizable *bla*_NDM-1_pHB5 (IncFIB_(pQil)_, AA019/AH565), homologous to CP014757, occurred in only five related ST147, suggesting clonal transmission of this strain-plasmid complex (Supplementary Fig. S7).

The *bla*_NDM-5_ plasmid pHB4 (IncFIB_(pNDM-Mar)_-IncHI1B_(pNDM-MAR)_, AA405/AI436) showed close homology to plasmids CP091814 (Qatar, ST383), CP034201 (UK, ST383), and CP137386 (UK, ST147) (>87% coverage and >98% identity). These conjugative plasmids harbor multiple resistance (*bla*_NDM-5_, *bla*_CTX-M-15_, *armA*, *qnrS*, *aac(6’)*) and virulence genes (*iucABCD-iutA, rmpA*, *rmpA2*). In 2022, pHB4 was identified from CPK sepsis isolates in ST147, ST29, and ST101 (Supplementary Fig. S10). Moreover, highly homologous plasmids to pHB4 occurred throughout the NCBI assemblies related to Tunisian isolates (Supplementary data 1).

In aggregate, *bla*_OXA-48_ IncL/M plasmid pHB1, *bla*_NDM-1_ IncFII_K2_+FIB_(pQil)_ pHB3, and *bla*_NDM-5_ IncHI1B+FIB_(pNDM-Mar)_ pHB4 were the most prevalent and promiscuous plasmids within the HBH CPK population (Fig. 8) and the HBH-related NCBI assemblies (Supplementary data 1).

**Fig. 8:**
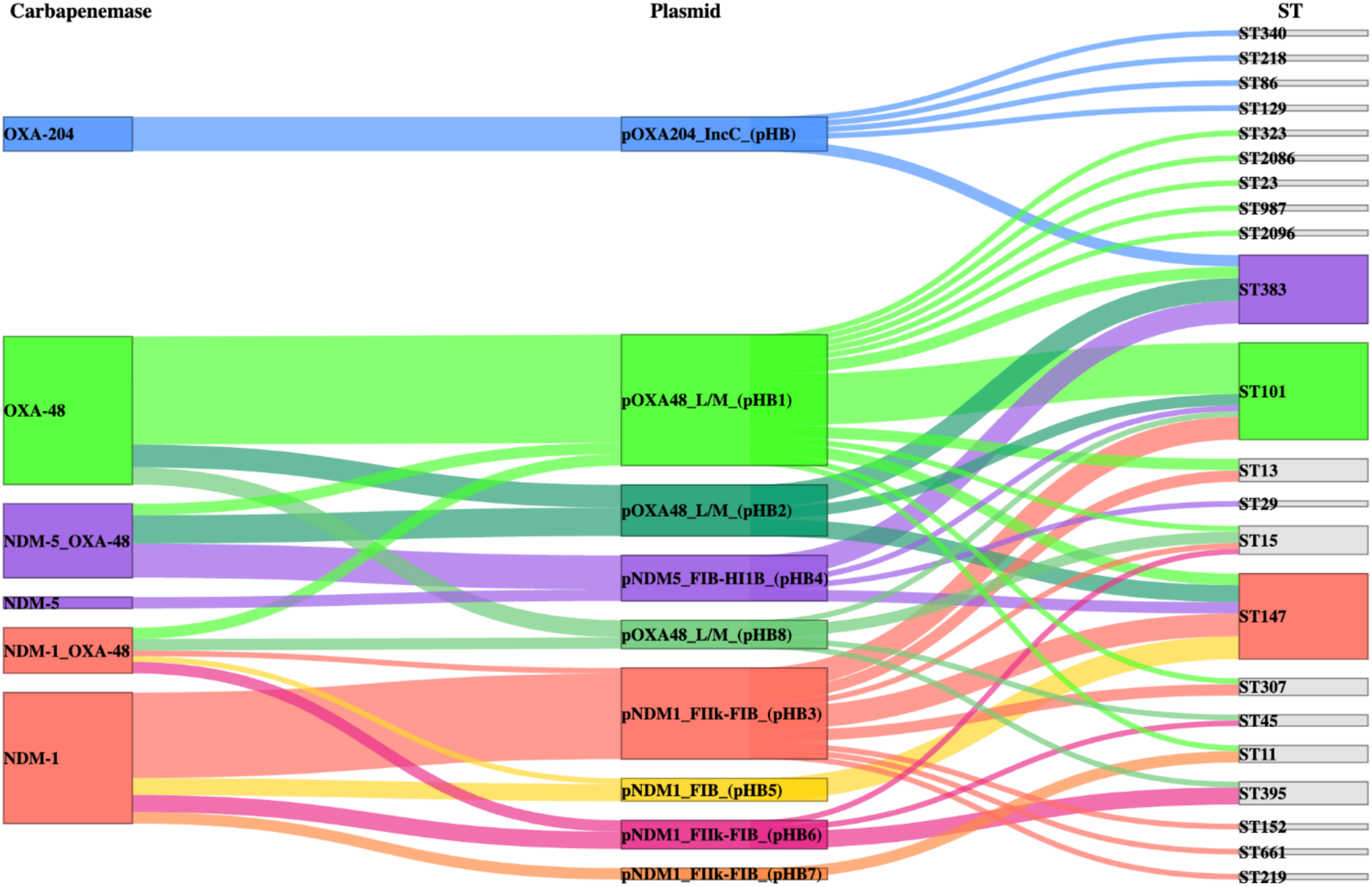
Sankey diagram illustrating relationships among the carbapenemase genes on the left, plasmid incompatibility and group types in the middle, and sequence types (ST) on the right. Line widths are proportional to the number of isolates and are colored based on carbapenemase-carrying plasmids noted in the middle section.

## Discussion

Genomic-epidemiologic analyses of patient-infecting CPK enable high-resolution understanding of their entry, dynamics, and mechanisms of spread within nosocomial environments. This information is critical to implement programs in infection prevention and control. We provide a high-resolution understanding of the dynamics of CPK over a 14-year period within a large Tunisian medical center. Given limited resources for genomic analyses, we leveraged clinical microbiologic phenotypic information with PFGE to select representative strains for genomic-epidemiologic investigations. Our data revealed sustained increases in CPK incidence, consistent with national reports from Tunisia, other resource-limited countries, and with global alerts from WHO.^1,2,15^ This rise underlines the need to develop tailored IPC programs for these settings.

While carriage of OXA-48-like carbapenemases occurred most frequently, we identified unexpected increases in NDM carriage, particularly of NDM-5.^19^ Surprisingly, we did not detect KPC enzymes by genomics or clinical PCR testing, nor the globally dominant ST258 clone, suggesting that our setting diverges from global epidemiological trends where ST258-KPC continues to predominate.^4^

Analyses of sepsis CPK isolates revealed dynamic shifts in CPK populations, particularly among three lineages, ST101, ST147, and ST383 that carried OXA-48, NDM-1, and NDM-5+OXA-48, respectively. Comparative genomic analyses of these lineages identified diverse isolates. Although PFGE analysis suggested substantial intra-hospital transmission of these clones, particularly among ST101, analyses within NCBI Pathogen Detection revealed that 21 of 48 strains in putative PFGE-identified clones (44%) were not clonal but instead related to strains from other countries, raising suspicion for sporadic reintroduction from external sources. These findings inform the need for IPC programs to consider entry and transmission by colonized patients who may have acquired strains in neighboring healthcare institutions or travel to other countries. While PFGE demonstrated less discriminatory power than genomic approaches, in a resource-constrained setting, it still provides useful information in selecting potentially related strains for genomic analyses. We also verified PFGE’s capacity to predict clonal associations among outbreak clusters of ST147 and ST383, particularly in settings where resources to conduct genomic analyses are limited, allowing use of this information to inform infection control and prevention strategies. However, continued analysis and vigilance is required to ensure the predictive capacity of PFGE to identify putative transmission chains.

MDR plasmids transmitted the CPK carbapenemases. pBH3 IncFII_K2_+FIB_(pQil)_ carried *bla*_NDM-1_ among 30 isolates in 6 strain clusters, while pHB4 IncFIB_HI1B hybrid plasmid transmitted *bla*_NDM-5_ among 13 isolates in 4 clonal clusters. Two IncL/M plasmids, pHB1 and pHB2, transmitted *bla*_OXA-48_ across all lineages, suggesting chronic persistence of these replicons within HBH. Global contextualization of HBH strains and AMR-transmitting plasmids further identified Mediterranean-focused transmission of *bla*_NDM-1_ by FII_K2_+FIB_(pQil)_, pKpQIL-like plasmids,^18^ while *bla*_OXA-48_ transmission occurred globally via IncL/M replicons (Supplementary data 1).^20^ In contrast, among HBH’s genomically analyzed CPKs, we did not detect IncX, and rarely IncC, which facilitate *bla*_NDM_ transmission in other regions.^19^

Our study confirmed the spread of high-risk MLSTs ST101 and ST147.^4,18^ We further identified emerging global, high-risk clusters of ST383 carrying *bla*_NDM-5_, *bla*_OXA-48_, on known virulence plasmids. These latter strains accounted for 43% of HBH’s CPK sepsis isolates from 2019-2022.

The MDR ST383 strains were first detected in 2016 in Qatar,^21^ followed by reports in the UK and Lebanon in 2018,^22,23^ then Italy, Germany, France, Tunisia, Libya, Egypt, Saudi Arabia, Singapore, and the US over 2019-2023.^24,25^ Genomic analyses of these isolates suggest that *bla*_NDM-5_-carriage occurs in emerging and highly drug-resistant epidemic clones (clades C and D1). In addition, the recent increases in ST383 global prevalence correlate with carriage of OXA-48 and NDM-5, while the earlier clade D strains, which lack NDM, have decreased in prevalence. These ST383 provide a strong working example where international collaboration among low-resource African and Mediterranean countries has defined an emerging health threat.

While our analyses identified plasmid-associated virulence markers among 26% of our African CPKs, their contribution to patient sepsis and outcomes remains unclear, given the absence of clinical syndromes associated with hypervirulent *K. pneumoniae,* such as liver abscesses, endophthalmitis, or additional tissue-based, metastatic infections. These findings support the results of Kochan et al., which suggest that convergent strains do not behave as classical hypervirulent *K. pneumoniae*, potentially due to the absence of the full virulence repertoire, differences in capsule types, or other factors.^26^

Our findings identified a complex landscape of endemic strains with repeated sporadic introduction of CPK into HBH, with capacity for entering strains to become established in the hospital. These findings highlight the need to screen high-risk patients for CPK carriage to inform local IPC strategies. In particular, ICU admission provides an established focal point to conduct CPK surveillance for this at-risk population. In addition, the clinical microbiology laboratory’s PCR-based testing for carbapenemases, with additional rapid molecular strain typing, can inform cohorting patients colonized with the high-risk lineages—ST101, ST147, and ST383 to provide adaptable IPC strategies in hospital environments seen in resource-limited countries.

Limitations of our studies include the focus on bloodstream isolates, which represented 25% of all CPK isolates seen at HBH. While phenotypic and PFGE analyses indicated the sepsis isolates to be reflective of our CPK population, this approach could have detected additional circulating clones. Nevertheless, our study provides a working example for how high-resolution genomic analyses in resource-limited countries are enabled by publicly available resources and validated bioinformatic tools to achieve robust epidemiological surveillance. This approach underscores the need for global genomic surveillance to guide IPC strategies at hospital through international levels.

## Methods

### Clinical setting and study design

HBH is a referral tertiary 500-bed hospital with 22,000 admissions per year, serving a population of 1 million in South Tunisia. HBH includes 10 surgical and 3 medical wards, one medical intensive care unit (ICU) (22 beds), and one surgical ICU (5 beds). The SUD Committee of Persons’ Protection (C.P.P.SUD) approved the study (reference number 0192/2019). Patient cultures with the demographic information, including age, gender, and hospital admission, were retrieved from hospital records. We confirmed carbapenemase carriage clinically in all *K. pneumoniae* isolates with ertapenem minimal inhibitory concentrations (MICs) ζ 0·5 mg/L and PCR. Clonal relationships among blood CPK isolates were further evaluated by pulse-field gel electrophoresis (PFGE).^10^ Strains were selected by year from each pulsotype, phenotypic, and PCR profile for whole-genome sequencing (WGS).

### Antimicrobial susceptibility testing, PCR screening, and string test

Antimicrobial susceptibility testing was performed by disk diffusion methods or broth microdilution per the European Committee on Antimicrobial Susceptibility Testing. PCR-based analyses evaluated carriage of carbapenem resistance genes (*bla*_NDM,_ *bla*_IMP_, *bla*_KPC,_ *bla*_VIM_, and *bla*_OXA-48-like_), aminoglycoside methylase genes (*armA*, *rmtA*, *rmtB*, *rmtC*, *rmtD*, *rmtE*, *rmtF*), and virulence genes (*rmpA*, *rmpA2*, *iutA*, *ybtS*, *mrkD*).^27,28^ String test for the hypermucoviscous phenotype used colonies cultured on MacConkey agar plates as described.^29^

### Conjugation assays

*In vitro* assays for transfer of carbapenemase genes from blood CPK isolates used rifampicin-resistant *Escherichia coli* J53-2 recipients. Transconjugants were selected on imipenem (2 μg /ml) and rifampin (250 μg/ml) and confirmed by PCR. We classified plasmids according to their incompatibility group by using the PCR-based replicon typing method as described previously.^30^

### Genomic analyses

Genomic DNA was extracted using QIAamp DNA Mini Kit (Qiagen) and sequenced by 150bp paired-end Illumina NovaSeq. Raw data were quality-checked with FastQC.^31^ and *de novo* assembled with SPAdes (v.3.14.0).^32^ Kleborate v2.1, using default settings, identified strain multilocus sequence and capsule locus types, AMR, and virulence genes.^33^

For cluster and outbreak analyses, we used the FDA Center for Food Safety and Nutrition’s (CFSAN) Single Nucleotide Polymorphism (SNP) Pipeline in GalaxyTrakr.^34,35^ Strain SNP alignments were mapped to the study isolate with the highest N50 score, and used an established cutoff of 21 SNPs to define clonal clusters.^36^ Phylogenetic trees were constructed using FastTree 2.0, visualized and annotated using iTOL (v6).^37^ Sequencing reads were deposited to NCBI’s Sequence Read Archive (Bioproject PRJNA1160997) for rapid SNP clustering in NCBI Pathogen Detection to place strains within an international context, and call resistance genes.^38,39^

To validate the findings of the GalaxyTrakr SNP Pipeline and the NCBI Pathogen Detection tools, we conducted whole-genome MLST (wgMLST) and SNP analyses of the HBH isolates alongside the 100 closest genomes from the NCBI SRA Supplementary data 2 Table S8). From NCBI’s nucleotide database, 86,290 putative *Klebsiella pneumoniae* isolate genomes were downloaded as identified in the Pathogen Isolates browser on February 13^th^, 2025 (Supplementary data 2 Table S8).^38^ From these, 84,393 were designated as *Klebsiella* by the program MLST, which was used to assign MLST types according to the Pasteur Institute typing scheme.^40–42^ For assembly quality control, two criteria were used: 1) 80 contigs or fewer contained more than 90% of the sequence information; 2) total sequence length between 5.0378 and 6.2526 MB, approximating three standard deviations from the median sequence length plus and minus of available assemblies. Using these cutoffs, 80,252 sequence assemblies were used for phylogenetic analyses [Supplementary data 2 Table S8].

Gene coding sequences (n = 5316) from the reference genome CP003200.1 were used to create a wgMLST scheme.^43^ BLAST was used to identify gene sequence alignments that had an e-value under 0.001 and were all gapless, unique, and full-length.^44^ Distinct gene sequences were given a sequence type identity for wgMLST analyses.^45^ For phylogenetic estimation, SNP matrices were composed based off these sequences, replacing missing sequences with ‘N’ characters. Trees were reduced to only sequence positions containing SNPs. Trees were calculated using RAxML-ng and its GTR+G substitution model with automatic bootstrap analysis to 1000 bootstraps, and transfer bootstrap expectation as the branch confidence metric.^46^

Closely related strains were identified with ranking by perfect-match wgMLST allele counts relative to a selected seed strain. For SNP clustering analysis, the list of ranked strains included was expanded from the seeded strain until 100 strains submitted outside this study were included. For ST383 analysis, a seed was chosen and the list expanded until the total number of genomes was 400, which was confirmed to include all 248 ST383-typed assemblies. This set of 400 was pruned to just those identified by outgroup analysis using a cluster of six ST376 assemblies (Supplementary data 1, Tree 42). Phylogenetic tree visualizations were created using the Interactive Tree of Life with minor cosmetic alterations.^37^ Trees without known outgroups were analyzed using IToL midpoint rooting. For ST383 outgroup identification, several ML trees produced by RAxML without bootstrapping analysis were analyzed, which all indicated that a cluster of ST376 genomes was an appropriate outgroup. Global comparative genomic analysis of ST383 isolates was performed using RAxML-ng.

### Plasmid Analyses

Plasmid replicon types were identified with PlasmidFinder 2.1.^47^ and MOB-suite using the MOB-recon and MOB-typer modules to identify carbapenemase-carrying plasmids from the draft assemblies,^48^ per homology to plasmid markers (replicons, relaxases) and plasmids in the reference NCBI plasmid database.^49^ To aid in the visualization of plasmid alignments to a reference plasmid, we developed a program named PlasMap, available at https://github.com/worleyjn/PlasMap. This program uses the Genome Diagram package implemented in BioPython to visualize where contigs have aligned to reference plasmids, visualize the reference plasmids, visualize contigs with sequence not represented on the reference plasmid, and simultaneously observe plasmid gene content.^50,51^ Reference plasmids and plasmid feature calls were downloaded from the NCBI Nucleotide database (Supplementary data 2 Table S7). BLASTn was used to align draft genome contigs to reference plasmid sequences with cutoffs of an e-value < 0.001 and length > 500.^44^ Plasmid-associated draft assembly contigs with any features < 60% represented in sequence alignments were analyzed for unique gene content. Files produced in PDF format were edited when necessary for clarity.

### Statistics and visualization

Significant differences in counts-based categorical variables were evaluated in R (v4.4.2) by Chi-squared or Fisher’s exact test with use of tidyverse and ggsankey for visualization.^52^ Phylogenetic trees were visualized in ITOL v6.^37^ NCBI’s Pathogen Detection Isolates Browser evaluated the global context of dominant clones seen in HBH. PlasMap generated visual representations of the strain contigs aligned to homologous reference plasmids.^38,49^

## Data availability

WGS data can be found under NCBI BioProject PRJNA1160997

## Funding statement/Acknowledgements

Analyses were supported by the Research Funds of the Ministry of Health for the Research Laboratory « Micro-organismes et Pathologies Humaines » of Habib Bourguiba University Hospital and NIDDK grant P30 DK034854 (Bry). The work of Jay Worley was supported by the National Center for Biotechnology Information of the National Library of Medicine (NLM), and the National Institute of Allergy and Infectious Diseases, National Institutes of Health. We thank Drs. Nicole Pecora, Michael Monuteaux, and Marc Allard for critical reading of the manuscript and helpful insights.

## Author contributions

B.M., F.M., and A.H. contributed to data collection. B.M. and N.B.M. conducted experiments. B.M and J.N.W. performed data analysis and interpreted the results under the scientific guidance of L.B. B.M. and J.N.W. drafted and edited the paper, and L.B. contributed to the structuring and editing of the paper. All authors read and approved the final draft.

## Competing interests

The authors declare no competing interests.

## Notes

### Competing Interest Statement

The authors have declared no competing interest.

### Author Declarations

The SUD Committee of Persons Protection (C.P.P.SUD) approved the study (reference number 0192/2019)

