## Supplementary for "Leveraging the global genomic epidemiology of carbapenemase-producing *Klebsiella pneumoniae* to inform infection prevention in Tunisian hospitals"

### Supplementary Figures

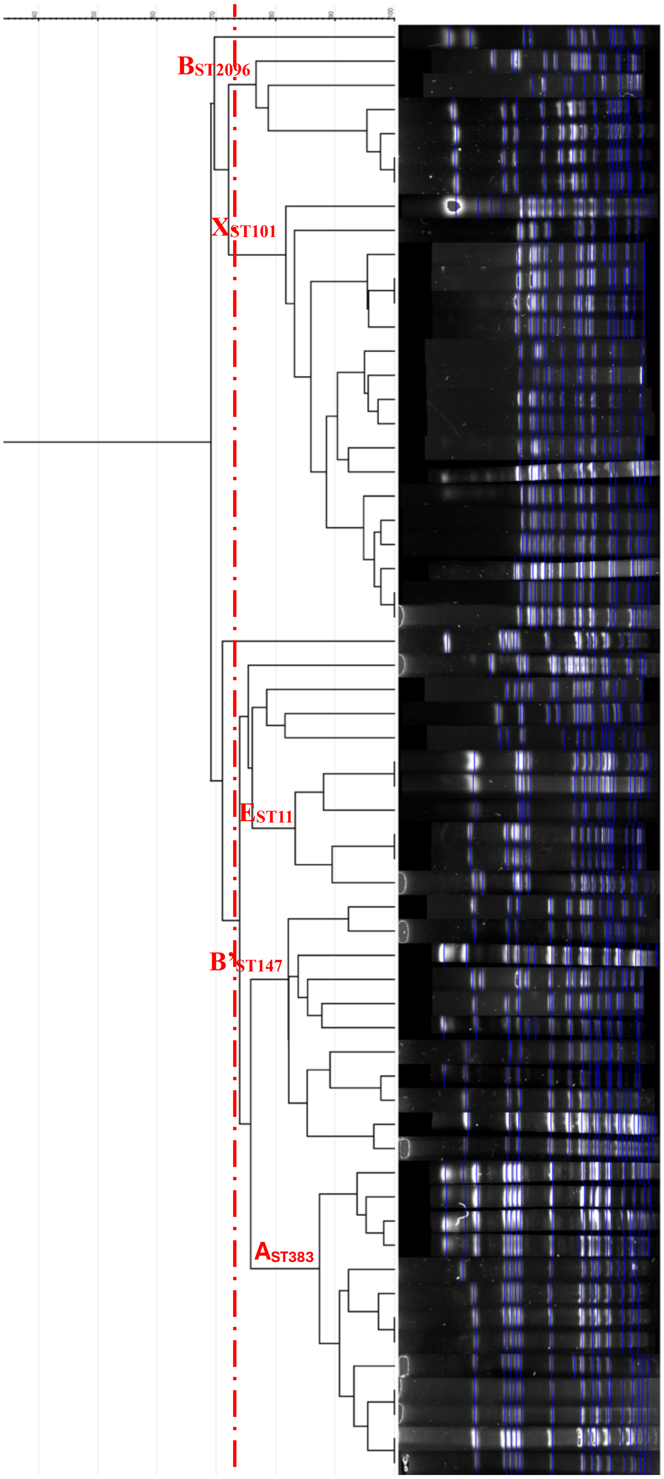

| Strain Number | WARD | CLONE | WZI | OXA-48_NDM | ArmA |
|---|---|---|---|---|---|
| R2991/17 | ICU | DIF | null | NDM | - |
| R919/16 | ICU | J | null | NDM | - |
| R2101/18 | ICU | DIF | null | NDM | - |
| R3501/20 | ICU | B | 64 | OXA-48 | ArmA |
| R3820/20 | ICU | B | 64 | OXA-48 | ArmA |
| R3694/20 | ICU | B | 64 | OXA-48 | ArmA |
| R3672/20 | ICU | B | 64 | OXA-48 | ArmA |
| Z177/17 | SURG | X | 137 | OXA-48 | - |
| R3900/20 | ICU | X | 137 | OXA-48 | - |
| Z22/16 | SURG | X | 137 | OXA-48 | - |
| R316/16 | ICU | X | 137 | OXA-48 | - |
| R275/16 | ICU | X | 137 | OXA-48 | - |
| R4562/16 | ICU | X | 137 | OXA-48 | - |
| R2670/18 | ICU | X | 137 | OXA-48 | - |
| R3147/18 | null | X | 137 | OXA-48 | - |
| R2052/18 | ICU | X | 137 | NDM | - |
| O43/18 | ORTHO | X | 137 | OXA-48 | - |
| R4008/18 | ICU | X | 137 | OXA-48 | - |
| Z691/19 | SURG | X | 137 | OXA-48 | - |
| R3230/17 | ICU | X | 137 | OXA-48 | - |
| S17/17 | SAMU | X | 137 | OXA-48 | - |
| U3299/17 | URO | X | 137 | OXA-48 | - |
| U1847/19 | URO | X | 137 | OXA-48 | - |
| U2721/17 | URO | X | 137 | OXA-48 | - |
| Z859/21 | SURG | X | 137 | NDM | - |
| R3788/20 | ICU | DIF | 705 | OXA-48+NDM | ArmA |
| S329/21 | SAMU | DIF | null | OXA-48+NDM | ArmA |
| R363/18 | ICU | DIF | 24 | OXA-48 | - |
| R2648/17 | ICU | DIF | 24 | NDM | - |
| R7/18 | ICU | DIF | 177 | NDM | - |
| Z658/17 | SURG | E | 24 | OXA-48 | - |
| S996/17 | SAMU | E | 24 | NDM | - |
| R40/17 | ICU | E | 24 | NDM | - |
| R4427/16 | ICU | E | 24 | NDM | - |
| R743/16 | ICU | E | 24 | NDM | - |
| UB61/21 | BURNS | E | 24 | NDM | - |
| R428/18 | ICU | B' | 64 | OXA-48+NDM | - |
| RH16/21 | SICU | B' | 64 | NDM | - |
| R2381/19 | ICU | B' | 64 | NDM | - |
| R3610/16 | ICU | B' | 64 | NDM | - |
| U271/16 | URO | B' | 64 | NDM | - |
| R556/19 | ICU | B' | 64 | OXA-48+NDM | - |
| R110/20 | ICU | B' | 64 | OXA-48+NDM | - |
| R144/19 | ICU | B' | 64 | NDM | ArmA |
| R3309/20 | ICU | B' | 64 | NDM | ArmA |
| RH147/19 | SICU | B' | 64 | NDM | ArmA |
| R2820/21 | ICU | B' | 64 | NDM | ArmA |
| R487/19 | ICU | A | 705 | OXA-48+NDM | ArmA |
| R3731/19 | ICU | A | 705 | OXA-48 | - |
| R1312/19 | ICU | A | 705 | OXA-48+BDM | ArmA |
| Z144/19 | SURG | A | 705 | OXA-48+NDM | ArmA |
| R1789/20 | ICU | A | 705 | OXA-48+NDM | ArmA |
| R3981/20 | ICU | A | 705 | OXA-48+NDM | ArmA |
| R3959/20 | ICU | A | 705 | OXA-48+NDM | ArmA |
| R3741/20 | ICU | A | 705 | OXA-48+NDM | ArmA |
| Z81/21 | SURG | A | 705 | OXA-48+NDM | ArmA |
| R211/21 | ICU | A | 705 | OXA-48+NDM | ArmA |
| R1428/21 | ICU | A | 705 | OXA-48+NDM | ArmA |
| R627/21 | ICU | A | 705 | OXA-48+NDM | ArmA |
| R1655/21 | ICU | A | 705 | OXA-48+NDM | ArmA |

**Fig. S1: PFGE profiles of  Sepsis CPK isolates**

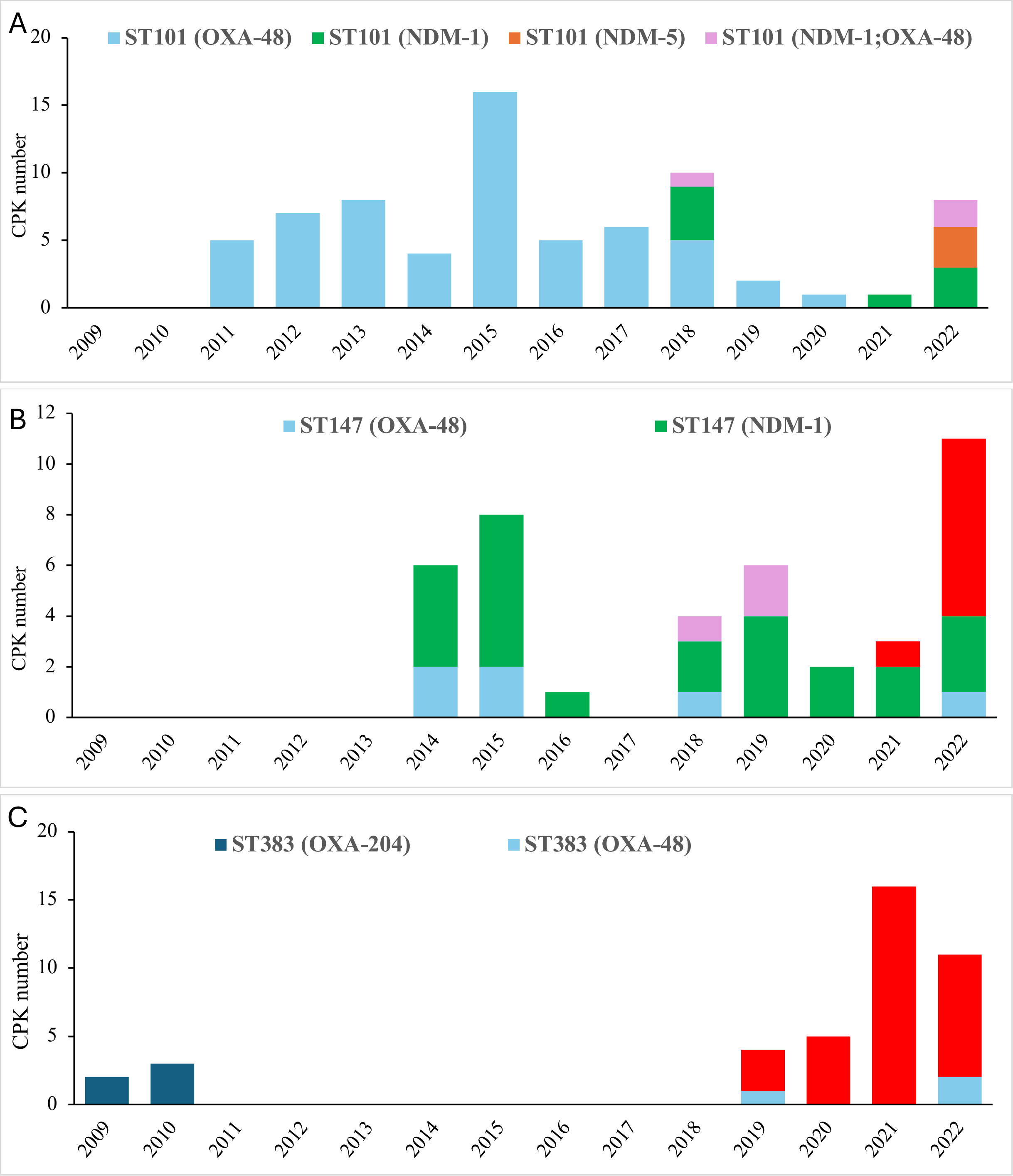

**Fig. S2: Temporal trends of ST101, ST147, and ST383 in HBH, 2009–2022.**

Legend: Frequency of dominant CPK sepsis clones over time, A: ST101, B, ST147, C: ST383, in Habib Bourguiba Hospital, Sfax, Tunisia. X axis shows years; Y axis shows the number of CPK isolated by year. Sample key shows the ST-carbapenemase type association.

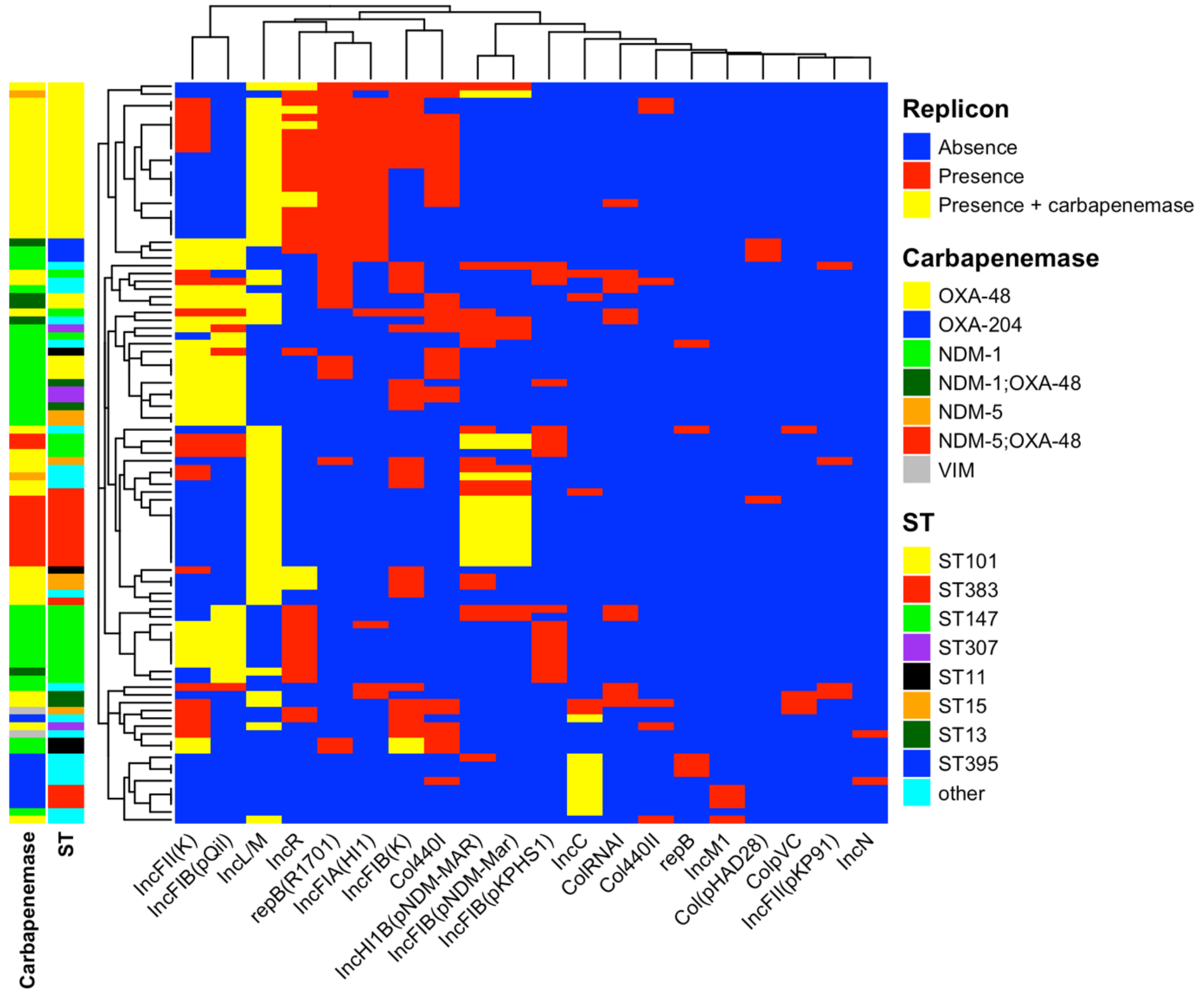

**Fig. S3: Heatmap demonstrating replicon distribution across CPK isolates by sequence type**

(ST - right-most column on the left-hand side) and carbapenemase type (left-most column on the left-hand side). Color key shows the main STs and carbapenemases. Red boxes in the 2D matrix represent presence of the replicon noted along the bottom axis, yellow the replicon harboring the carbapenemases, and blue the absence of the given replicon.

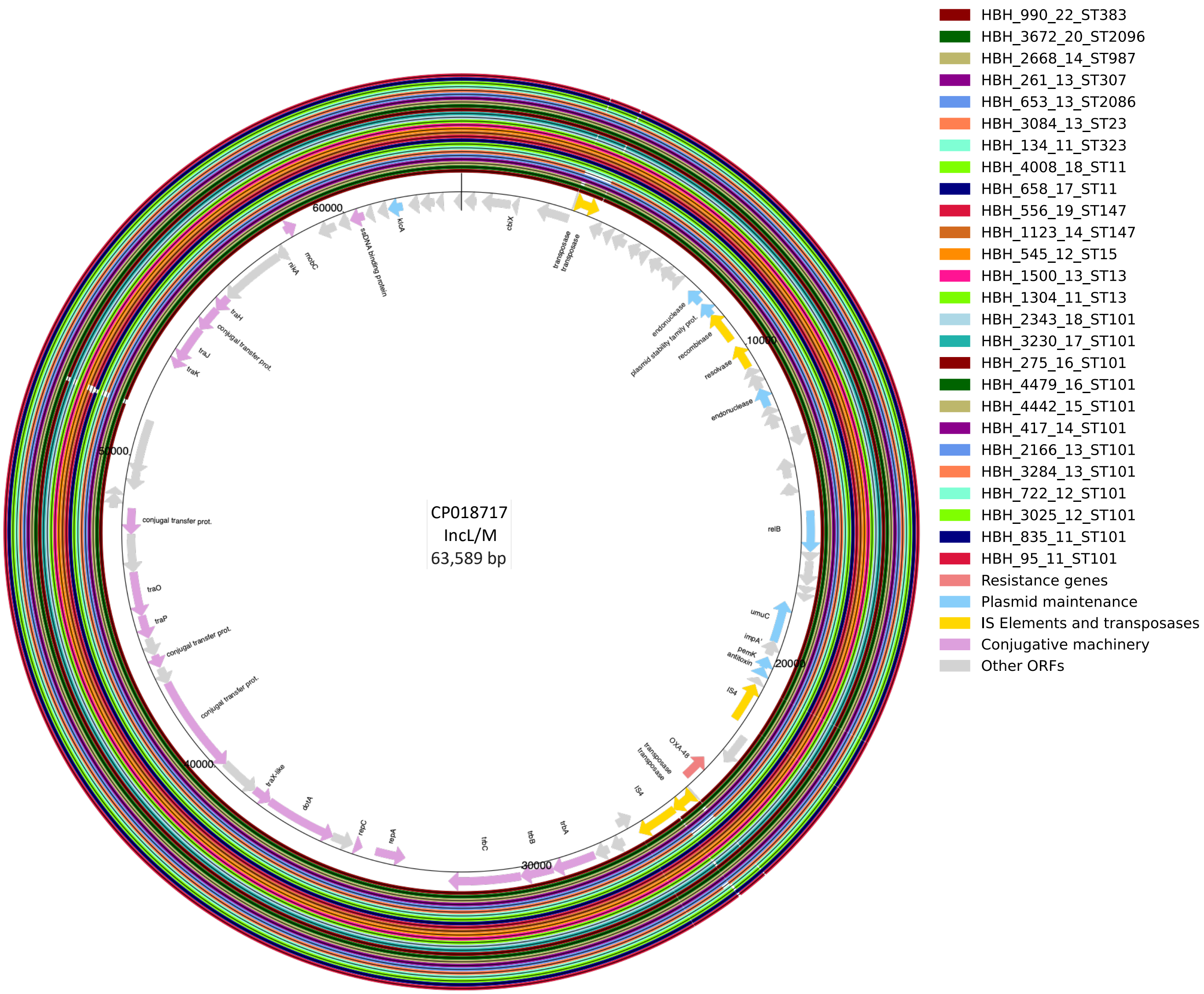

**Fig. S4: *bla*$_{OXA-48}$ IncL/M (Mobsuite ID AA018/AH562) plasmids from study isolates.**
Each ring corresponds to a plasmid from HBH CPK **(n=26 plasmids)**, identified on the right side of the figure along with the color code indicating, in order: strain identification, year of isolation, and sequence type (ST). Plasmid CP018717 was used as the reference. The genes are shown in the inner ring, represented by arrows indicating the direction of transcription.
PlasMap analyses show reads with >99% coverage and >99% identity for the IncL/M CP018717 plasmid.

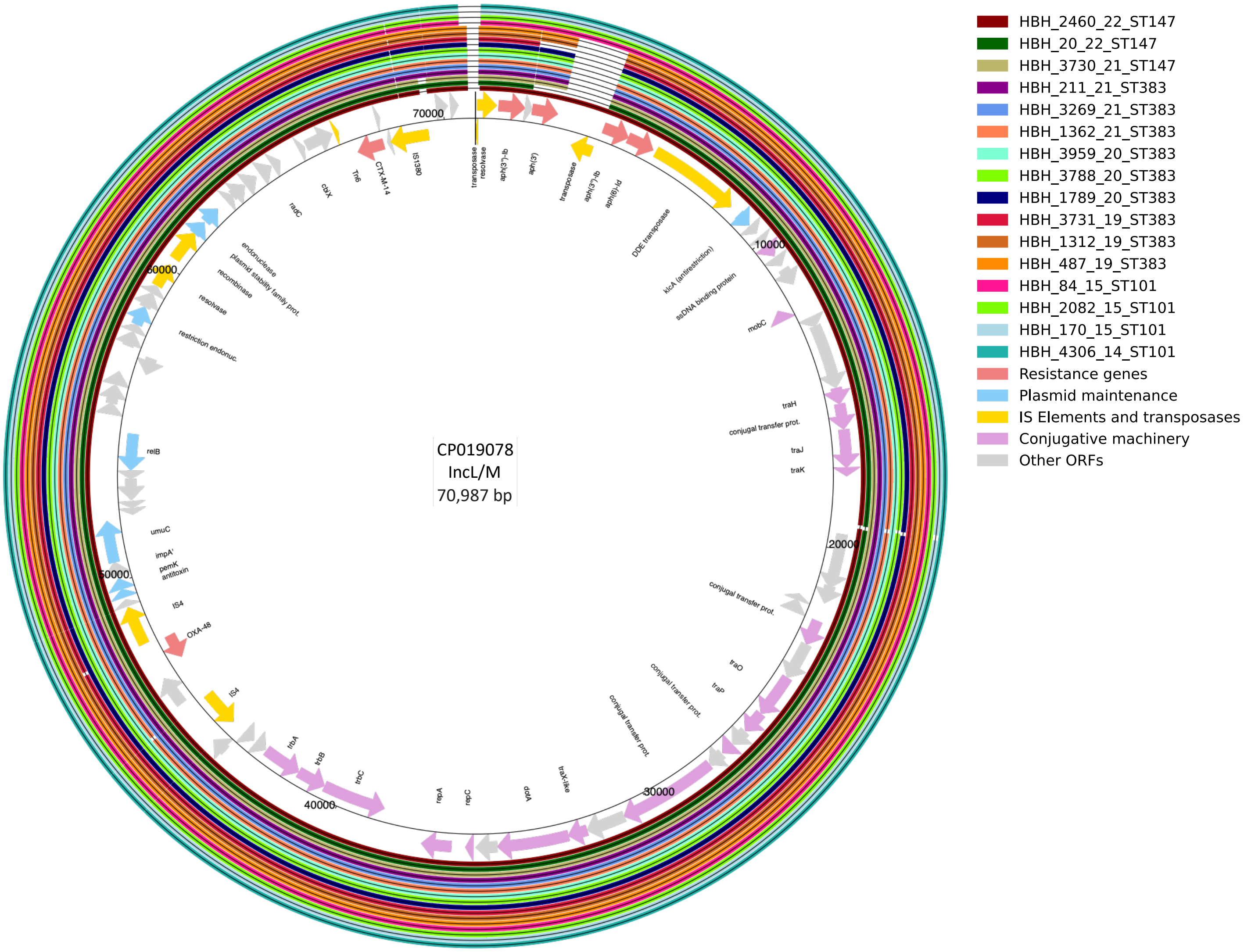

**Fig. S5: *bla*$_{OXA-48}$ IncL/M (Mobsuite ID AA002/AH529) plasmids from study isolates.**
Each ring corresponds to a plasmid from HBH CPK **(n=16 plasmids)**, identified on the right side of the figure along with the color code indicating, in order: strain identification, year of isolation, and sequence type (ST). Plasmid CP019078 was used as the reference. The genes are shown in the inner ring, represented by arrows indicating the direction of transcription.

PlasMap analyses show reads with >99% coverage and >99% identity for the IncL/M CP019078 plasmid.

A

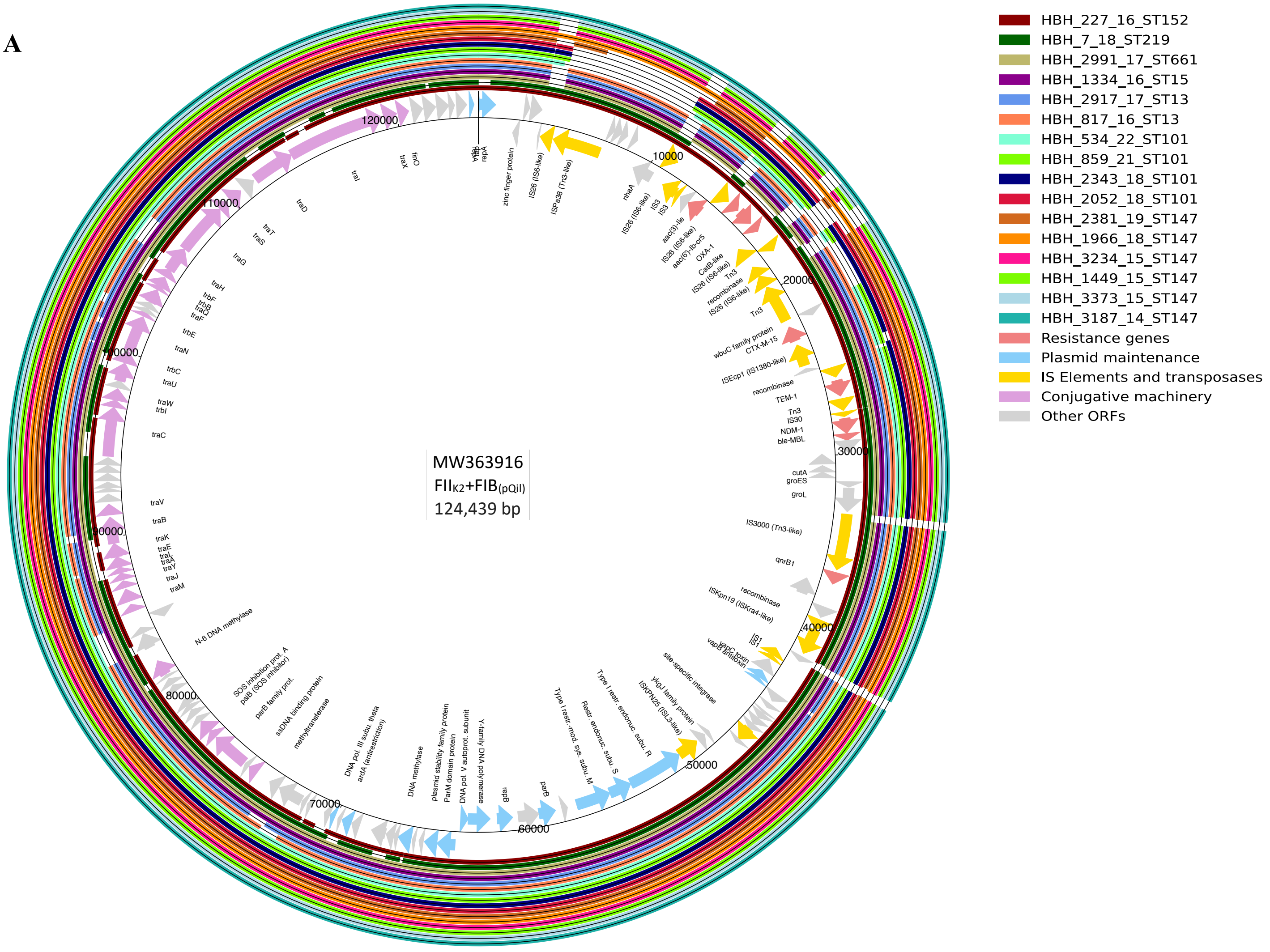

B

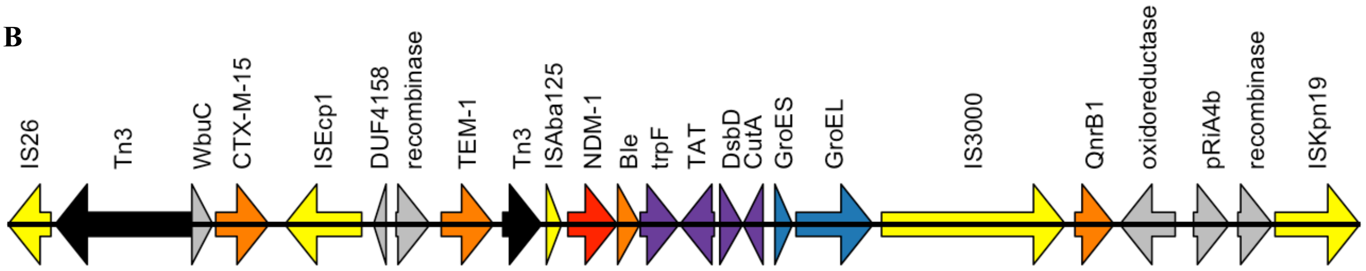

**Fig. S6: $FII_{K2}$+$FIB_{(pQil)}$ *bla*$_{NDM-1}$ plasmids (Mobsuite ID AA018/AH562) from study isolates.**
**A: $FII_{K2}$+$FIB_{(pQil)}$ *bla*NDM-1 plasmids (n=16 plasmids)**. Each ring corresponds to a plasmid from HBH CPK, identified on the right side of the figure along with the color code indicating, in order: strain identification, year of isolation, and sequence type (ST). Plasmid **MW363916** was used as the reference. The genes are shown in the inner ring, represented by arrows indicating the direction of transcription. **B: Genetic context of *bla*$_{NDM-1}$ in $FII_{K2}$+$FIB_{(pQil)}$ plasmids (AA018/AH562)**
PlasMap analyses show reads with >90% coverage and >99% identity for the IncFII$_{K2}$+FIB$_{K}$ MW363916 plasmid.

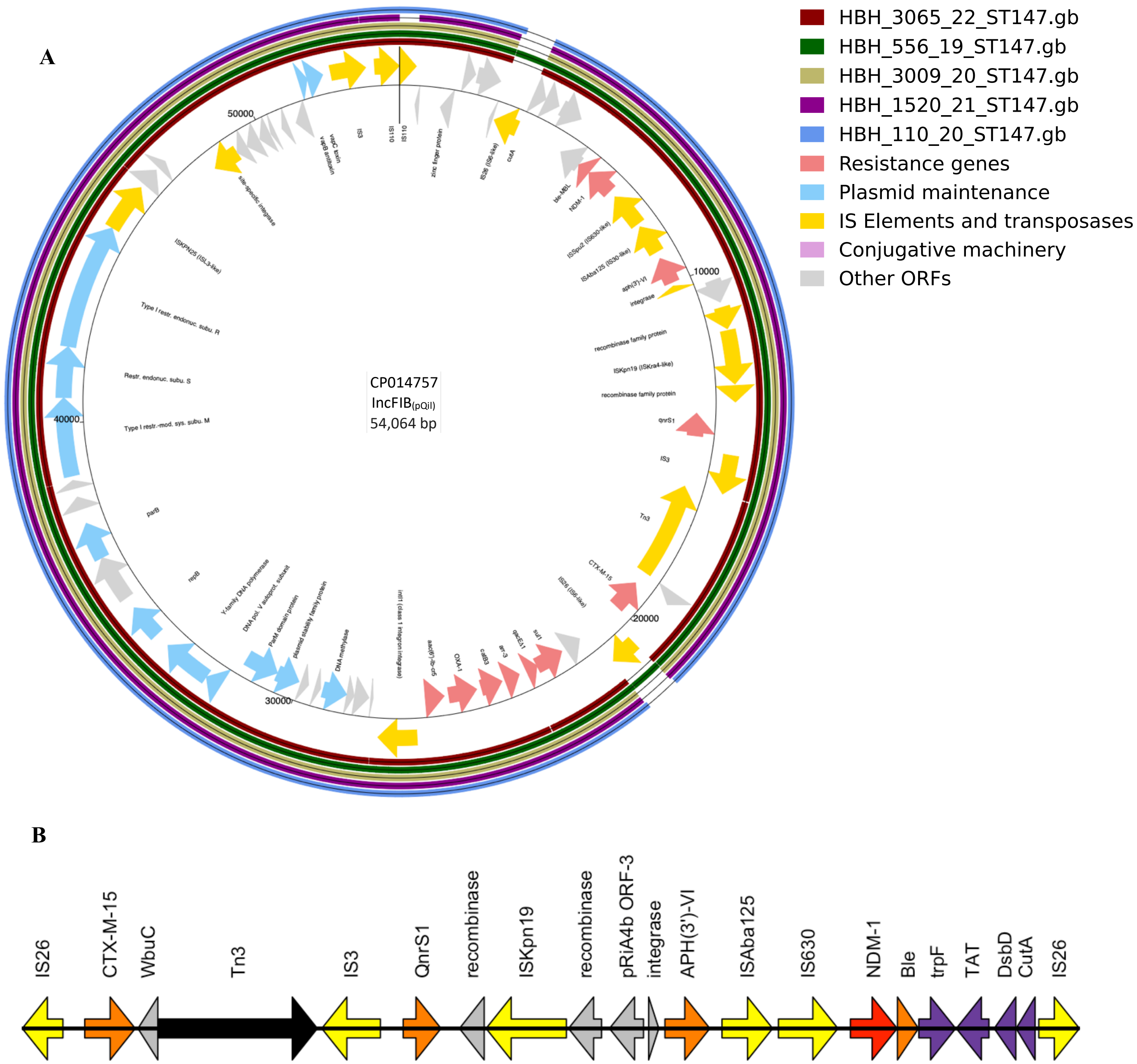

**Fig. S7: *bla*$_{NDM-1}$ IncFIB$_{(pQil)}$ (AA019/AH565) from study isolates.**
**A: *bla*$_{NDM-1}$ IncFIB$_{(pQil)}$ (n=5 plasmids)**. Each ring corresponds to a plasmid from HBH CPK, identified on the right side of the figure along with the color code indicating, in order: strain identification, year of isolation, and sequence type (ST). Plasmid **CP014757** was used as the reference. The genes are shown in the inner ring, represented by arrows indicating the direction of transcription. **B: Genetic context of *bla*$_{NDM-1}$ in IncFIB(pQil) plasmids.**
PlasMap analyses show reads with >96% coverage and >99% identity for the IncFIB$_{pQil}$ CP014757 plasmid.

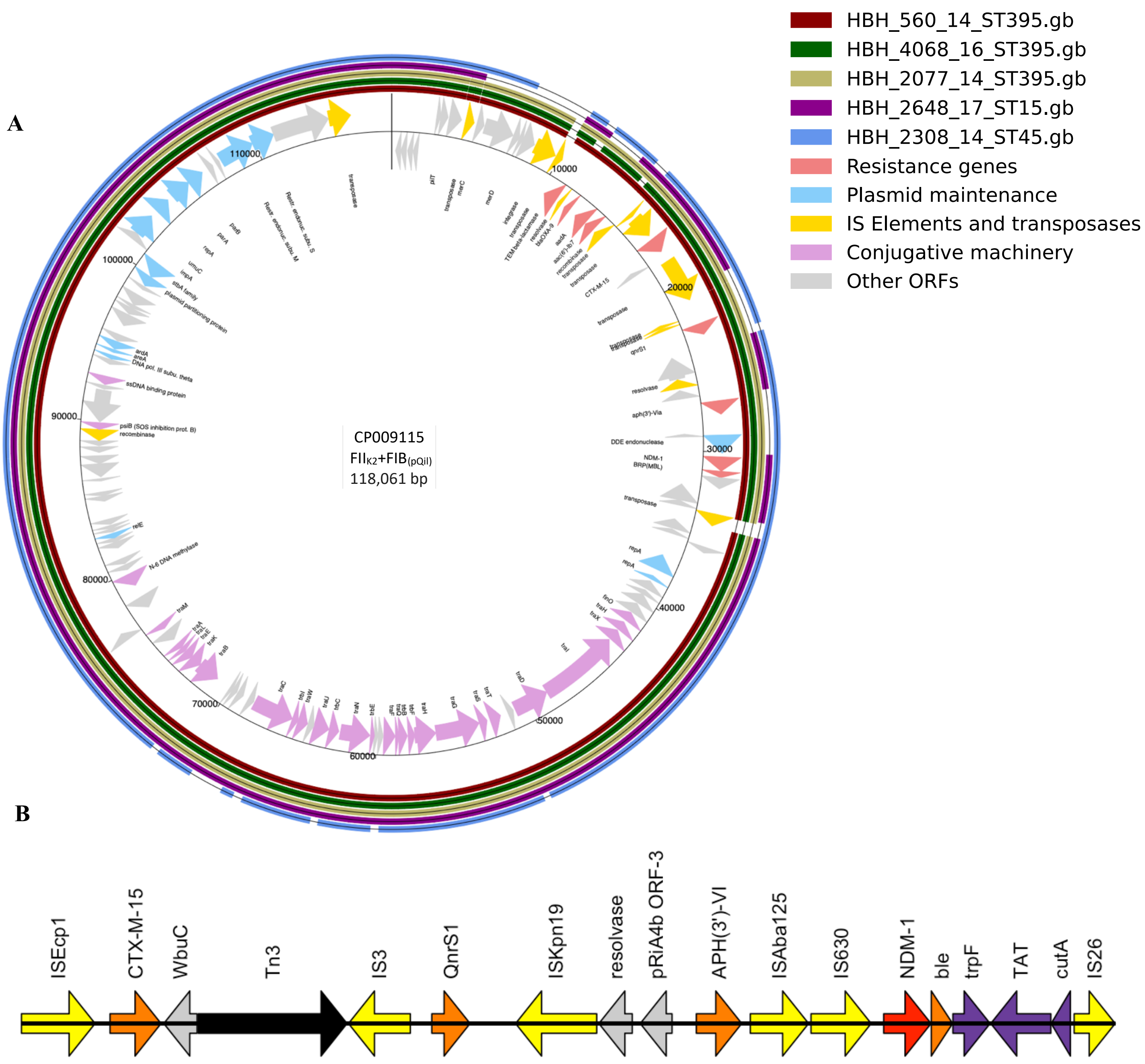

**Fig. S8: FII$_{K2}$+FIB$_{(pQil)}$ *bla*$_{NDM-1}$ plasmids (Mobsuite ID AA018/AH560) from study isolates.**
**A: FII$_{K2}$+FIB$_{(pQil)}$ *bla*$_{NDM-1}$ plasmids (n=5 plasmids)**. Each ring corresponds to a plasmid from HBH CPK, identified on the right side of the figure along with the color code indicating, in order: strain identification, year of isolation, and sequence type (ST). Plasmid **CP009115** (inner ring) was used as the reference. The genes are shown in the inner ring, represented by arrows indicating the direction of transcription. **B: Genetic context of *bla*$_{NDM-1}$ in FII$_{K2}$+FIB$_{(pQil)}$ plasmids (AA018/AH560)**
PlasMap analyses show reads with >87% coverage and >99% identity for the IncFII$_{K2}$+ FIB$_{(pQil)}$ CP009115 plasmid.

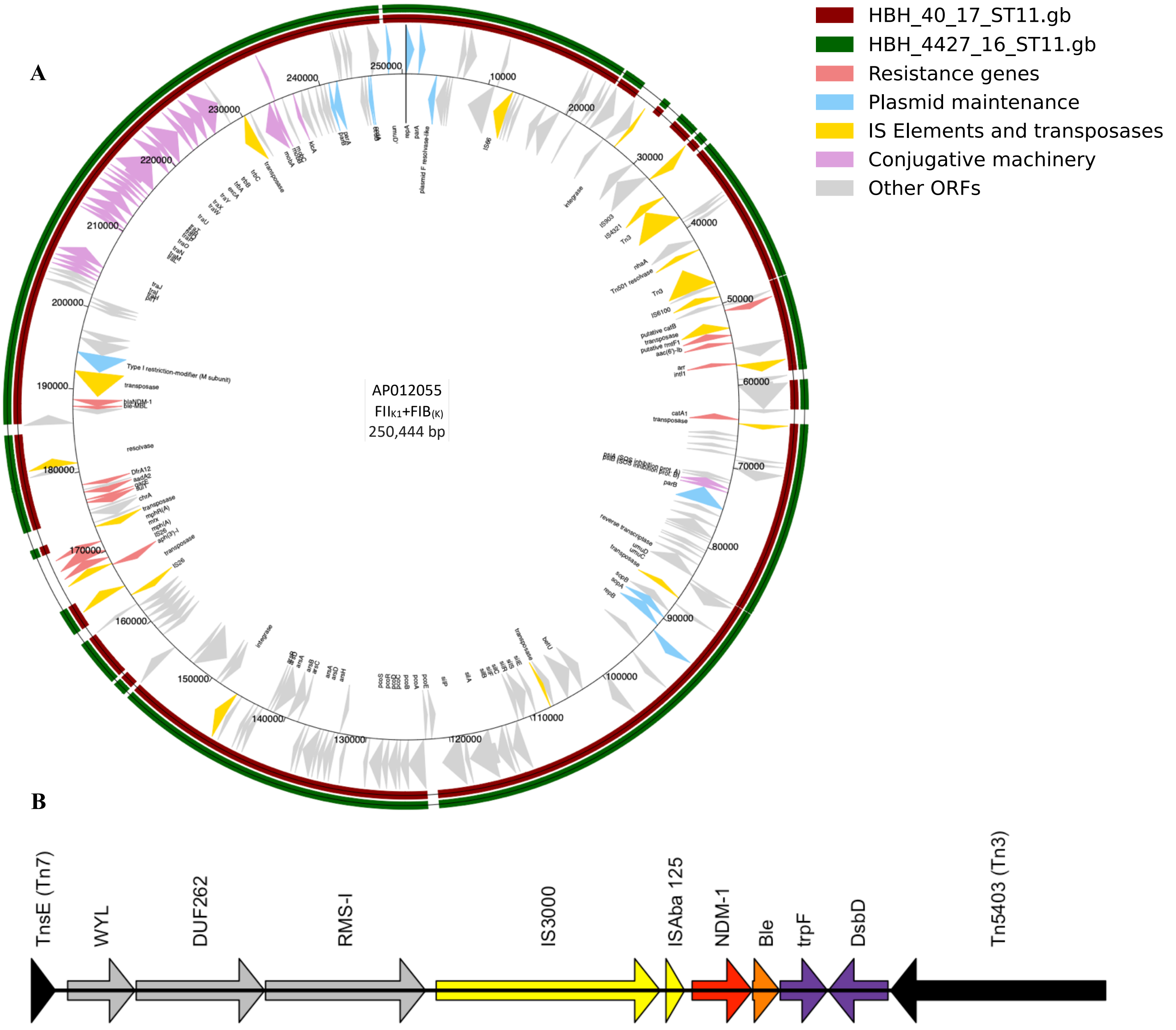

**Fig. S9: FII$_{K1}$+FIB$_{(K)}$ *bla*$_{NDM-1}$ plasmids (Mobsuite ID AA274/AI071) from study isolates.**

**A: FII$_{K1}$+FIB$_{(K)}$ *bla*$_{NDM-1}$ plasmids (n=2 plasmids)**. Each ring corresponds to a plasmid from HBH CPK, identified on the right side of the figure along with the color code indicating, in order: strain identification, year of isolation, and sequence type (ST). Plasmid **AP012055** (inner ring) was used as the reference. The genes are shown in the inner ring, represented by arrows indicating the direction of transcription.

**B: Genetic context of *bla*$_{NDM-1}$ in FII$_{K1}$+FIB$_{(K)}$ plasmids (AA274/AI071)**

PlasMap analyses show reads with >91% coverage and >99% identity for the IncFII$_{K2}$+FIB$_{K}$ AP012055 plasmid.

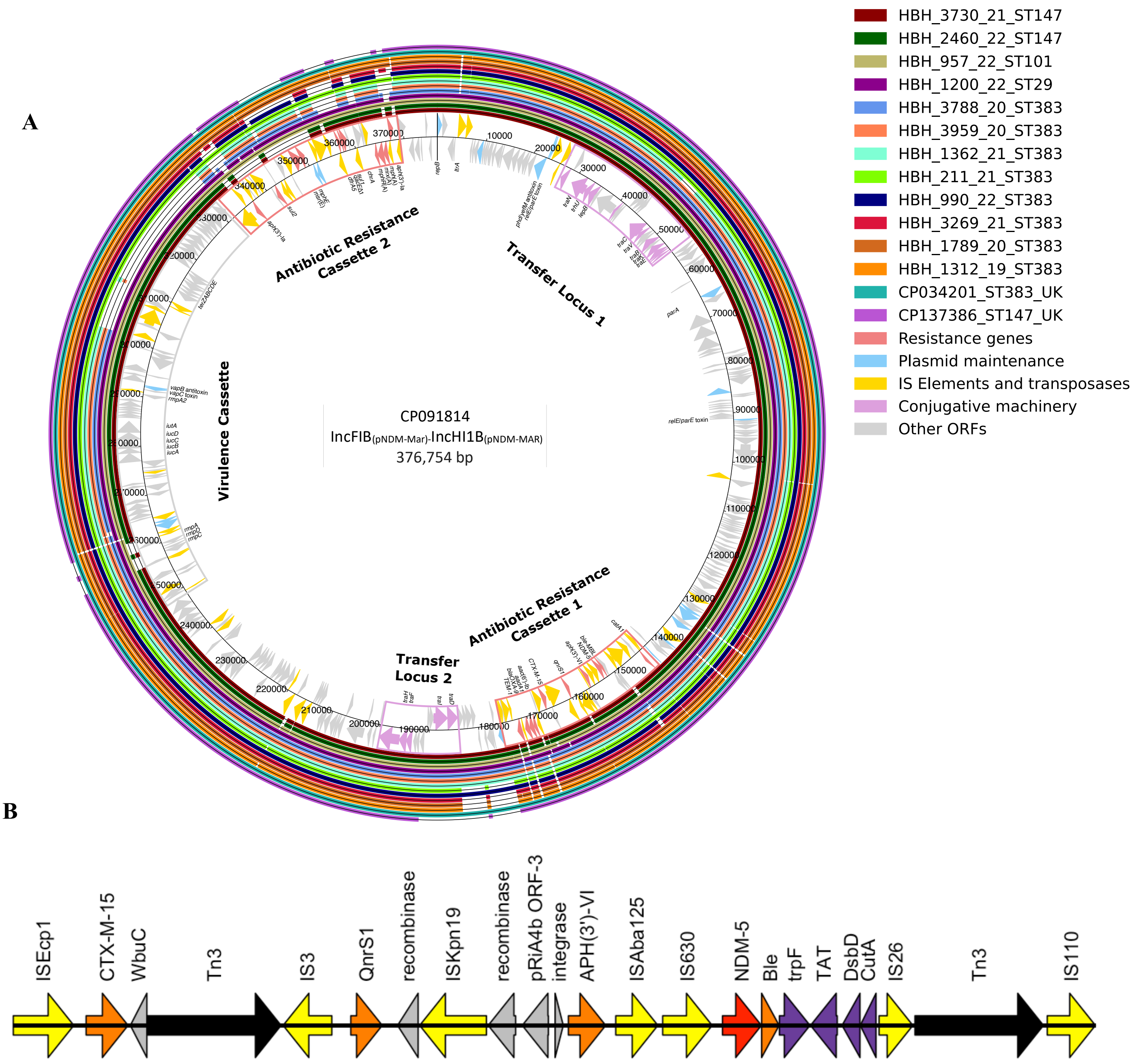

**Fig. S10: *bla*$_{NDM-5}$ IncFIB(pNDM-Mar)-IncHI1B(pNDM-MAR) plasmids from study isolates.**
**A: *bla*$_{NDM-5}$ IncFIB(pNDM-Mar)-IncHI1B(pNDM-MAR) plasmids (n=12 plasmids)**. Each ring corresponds to a plasmid from HBH CPK, identified on the right side of the figure along with the color code indicating, in order: strain identification, year of isolation, and sequence type (ST). Plasmid **CP091814** (inner ring) was used as the reference. The genes are shown in the inner ring, represented by arrows indicating the direction of transcription.
**B: Genetic context of *bla*$_{NDM-5}$ in IncFIB(pNDM-Mar)-IncHI1B(pNDM-MAR) plasmids**
PlasMap analyses show reads with >86% coverage and >98% identity for the IncFIB-IncHI1B CP091814 plasmid.

#### Supplementary Tables

##### Table S1: Carbapenemase and virulence gene distribution by culture site

| | Total | Carbapenemase genes | | | | Virulence genes | | |
|---|---|---|---|---|---|---|---|---|
| | | *bla*NDM | *bla*OXA48 | *bla*NDM + *bla*OXA4 | *bla*VIM | *rmpA2* | *rmpA* | *iuc* |
| Urine | 380 | 117 (30.8) | 156 (41.1) | 107 (28.2) | | 90 (23.7) | 66 (17.4) | 86 (22.6) |
| Blood | 220 | 64 (29.1) | 102 (46.4) | 52 (23.2) | 2 | 61 (27.7) | 45 (20.5) | 56 (25.5) |
| Sputum | 152 | 23 (15.1) | 80 (52.6) | 49 (32.2) | | 50 (32.9) | 36 (23.7) | 46 (30.3) |
| Skin, soft tissue | 189 | 50 (25.3) | 97 (49.0) | 51 (25.8) | | 45 (23.8) | 34 (18.0) | 39 (20.6) |
| Other | 63 | 27 (42.9) | 22 (34.9) | 14 (22.2) | | 17 (27.0) | 15 (23.8) | 15 (23.8) |
| P value | | 0.0002 | 0.03901 | 0.3603 | | 0.1782 | 0.3591 | 0.2039 |

P-values show the comparison of carbapenemase or virulence genes between the different types of culture sites.

**Table S2: PFGE pulsotypes of Sepsis CPK isolates**

| Year | CKP Number | Number of strains per PFGE profile (number of sequenced strains) | Number of sequenced strains per year |
|---|---|---|---|
| 2009 | 3 | 2 $F_{ST383}$ (1), 1 $V_{ST340}$ (1) | 2 |
| 2010 | 7 | 3 $F_{ST383}$ (1), 2H $_{ST218}$ (2), 1 $V_{ST340}$ (1), 1 unique (1) | 5 |
| 2011 | 8 | 3 $A'_{ST101}$ (1), 2 $D_{ST101}$ (1), 2 S (1), unique (1) | 4 |
| 2012 | 11 | 7 $A'_{ST101}$ (2), 3 unique (2), 1 NT (1) | 5 |
| 2013 | 15 | 4 $A_{ST101}$ (1), 4 $D_{ST101}$ (2), 6 unique (6) | 9 |
| 2014 | 17 | 3 $D_{ST101}$ (1), 5 $B'_{ST147}$ (1), 2 $K_{ST395}$ (2), 2 unique (2), 3 NT (2) | 9 |
| 2015 | 32 | 12 $D_{ST101}$ (3), 3 $A_{ST101}$ (1), 7 $B'_{ST147}$ (3), 2 $Q_{ST11}$ (1), 6 unique (3) | 10 |
| 2016 | 17 | 3 $A_{ST101}$ (1) , 2 $X''_{ST101}$ (1), $B'_{ST147}$ (1), 2 $H_{ST152}$ (1), 3 E $_{ST11}$ (1), 3 $J_{ST15}$ (1), 1 K $_{ST395}$ (1), 2 unique (2) | 8 |
| 2017 | 12 | 5 $X_{ST101}$ (1), 1 $X''_{ST101}$ (1), 2 $E_{ST11}$ (1), 1 $E'_{ST11}$ (1), 1 $J_{ST15}$ (1), 2 unique (2) | 7 |
| 2018 | 16 | 7 $X_{ST101}$ (2), 1 $X''_{ST101}$ (1), 2 $B'_{ST147}$ (1), 3 unique (2), 3 NT (2) | 8 |
| 2019 | 13 | 6 $B'_{ST147}$ (2), 4 $A_{ST383}$ (3), 2 $X_{ST101}$, 1 NT | 5 |
| 2020 | 13 | 5 $A_{ST383}$ (3), 5 $B_{ST1096}$ (1) , 2 $B'_{ST147}$ (2), 1 unique (1) | 7 |
| 2021 | 21 | 15 $A_{ST383}$ (3) , 2 $B'_{ST147}$ (1), 1 $D_{ST147}$ (1), 1 $X_{ST101}$ (1), 2 unique (1) | 7 |
| 2022 | 35 | 11 $A_{ST383}$ (2), 3 $B'_{ST147}$ (1), 8 $D_{ST147}$ (2), 3 $X''_{ST101}$ (1), 5 $X_{ST101}$ (1), 5 unique (1) | 8 |

The table summarizes the main PFGE profiles, the number of strains per profile, and the unique pulsotypes, indicating in parentheses the number of sequenced strains per PFGE.

**Table S3: Characteristics of the ST383 *K. pneumoniae* isolates reported in the 42 studies published in PubMed by June 5, 2025**

| Reference | Country | Setting | Study isolates | Year | Collection site | ST383 isolates | | | | | |
|---|---|---|---|---|---|---|---|---|---|---|---|
| | | | | | | Number | ST383 % among CRK | ST383 % among all KPN | ST383 Outbreak | Carbapenemase types | Virulence markers (*rmpA/iuc*) |
| **Rotondo 2024** [1] | **Italy** | **19 hospitals in the Lazio region** | **126 NDM-Kpn isolates** | **2020 and 2023** | **diverse** | **6** | **4.8%** | **-** | **-** | **NDM5 (2); NDM1 (4)** | **1 (16 %)** |
| Padovani 2023 [2] | Italy | Brescia | 6 Kpn resistant to cefiderocol | 2021 and 2022 | diverse | 2 | - | - | - | NDM1-OXA48 (2) | 0 |
| Spaziante 2021 [3] | Italy | Lazio region hospitals | all inpatients with NDM-Kpn strain (17 NDM-Kpn) | January 2019 and June 2020 | diverse | 4 | 23.5% among NDM-Kpn | - | - | NDM5 (1), NDM1-OXA48 (3) | NS |
| **Lorenzin 2022** [4] | **Italy** | **IRCCS San Raffaele Scientific Institute, Milan** | **4 XDR Hyperviruent Kpn isolates** | **2019** | **recta swab** | **2** | **-** | **-** | **-** | **NDM5-OXA48 (1) ; NDM1-OXA48 (1)** | **2 (100 %)** |
| **Ventura 2022** [5] | **Italy** | **Verona Hospital** | **19 hypermucoviscous Kpn isolates** | **2021** | **blood cultures and abscesses** | **1** | **-** | **-** | **-** | **VIM1 + NDM5 (1)** | **1 (100 %)** |
| **Turton 2019** [6] | **UK** | **South East England** | **12 CPK with hybrid virulence plasmids** | **2016-2017** | **diverse** | **3** | **-** | **-** | **-** | **NDM5-OXA48 (2), OXA48 (1)** | **3 (100 %)** |
| **Hammad 2025** [7] | **Egypt** | **ICU at Assiut University Hospital** | **case report** | **2015** | **endotracheal aspirates** | **1** | **-** | **-** | **-** | **NDM5-OXA48 (1)** | **1 (100 %)** |
| **Attalla 2023** [8] | **Egypt** | **ICUs in Alexandria** | **17 colistin-resistant Kpn isolates** | **2020 (6 months)** | **diverse** | **7** | **-** | **-** | **-** | **NDM5-OXA48 (7)** | **6 (85 %)** |
| **Attalla 2024** [9] | **Egypt** | **Alexandria Main University Hospita** | **7 seleceted colistin-resistant Kpn isolates** | **2021** | **diverse** | **2** | **-** | **-** | **-** | **NDM5-OXA48 (2)** | **NS** |
| **Abdelsalam 2024** [10] | **Egypt** | **Microbiology laboratory in Alexandria** | **19 selected CRK** | **August 2020, and April 2021** | **diverse** | **5** | **26.3 %** | **-** | **-** | **NDM5-OXA48 (3) ; NDM1-OXA48 (2)** | **5 (100%)** |
| **Gamaleldin 2024** [11] | **Egypt** | **Alexandria Main University Hospital** | **27 sequenced among 56 MDR Kpn isolates** | **2019 et 2021** | **diverse** | **4** | **-** | **14.8 % among MDR-Kpn** | **-** | **NDM1 (1); OXA48 (1); NDM5-OXA48 (1)** | **NS** |

| | | | | | | | | | | | |
|---|---|---|---|---|---|---|---|---|---|---|---|
| Edward 2022 [12] | Egypt | Mabaret Al-Asafra Hospitals | 23 3GC resistant Kpn isolates, one selected for WGS | 2020 | diverse | 1 | - | - | - | OXA48 (1); NDM5-OXA48 (1) | NS |
| AhmedMAEE 2021 [13] | Egypt | Demerdash Hospital (Cairo, Egypt) | 34 Kpn isolates | June 2017 and March 2018 | Blood | 5 | | 14.7 % among sepsis-Kpn | - | NP | NS |
| Osman 2023 [14] | Sudan | 5 hospitals in Khartoum | 86 Kpn isolates (68 MDR) | 2016- 2020 | diverse | 5 | - | - | - | NDM5-OXA48 (4) | 2 (20 %) |
| Sobh 2024 [15] | Lebanon | American University of Beirut Medical Center | 34 ceftazidime-avibactam resistant Kpn isolates (17 sequenced) | 2019 and 2021 | diverse | 12 | - | - | - | NDM5-OXA48 (12) | 3 (25 %) |
| Dagher 2019 [16] | Lebanon | Saint George Hospital in Beirut | 5 cases of MDR Kpn isolates | 2017 | diverse | 5 | - | - | - | NDM5 (5) | NS |
| Elgriw 2023 [17] | Lybia | 1 hospital (TUH) | 44 CRK | 2019 and 2021 | diverse | 6 | 13.6 % | - | - | OXA48 (2); NDM5-OXA48 (3) | yes (2 sequenced) |
| Eltai 2020 [18] | Qatar | Hamad Medical Corporation | 18 colistin-resistant Kpn isolates | NP | diverse | 3 | - | - | - | OXA48 (1); NDM5-OXA48 (1) | |
| Tsui 2023 [19] | Qatar | Hamad Medical Corporation | 95 CPK | April 2016 to October 2017 | diverse | 4 | 4.2 % | - | - | 4 NDM5-OXA48 (4) | 4 (100 %) |
| Abid 2021 [20] | Qatar | Hamad Medical Corporation | 81 CPK | April 2014 toNovember 2017 | diverse | 4 | 4.9 % | - | - | OXA48 (1); NDM5 + OXA48 (3) | NS |
| SidAhmed 2024 [21] | Qatar | Hamad Medical Corporation | 3 XDR Kpn isolates | NP | urine and Respiratory tract | 1 | - | - | YES | NDM5-OXA48 (1) | NS |
| Huang 2024 [22] | Saudi Arabia | 34 KSA hospitals | 352 MDR Kpn isolates | January 2022 and April 2023 | Blood and urine | 2 | - | 0.6 % among MDR-Kpn | - | NDM5-OXA48 (2) | 2 (100 %) |
| Al-Zahrani [23] | Saudi Arabia | tertiary hospital in Jeddah | 29 selected CPK | NA | diverse | 2 | - | - | - | NDM5-OXA48 (2) | 2 (100 %) |
| Alghoribi 2020 [24] | Saudi Arabia | King Khalid University Hospital | case report | NP | wound | 1 | - | - | - | KPC2 (1) | 2 (100 %) |

| | | | | | | | | | | | |
|---|---|---|---|---|---|---|---|---|---|---|---|
| Chiarelli 2020 [25] | France | Bicetre Hospital | one strain selected for in vitro studies | 2017 | Blood | 1 | - | - | - | KPC2 (1) | 0 |
| Bonnin 2020 [26] | France | France's National Reference Center for Antimicrobial Resistance | 63 nonduplicate KPC-*Kp* | 2018 | diverse | 4 | 6.4% among KPC-Kpn | - | - | KPC2 (4) | 0 |
| Sabirova JS [27] | Greece | Tzaneio General Hospital (TGH) | 12 CPK | 2010–13 | diverse | 12 | - | - | - | KPC2 (7); VIM19 (11) | 0 |
| Mavroidi 2016 [28] | Greece | Konstantopouleio-Patission hospital | 19 colistin and carbapenem-resistant Kpn isolates among 135 Kpn | July 2012 to December 2013 | diverse | 1 | - | - | - | KPC2 (1) | 0 |
| Papagiannitsis 2016 [29] | Greece | NP | Case report of VIM-19 in IncA/C | 2015 | NP | 1 | - | - | - | VIM19 (1) | NS |
| Pitt 2018 [30] | Greece and Brazil | Hygeia General Hospital, Athen and Instituto Dante Pazzanese de Cardiologia, Brazil | 19 colistin-resistant Kpn isolates | 2012-2014 | diverse | 2 | - | - | - | - | 0 |
| Xanthopoulou 2022 [31] | Germany | 6 hospitals | 39 CRK | 2016–2018 | diverse | 1 | 2.6 % | - | - | VIM-19 (1) | 0 |
| Giakkoupi 2010 [32] | Greece | 40 Greek hospitals | 378 KPC-2 Kpn isolates | January 2009-April 2010 | diverse | 9 | 2.4 % among KPC-Kpn | - | - | KPC2 (2) | NP |
| Papagiannitsis 2010 [33] | Greece | Greece | Case report of ST383 producing VIM-4, KPC-2 and CMY-4 | 2009 | NP | 1 | - | - | - | VIM4 + KPC2 (1) | NS |
| Afolayan 2023 [34] | Greece | tertiary hospital in Athens | 211 CRK | 2003 and 2018 | diverse | 13 | 6.2 % | - | - | VIM-19 (13) | 0 |
| Baraniak 2015 [35] | Europe and Israel | multicentre project | Colonization with 110 KPC-Kp | 2008-2011 | fecal swabs | 3 | 2.7 % | | - | KPC2 (3) | NS |
| Wang 2021 [36] | China | multicentre resistance monitoring project | 34 selected OX48-Kpn isolates | 2013–2018 | diverse | 10 | 29.4% among OXA48-Kpn | - | - | OXA48 (10) | 6 (60 %) |
| Guo 2016 [37] | China | respiratory ICU in Beijing | 37 OXA48 Kpn isolates | 2013 and 2014 | diverse | 27 | - | - | **YES** | OXA48 (10) | NS |

| | | | | | | | | | | | |
|---|---|---|---|---|---|---|---|---|---|---|---|
| Palmieri 2019 [38] | China | 4000-bed Hospital in Beijing | 200 Kpn isolates | 2002–2016 | diverse | 16 | - | 8% | **YES** | OXA48 | 8 (50 %) |
| Gan 2022 [39] | China | 9 provinces of China | 232 Kpn isolates | 2013 to 2020 | liver abscess and pneumonia | 8 | - | 3.5% | - | OXA48 (6) | 8 (100 %) |
| Potron 2013 [40] | France | NP | one patient who had been hospitalized in Tunis | 2013 | urine | 1 | - | - | - | OXA204 (1) | NS |
| Österblad 2012 [41] | Finland | National Institute for Health and Welfare | All CPE send to the center (26 Kpn isolates) | 2008-11 | diverse | 1 | - | - | - | VIM (1) | NS |
| Samuelsen 2011 [42] | Scandinavia | Scandinavia | 8 VIM-Kpn isolates | 2005-2008 | diverse | 1 | - | - | - | VIM1 (1) | NS |

NP: not specified, Kpn: *K. pneumoniae*

**Table S4: Distribution of *bla*$_{OXA-48}$ plasmids by ST of blood CPK isolates**

| rep_type(s) | IncL/M* (pHB1) | IncL/M (pHB2) | IncL/M (+/- IncR,IncHI1B) (pHB8) | IncL/M | IncL/M | IncL/M | IncL/M,IncR | IncL/M |
|---|---|---|---|---|---|---|---|---|
| **Rep_type_accession(s)** | **JN626286** | **JN626286** | **JN626286 (+/- 000204_CP008701_00115, JN420336)** | **JN626286** | **JN626286** | **JN626286** | **JN626286, 000204_CP008701_00115** | **U27345** |
| **Mash_nearest_ neighbor** | **CP018717** | **CP019078** | **KX523901** | **KY215945** | **KY213890** | **KX636096** | **LN864819** | **KP025948** |
| **Mash distance mean (SD)** | 0,0023 (0.003) | 0.0031 (0.0027) | 0.0064 (0.0055) | 0.012 (0.002) | 0.01355 | 0.0007 | 0.0113 | 0.0015 |
| **Primary/ Secondary_cluster_id** | AA002 /AH539 | AA002 /AH529 | AA002 /AH539 | AA002 /AH539 | AA002 /AH539 | AA002 /AH539 | AA002 /AH539 | AA002 /AH529 |
| **Relaxase_type** | MOBP | MOBP | MOBP | MOBP | MOBP | MOBP | MOBP | MOBP |
| **MPF_type** | MPF_I | MPF_I | MPF_I | MPF_I | MPF_I | MPF_I | MPF_I | MPF_I |
| **Predicted_mobility** | conjugative | conjugative | conjugative | conjugative | conjugative | conjugative | conjugative | conjugative |
| **Plasmid size pb Mean (SD)** | 65917.1 (9438.8) | 71823.3 (8111.1) | 79955.6 (17049.9) | 44511 (5545.1) | 41882 | 59211 | 92377 | 71108 |
| **Resistance genes in plasmids** | - | CTX-M-14 | - | - | - | - | - | - |
| **Years of identification** | **2011-2022** | **2014-2022** | 2013-2020 | 2018 | 2022 | 2013 | 2017 | 2013 |
| **ST (transconjugant number)** | | | | | | | | |
| **ST147** | 2 (1) | 3 (1) | | 1 (0) | | | | |
| **ST101** | 13 (12) | 4 (3) | 1 (1) | | | 1 (1) | 1 (1) | |
| **ST13** | 2 (2) | | | | | | | |
| **ST15** | 1 (1) | | 2 (2) | 1 (1) | | | | |

| | | | | | | | | |
|---|---|---|---|---|---|---|---|---|
| **ST383** | 2 (2) | 8 (1) | | | 1 (0) | | | |
| **ST11** | 1 (1) | | | | | | | |
| **ST395** | | | 1 (1) | | | | | |
| **ST307** | 1 (1) | | | | | | | |
| **ST45** | | | 1 (1) | | | | | |
| **ST2086** | 1 (1) | | | | | | | |
| **ST2096** | 1 (1) | | | | | | | |
| **ST23** | 1 (0) | | | | | | | |
| **ST323** | 1 (1) | | | | | | | |
| **ST514** | | | | | | | | 1 (1) |
| **ST987** | 1 (1) | | | | | | | |
| **Total** | **27 (24)** | **15 (5)** | **5 (5)** | **2 (1)** | **1 (0)** | **1 (1)** | **1 (1)** | **1 (1)** |

*: 3 IncL/M plasmids were associated with IncR replicon: 000204_CP008701_00115

The table summarizes the features of the study plasmids identified by MOB-suite, which groups plasmids into primary and secondary clusters with pairwise Mash distances. Values obtained from MOB-suite indicate the replicon types (rep_type(s)), the Mash nearest neighbor, and the corresponding mash distance. MOB-suite classifies the Mobility of plasmids based on the presence of relaxase (mobilizable) and/or MPF proteins (conjugative) or absence of both (non-mobilizable)

#### Tables S5: Distribution of *bla*$_{\text{NDM-1.}}$ plasmids by ST of blood CPK isolates

| **Plasmids** | **IncFIB(pQil); IncFII(K)** | | | **IncFIB(pQil)** | **IncFIB(K); IncFII(K)** | **IncC** | | **IND** |
|---|---|---|---|---|---|---|---|---|
| **Rep_type(s)** | **IncFIB(pQil); IncFII(K:2) (pHB3)** | **IncFIB(pQil); IncFII(K:2)** | **IncFIB(pQil); IncFII(K:2) (pHB6)** | **IncFIB(pQil) (pHB5)** | **IncFIB(K); IncFII(K:1) (pHB7)** | **IncC:3** | **IncC:3** | IND |
| **Mash_nearest_neighbor** | **MW363916** | **MW363914** | **CP009115** | **CP014757** | **AP012055** | **MG450360** | **CP043190** | |
| **Mash distance mean (SD)** | 0.00208 (0.0014) | 0.0069 (0.0015) | 0.00266 (0.00198) | 0.0077 (0.006) | 0.01039 | 0.00528 | 0.01104 | |
| **Primary/secondary_cluster_ID** | AA018/AH562 | AA018/AH562 | AA018/AH560 | AA019/AH565 | AA274/AI071 | AA860/AJ275 | AA860/AJ275 | |
| **Relaxase_type** | MOBF,MOBF | MOBF,MOBF | MOBF,MOBF | - | MOBF,MOBF,MOBP | MOBH,MOBH | MOBH,MOBH | |
| **MPF_type** | MPF_F | MPF_F | MPF_F | - | MPF_F | MPF_F | MPF_F | |
| **Predicted_mobility** | conjugative | conjugative | conjugative | non-mobilizable | conjugative | conjugative | conjugative | |
| **Plasmid size pb Mean (SD)** | 112717 (8315.2) | 82143.5 (4891.1) | 113096.2 (8720.7) | 40651.2 (10936.2) | 298673.5 (2935.2) | 202974 | 54109 | |
| **Years of identification** | 2014-2022 | 2014-2015 | 2014-2017 | 2019-2022 | 2016-2017 | 2018 | 2021 | 2015-2016 |
| **Resistance genes in plasmids** | NDM1-M15-AAC6-AAC3-OXA1-TEM1-CATB-QNRB1 | NDM1-QNRB1 | NDM1-M15-TEM-APH3VI-OXA9-AAD-QNRS1 | NDM1-M15-QNRS1-AAR3-CATB-AAC6-APH3 | NDM1-M15-OXA1-RMTF-CATB-ARR-SUL1-DFR-AAC3-AAD-AAC6 | NDM1-CMY-AAC-AAD-ARM-APH3-APH6-OX1-CATB-SUL1-SUL2-TET-QNRA6-AAR | | |
| **ST (transconjugant nb)** | | | | | | | | |
| **ST147** | **6 (5)** | | | **5 (0)** | | | | |
| **ST101** | **4 (2)** | | | | | **1 (1)** | | |
| **ST13** | **2 (2)** | | | | | | | |
| **ST15** | **1 (1)** | | **1 (1)** | | | | | |
| **ST152** | **1 (1)** | | | | | | | |
| **ST219** | **1 (1)** | | | | | | | |
| **ST661** | **1 (1)** | | | | | | | |

| | | | | | | | | |
|---|---|---|---|---|---|---|---|---|
| **ST11** | | | | | **2 (0)** | | | **1 (1)** |
| **ST395** | | | **3 (0)** | | | | | |
| **ST307** | | **2 (1)** | | | | | | **1 (1)** |
| **ST45** | | | **1 (1)** | | | | | **1 (0)** |
| **ST1418** | | | | | | | **1 (0)** | |
| **Total** | **16 (13)** | **2 (1)** | **5 (2)** | **5 (0)** | **2 (0)** | **1 (1)** | **1(0)** | **3 (2)** |

The table summarizes the features of the study plasmids identified by MOB-suite, which groups plasmids into primary and secondary clusters with pairwise Mash distances. Values obtained from MOB-suite indicate the replicon types (rep_type(s)), the Mash nearest neighbor, and the corresponding mash distance. MOB-suite classifies the Mobility of plasmids based on the presence of relaxase (mobilizable) and/or MPF proteins (conjugative) or absence of both (non-mobilizable).

**Table S6 (Supplementary Data 2, p1): Sequence records of the NCBI Reference Plasmids used in the study.**

**Table S7 (Supplementary Data 2, p2): Alignment summaries of the study plasmids to the reference plasmids**

Results showed as percentages of coverage and identity to the reference plasmids.

**Table S8 (Supplementary Data 2, p3): Sequence records of the NCBI *Klebsiella pneumoniae* isolates used in the study.**
