## Supplementary_Data_1 for "Leveraging the global genomic epidemiology of carbapenemase-producing *Klebsiella pneumoniae* to inform infection prevention in Tunisian hospitals"

### Supplementary Data 1: Additional phylogenetic dendrograms

Dendrograms in this appendix were created using RAxML-ng on a wgMLST derived SNP matrix containing positions with substitutions. Dendrogram members were selected by those with the most perfect wgMLST matches. The initial starting pool of genomes numbered 80,252.

Isolates submitted as part of this study have their Isolate and MLST colored dark red, and the isolate font is enlarged.

Countries are color coded by continent.

Gene content is blue when present, white when absent. KPC and VIM include all alleles, and the identities of all genes are available through the interactive trees at the link below.

Plasmid presence is a gradient from white (0% present) to blue (100% present). These correspond to the amount of the plasmid covered by unique blast hits longer than 500 b and with an e-value < 0.001.

Nodes and branches are colored based on their transfer bootstrap expectation, with red = 0 and green = 1.

Interactive versions of these trees and SNP difference matrices are available at:
https://itol.embl.de/shared/pUTb8g8Zw3eX

NCBI Pathogen Isolates database access date: February 13th, 2025.

The table of contents lists the MLSTs, SNP clusters, and important stats for each of the dendrograms.

### Table of contents

| Tree # | Tree | Seed | # HBH Isolates | # Non-HBH Isolates | MLSTs | SNP Clusters | Matrix length (b) |
|---|---|---|---|---|---|---|---|
| 1 | GCA_043858345 | GCA_043858345 | 4 | 100 | 15, 2147 | PDS000036279.2, PDS000041726.21, PDS000045329.17, PDS000046916.2, PDS000060613.1, PDS000066127.3, PDS000075194.4, PDS000100772.1, PDS000104501.4, PDS000106381.4, PDS000156654.1, PDS000179049.7, PDS000184430.3, PDS000199428.1, PDS000214445.1 | 3379 |
| 2 | GCA_043859295 | GCA_043859295 | 1 | 100 | 323 | PDS000007156.3, PDS000017083.21, PDS000041776.5, PDS000041777.3, PDS000064191.2, PDS000079407.1, PDS000093251.1, PDS000096681.1, PDS000103183.2, PDS000106604.3, PDS000107667.1, PDS000121548.1, PDS000170482.1, PDS000216815.1, PDS000216816.1 | 2931 |
| 3 | GCA_043859305 | GCA_043859305 | 1 | 100 | 14 | PDS000048890.1, PDS000063451.2, PDS000073429.1, PDS000074446.1, PDS000085075.2, PDS000098764.2, PDS000102332.1, PDS000104978.1, PDS000139927.2, PDS000146621.4, PDS000149382.2, PDS000164521.1, PDS000169108.4, PDS000172308.1, PDS000192187.1, PDS000194032.1, PDS000198392.1, PDS000198406.1, PDS000213373.1, PDS000215014.1, PDS000215017.1 | 7842 |
| 4 | GCA_043859365 | GCA_043859365 | 4 | 100 | 129, 395, 1082, 1625, 1823 | PDS000036210.15, PDS000040454.8, PDS000041774.4, PDS000044310.6, PDS000045400.2, PDS000053786.7, PDS000072337.1, PDS000076869.2, PDS000088522.1, PDS000095854.2, PDS000101907.1, PDS000114341.1, PDS000122140.2, PDS000149535.1, PDS000171747.1, PDS000171750.1, PDS000185317.1, PDS000190144.1, PDS000197597.1, PDS000201458.1 | 45842 |
| 5 | GCA_043859495 | GCA_043859495 | 2 | 100 | 13 | PDS000041826.3, PDS000041857.2, PDS000053017.24, PDS000056122.9, PDS000098778.2, PDS000104514.1, PDS000110898.1, PDS000121641.1, PDS000140054.5, PDS000155185.4, PDS000185322.1, PDS000194059.1, PDS000197052.1, PDS000217867.1 | 5125 |
| 6 | GCA_043859615 | GCA_043859615 | 13 | 100 | 101, 2502 | PDS000045320.1, PDS000045324.14, PDS000045328.1, PDS000054005.14, PDS000060649.2, PDS000080193.2, PDS000104479.25, PDS000166495.5, PDS000199426.1 | 1135 |
| 7 | GCA_043859815 | GCA_043859815 | 1 | 100 | 23, 57 | PDS000045398.1, PDS000060603.1, PDS000095240.1, PDS000100774.1, PDS000111295.2, PDS000111758.3, PDS000144603.1, PDS000160830.3, PDS000164559.2, PDS000168933.1, PDS000168937.2, PDS000183478.1, PDS000183493.1, PDS000183496.1, PDS000187849.1, PDS000192486.2, PDS000199499.1 | 3930 |
| 8 | GCA_043859855 | GCA_043859855 | 1 | 101 | 44, 107, 219, 305, 514, 2449, 3636, 5756 | PDS000036289.4, PDS000074438.1, PDS000074448.1, PDS000074449.1, PDS000112610.2, PDS000112630.1, PDS000114410.2, PDS000121675.1, PDS000148766.7, PDS000150182.3, PDS000150194.3, PDS000157999.11, PDS000170612.1, PDS000176509.1, PDS000181291.1, PDS000198446.1, PDS000215654.1 | 54969 |
| 9 | GCA_043859995 | GCA_043859995 | 1 | 101 | 5, 504, 540, 1013, 1087, 2086, 3319, 4510, 6245 | PDS000036226.2, PDS000083576.2, PDS000090692.1, PDS000108803.2, PDS000112629.2, PDS000123677.1, PDS000130868.1, PDS000140723.2, PDS000145396.1, PDS000165203.1, PDS000170846.1, PDS000170904.1, PDS000175697.1 | 36809 |
| 10 | GCA_043860015 | GCA_043860015 | 1 | 100 | 147 | PDS000006578.119, PDS000006642.4, PDS000009779.14, PDS000036185.10, PDS000040466.3, PDS000041784.1, PDS000052089.184, PDS000052092.1, PDS000060612.1, PDS000067059.3, PDS000076259.2, PDS000077015.34, PDS000083378.2, PDS000084451.3, PDS000096678.1, PDS000099727.3, PDS000104546.2, PDS000111778.2, PDS000129321.8, PDS000130855.1, PDS000132251.1, PDS000136742.4, PDS000156049.11, PDS000165990.1, PDS000187661.2 | 4189 |
| 11 | GCA_043860075 | GCA_043860075 | 1 | 103 | 86, 3509, 5559 | PDS000042808.1, PDS000074639.1, PDS000097499.1, PDS000100761.1, PDS000124950.1, PDS000168656.1, PDS000182304.3, PDS000183471.1, PDS000183505.1, PDS000197439.1, PDS000214453.1 | 5774 |
| 12 | GCA_043860195 | GCA_043860195 | 1 | 103 | 163, 485, 496, 815, 914, 987, 2719, 3649, 4064, 6114 | PDS000038919.5, PDS000053037.8, PDS000054678.2, PDS000059976.3, PDS000071977.4, PDS000080493.2, PDS000085062.4, PDS000105717.1, PDS000107774.2, PDS000112639.1, PDS000160856.1, PDS000161308.3, PDS000175698.1, PDS000178659.1, PDS000183523.1, PDS000183592.1, PDS000184186.2, PDS000197593.1, PDS000199202.1 | 40282 |

### Table of contents, contniued

| Tree # | Tree | Seed | # HBH Isolates | # Non-HBH Isolates | MLSTs | SNP Clusters | Matrix length (b) |
|---|---|---|---|---|---|---|---|
| 13 | GCA_043861645 | GCA_043861645 | 3 | 100 | 11, 1640 | PDS000045343.1, PDS000045344.10, PDS000045350.2, PDS000046910.86, PDS000046967.1, PDS000060651.29, PDS000079304.4, PDS000102670.3, PDS000103169.12, PDS000112591.1, PDS000138389.1, PDS000146165.3, PDS000148349.1, PDS000149983.1, PDS000165202.2, PDS000173787.1, PDS000185312.1, PDS000216219.1 | 3372 |
| 14 | GCA_043863025 | GCA_043863025 | 4 | 101 | 307 | PDS000038607.20, PDS000045377.17, PDS000045391.1, PDS000045393.5, PDS000051246.2, PDS000054607.22, PDS000056158.28, PDS000072661.1, PDS000073377.3, PDS000074878.9, PDS000075200.1, PDS000082283.3, PDS000104496.1, PDS000176502.1, PDS000189621.16, PDS000199421.1, PDS000199423.1, PDS000201306.1, PDS000201459.1, PDS000213377.1 | 2897 |
| 15 | GCA_043863145 | GCA_043863145 | 2 | 100 | 45, 3031 | PDS000006905.10, PDS000007035.4, PDS000008541.10, PDS000052944.18, PDS000067092.3, PDS000070441.1, PDS000074716.5, PDS000074859.1, PDS000078891.2, PDS000100089.1, PDS000100594.2, PDS000104481.1, PDS000104562.1, PDS000105716.1, PDS000106393.3, PDS000106675.2, PDS000109086.2, PDS000146617.5, PDS000162236.1, PDS000175704.1, PDS000178660.3, PDS000179055.2, PDS000192605.1, PDS000198705.1, PDS000206449.1, PDS000212921.2, PDS000214277.1, PDS000215653.1 | 5431 |
| 16 | GCA_043863165 | GCA_043863165 | 3 | 100 | 395 | PDS000036210.15, PDS000040454.8, PDS000041774.4, PDS000044310.6, PDS000045400.2, PDS000053740.1, PDS000053786.7, PDS000072337.1, PDS000077831.2, PDS000095854.2, PDS000112540.1, PDS000114341.1, PDS000156671.1, PDS000190144.1 | 3892 |
| 17 | GCA_043863205 | GCA_043863205 | 2 | 101 | 218, 5939, 5943 | PDS000074977.2, PDS000076457.1, PDS000079882.2, PDS000097290.2, PDS000100586.2, PDS000117560.1, PDS000183511.1, PDS000183580.1, PDS000190124.1, PDS000199420.1 | 5471 |
| 18 | GCA_043863245 | GCA_043863245 | 1 | 101 | 152, 4513 | PDS000036263.5, PDS000044881.3, PDS000071072.5, PDS000074978.1, PDS000075204.9, PDS000076193.10, PDS000088478.25, PDS000096689.1, PDS000100167.3, PDS000102786.1, PDS000106014.1, PDS000110342.1, PDS000170201.1, PDS000185318.1, PDS000186854.1, PDS000197056.1, PDS000197588.1, PDS000198509.1, PDS000201460.1, PDS000213526.2 | 11945 |
| 19 | GCA_043863305 | GCA_043863305 | 1 | 100 | 661, 3409, 3453, 3655 | PDS000041757.1, PDS000052192.6, PDS000053783.5, PDS000090686.2, PDS000093436.1, PDS000108371.1, PDS000142414.5, PDS000148773.2, PDS000160820.1, PDS000169090.1, PDS000170208.1, PDS000172294.1, PDS000185324.1, PDS000197609.1, PDS000198414.1, PDS000198827.1, PDS000203214.1 | 22027 |
| 20 | GCA_043863325 | GCA_043863325 | 3 | 100 | 15 | PDS000036279.2, PDS000041726.21, PDS000045329.17, PDS000045331.1, PDS000046916.2, PDS000053782.4, PDS000062513.1, PDS000066127.3, PDS000072336.1, PDS000075194.4, PDS000083760.2, PDS000104501.4, PDS000106761.1, PDS000115079.3, PDS000124952.2, PDS000170844.3, PDS000179049.7, PDS000186880.1, PDS000199428.1, PDS000214445.1 | 4301 |
| 21 | GCA_043863345 | GCA_043863345 | 1 | 100 | 11, 3666 | PDS000005579.7, PDS000013916.2, PDS000036336.53, PDS000045420.7, PDS000045421.52, PDS000065468.2, PDS000072335.6, PDS000077811.10, PDS000084821.3, PDS000092775.2, PDS000103182.2, PDS000113930.22, PDS000140731.18, PDS000156641.12 | 3532 |
| 22 | GCA_043863385 | GCA_043863385 | 2 | 100 | 11 | PDS000046910.86, PDS000060651.29, PDS000154385.11, PDS000165202.2, PDS000185312.1 | 2249 |
| 23 | GCA_043863405 | GCA_043863405 | 3 | 100 | 13 | PDS000046986.1, PDS000053017.24, PDS000056122.9, PDS000098778.2, PDS000157997.9, PDS000161927.6, PDS000173925.11, PDS000194059.1, PDS000197052.1, PDS000199430.1, PDS000211070.1 | 4337 |
| 24 | GCA_043863445 | GCA_043863445 | 1 | 101 | 219 | PDS000012112.164, PDS000179977.1, PDS000217221.1 | 3691 |
| 25 | GCA_043863535 | GCA_043863535 | 1 | 100 | 147 | PDS000006578.119, PDS000006642.4, PDS000009779.14, PDS000036182.26, PDS000036186.4, PDS000036295.1, PDS000040359.4, PDS000045262.1, PDS000052092.1, PDS000053519.1, PDS000065404.6, PDS000074889.1, PDS000075223.4, PDS000080300.2, PDS000083378.2, PDS000084451.3, PDS000097792.5, PDS000099727.3, PDS000100083.5, PDS000100389.1, PDS000104526.1, PDS000106769.6, PDS000130855.1, PDS000132251.1, PDS000156049.11, PDS000157007.2, PDS000164093.1, PDS000165990.1, PDS000197072.1 | 4594 |
| 26 | GCA_043904805 | GCA_043904805 | 4 | 100 | 15, 2147 | PDS000041726.21, PDS000045329.17, PDS000046916.2, PDS000060613.1, PDS000097194.4, PDS000100772.1, PDS000106381.4, PDS000124938.1, PDS000156654.1, PDS000179049.7, PDS000184430.3, PDS000188634.2, PDS000199428.1, PDS000214445.1 | 3822 |

### Table of contents, continued

| Tree # | Tree | Seed | # HBH Isolates | # Non-HBH Isolates | MLSTs | SNP Clusters | Matrix length (b) |
|---|---|---|---|---|---|---|---|
| 27 | GCA_043904845 | GCA_043904845 | 11 | 100 | 383, 6118 | PDS000060640.1, PDS000061038.3, PDS000070357.5, PDS000085592.11, PDS000098809.4, PDS000106324.3, PDS000106730.2, PDS000108392.5, PDS000133910.8, PDS000171211.1, PDS000175841.1, PDS000179530.2, PDS000180184.1, PDS000192050.8, PDS000202475.1, PDS000217883.1 | 2177 |
| 28 | GCA_043904865 | GCA_043904865 | 1 | 100 | 340 | PDS000017086.10, PDS000036321.13, PDS000083373.14, PDS000130841.1, PDS000130869.1 | 2174 |
| 29 | GCA_043904885 | GCA_043904885 | 6 | 101 | 147 | PDS000009779.14, PDS000026421.12, PDS000045135.24, PDS000045262.1, PDS000071979.8, PDS000080114.2, PDS000092779.3, PDS000104516.1 | 1031 |
| 30 | GCA_043905265 | GCA_043905265 | 1 | 100 | 2096 | PDS000060581.66 | 667 |
| 31 | GCA_043905305 | GCA_043905305 | 6 | 101 | 383 | PDS000007455.4, PDS000060640.1, PDS000070357.5, PDS000085592.11, PDS000106324.3, PDS000108392.5, PDS000133910.8, PDS000176495.1, PDS000199419.2, PDS000202475.1 | 1735 |
| 32 | GCA_043905425 | GCA_043905425 | 1 | 100 | 147 | PDS000091501.176, PDS000201182.1 | 226 |
| 33 | GCA_043905485 | GCA_043905485 | 12 | 100 | 101, 2502, 3367 | PDS000041735.6, PDS000045311.30, PDS000045312.1, PDS000045320.1, PDS000055633.53, PDS000080305.6, PDS000088521.1, PDS000188993.3, PDS000199424.2, PDS000199425.1 | 2177 |
| 34 | GCA_043905525 | GCA_043905525 | 1 | 101 | 13 | PDS000053017.24, PDS000056122.9, PDS000098778.2, PDS000173925.11, PDS000194059.1, PDS000197052.1, PDS000211070.1 | 2316 |
| 35 | GCA_043905545 | GCA_043905545 | 1 | 101 | 29, 714, 5832 | PDS000070366.1, PDS000101853.1, PDS000108354.1, PDS000122861.1, PDS000122913.1, PDS000133597.1, PDS000143149.1, PDS000143192.1, PDS000156350.1, PDS000169967.2, PDS000173086.1, PDS000182318.2, PDS000183535.1, PDS000183538.1, PDS000183548.1, PDS000188010.2, PDS000188027.1, PDS000198821.1, PDS000201454.1, PDS000214440.1 | 10581 |
| 36 | GCA_043905565 | GCA_043905565 | 1 | 100 | 45, 1418, 2954, 3098 | PDS000006905.10, PDS000052944.18, PDS000054827.4, PDS000060037.37, PDS000074703.6, PDS000074716.5, PDS000075208.5, PDS000093208.1, PDS000100594.2, PDS000101897.1, PDS000105716.1, PDS000106393.3, PDS000106675.2, PDS000127598.3, PDS000178660.3, PDS000179055.2, PDS000190165.1, PDS000198705.1, PDS000208562.1, PDS000212921.2, PDS000213518.2, PDS000214277.1 | 12155 |
| 37 | GCA_043905585 | GCA_043905585 | 12 | 100 | 101, 2502 | PDS000045324.14, PDS000045328.1, PDS000054005.14, PDS000060649.2, PDS000080193.2, PDS000104479.25, PDS000166495.5, PDS000199426.1 | 1107 |
| 38 | GCA_043905605 | GCA_043905605 | 11 | 101 | 383, 6118 | PDS000060640.1, PDS000061038.3, PDS000070357.5, PDS000085592.11, PDS000098809.4, PDS000106324.3, PDS000106730.2, PDS000108392.5, PDS000125001.14, PDS000133910.8, PDS000171211.1, PDS000175841.1, PDS000179530.2, PDS000180184.1, PDS000192050.8, PDS000202475.1, PDS000217883.1 | 2222 |
| 39 | GCA_043905625 | GCA_043905625 | 1 | 100 | 147 | PDS000091501.176, PDS000199417.1, PDS000210545.1 | 252 |
| 40 | GCA_043905645 | GCA_043905645 | 5 | 100 | 147, 4843 | PDS000052089.184, PDS000056180.7, PDS000077015.34, PDS000090167.14, PDS000103185.4, PDS000104520.1, PDS000106697.6, PDS000129332.6, PDS000136743.8, PDS000156049.11, PDS000199427.1, PDS000201444.2, PDS000205531.1 | 1758 |
| 41 | GCA_043905665 | GCA_043905665 | 1 | 100 | 147 | PDS000091501.176 | 252 |
| 42 | MLST383_top400 .raxml.support | GCA_043905605 | 15 | 385 | 42, 110, 231, 376, 383, 413, 750, 1190, 1588, 2947, 4295, 4853, 5079, 5410, 5875, 6118, 6125 | PDS000007455.4, PDS000041770.3, PDS000050643.3, PDS000052267.2, PDS000056155.1, PDS000059234.2, PDS000060640.1, PDS000061038.3, PDS000065279.1, PDS000065282.1, PDS000070357.5, PDS000071539.1, PDS000072925.3, PDS000074875.1, PDS000074905.1, PDS000074991.1, PDS000077168.1, PDS000077820.1, PDS000078846.2, PDS000083580.1, PDS000085591.7, PDS000085592.11, PDS000093234.4, PDS000098809.4, PDS000101835.1, PDS000104969.1, PDS000106030.1, PDS000106324.3, PDS000106730.2, PDS000106886.14, PDS000107419.2, PDS000108392.5, PDS000125001.14, PDS000133910.8, PDS000141863.2, PDS000143218.1, PDS000149358.4, PDS000161102.14, PDS000166706.2, PDS000171211.1, PDS000171212.1, PDS000172292.1, PDS000172301.1, PDS000175841.1, PDS000176495.1, PDS000179530.2, PDS000180184.1, PDS000186585.1, PDS000188960.4, PDS000192050.8, PDS000192602.1, PDS000194043.1, PDS000194063.1, PDS000198706.1, PDS000198822.1, PDS000199419.2, PDS000202475.1, PDS000205525.1, PDS000210726.1, PDS000217883.1 | 89712 |
| 43 | ST383_ST376 | GCA_043905605 | 15 | 249 | 376, 383, 4853, 5410, 6118 | PDS000007455.4, PDS000050643.3, PDS000060640.1, PDS000061038.3, PDS000070357.5, PDS000077168.1, PDS000085591.7, PDS000085592.11, PDS000098809.4, PDS000106324.3, PDS000106730.2, PDS000108392.5, PDS000125001.14, PDS000133910.8, PDS000141863.2, PDS000161102.14, PDS000171211.1, PDS000171212.1, PDS000175841.1, PDS000176495.1, PDS000179530.2, PDS000180184.1, PDS000188960.4, PDS000192050.8, PDS000192602.1, PDS000199419.2, PDS000202475.1, PDS000205525.1, PDS000217883.1 | 9974 |

### Tree 1: GCA_043858345

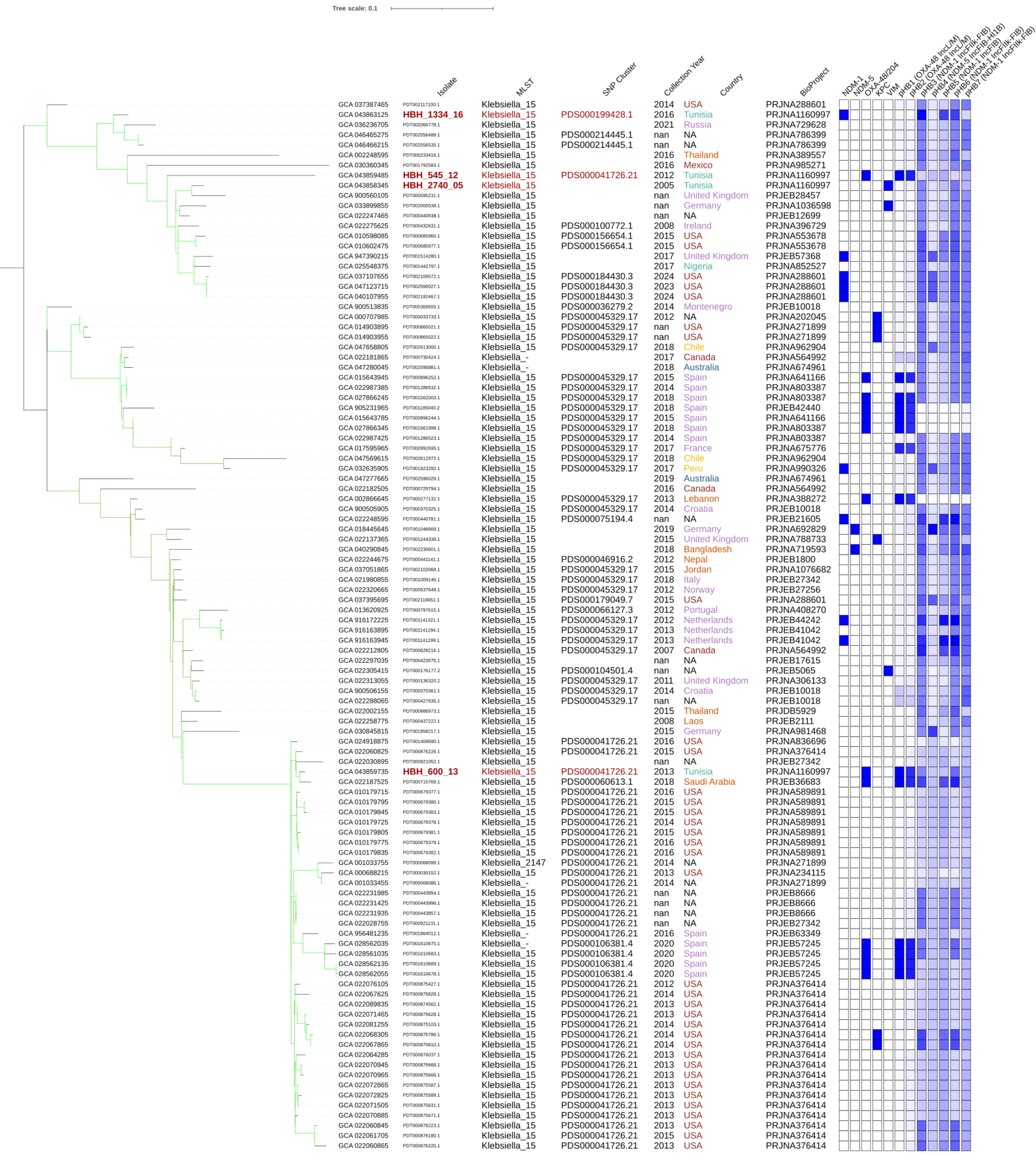

### Tree 2: GCA_043859295

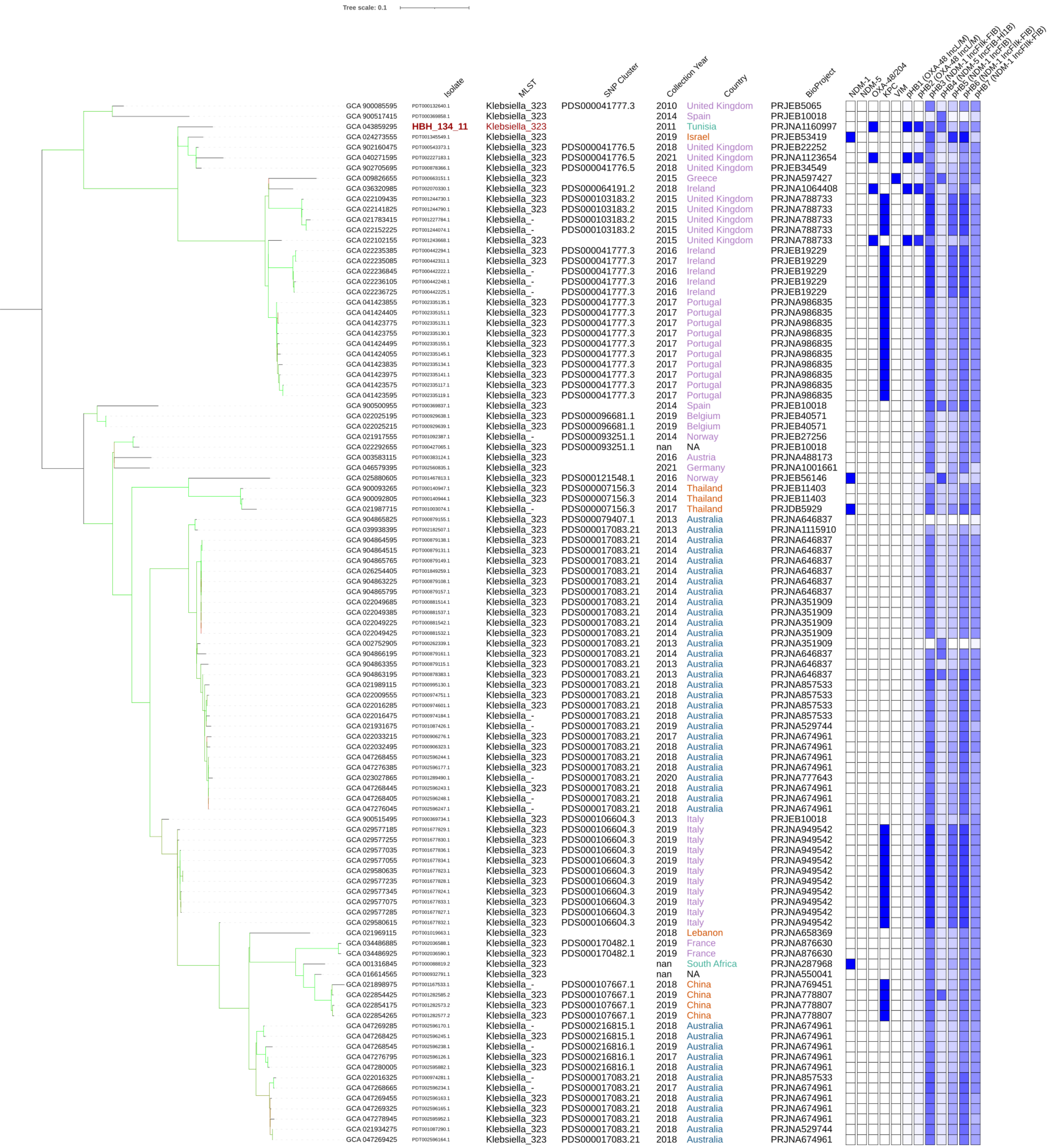

### Tree 3: GCA_043859305

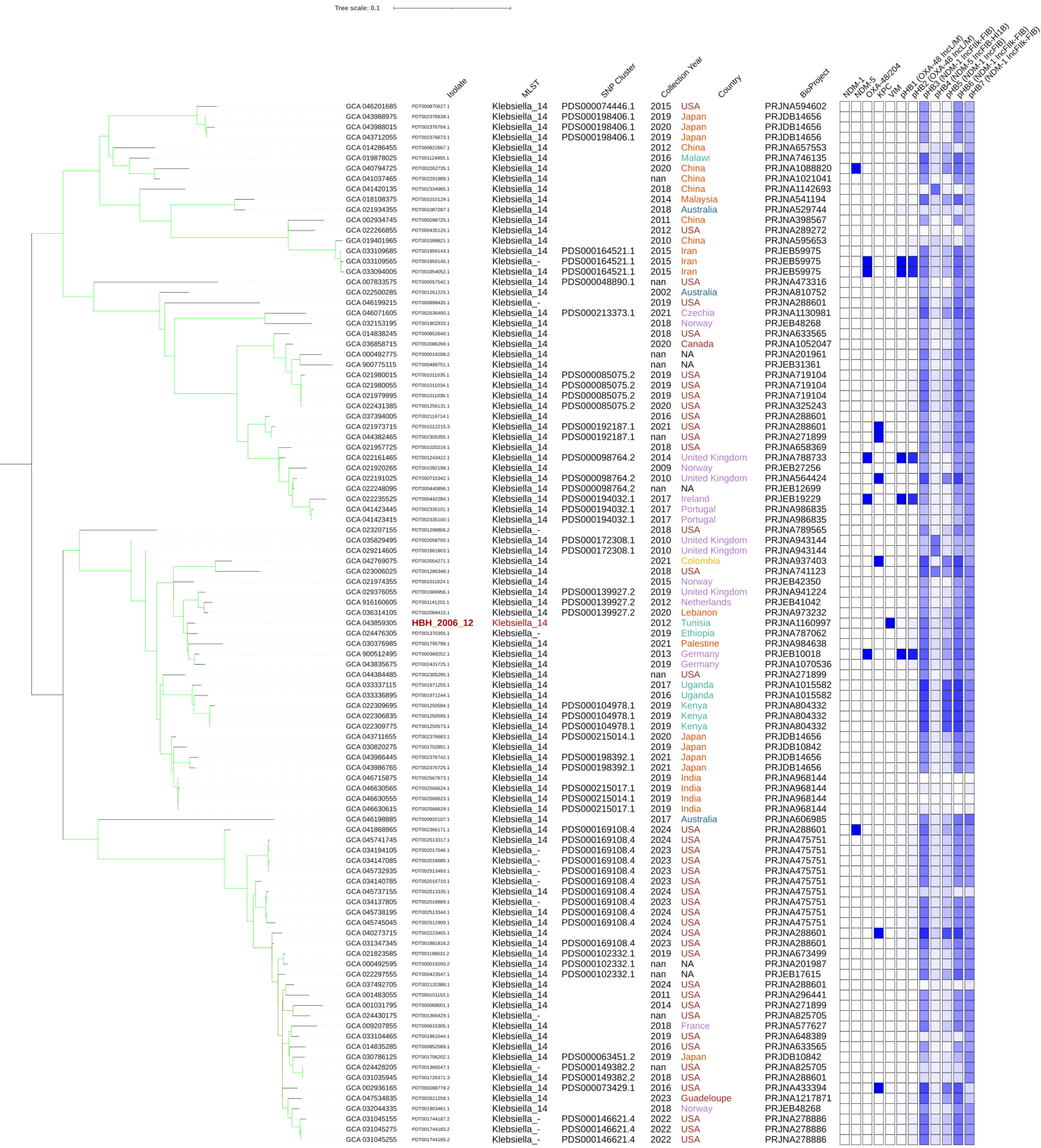

### Tree 4: GCA_043859365

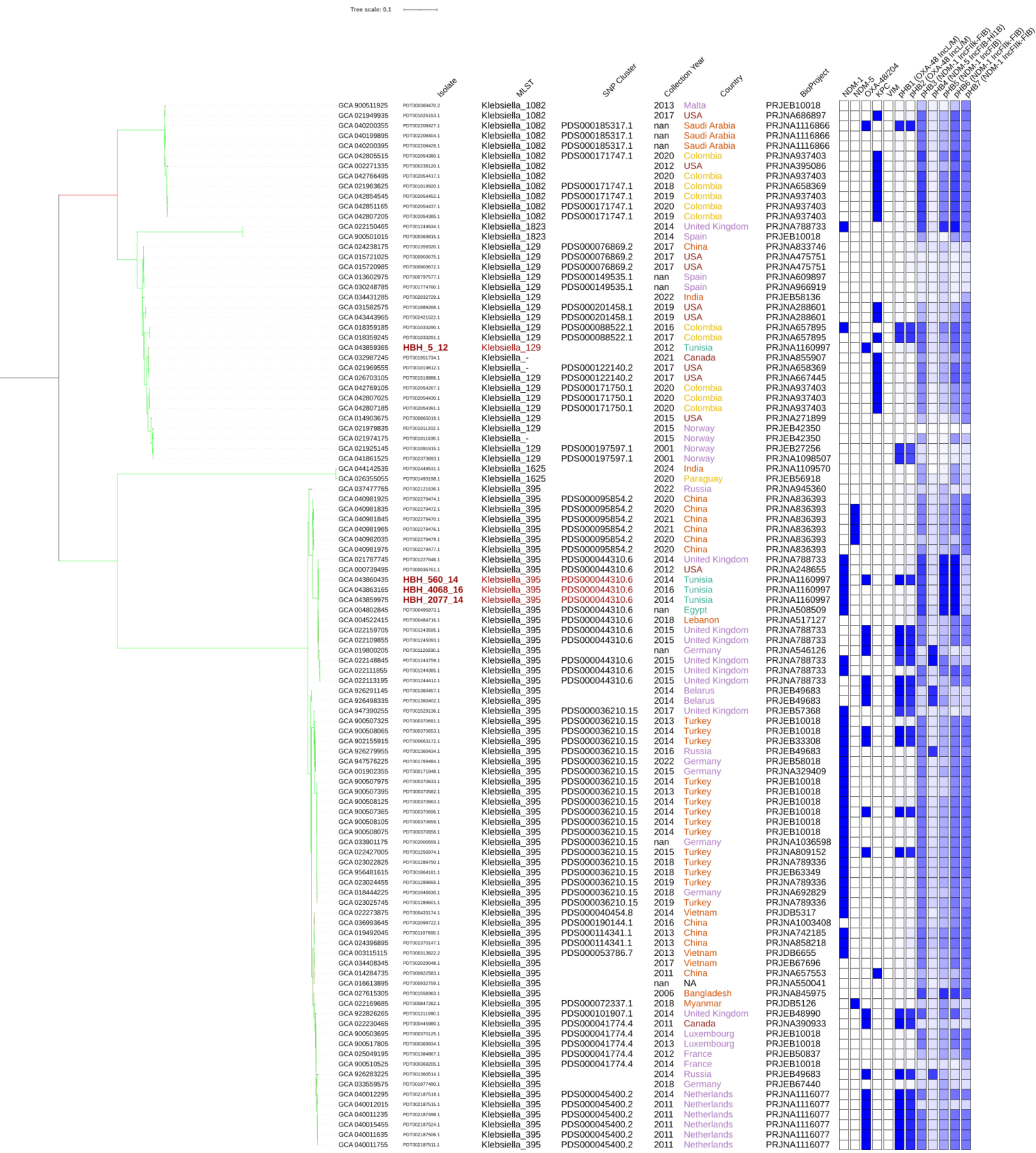


| | Isolate | MLST | SNP Cluster | Collection Year | Country | BioProject |
|---|---|---|---|---|---|---|
| GCA 900511925 | PDT000369470.2 | Klebsiella_1082 | | 2013 | Malta | PRJEB10018 |
| GCA 021949935 | PDT001025153.1 | Klebsiella_1082 | | 2017 | USA | PRJNA686897 |
| GCA 040200355 | PDT002208427.1 | Klebsiella_1082 | PDS000185317.1 | nan | Saudi Arabia | PRJNA1116866 |
| GCA 040199895 | PDT002208404.1 | Klebsiella_1082 | PDS000185317.1 | nan | Saudi Arabia | PRJNA1116866 |
| GCA 040200395 | PDT002208429.1 | Klebsiella_1082 | PDS000185317.1 | nan | Saudi Arabia | PRJNA1116866 |
| GCA 042805515 | PDT002054380.1 | Klebsiella_1082 | PDS000171747.1 | 2020 | Colombia | PRJNA937403 |
| GCA 002271335 | PDT000238120.1 | Klebsiella_1082 | | 2012 | USA | PRJNA395086 |
| GCA 042766495 | PDT002054417.1 | Klebsiella_1082 | | 2020 | Colombia | PRJNA937403 |
| GCA 021963625 | PDT001019920.1 | Klebsiella_1082 | PDS000171747.1 | 2018 | Colombia | PRJNA658369 |
| GCA 042854545 | PDT002054452.1 | Klebsiella_1082 | PDS000171747.1 | 2019 | Colombia | PRJNA937403 |
| GCA 042851165 | PDT002054437.1 | Klebsiella_1082 | PDS000171747.1 | 2020 | Colombia | PRJNA937403 |
| GCA 042807205 | PDT002054385.1 | Klebsiella_1082 | PDS000171747.1 | 2019 | Colombia | PRJNA937403 |
| GCA 022150465 | PDT001244634.1 | Klebsiella_1823 | | 2014 | United Kingdom | PRJNA788733 |
| GCA 900501015 | PDT000369815.1 | Klebsiella_1823 | | 2014 | Spain | PRJEB10018 |
| GCA 024238175 | PDT001359320.1 | Klebsiella_129 | PDS000076869.2 | 2017 | China | PRJNA833746 |
| GCA 015721025 | PDT000903675.1 | Klebsiella_129 | PDS000076869.2 | 2017 | USA | PRJNA475751 |
| GCA 015720985 | PDT000903672.1 | Klebsiella_129 | PDS000076869.2 | 2017 | USA | PRJNA475751 |
| GCA 013602975 | PDT000797577.1 | Klebsiella_129 | PDS000149535.1 | nan | Spain | PRJNA609897 |
| GCA 030248785 | PDT001774760.1 | Klebsiella_129 | PDS000149535.1 | nan | Spain | PRJNA966919 |
| GCA 034431285 | PDT002032729.1 | Klebsiella_129 | | 2022 | India | PRJEB58136 |
| GCA 031582575 | PDT001889268.1 | Klebsiella_129 | PDS000201458.1 | 2019 | USA | PRJNA288601 |
| GCA 043443965 | PDT002421522.1 | Klebsiella_129 | PDS000201458.1 | 2019 | USA | PRJNA288601 |
| GCA 018359185 | PDT001033290.1 | Klebsiella_129 | PDS000088522.1 | 2016 | Colombia | PRJNA657895 |
| GCA 018359245 | PDT001033291.1 | Klebsiella_129 | PDS000088522.1 | 2017 | Colombia | PRJNA657895 |
| GCA 043859365 | **HBH_5_12** | Klebsiella_129 | | 2012 | Tunisia | PRJNA1160997 |
| GCA 032987245 | PDT001951734.1 | Klebsiella_- | | 2021 | Canada | PRJNA855907 |
| GCA 021969555 | PDT001019612.1 | Klebsiella_- | PDS000122140.2 | 2017 | USA | PRJNA658369 |
| GCA 026703105 | PDT001518886.1 | Klebsiella_129 | PDS000122140.2 | 2017 | USA | PRJNA667445 |
| GCA 042769105 | PDT002054267.1 | Klebsiella_129 | PDS000171750.1 | 2020 | Colombia | PRJNA937403 |
| GCA 042807025 | PDT002054430.1 | Klebsiella_129 | PDS000171750.1 | 2020 | Colombia | PRJNA937403 |
| GCA 042807185 | PDT002054391.1 | Klebsiella_129 | PDS000171750.1 | 2020 | Colombia | PRJNA937403 |
| GCA 014903675 | PDT000865019.1 | Klebsiella_129 | | 2015 | USA | PRJNA271899 |
| GCA 021979835 | PDT001011202.1 | Klebsiella_129 | | 2015 | Norway | PRJEB42350 |
| GCA 021974175 | PDT001011639.1 | Klebsiella_- | | 2015 | Norway | PRJEB42350 |
| GCA 021925145 | PDT001091915.1 | Klebsiella_129 | PDS000197597.1 | 2001 | Norway | PRJEB27256 |
| GCA 041861525 | PDT002373693.1 | Klebsiella_129 | PDS000197597.1 | 2001 | Norway | PRJNA1098507 |
| GCA 044142535 | PDT002448531.1 | Klebsiella_1625 | | 2024 | India | PRJNA1109570 |
| GCA 026355055 | PDT001493198.1 | Klebsiella_1625 | | 2020 | Paraguay | PRJEB56918 |
| GCA 037477765 | PDT002121536.1 | Klebsiella_395 | | 2022 | Russia | PRJNA945360 |
| GCA 040981925 | PDT002279474.1 | Klebsiella_395 | PDS000095854.2 | 2020 | China | PRJNA836393 |
| GCA 040981835 | PDT002279472.1 | Klebsiella_395 | PDS000095854.2 | 2020 | China | PRJNA836393 |
| GCA 040981845 | PDT002279470.1 | Klebsiella_395 | PDS000095854.2 | 2021 | China | PRJNA836393 |
| GCA 040981965 | PDT002279476.1 | Klebsiella_395 | PDS000095854.2 | 2021 | China | PRJNA836393 |
| GCA 040982035 | PDT002279479.1 | Klebsiella_395 | PDS000095854.2 | 2020 | China | PRJNA836393 |
| GCA 040981975 | PDT002279477.1 | Klebsiella_395 | PDS000095854.2 | 2020 | China | PRJNA836393 |
| GCA 021787745 | PDT001227646.1 | Klebsiella_395 | PDS000044310.6 | 2014 | United Kingdom | PRJNA788733 |
| GCA 000739495 | PDT000036761.1 | Klebsiella_395 | PDS000044310.6 | 2012 | USA | PRJNA248655 |
| GCA 043860435 | **HBH_560_14** | Klebsiella_395 | PDS000044310.6 | 2014 | Tunisia | PRJNA1160997 |
| GCA 043863165 | **HBH_4068_16** | Klebsiella_395 | PDS000044310.6 | 2016 | Tunisia | PRJNA1160997 |
| GCA 043859975 | **HBH_2077_14** | Klebsiella_395 | PDS000044310.6 | 2014 | Tunisia | PRJNA1160997 |
| GCA 004802845 | PDT000495873.1 | Klebsiella_395 | PDS000044310.6 | nan | Egypt | PRJNA508509 |
| GCA 004522415 | PDT000484716.1 | Klebsiella_395 | PDS000044310.6 | 2018 | Lebanon | PRJNA517127 |
| GCA 022159705 | PDT001243595.1 | Klebsiella_395 | PDS000044310.6 | 2015 | United Kingdom | PRJNA788733 |
| GCA 022109855 | PDT001245093.1 | Klebsiella_395 | PDS000044310.6 | 2015 | United Kingdom | PRJNA788733 |
| GCA 019800205 | PDT001120290.1 | Klebsiella_395 | | nan | Germany | PRJNA546126 |
| GCA 022148845 | PDT001244759.1 | Klebsiella_395 | PDS000044310.6 | 2015 | United Kingdom | PRJNA788733 |
| GCA 022111855 | PDT001244385.1 | Klebsiella_395 | PDS000044310.6 | 2015 | United Kingdom | PRJNA788733 |
| GCA 022113195 | PDT001244412.1 | Klebsiella_395 | PDS000044310.6 | 2015 | United Kingdom | PRJNA788733 |
| GCA 926291145 | PDT001360457.1 | Klebsiella_395 | | 2014 | Belarus | PRJEB49683 |
| GCA 926498335 | PDT001360402.1 | Klebsiella_395 | | 2014 | Belarus | PRJEB49683 |
| GCA 947390255 | PDT001529136.1 | Klebsiella_395 | PDS000036210.15 | 2017 | United Kingdom | PRJEB57368 |
| GCA 900507325 | PDT000370691.1 | Klebsiella_395 | PDS000036210.15 | 2013 | Turkey | PRJEB10018 |
| GCA 900508065 | PDT000370653.1 | Klebsiella_395 | PDS000036210.15 | 2014 | Turkey | PRJEB10018 |
| GCA 902155915 | PDT000663172.1 | Klebsiella_395 | PDS000036210.15 | 2014 | Turkey | PRJEB33308 |
| GCA 926279955 | PDT001360434.1 | Klebsiella_395 | PDS000036210.15 | 2016 | Russia | PRJEB49683 |
| GCA 947576225 | PDT001769484.1 | Klebsiella_395 | PDS000036210.15 | 2022 | Germany | PRJEB58018 |
| GCA 001902355 | PDT000171848.1 | Klebsiella_395 | PDS000036210.15 | 2015 | Germany | PRJNA329409 |
| GCA 900507975 | PDT000370633.1 | Klebsiella_395 | PDS000036210.15 | 2014 | Turkey | PRJEB10018 |
| GCA 900507395 | PDT000370582.1 | Klebsiella_395 | PDS000036210.15 | 2013 | Turkey | PRJEB10018 |
| GCA 900508125 | PDT000370663.1 | Klebsiella_395 | PDS000036210.15 | 2014 | Turkey | PRJEB10018 |
| GCA 900507365 | PDT000370696.1 | Klebsiella_395 | PDS000036210.15 | 2014 | Turkey | PRJEB10018 |
| GCA 900508105 | PDT000370659.1 | Klebsiella_395 | PDS000036210.15 | 2014 | Turkey | PRJEB10018 |
| GCA 900508075 | PDT000370656.1 | Klebsiella_395 | PDS000036210.15 | 2014 | Turkey | PRJEB10018 |
| GCA 033901175 | PDT002000559.1 | Klebsiella_395 | PDS000036210.15 | nan | Germany | PRJNA1036598 |
| GCA 022427005 | PDT001256974.1 | Klebsiella_395 | PDS000036210.15 | 2015 | Turkey | PRJNA809152 |
| GCA 023022825 | PDT001289750.1 | Klebsiella_395 | PDS000036210.15 | 2018 | Turkey | PRJNA789336 |
| GCA 956481615 | PDT001864181.1 | Klebsiella_395 | PDS000036210.15 | 2018 | Turkey | PRJEB63349 |
| GCA 023024455 | PDT001289655.1 | Klebsiella_395 | PDS000036210.15 | 2019 | Turkey | PRJNA789336 |
| GCA 018444225 | PDT001046630.1 | Klebsiella_395 | PDS000036210.15 | 2018 | Germany | PRJNA692829 |
| GCA 023025745 | PDT001289601.1 | Klebsiella_395 | PDS000036210.15 | 2019 | Turkey | PRJNA789336 |
| GCA 022273875 | PDT000433174.1 | Klebsiella_395 | PDS000040454.8 | 2014 | Vietnam | PRJDB5317 |
| GCA 036993645 | PDT002098722.1 | Klebsiella_395 | PDS000190144.1 | 2016 | China | PRJNA1003408 |
| GCA 019492045 | PDT001107669.1 | Klebsiella_395 | PDS000114341.1 | 2013 | China | PRJNA742185 |
| GCA 024396895 | PDT001370147.1 | Klebsiella_395 | PDS000114341.1 | 2013 | China | PRJNA858218 |
| GCA 003115115 | PDT000313822.2 | Klebsiella_395 | PDS000053786.7 | 2013 | Vietnam | PRJDB6655 |
| GCA 034408345 | PDT002029048.1 | Klebsiella_395 | | 2017 | Vietnam | PRJEB67696 |
| GCA 014284735 | PDT000822583.1 | Klebsiella_395 | | 2011 | China | PRJNA657553 |
| GCA 016613895 | PDT000932759.1 | Klebsiella_395 | | nan | NA | PRJNA550041 |
| GCA 027615305 | PDT001558363.1 | Klebsiella_395 | | 2006 | Bangladesh | PRJNA845975 |
| GCA 022169685 | PDT000847262.1 | Klebsiella_395 | PDS000072337.1 | 2018 | Myanmar | PRJDB5126 |
| GCA 922826265 | PDT001211080.1 | Klebsiella_395 | PDS000101907.1 | 2014 | United Kingdom | PRJEB48990 |
| GCA 022230465 | PDT000445980.1 | Klebsiella_395 | PDS000041774.4 | 2011 | Canada | PRJNA390933 |
| GCA 900503695 | PDT000370125.1 | Klebsiella_395 | PDS000041774.4 | 2014 | Luxembourg | PRJEB10018 |
| GCA 900517805 | PDT000369934.1 | Klebsiella_395 | PDS000041774.4 | 2013 | Luxembourg | PRJEB10018 |
| GCA 025049195 | PDT001384867.1 | Klebsiella_395 | PDS000041774.4 | 2012 | France | PRJEB50837 |
| GCA 900510525 | PDT000369205.1 | Klebsiella_395 | PDS000041774.4 | 2014 | France | PRJEB10018 |
| GCA 926283225 | PDT001360514.1 | Klebsiella_395 | | 2014 | Russia | PRJEB49683 |
| GCA 033559575 | PDT001977490.1 | Klebsiella_395 | | 2018 | Germany | PRJEB67440 |
| GCA 040012295 | PDT002187519.1 | Klebsiella_395 | PDS000045400.2 | 2014 | Netherlands | PRJNA1116077 |
| GCA 040012015 | PDT002187515.1 | Klebsiella_395 | PDS000045400.2 | 2011 | Netherlands | PRJNA1116077 |
| GCA 040011235 | PDT002187498.1 | Klebsiella_395 | PDS000045400.2 | 2011 | Netherlands | PRJNA1116077 |
| GCA 040015455 | PDT002187524.1 | Klebsiella_395 | PDS000045400.2 | 2011 | Netherlands | PRJNA1116077 |
| GCA 040011635 | PDT002187506.1 | Klebsiella_395 | PDS000045400.2 | 2011 | Netherlands | PRJNA1116077 |
| GCA 040011755 | PDT002187511.1 | Klebsiella_395 | PDS000045400.2 | 2011 | Netherlands | PRJNA1116077 |

### Tree 5: GCA_043859495

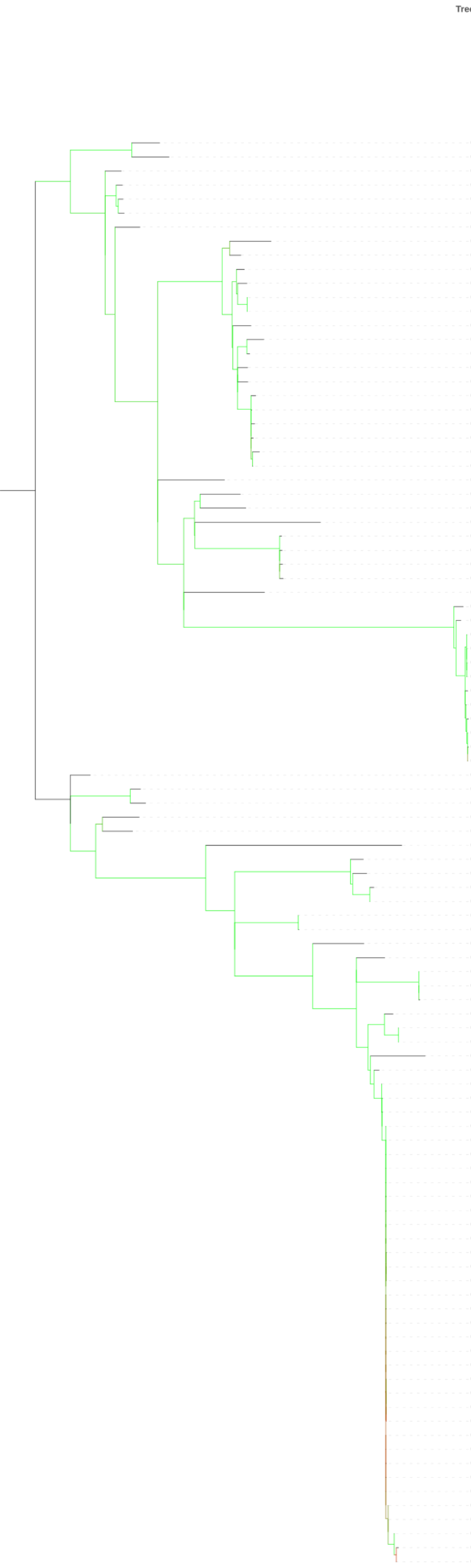


| | Isolate | MLST | SNP Cluster | Collection Year | Country | BioProject |
|---|---|---|---|---|---|---|
| GCA 033865535 | PDT001992543.1 | Klebsiella_13 | | 2016 | Canada | PRJNA532924 |
| GCA 034163965 | PDT002016309.1 | Klebsiella_13 | | 2023 | USA | PRJNA475751 |
| GCA 040364825 | PDT002233356.1 | Klebsiella_13 | | 2015 | Japan | PRJDB17849 |
| GCA 021939535 | PDT001049127.1 | Klebsiella_13 | | 2018 | Australia | PRJNA857533 |
| GCA 030715715 | PDT001849300.1 | Klebsiella_13 | PDS000185322.1 | 2019 | Oman | PRJNA999478 |
| GCA 040198915 | PDT002208356.1 | Klebsiella_13 | PDS000185322.1 | nan | Saudi Arabia | PRJNA1116866 |
| GCA 022292195 | PDT000427130.1 | Klebsiella_13 | | nan | NA | PRJEB10018 |
| GCA 905331375 | PDT001185214.2 | Klebsiella_13 | | 2018 | Spain | PRJEB42440 |
| GCA 013604195 | PDT000797598.1 | Klebsiella_13 | | nan | Spain | PRJNA609897 |
| GCA 031038295 | PDT001717196.3 | Klebsiella_13 | | 2023 | USA | PRJNA288601 |
| GCA 024489195 | PDT001373413.1 | Klebsiella_13 | | 2019 | Spain | PRJEB50822 |
| GCA 033122965 | PDT001958059.1 | Klebsiella_13 | PDS000197052.1 | 2015 | Norway | PRJEB27256 |
| GCA 041765315 | PDT002366259.1 | Klebsiella_13 | PDS000197052.1 | 2015 | Norway | PRJNA1098507 |
| GCA 043905525 | **HBH_1304_11** | Klebsiella_13 | | 2011 | Tunisia | PRJNA1160997 |
| GCA 041423215 | PDT002335078.1 | Klebsiella_13 | PDS000194059.1 | 2017 | Portugal | PRJNA986835 |
| GCA 041422095 | PDT002335021.1 | Klebsiella_13 | PDS000194059.1 | 2017 | Portugal | PRJNA986835 |
| GCA 044382055 | PDT002456491.1 | Klebsiella_13 | | nan | Saudi Arabia | PRJNA1116866 |
| GCA 900513595 | PDT000369532.1 | Klebsiella_13 | | 2014 | Israel | PRJEB10018 |
| GCA 041424895 | PDT002335176.1 | Klebsiella_13 | PDS000056122.9 | 2017 | Portugal | PRJNA986835 |
| GCA 016643445 | PDT000933591.1 | Klebsiella_13 | PDS000056122.9 | nan | Portugal | PRJNA602991 |
| GCA 016643375 | PDT000933588.1 | Klebsiella_13 | PDS000056122.9 | nan | Portugal | PRJNA602991 |
| GCA 016643475 | PDT000933593.1 | Klebsiella_13 | PDS000056122.9 | nan | Portugal | PRJNA602991 |
| GCA 041422725 | PDT002335053.1 | Klebsiella_13 | PDS000056122.9 | 2017 | Portugal | PRJNA986835 |
| GCA 041423075 | PDT002335070.1 | Klebsiella_13 | PDS000056122.9 | 2017 | Portugal | PRJNA986835 |
| GCA 037397895 | PDT002117173.1 | Klebsiella_13 | | 2014 | USA | PRJNA288601 |
| GCA 900517715 | PDT000369921.1 | Klebsiella_13 | | 2014 | Spain | PRJEB10018 |
| GCA 041353175 | PDT002331172.1 | Klebsiella_13 | | nan | Spain | PRJNA985214 |
| GCA 043859495 | **HBH_1500_13** | Klebsiella_13 | | 2013 | Tunisia | PRJNA1160997 |
| GCA 025891035 | PDT001469143.1 | Klebsiella_13 | PDS000121641.1 | 2016 | Malaysia | PRJEB51966 |
| GCA 025888615 | PDT001469162.1 | Klebsiella_13 | PDS000121641.1 | 2016 | Malaysia | PRJEB51966 |
| GCA 025889755 | PDT001469144.1 | Klebsiella_13 | PDS000121641.1 | 2016 | Malaysia | PRJEB51966 |
| GCA 025889235 | PDT001469142.1 | Klebsiella_13 | PDS000121641.1 | 2016 | Malaysia | PRJEB51966 |
| GCA 002851195 | PDT000276054.1 | Klebsiella_13 | | 2004 | Colombia | PRJNA378654 |
| GCA 046198465 | PDT000995918.1 | Klebsiella_13 | PDS000041857.2 | 2006 | Germany | PRJNA716285 |
| GCA 046198385 | PDT000995919.1 | Klebsiella_13 | PDS000041857.2 | 2006 | Germany | PRJNA716285 |
| GCA 022246385 | PDT000441020.1 | Klebsiella_13 | PDS000041857.2 | nan | NA | PRJEB12699 |
| GCA 022246345 | PDT000441022.1 | Klebsiella_- | PDS000041857.2 | nan | NA | PRJEB12699 |
| GCA 022246415 | PDT000441019.1 | Klebsiella_- | PDS000041857.2 | nan | NA | PRJEB12699 |
| GCA 022246445 | PDT000441018.1 | Klebsiella_- | PDS000041857.2 | nan | NA | PRJEB12699 |
| GCA 022246885 | PDT000440994.1 | Klebsiella_- | PDS000041857.2 | nan | NA | PRJEB12699 |
| GCA 022246465 | PDT000441017.1 | Klebsiella_- | PDS000041857.2 | nan | NA | PRJEB12699 |
| GCA 022246745 | PDT000441003.1 | Klebsiella_- | PDS000041857.2 | nan | NA | PRJEB12699 |
| GCA 022246405 | PDT000441024.1 | Klebsiella_- | PDS000041857.2 | nan | NA | PRJEB12699 |
| GCA 022246565 | PDT000441016.1 | Klebsiella_- | PDS000041857.2 | nan | NA | PRJEB12699 |
| GCA 022246305 | PDT000441023.1 | Klebsiella_- | PDS000041857.2 | nan | NA | PRJEB12699 |
| GCA 030874885 | PDT001701705.1 | Klebsiella_13 | | 2019 | Japan | PRJDB10842 |
| GCA 029813975 | PDT001693193.1 | Klebsiella_13 | | 2017 | Australia | PRJNA916637 |
| GCA 021968835 | PDT001019617.1 | Klebsiella_13 | | 2018 | Lebanon | PRJNA658369 |
| GCA 021773515 | PDT001233313.3 | Klebsiella_13 | PDS000140054.5 | 2021 | USA | PRJNA288601 |
| GCA 023677665 | PDT001323713.1 | Klebsiella_13 | PDS000110898.1 | 2018 | Australia | PRJNA797179 |
| GCA 916164085 | PDT001141304.1 | Klebsiella_13 | | 2014 | Netherlands | PRJEB41042 |
| GCA 022306535 | PDT000176097.2 | Klebsiella_13 | | nan | NA | PRJEB5065 |
| GCA 024430215 | PDT001366426.1 | Klebsiella_13 | PDS000053017.24 | nan | USA | PRJNA825705 |
| GCA 900503065 | PDT000370229.1 | Klebsiella_13 | PDS000098778.2 | 2014 | United Kingdom | PRJEB10018 |
| GCA 022194405 | PDT000715140.1 | Klebsiella_- | PDS000098778.2 | 2014 | United Kingdom | PRJNA564424 |
| GCA 047564655 | PDT002613266.1 | Klebsiella_- | PDS000217867.1 | 2019 | Chile | PRJNA962904 |
| GCA 047564485 | PDT002613317.1 | Klebsiella_- | PDS000217867.1 | 2019 | Chile | PRJNA962904 |
| GCA 022211965 | PDT000630400.1 | Klebsiella_- | | 2011 | Canada | PRJNA564992 |
| GCA 043272185 | PDT002271131.1 | Klebsiella_13 | | 2018 | Netherlands | PRJEB45369 |
| GCA 031010105 | PDT001844771.2 | Klebsiella_13 | PDS000155185.4 | 2023 | USA | PRJNA288601 |
| GCA 031014445 | PDT001826040.1 | Klebsiella_13 | PDS000155185.4 | 2023 | USA | PRJNA288601 |
| GCA 033132915 | PDT001961246.2 | Klebsiella_13 | PDS000155185.4 | 2023 | USA | PRJNA288601 |
| GCA 031047195 | PDT001726509.3 | Klebsiella_- | | 2018 | USA | PRJNA288601 |
| GCA 022159105 | PDT001243663.1 | Klebsiella_13 | PDS000104514.1 | 2015 | United Kingdom | PRJNA788733 |
| GCA 022157105 | PDT001243866.1 | Klebsiella_13 | PDS000104514.1 | 2015 | United Kingdom | PRJNA788733 |
| GCA 009907185 | PDT000668954.1 | Klebsiella_13 | | 2015 | Nigeria | PRJNA600702 |
| GCA 022297715 | PDT000423540.1 | Klebsiella_- | | nan | NA | PRJEB17615 |
| GCA 022232165 | PDT000443766.1 | Klebsiella_- | PDS000041826.3 | nan | NA | PRJEB8666 |
| GCA 022232185 | PDT000443764.1 | Klebsiella_13 | PDS000041826.3 | nan | NA | PRJEB8666 |
| GCA 022233005 | PDT000443780.1 | Klebsiella_- | PDS000041826.3 | nan | NA | PRJEB8666 |
| GCA 022231245 | PDT000444040.1 | Klebsiella_13 | PDS000041826.3 | nan | NA | PRJEB8666 |
| GCA 022030155 | PDT000921125.1 | Klebsiella_- | PDS000041826.3 | nan | NA | PRJEB27342 |
| GCA 022232205 | PDT000443761.1 | Klebsiella_- | PDS000041826.3 | nan | NA | PRJEB8666 |
| GCA 022029395 | PDT000921122.1 | Klebsiella_- | PDS000041826.3 | nan | NA | PRJEB27342 |
| GCA 022231625 | PDT000443953.1 | Klebsiella_- | PDS000041826.3 | nan | NA | PRJEB8666 |
| GCA 022232945 | PDT000443789.1 | Klebsiella_- | PDS000041826.3 | nan | NA | PRJEB8666 |
| GCA 022231765 | PDT000443935.1 | Klebsiella_13 | PDS000041826.3 | nan | NA | PRJEB8666 |
| GCA 022231685 | PDT000443945.1 | Klebsiella_13 | PDS000041826.3 | nan | NA | PRJEB8666 |
| GCA 022029555 | PDT000921113.1 | Klebsiella_- | PDS000041826.3 | nan | NA | PRJEB27342 |
| GCA 022232325 | PDT000443743.1 | Klebsiella_- | PDS000041826.3 | nan | NA | PRJEB8666 |
| GCA 022029455 | PDT000921118.1 | Klebsiella_- | PDS000041826.3 | nan | NA | PRJEB27342 |
| GCA 022232285 | PDT000443750.1 | Klebsiella_13 | PDS000041826.3 | nan | NA | PRJEB8666 |
| GCA 022232245 | PDT000443758.1 | Klebsiella_- | PDS000041826.3 | nan | NA | PRJEB8666 |
| GCA 022231705 | PDT000443947.1 | Klebsiella_13 | PDS000041826.3 | nan | NA | PRJEB8666 |
| GCA 022231665 | PDT000443950.1 | Klebsiella_- | PDS000041826.3 | nan | NA | PRJEB8666 |
| GCA 022231725 | PDT000443944.1 | Klebsiella_- | PDS000041826.3 | nan | NA | PRJEB8666 |
| GCA 022029335 | PDT000921123.1 | Klebsiella_- | PDS000041826.3 | nan | NA | PRJEB27342 |
| GCA 022029515 | PDT000921115.1 | Klebsiella_- | PDS000041826.3 | nan | NA | PRJEB27342 |
| GCA 022028855 | PDT000921126.1 | Klebsiella_- | PDS000041826.3 | nan | NA | PRJEB27342 |
| GCA 022231545 | PDT000443956.1 | Klebsiella_- | PDS000041826.3 | nan | NA | PRJEB8666 |
| GCA 022232925 | PDT000443784.1 | Klebsiella_- | PDS000041826.3 | nan | NA | PRJEB8666 |
| GCA 022231585 | PDT000443955.1 | Klebsiella_- | PDS000041826.3 | nan | NA | PRJEB8666 |
| GCA 022029295 | PDT000921124.1 | Klebsiella_- | PDS000041826.3 | nan | NA | PRJEB27342 |
| GCA 022232145 | PDT000443769.1 | Klebsiella_13 | PDS000041826.3 | nan | NA | PRJEB8666 |
| GCA 022231645 | PDT000443952.1 | Klebsiella_13 | PDS000041826.3 | nan | NA | PRJEB8666 |
| GCA 022232225 | PDT000443759.1 | Klebsiella_13 | PDS000041826.3 | nan | NA | PRJEB8666 |
| GCA 022232385 | PDT000443731.1 | Klebsiella_- | PDS000041826.3 | nan | NA | PRJEB8666 |
| GCA 022231885 | PDT000443903.1 | Klebsiella_- | PDS000041826.3 | nan | NA | PRJEB8666 |
| GCA 022232265 | PDT000443751.1 | Klebsiella_13 | PDS000041826.3 | nan | NA | PRJEB8666 |
| GCA 022029375 | PDT000921120.1 | Klebsiella_13 | PDS000041826.3 | nan | NA | PRJEB27342 |
| GCA 022232105 | PDT000443779.1 | Klebsiella_- | PDS000041826.3 | nan | NA | PRJEB8666 |
| GCA 022231745 | PDT000443942.1 | Klebsiella_- | PDS000041826.3 | nan | NA | PRJEB8666 |

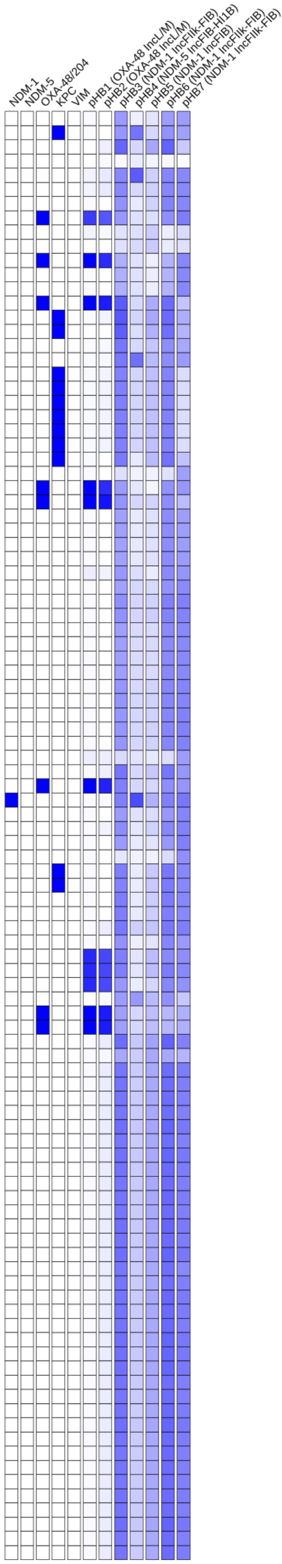

### Tree 6: GCA_043859615

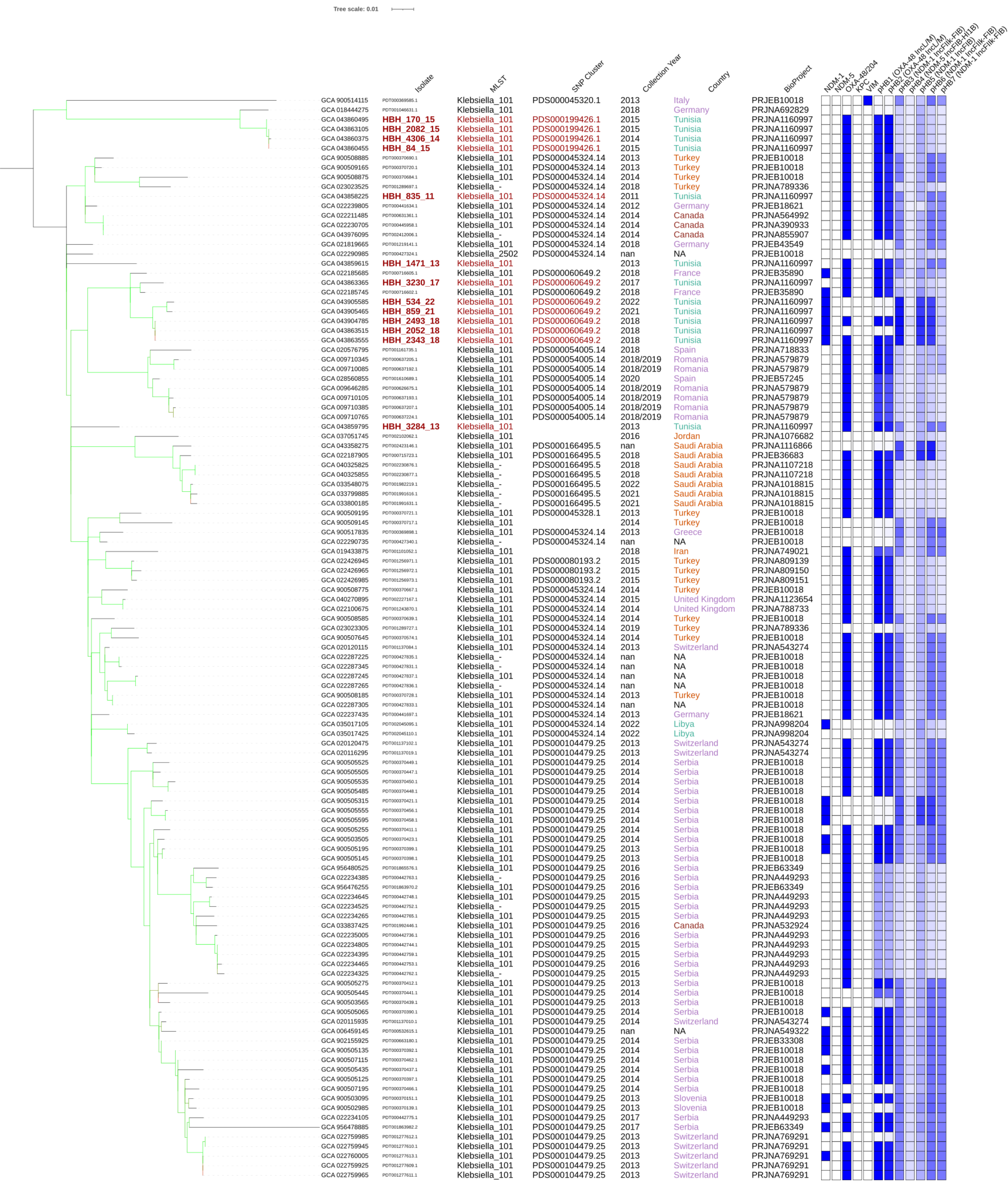

### Tree 7: GCA_043859815

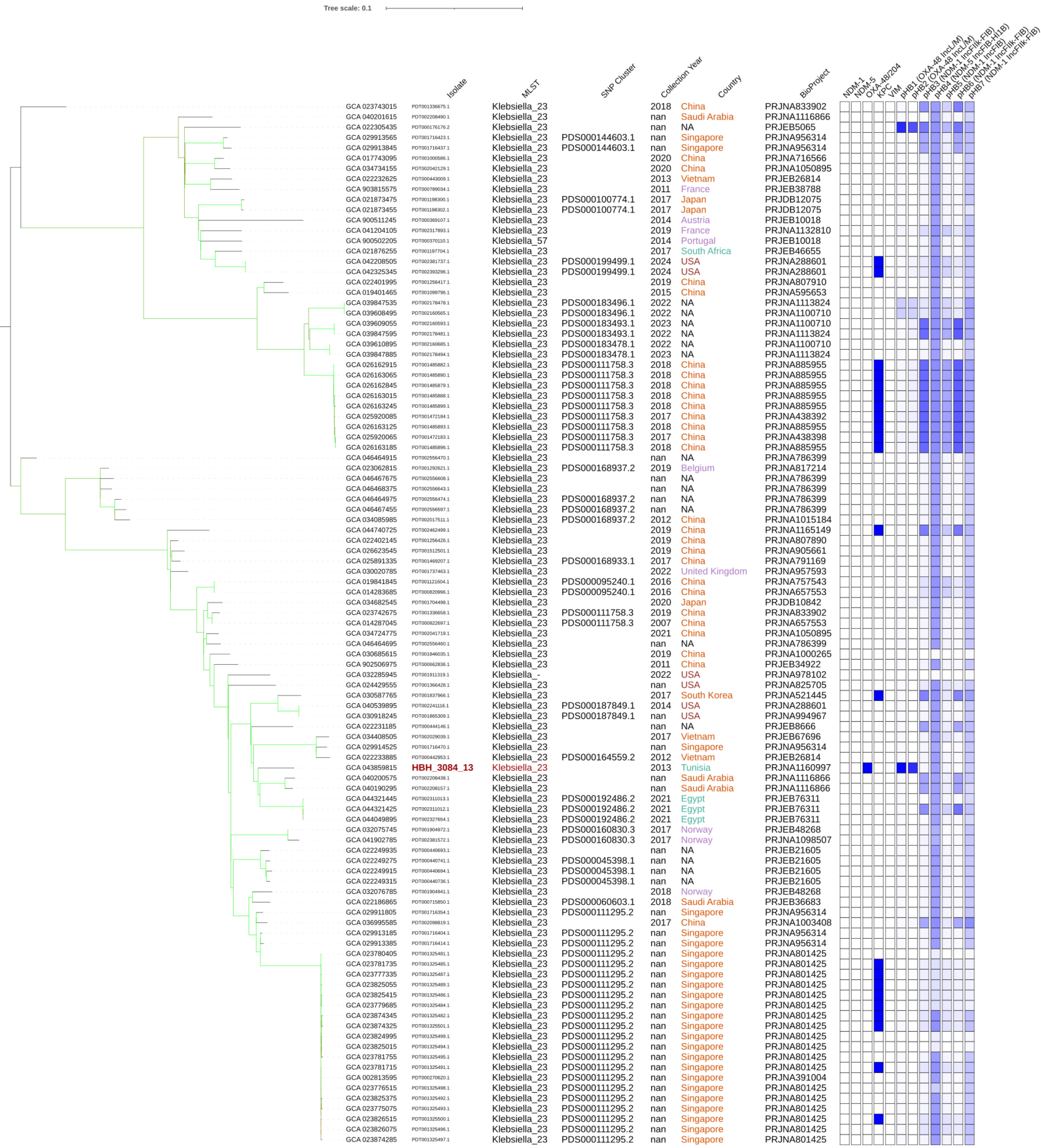

### Tree 8: GCA_043859855

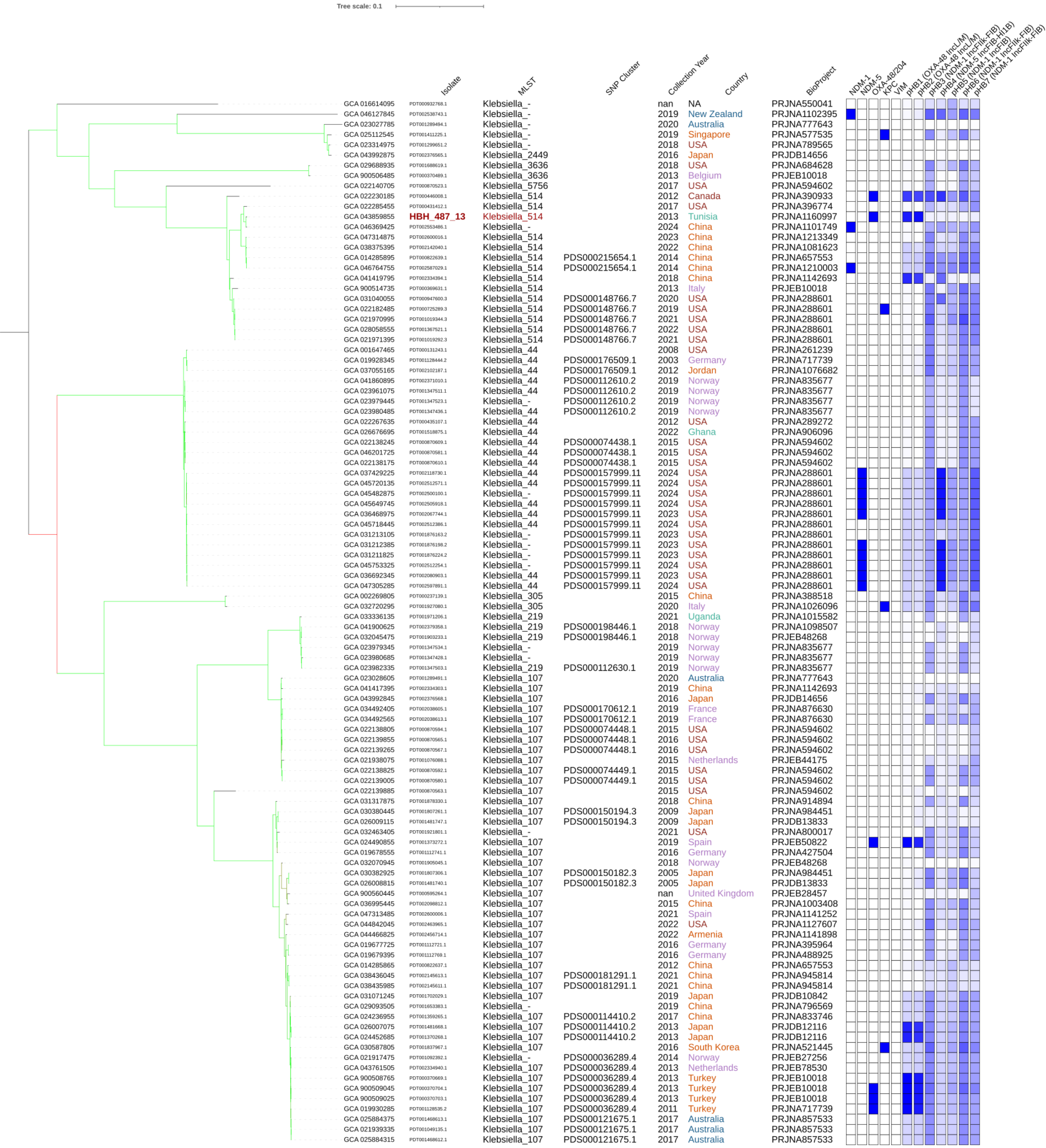

### Tree 9: GCA_043859995

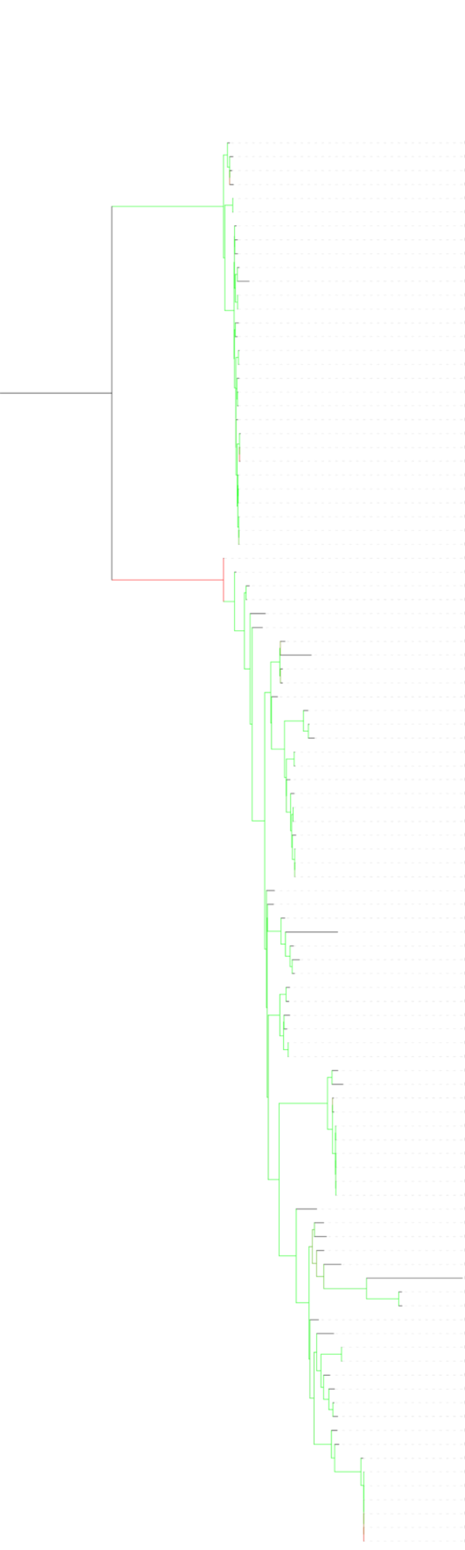


| | Isolate | MLST | SNP Cluster | Collection Year | Country | BioProject |
|---|---|---|---|---|---|---|
| GCA 900515165 | PDT000369408.1 | Klebsiella_1013 | | 2014 | Italy | PRJEB10018 |
| GCA 026335915 | PDT001494223.1 | Klebsiella_1013 | | 2020 | Germany | PRJEB56918 |
| GCA 900498695 | PDT000361084.1 | Klebsiella_1013 | | 2014 | Germany | PRJEB27975 |
| GCA 022267655 | PDT000435112.1 | Klebsiella_1013 | | 2012 | USA | PRJNA289272 |
| GCA 900506315 | PDT000370473.1 | Klebsiella_5 | PDS000036226.2 | 2014 | Ireland | PRJEB10018 |
| GCA 900506325 | PDT000370476.1 | Klebsiella_5 | PDS000036226.2 | 2014 | Ireland | PRJEB10018 |
| GCA 027152215 | PDT001543585.1 | Klebsiella_5 | | 2016 | Italy | PRJNA781811 |
| GCA 035618615 | PDT002052413.1 | Klebsiella_5 | | 2021 | Italy | PRJNA814467 |
| GCA 036949905 | PDT002096688.2 | Klebsiella_5 | | 2019 | Belgium | PRJNA927131 |
| GCA 024433395 | PDT001366221.1 | Klebsiella_5 | | 2019 | Guadeloupe | PRJNA778230 |
| GCA 900085255 | PDT000131445.1 | Klebsiella_5 | | 2008 | United Kingdom | PRJEB5065 |
| GCA 029955985 | PDT001727354.1 | Klebsiella_5 | PDS000145396.1 | 2022 | Ecuador | PRJNA953974 |
| GCA 029956045 | PDT001727355.1 | Klebsiella_5 | PDS000145396.1 | 2022 | Ecuador | PRJNA953974 |
| GCA 036951305 | PDT002096711.2 | Klebsiella_5 | PDS000175697.1 | 2018 | Belgium | PRJNA927131 |
| GCA 900503005 | PDT000370142.1 | Klebsiella_5 | | 2014 | Slovenia | PRJEB10018 |
| GCA 040988785 | PDT002279750.1 | Klebsiella_5 | | nan | China | PRJNA1021041 |
| GCA 046464945 | PDT002556471.1 | Klebsiella_5 | | nan | NA | PRJNA786399 |
| GCA 016614895 | PDT000932803.1 | Klebsiella_5 | | nan | NA | PRJNA550041 |
| GCA 034739495 | PDT002042380.1 | Klebsiella_5 | | 2021 | China | PRJNA1050895 |
| GCA 034725675 | PDT002041742.1 | Klebsiella_5 | | 2021 | China | PRJNA1050895 |
| GCA 034731975 | PDT002042049.1 | Klebsiella_5 | | 2020 | China | PRJNA1050895 |
| GCA 034737775 | PDT002042295.1 | Klebsiella_5 | PDS000170846.1 | 2021 | China | PRJNA1050895 |
| GCA 034732975 | PDT002042074.1 | Klebsiella_5 | | 2020 | China | PRJNA1050895 |
| GCA 034738695 | PDT002042341.1 | Klebsiella_5 | | 2021 | China | PRJNA1050895 |
| GCA 034734315 | PDT002042137.1 | Klebsiella_5 | PDS000170904.1 | 2021 | China | PRJNA1050895 |
| GCA 034726715 | PDT002041775.1 | Klebsiella_5 | PDS000170904.1 | 2021 | China | PRJNA1050895 |
| GCA 034726755 | PDT002041777.1 | Klebsiella_5 | PDS000170904.1 | 2021 | China | PRJNA1050895 |
| GCA 021939095 | PDT001049144.1 | Klebsiella_4510 | PDS000090692.1 | 2018 | Australia | PRJNA857533 |
| GCA 021939135 | PDT001049143.1 | Klebsiella_4510 | PDS000090692.1 | 2018 | Australia | PRJNA857533 |
| GCA 021939175 | PDT001049141.1 | Klebsiella_4510 | PDS000090692.1 | 2018 | Australia | PRJNA857533 |
| GCA 000567125 | PDT000022744.1 | Klebsiella_540 | | 2012 | USA | PRJNA219254 |
| GCA 022231345 | PDT000444017.1 | Klebsiella_540 | | nan | NA | PRJEB8666 |
| GCA 033336295 | PDT001971213.1 | Klebsiella_540 | PDS000165203.1 | 2021 | Uganda | PRJNA1015582 |
| GCA 021993315 | PDT000989500.1 | Klebsiella_540 | | nan | NA | PRJEB27342 |
| GCA 030520285 | PDT001827421.1 | Klebsiella_- | | 2022 | Uganda | PRJNA985213 |
| GCA 004312705 | PDT000471500.1 | Klebsiella_540 | | 2014 | Japan | PRJDB7163 |
| GCA 037054045 | PDT002102131.1 | Klebsiella_540 | | 2012 | Jordan | PRJNA1076682 |
| GCA 021977835 | PDT001011381.1 | Klebsiella_540 | | 2015 | Norway | PRJEB42350 |
| GCA 900498885 | PDT000361083.1 | Klebsiella_540 | | 2014 | Germany | PRJEB27975 |
| GCA 022258355 | PDT000437251.1 | Klebsiella_540 | | 2004 | Vietnam | PRJEB2111 |
| GCA 021970755 | PDT001019558.1 | Klebsiella_540 | | 2017 | USA | PRJNA658369 |
| GCA 021264305 | PDT001211222.1 | Klebsiella_- | | nan | USA | PRJNA788509 |
| GCA 045487615 | PDT002191104.1 | Klebsiella_540 | | 2017 | USA | PRJNA1054115 |
| GCA 022267595 | PDT000435111.1 | Klebsiella_540 | | 2012 | USA | PRJNA289272 |
| GCA 040293125 | PDT002230728.1 | Klebsiella_540 | | 2017 | Bangladesh | PRJNA719593 |
| GCA 027893015 | PDT001557941.1 | Klebsiella_540 | | 2020 | Bangladesh | PRJNA845975 |
| GCA 043857945 | PDT002431738.1 | Klebsiella_540 | | 2018 | China | PRJNA1175053 |
| GCA 021906335 | PDT001141058.1 | Klebsiella_540 | | 2017 | Vietnam | PRJNA641536 |
| GCA 021999495 | PDT000987036.1 | Klebsiella_540 | | 2016 | Thailand | PRJDB5929 |
| GCA 900493105 | PDT000354553.1 | Klebsiella_540 | | 2016 | Thailand | PRJEB19226 |
| GCA 022250115 | PDT000440685.1 | Klebsiella_540 | | nan | NA | PRJEB21605 |
| GCA 045575395 | PDT002509443.1 | Klebsiella_540 | PDS000140723.2 | nan | Singapore | PRJNA1171009 |
| GCA 029493505 | PDT001672815.1 | Klebsiella_540 | PDS000140723.2 | nan | Singapore | PRJNA757551 |
| GCA 022266055 | PDT000435200.1 | Klebsiella_540 | PDS000140723.2 | 2014 | Singapore | PRJNA342893 |
| GCA 027615845 | PDT001558316.1 | Klebsiella_540 | | 2006 | Bangladesh | PRJNA845975 |
| GCA 900452445 | PDT000357789.1 | Klebsiella_504 | | 1900/1952 | NA | PRJEB6403 |
| GCA 021920715 | PDT001092161.1 | Klebsiella_504 | | 2009 | Norway | PRJEB27256 |
| GCA 043570105 | PDT002347458.1 | Klebsiella_504 | | 2015 | Norway | PRJEB27256 |
| GCA 900508795 | PDT000370675.1 | Klebsiella_504 | | 2014 | Turkey | PRJEB10018 |
| GCA 901421135 | PDT000526521.1 | Klebsiella_504 | | 1953 | USA | PRJEB6403 |
| GCA 022203745 | PDT000674645.1 | Klebsiella_504 | | 2018 | Italy | PRJEB27342 |
| GCA 014883955 | PDT000863725.1 | Klebsiella_504 | | 2015 | Colombia | PRJNA665232 |
| GCA 022392705 | PDT001254598.1 | Klebsiella_3319 | | 2015 | Madagascar | PRJNA580525 |
| GCA 956481305 | PDT001865613.1 | Klebsiella_2086 | | 2016 | Spain | PRJEB63349 |
| GCA 043859995 | **HBH_653_13** | Klebsiella_2086 | | 2013 | Tunisia | PRJNA1160997 |
| GCA 023208335 | PDT001296765.1 | Klebsiella_2086 | PDS000108803.2 | 2018 | USA | PRJNA789565 |
| GCA 023208275 | PDT001296768.2 | Klebsiella_2086 | PDS000108803.2 | 2018 | USA | PRJNA789565 |
| GCA 022306595 | PDT001250702.1 | Klebsiella_540 | | 2019 | Kenya | PRJNA804332 |
| GCA 033802105 | PDT001991727.1 | Klebsiella_540 | | 2020 | Saudi Arabia | PRJNA1018815 |
| GCA 903684355 | PDT001251919.1 | Klebsiella_540 | | 2017 | Ghana | PRJEB38442 |
| GCA 046465345 | PDT002556493.1 | Klebsiella_540 | | nan | NA | PRJNA786399 |
| GCA 023978955 | PDT001347551.1 | Klebsiella_540 | PDS000112629.2 | 2018 | USA | PRJNA842739 |
| GCA 046119775 | PDT002539599.1 | Klebsiella_540 | PDS000112629.2 | 2019 | USA | PRJNA858823 |
| GCA 046119535 | PDT002539626.1 | Klebsiella_540 | PDS000112629.2 | 2017 | USA | PRJNA858823 |
| GCA 023978835 | PDT001347557.1 | Klebsiella_540 | PDS000112629.2 | 2017 | USA | PRJNA842739 |
| GCA 046119875 | PDT002539594.1 | Klebsiella_540 | PDS000112629.2 | 2017 | USA | PRJNA858823 |
| GCA 023978795 | PDT001347559.1 | Klebsiella_540 | PDS000112629.2 | 2017 | USA | PRJNA842739 |
| GCA 028062295 | PDT001175579.3 | Klebsiella_- | | 2021 | USA | PRJNA288601 |
| GCA 039719115 | PDT002172146.1 | Klebsiella_1087 | | 2023 | India | PRJNA1109570 |
| GCA 033105385 | PDT001960999.1 | Klebsiella_- | | 2021 | USA | PRJNA648389 |
| GCA 029526625 | PDT001676560.1 | Klebsiella_1087 | | 2019 | China | PRJNA810290 |
| GCA 016918985 | PDT000971346.1 | Klebsiella_1087 | | 2016 | India | PRJNA694019 |
| GCA 016761975 | PDT000945127.2 | Klebsiella_- | | 2018 | Bangladesh | PRJNA659108 |
| GCA 027892215 | PDT001557972.1 | Klebsiella_- | PDS000130868.1 | 2009 | Bangladesh | PRJNA845975 |
| GCA 027828125 | PDT001557994.1 | Klebsiella_- | | 2012 | Bangladesh | PRJNA845975 |
| GCA 031069165 | PDT001702406.1 | Klebsiella_1087 | | 2020 | Japan | PRJDB10842 |
| GCA 043000145 | PDT002129370.1 | Klebsiella_1087 | | 2019 | Egypt | PRJNA1071125 |
| GCA 026223775 | PDT001490502.1 | Klebsiella_6245 | PDS000123677.1 | 2020 | Brazil | PRJNA879417 |
| GCA 026223715 | PDT001490499.1 | Klebsiella_6245 | PDS000123677.1 | 2020 | Brazil | PRJNA879417 |
| GCA 036993625 | PDT002098721.1 | Klebsiella_1087 | | 2016 | China | PRJNA1003408 |
| GCA 034744975 | PDT002043131.1 | Klebsiella_1087 | | 2021 | China | PRJNA1050895 |
| GCA 034742855 | PDT002043026.1 | Klebsiella_1087 | | 2021 | China | PRJNA1050895 |
| GCA 039960775 | PDT002182482.1 | Klebsiella_1087 | | 2024 | USA | PRJNA288601 |
| GCA 016616695 | PDT000932866.1 | Klebsiella_1087 | | nan | NA | PRJNA550041 |
| GCA 021920405 | PDT001092188.1 | Klebsiella_1087 | | 2009 | Norway | PRJEB27256 |
| GCA 022323765 | PDT000535798.1 | Klebsiella_1087 | | 2016 | India | PRJEB30913 |
| GCA 021992575 | PDT000989525.1 | Klebsiella_1087 | PDS000083576.2 | nan | NA | PRJEB27342 |
| GCA 021989035 | PDT000989606.1 | Klebsiella_1087 | PDS000083576.2 | nan | NA | PRJEB27342 |
| GCA 021996135 | PDT000989423.1 | Klebsiella_- | PDS000083576.2 | nan | NA | PRJEB27342 |
| GCA 021993015 | PDT000989508.1 | Klebsiella_- | PDS000083576.2 | nan | NA | PRJEB27342 |
| GCA 021994195 | PDT000989471.1 | Klebsiella_1087 | PDS000083576.2 | nan | NA | PRJEB27342 |
| GCA 021993275 | PDT000989505.1 | Klebsiella_- | PDS000083576.2 | nan | NA | PRJEB27342 |

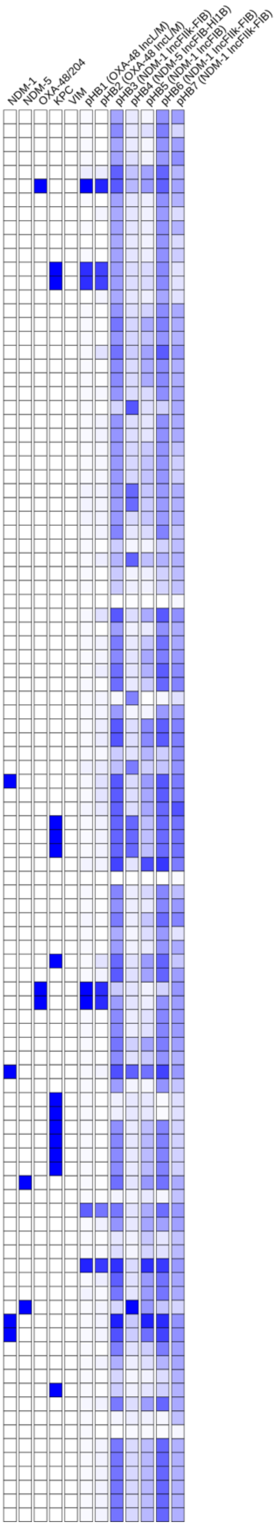

### Tree 10: GCA_043860015

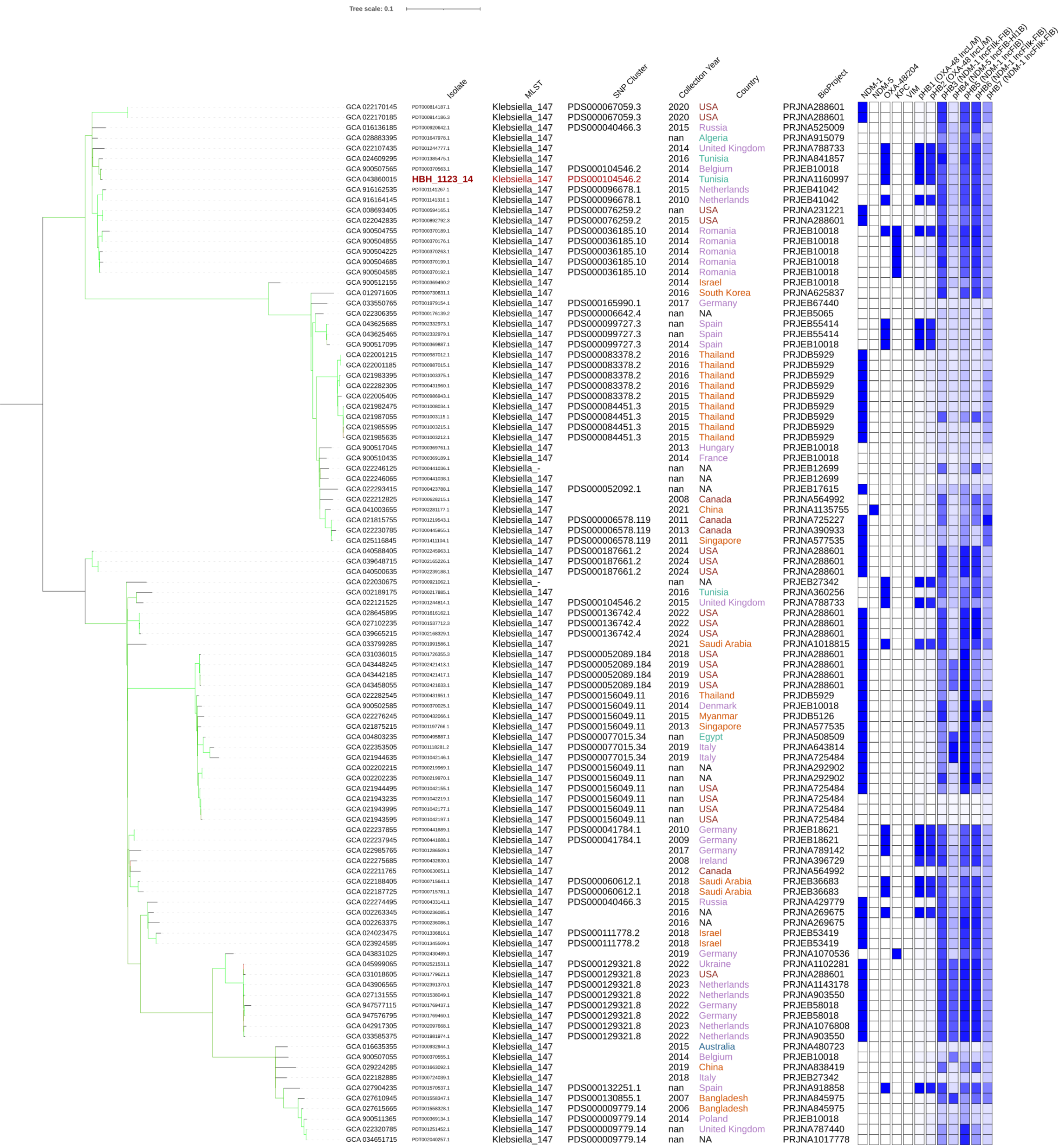

### Tree 11: GCA_043860075

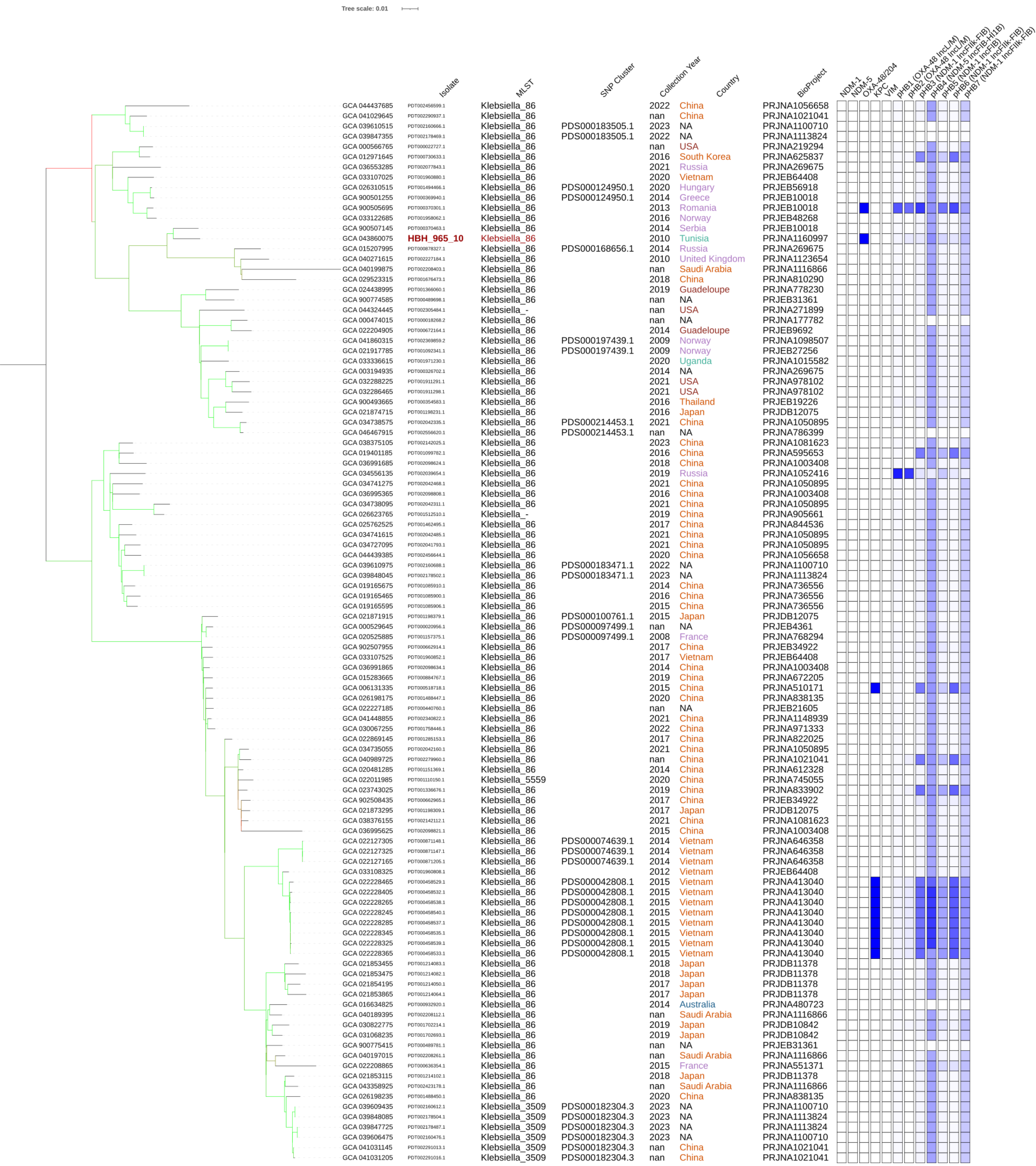

### Tree 12: GCA_043860195

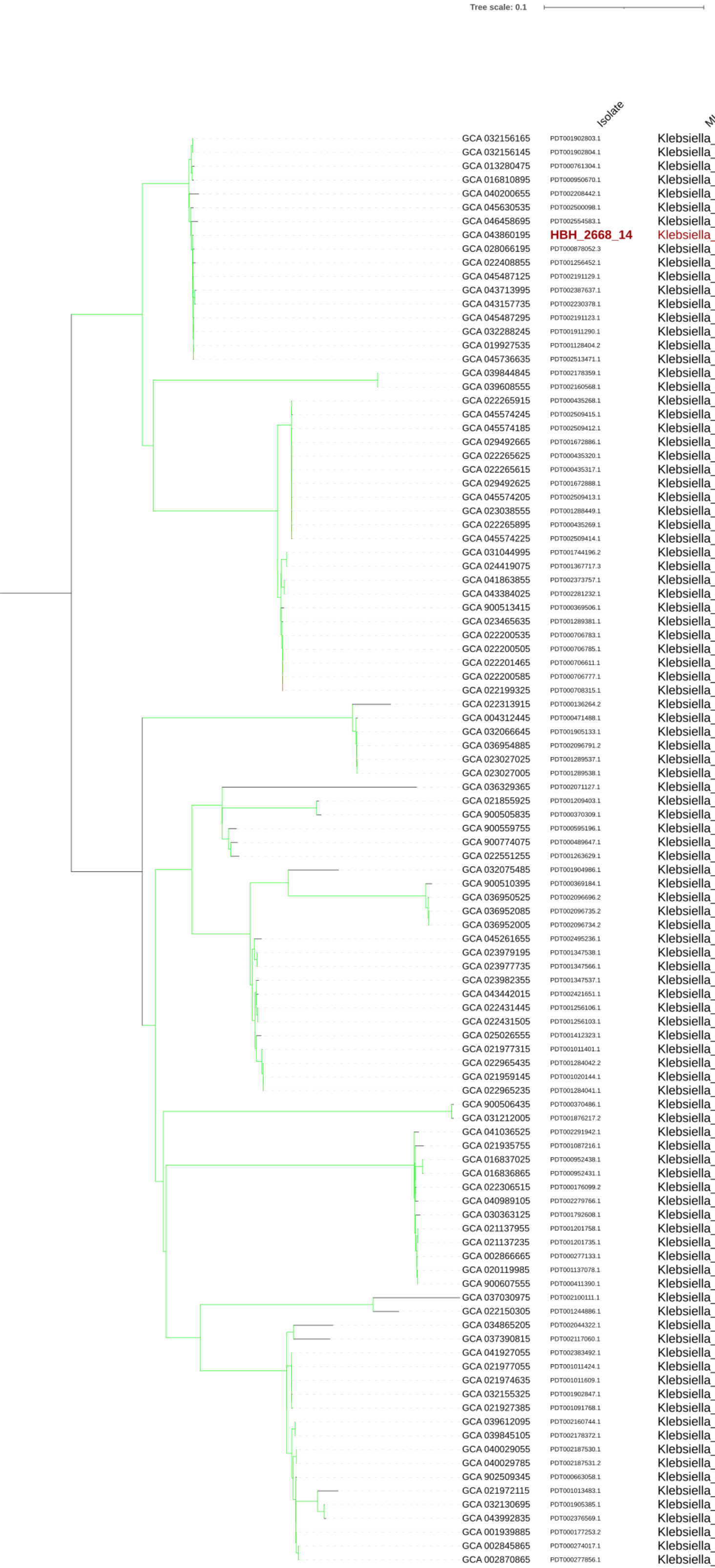


| | Isolate | MLST | SNP Cluster | Collection Year | Country | BioProject |
|---|---|---|---|---|---|---|
| GCA 032156165 | PDT001902803.1 | Klebsiella_987 | PDS000160856.1 | 2018 | Norway | PRJEB48268 |
| GCA 032156145 | PDT001902804.1 | Klebsiella_987 | PDS000160856.1 | 2018 | Norway | PRJEB48268 |
| GCA 013280475 | PDT000761304.1 | Klebsiella_987 | | 2019 | Lebanon | PRJNA634137 |
| GCA 016810895 | PDT000950670.1 | Klebsiella_987 | | 2017 | Qatar | PRJNA690895 |
| GCA 040200655 | PDT002208442.1 | Klebsiella_987 | | nan | Saudi Arabia | PRJNA1116866 |
| GCA 045630535 | PDT002500098.1 | Klebsiella_987 | | 2024 | USA | PRJNA288601 |
| GCA 046458695 | PDT002554583.1 | Klebsiella_987 | | 2019 | Qatar | PRJNA1005941 |
| GCA 043860195 | **HBH_2668_14** | Klebsiella_987 | | 2014 | Tunisia | PRJNA1160997 |
| GCA 028066195 | PDT000878052.3 | Klebsiella_987 | | 2018 | USA | PRJNA288601 |
| GCA 022408855 | PDT001256452.1 | Klebsiella_987 | | nan | USA | PRJNA786742 |
| GCA 045487125 | PDT002191129.1 | Klebsiella_987 | PDS000161308.3 | 2018 | USA | PRJNA1054115 |
| GCA 043713995 | PDT002387637.1 | Klebsiella_987 | PDS000199202.1 | 2024 | USA | PRJNA1127607 |
| GCA 043157735 | PDT002230378.1 | Klebsiella_987 | PDS000199202.1 | 2024 | USA | PRJNA1127607 |
| GCA 045487295 | PDT002191123.1 | Klebsiella_987 | PDS000161308.3 | 2019 | USA | PRJNA1054115 |
| GCA 032288245 | PDT001911290.1 | Klebsiella_987 | PDS000161308.3 | 2021 | USA | PRJNA978102 |
| GCA 019927535 | PDT001128404.2 | Klebsiella_987 | PDS000161308.3 | 2019 | USA | PRJNA717739 |
| GCA 045736635 | PDT002513471.1 | Klebsiella_987 | PDS000161308.3 | 2024 | USA | PRJNA475751 |
| GCA 039844845 | PDT002178359.1 | Klebsiella_- | PDS000183523.1 | 2023 | NA | PRJNA1113824 |
| GCA 039608555 | PDT002160568.1 | Klebsiella_- | PDS000183523.1 | 2022 | NA | PRJNA1100710 |
| GCA 022265915 | PDT000435268.1 | Klebsiella_496 | PDS000071977.4 | 2014 | Singapore | PRJNA342893 |
| GCA 045574245 | PDT002509415.1 | Klebsiella_496 | PDS000071977.4 | nan | Singapore | PRJNA1171009 |
| GCA 045574185 | PDT002509412.1 | Klebsiella_496 | PDS000071977.4 | nan | Singapore | PRJNA1171009 |
| GCA 029492665 | PDT001672886.1 | Klebsiella_496 | PDS000071977.4 | nan | Singapore | PRJNA765801 |
| GCA 022265625 | PDT000435320.1 | Klebsiella_496 | PDS000071977.4 | 2014 | Singapore | PRJNA342893 |
| GCA 022265615 | PDT000435317.1 | Klebsiella_496 | PDS000071977.4 | 2014 | Singapore | PRJNA342893 |
| GCA 029492625 | PDT001672888.1 | Klebsiella_496 | PDS000071977.4 | nan | Singapore | PRJNA765801 |
| GCA 045574205 | PDT002509413.1 | Klebsiella_496 | PDS000071977.4 | nan | Singapore | PRJNA1171009 |
| GCA 023038555 | PDT001288449.1 | Klebsiella_496 | PDS000071977.4 | nan | Singapore | PRJNA757551 |
| GCA 022265895 | PDT000435269.1 | Klebsiella_- | PDS000071977.4 | 2014 | Singapore | PRJNA342893 |
| GCA 045574225 | PDT002509414.1 | Klebsiella_496 | PDS000071977.4 | nan | Singapore | PRJNA1171009 |
| GCA 031044995 | PDT001744196.2 | Klebsiella_496 | PDS000178659.1 | 2022 | USA | PRJNA278886 |
| GCA 024419075 | PDT001367717.3 | Klebsiella_496 | PDS000178659.1 | 2022 | USA | PRJNA288601 |
| GCA 041863855 | PDT002373757.1 | Klebsiella_496 | PDS000197593.1 | 2009 | Norway | PRJNA1098507 |
| GCA 043384025 | PDT002281232.1 | Klebsiella_496 | PDS000197593.1 | 2009 | Norway | PRJEB27256 |
| GCA 900513415 | PDT000369506.1 | Klebsiella_4064 | | 2013 | Israel | PRJEB10018 |
| GCA 023465635 | PDT001289381.1 | Klebsiella_- | | 2020 | Australia | PRJNA777643 |
| GCA 022200535 | PDT000706783.1 | Klebsiella_- | PDS000059976.3 | 2018 | Italy | PRJEB27342 |
| GCA 022200505 | PDT000706785.1 | Klebsiella_496 | PDS000059976.3 | 2018 | Italy | PRJEB27342 |
| GCA 022201465 | PDT000706611.1 | Klebsiella_- | PDS000059976.3 | 2018 | Italy | PRJEB27342 |
| GCA 022200585 | PDT000706777.1 | Klebsiella_496 | PDS000059976.3 | 2018 | Italy | PRJEB27342 |
| GCA 022199325 | PDT000708315.1 | Klebsiella_- | PDS000059976.3 | 2018 | Italy | PRJEB27342 |
| GCA 022313915 | PDT000136264.2 | Klebsiella_485 | | 2012 | United Kingdom | PRJNA306133 |
| GCA 004312445 | PDT000471488.1 | Klebsiella_485 | | 2014 | Japan | PRJDB7163 |
| GCA 032066645 | PDT001905133.1 | Klebsiella_485 | PDS000053037.8 | 2018 | Norway | PRJEB48268 |
| GCA 036954885 | PDT002096791.2 | Klebsiella_485 | PDS000053037.8 | 2018 | Netherlands | PRJNA927131 |
| GCA 023027025 | PDT001289537.1 | Klebsiella_- | PDS000053037.8 | 2020 | Australia | PRJNA777643 |
| GCA 023027005 | PDT001289538.1 | Klebsiella_485 | PDS000053037.8 | 2020 | Australia | PRJNA777643 |
| GCA 036329365 | PDT002071127.1 | Klebsiella_485 | | 2020 | China | PRJNA930978 |
| GCA 021855925 | PDT001209403.1 | Klebsiella_163 | | 2018 | Italy | PRJEB27342 |
| GCA 900505835 | PDT000370309.1 | Klebsiella_163 | | 2013 | Croatia | PRJEB10018 |
| GCA 900559755 | PDT000595196.1 | Klebsiella_485 | | nan | United Kingdom | PRJEB28457 |
| GCA 900774075 | PDT000489647.1 | Klebsiella_485 | | nan | NA | PRJEB31361 |
| GCA 022551255 | PDT001263629.1 | Klebsiella_485 | | 2017 | USA | PRJNA726883 |
| GCA 032075485 | PDT001904986.1 | Klebsiella_485 | | 2017 | Norway | PRJEB48268 |
| GCA 900510395 | PDT000369184.1 | Klebsiella_3649 | | 2013 | France | PRJEB10018 |
| GCA 036950525 | PDT002096696.2 | Klebsiella_3649 | | 2019 | Belgium | PRJNA927131 |
| GCA 036952085 | PDT002096735.2 | Klebsiella_3649 | PDS000175698.1 | 2017 | Belgium | PRJNA927131 |
| GCA 036952005 | PDT002096734.2 | Klebsiella_3649 | PDS000175698.1 | 2017 | Belgium | PRJNA927131 |
| GCA 045261655 | PDT002495236.1 | Klebsiella_485 | | nan | NA | PRJNA1187231 |
| GCA 023979195 | PDT001347538.1 | Klebsiella_485 | PDS000112639.1 | 2012 | USA | PRJNA842739 |
| GCA 023977735 | PDT001347566.1 | Klebsiella_485 | PDS000112639.1 | 2012 | USA | PRJNA842739 |
| GCA 023982355 | PDT001347537.1 | Klebsiella_485 | | 2019 | Norway | PRJNA835677 |
| GCA 043442015 | PDT002421651.1 | Klebsiella_485 | | 2019 | USA | PRJNA288601 |
| GCA 022431445 | PDT001256106.1 | Klebsiella_485 | PDS000105717.1 | 2019 | USA | PRJNA325243 |
| GCA 022431505 | PDT001256103.1 | Klebsiella_485 | PDS000105717.1 | 2019 | USA | PRJNA325243 |
| GCA 025026555 | PDT001412323.1 | Klebsiella_485 | | 2019 | Norway | PRJNA835677 |
| GCA 021977315 | PDT001011401.1 | Klebsiella_- | | 2015 | Norway | PRJEB42350 |
| GCA 022965435 | PDT001284042.2 | Klebsiella_- | PDS000107774.2 | 2017 | USA | PRJNA789565 |
| GCA 021959145 | PDT001020144.1 | Klebsiella_- | PDS000107774.2 | 2017 | USA | PRJNA658369 |
| GCA 022965235 | PDT001284041.1 | Klebsiella_- | PDS000107774.2 | 2017 | USA | PRJNA789565 |
| GCA 900506435 | PDT000370486.1 | Klebsiella_485 | | 2014 | Ireland | PRJEB10018 |
| GCA 031212005 | PDT001876217.2 | Klebsiella_- | | 2023 | USA | PRJNA288601 |
| GCA 041036525 | PDT002291942.1 | Klebsiella_914 | | nan | China | PRJNA1021041 |
| GCA 021935755 | PDT001087216.1 | Klebsiella_914 | | 2018 | Australia | PRJNA529744 |
| GCA 016837025 | PDT000952438.1 | Klebsiella_914 | PDS000080493.2 | 2012 | Paraguay | PRJNA698801 |
| GCA 016836865 | PDT000952431.1 | Klebsiella_914 | PDS000080493.2 | 2012 | Paraguay | PRJNA698801 |
| GCA 022306515 | PDT000176099.2 | Klebsiella_914 | | nan | NA | PRJEB5065 |
| GCA 040989105 | PDT002279766.1 | Klebsiella_914 | | nan | China | PRJNA1021041 |
| GCA 030363125 | PDT001792608.1 | Klebsiella_6114 | | 2020 | China | PRJNA925808 |
| GCA 021137955 | PDT001201758.1 | Klebsiella_2719 | PDS000038919.5 | 2020 | Switzerland | PRJNA759850 |
| GCA 021137235 | PDT001201735.1 | Klebsiella_2719 | PDS000038919.5 | 2020 | Switzerland | PRJNA758223 |
| GCA 002866665 | PDT000277133.1 | Klebsiella_2719 | PDS000038919.5 | 2013 | Lebanon | PRJNA388273 |
| GCA 020119985 | PDT001137078.1 | Klebsiella_2719 | PDS000038919.5 | 2013 | Switzerland | PRJNA543274 |
| GCA 900607555 | PDT000411390.1 | Klebsiella_2719 | PDS000038919.5 | 2013 | Switzerland | PRJEB28660 |
| GCA 037030975 | PDT002100111.1 | Klebsiella_485 | | 2023 | Mexico | PRJNA1051187 |
| GCA 022150305 | PDT001244886.1 | Klebsiella_485 | | 2016 | United Kingdom | PRJNA788733 |
| GCA 034865205 | PDT002044322.1 | Klebsiella_485 | | 2014 | France | PRJNA1054041 |
| GCA 037390815 | PDT002117060.1 | Klebsiella_815 | | 2014 | USA | PRJNA288601 |
| GCA 041927055 | PDT002383492.1 | Klebsiella_485 | PDS000085062.4 | 2015 | Norway | PRJNA1098507 |
| GCA 021977055 | PDT001011424.1 | Klebsiella_485 | PDS000085062.4 | 2015 | Norway | PRJEB42350 |
| GCA 021974635 | PDT001011609.1 | Klebsiella_485 | PDS000085062.4 | 2015 | Norway | PRJEB42350 |
| GCA 032155325 | PDT001902847.1 | Klebsiella_485 | PDS000085062.4 | 2017 | Norway | PRJEB48268 |
| GCA 021927385 | PDT001091768.1 | Klebsiella_485 | PDS000085062.4 | 2015 | Norway | PRJEB27256 |
| GCA 039612095 | PDT002160744.1 | Klebsiella_485 | PDS000183592.1 | 2022 | NA | PRJNA1100710 |
| GCA 039845105 | PDT002178372.1 | Klebsiella_485 | PDS000183592.1 | 2023 | NA | PRJNA1113824 |
| GCA 040029055 | PDT002187530.1 | Klebsiella_485 | PDS000184186.2 | 2019 | China | PRJNA1116330 |
| GCA 040029785 | PDT002187531.2 | Klebsiella_485 | PDS000184186.2 | 2019 | China | PRJNA1116332 |
| GCA 902509345 | PDT000663058.1 | Klebsiella_485 | | 2017 | China | PRJEB34922 |
| GCA 021972115 | PDT001013483.1 | Klebsiella_485 | | 2018 | Denmark | PRJEB37711 |
| GCA 032130695 | PDT001905385.1 | Klebsiella_485 | | 2020 | Japan | PRJDB16654 |
| GCA 043992835 | PDT002376569.1 | Klebsiella_485 | | 2016 | Japan | PRJDB14656 |
| GCA 001939885 | PDT000177253.2 | Klebsiella_485 | PDS000054678.2 | 2015 | China | PRJNA349219 |
| GCA 002845865 | PDT000274017.1 | Klebsiella_485 | PDS000054678.2 | 2009 | China | PRJNA420654 |
| GCA 002870865 | PDT000277856.1 | Klebsiella_485 | PDS000054678.2 | 2011 | China | PRJNA428155 |

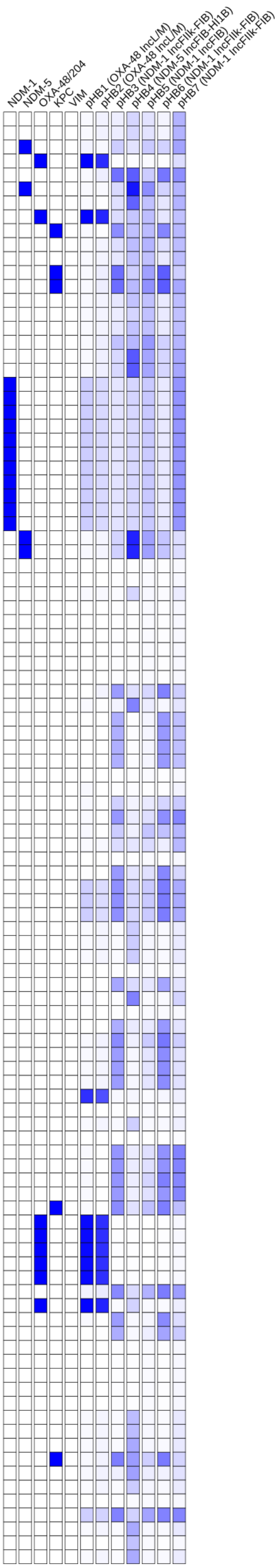

### Tree 13: GCA_043861645

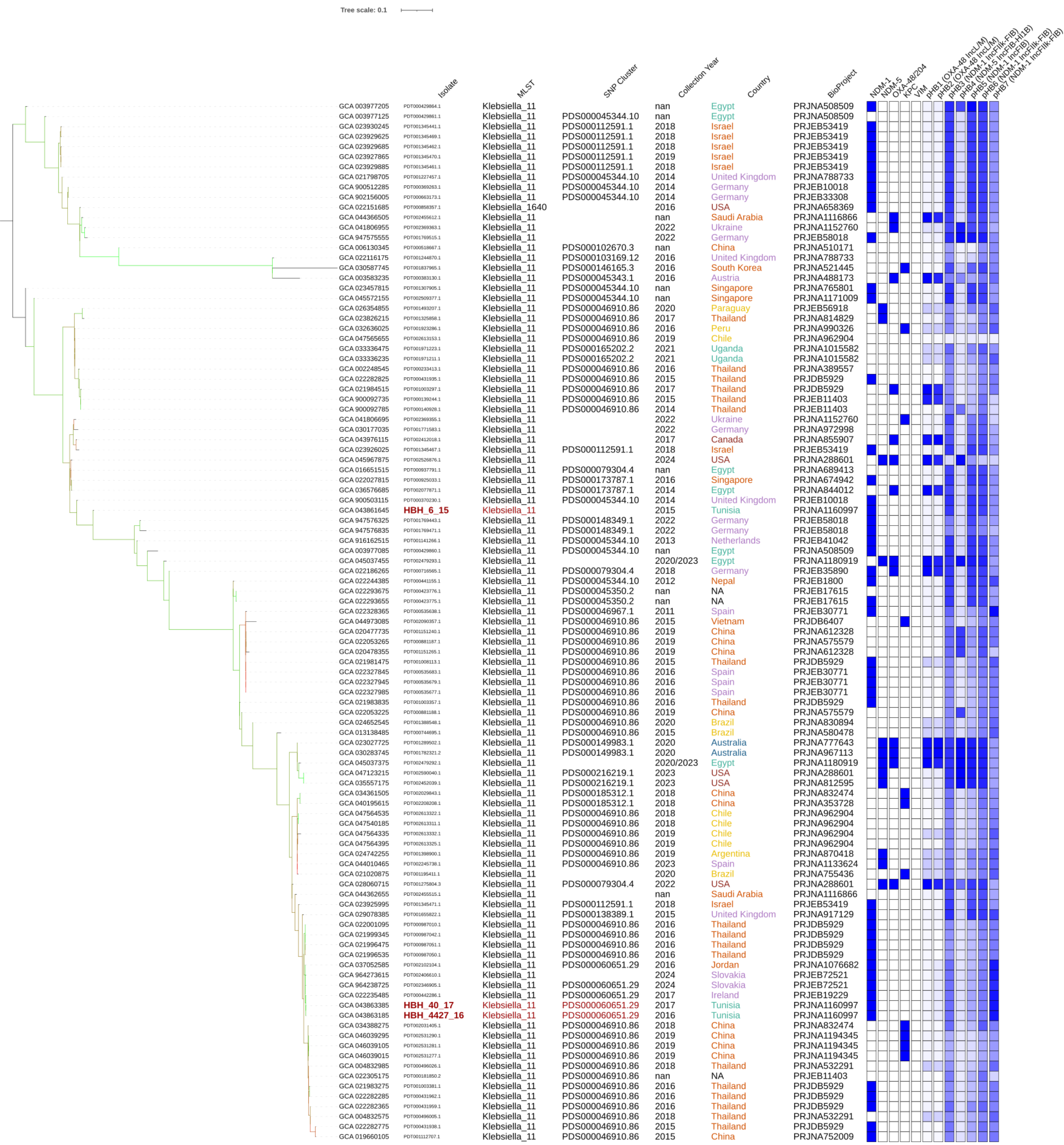

### Tree 14: GCA_043863025

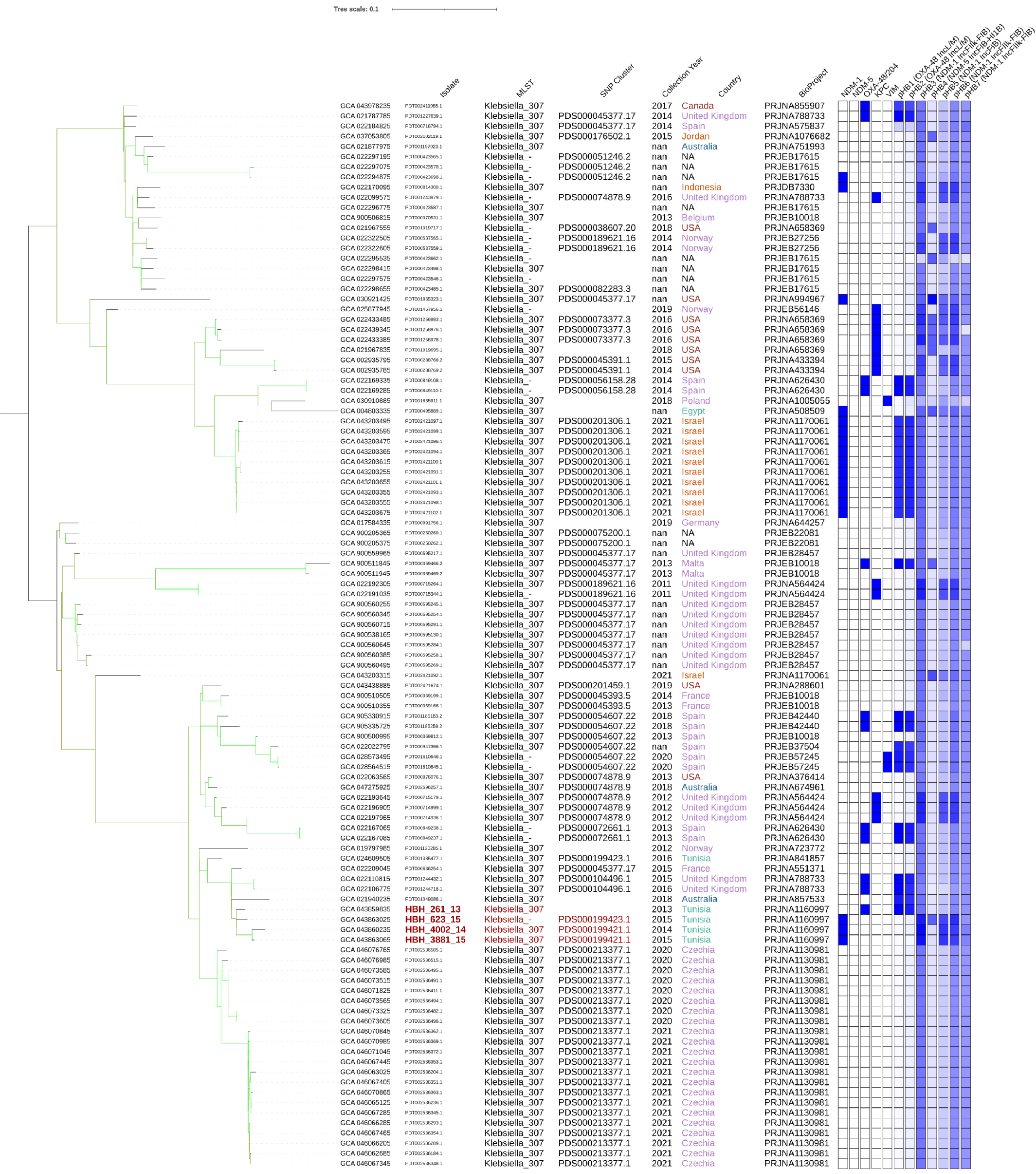

### Tree 15: GCA_043863145

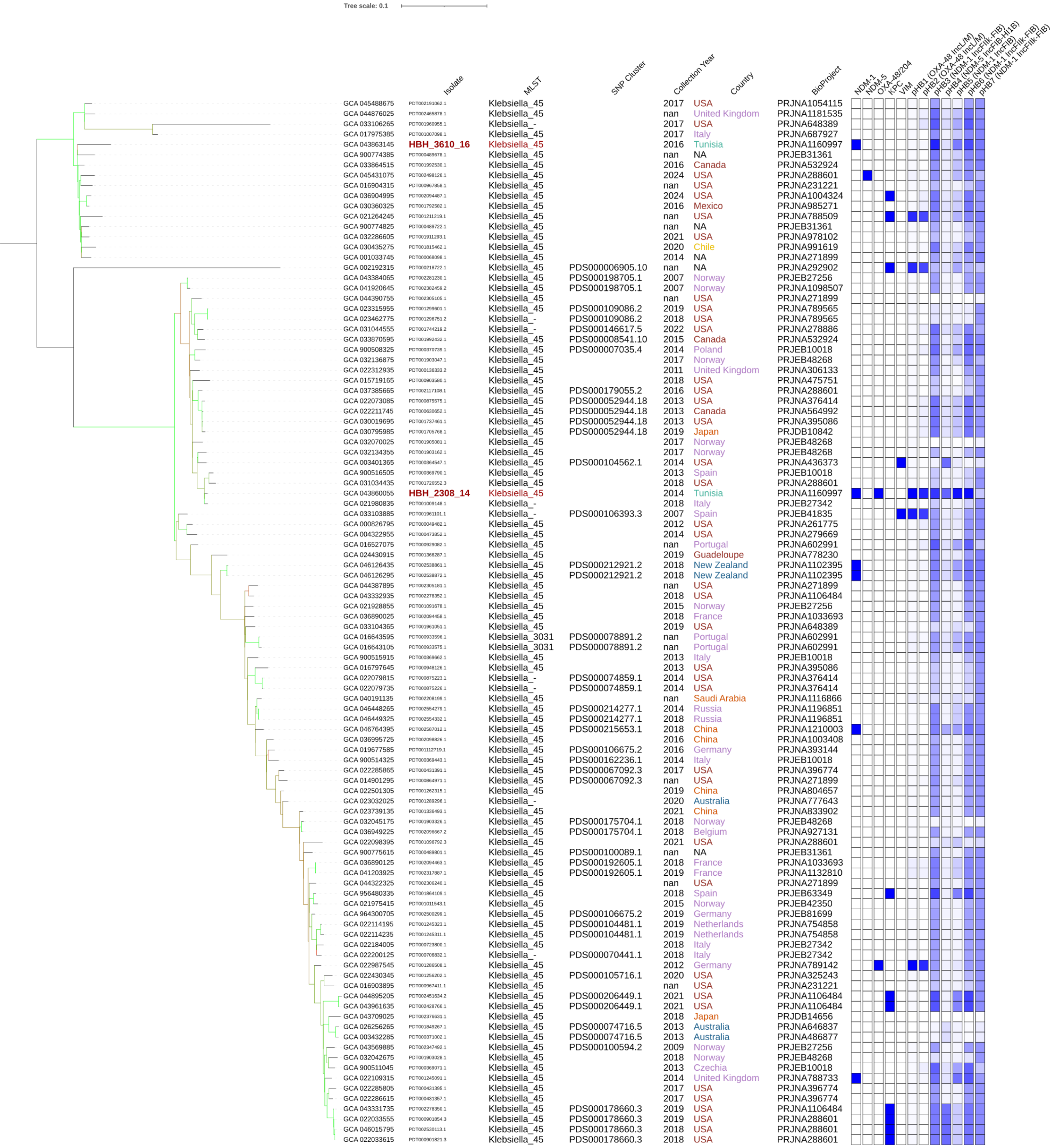

### Tree 16: GCA_043863165

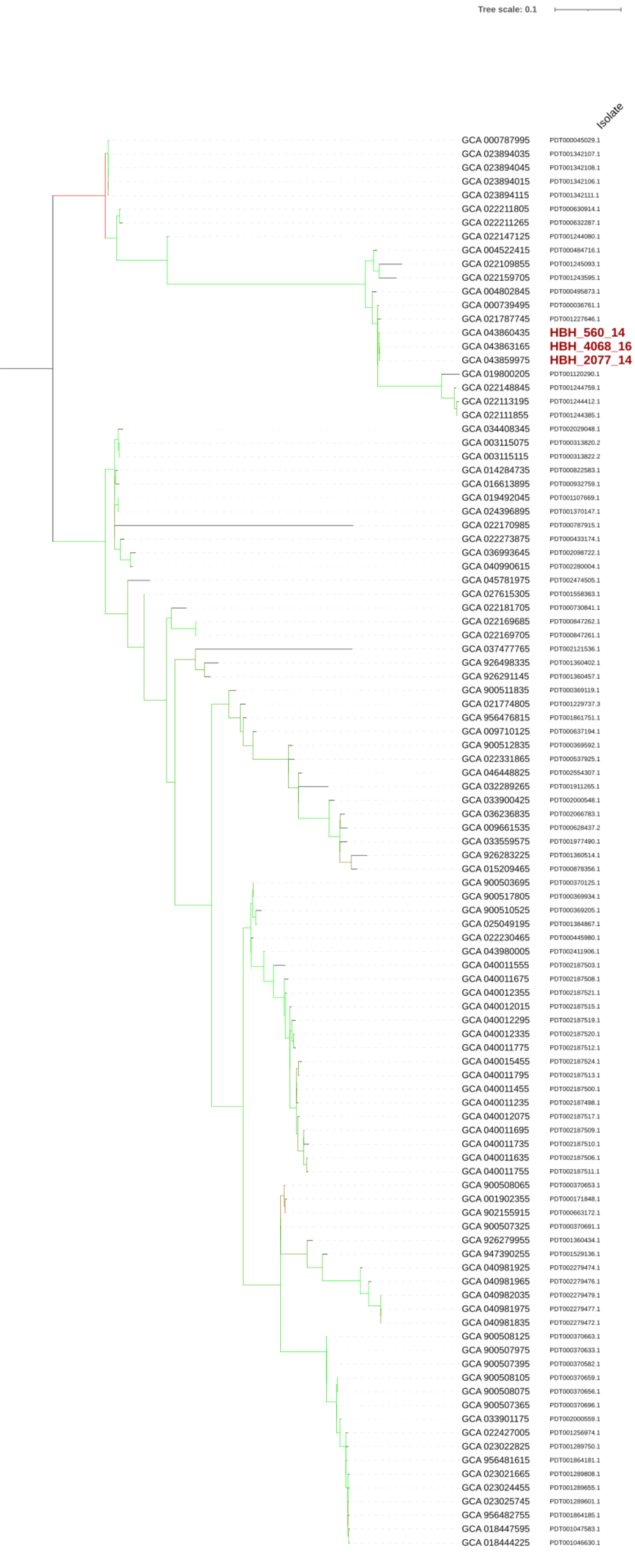


| Genome | Isolate | MLST | SNP Cluster | Collection Year | Country | BioProject |
|---|---|---|---|---|---|---|
| GCA 000787995 | | Klebsiella_395 | PDS000112540.1 | 2006 | USA | PRJNA261239 |
| GCA 023894035 | | Klebsiella_395 | PDS000112540.1 | 2006 | USA | PRJNA849970 |
| GCA 023894045 | | Klebsiella_395 | PDS000112540.1 | 2006 | USA | PRJNA849970 |
| GCA 023894015 | | Klebsiella_395 | PDS000112540.1 | 2006 | Germany | PRJNA849970 |
| GCA 023894115 | | Klebsiella_395 | PDS000112540.1 | 2006 | USA | PRJNA849970 |
| GCA 022211805 | | Klebsiella_395 | PDS000053740.1 | 2013 | Canada | PRJNA564992 |
| GCA 022211265 | | Klebsiella_395 | PDS000053740.1 | 2016 | Canada | PRJNA564992 |
| GCA 022147125 | | Klebsiella_395 | | 2015 | United Kingdom | PRJNA788733 |
| GCA 004522415 | | Klebsiella_395 | PDS000044310.6 | 2018 | Lebanon | PRJNA517127 |
| GCA 022109855 | | Klebsiella_395 | PDS000044310.6 | 2015 | United Kingdom | PRJNA788733 |
| GCA 022159705 | | Klebsiella_395 | PDS000044310.6 | 2015 | United Kingdom | PRJNA788733 |
| GCA 004802845 | | Klebsiella_395 | PDS000044310.6 | nan | Egypt | PRJNA508509 |
| GCA 000739495 | | Klebsiella_395 | PDS000044310.6 | 2012 | USA | PRJNA248655 |
| GCA 021787745 | | Klebsiella_395 | PDS000044310.6 | 2014 | United Kingdom | PRJNA788733 |
| GCA 043860435 | **HBH_560_14** | Klebsiella_395 | PDS000044310.6 | 2014 | Tunisia | PRJNA1160997 |
| GCA 043863165 | **HBH_4068_16** | Klebsiella_395 | PDS000044310.6 | 2016 | Tunisia | PRJNA1160997 |
| GCA 043859975 | **HBH_2077_14** | Klebsiella_395 | PDS000044310.6 | 2014 | Tunisia | PRJNA1160997 |
| GCA 019800205 | | Klebsiella_395 | | nan | Germany | PRJNA546126 |
| GCA 022148845 | | Klebsiella_395 | PDS000044310.6 | 2015 | United Kingdom | PRJNA788733 |
| GCA 022113195 | | Klebsiella_395 | PDS000044310.6 | 2015 | United Kingdom | PRJNA788733 |
| GCA 022111855 | | Klebsiella_395 | PDS000044310.6 | 2015 | United Kingdom | PRJNA788733 |
| GCA 034408345 | | Klebsiella_395 | | 2017 | Vietnam | PRJEB67696 |
| GCA 003115075 | | Klebsiella_395 | PDS000053786.7 | 2015 | Vietnam | PRJDB6655 |
| GCA 003115115 | | Klebsiella_395 | PDS000053786.7 | 2013 | Vietnam | PRJDB6655 |
| GCA 014284735 | | Klebsiella_395 | | 2011 | China | PRJNA657553 |
| GCA 016613895 | | Klebsiella_395 | | nan | NA | PRJNA550041 |
| GCA 019492045 | | Klebsiella_395 | PDS000114341.1 | 2013 | China | PRJNA742185 |
| GCA 024396895 | | Klebsiella_395 | PDS000114341.1 | 2013 | China | PRJNA858218 |
| GCA 022170985 | | Klebsiella_395 | | 2015 | China | PRJNA595047 |
| GCA 022273875 | | Klebsiella_395 | PDS000040454.8 | 2014 | Vietnam | PRJDB5317 |
| GCA 036993645 | | Klebsiella_395 | PDS000190144.1 | 2016 | China | PRJNA1003408 |
| GCA 040990615 | | Klebsiella_395 | PDS000190144.1 | nan | China | PRJNA1021041 |
| GCA 045781975 | | Klebsiella_395 | | 2017 | India | PRJNA939406 |
| GCA 027615305 | | Klebsiella_395 | | 2006 | Bangladesh | PRJNA845975 |
| GCA 022181705 | | Klebsiella_395 | | 2017 | Canada | PRJNA564992 |
| GCA 022169685 | | Klebsiella_395 | PDS000072337.1 | 2018 | Myanmar | PRJDB5126 |
| GCA 022169705 | | Klebsiella_395 | PDS000072337.1 | 2018 | Myanmar | PRJDB5126 |
| GCA 037477765 | | Klebsiella_395 | | 2022 | Russia | PRJNA945360 |
| GCA 926498335 | | Klebsiella_395 | | 2014 | Belarus | PRJEB49683 |
| GCA 926291145 | | Klebsiella_395 | | 2014 | Belarus | PRJEB49683 |
| GCA 900511835 | | Klebsiella_395 | PDS000077831.2 | 2014 | Estonia | PRJEB10018 |
| GCA 021774805 | | Klebsiella_395 | | 2021 | USA | PRJNA288601 |
| GCA 956476815 | | Klebsiella_395 | PDS000156671.1 | 2017 | Romania | PRJEB63349 |
| GCA 009710125 | | Klebsiella_395 | | 2018/2019 | Romania | PRJNA579879 |
| GCA 900512835 | | Klebsiella_395 | | 2013 | Italy | PRJEB10018 |
| GCA 022331865 | | Klebsiella_395 | | 2017 | Russia | PRJNA522420 |
| GCA 046448825 | | Klebsiella_395 | | 2014 | Russia | PRJNA1196851 |
| GCA 032289265 | | Klebsiella_395 | | 2021 | USA | PRJNA978102 |
| GCA 033900425 | | Klebsiella_395 | | nan | Germany | PRJNA1036598 |
| GCA 036236835 | | Klebsiella_395 | | 2022 | Russia | PRJNA729628 |
| GCA 009661535 | | Klebsiella_395 | | 2017 | Russia | PRJNA580263 |
| GCA 033559575 | | Klebsiella_395 | | 2018 | Germany | PRJEB67440 |
| GCA 926283225 | | Klebsiella_395 | | 2014 | Russia | PRJEB49683 |
| GCA 015209465 | | Klebsiella_395 | | 2013 | Russia | PRJNA269675 |
| GCA 900503695 | | Klebsiella_395 | PDS000041774.4 | 2014 | Luxembourg | PRJEB10018 |
| GCA 900517805 | | Klebsiella_395 | PDS000041774.4 | 2013 | Luxembourg | PRJEB10018 |
| GCA 900510525 | | Klebsiella_395 | PDS000041774.4 | 2014 | France | PRJEB10018 |
| GCA 025049195 | | Klebsiella_395 | PDS000041774.4 | 2012 | France | PRJEB50837 |
| GCA 022230465 | | Klebsiella_395 | PDS000041774.4 | 2011 | Canada | PRJNA390933 |
| GCA 043980005 | | Klebsiella_395 | PDS000041774.4 | 2015 | Canada | PRJNA855907 |
| GCA 040011555 | | Klebsiella_395 | PDS000045400.2 | 2011 | Netherlands | PRJNA1116077 |
| GCA 040011675 | | Klebsiella_395 | PDS000045400.2 | 2011 | Netherlands | PRJNA1116077 |
| GCA 040012355 | | Klebsiella_395 | PDS000045400.2 | 2011 | Netherlands | PRJNA1116077 |
| GCA 040012015 | | Klebsiella_395 | PDS000045400.2 | 2011 | Netherlands | PRJNA1116077 |
| GCA 040012295 | | Klebsiella_395 | PDS000045400.2 | 2014 | Netherlands | PRJNA1116077 |
| GCA 040012335 | | Klebsiella_395 | PDS000045400.2 | 2011 | Netherlands | PRJNA1116077 |
| GCA 040011775 | | Klebsiella_395 | PDS000045400.2 | 2011 | Netherlands | PRJNA1116077 |
| GCA 040015455 | | Klebsiella_395 | PDS000045400.2 | 2011 | Netherlands | PRJNA1116077 |
| GCA 040011795 | | Klebsiella_395 | PDS000045400.2 | 2011 | Netherlands | PRJNA1116077 |
| GCA 040011455 | | Klebsiella_395 | PDS000045400.2 | 2011 | Netherlands | PRJNA1116077 |
| GCA 040011235 | | Klebsiella_395 | PDS000045400.2 | 2011 | Netherlands | PRJNA1116077 |
| GCA 040012075 | | Klebsiella_395 | PDS000045400.2 | 2011 | Netherlands | PRJNA1116077 |
| GCA 040011695 | | Klebsiella_395 | PDS000045400.2 | 2011 | Netherlands | PRJNA1116077 |
| GCA 040011735 | | Klebsiella_395 | PDS000045400.2 | 2011 | Netherlands | PRJNA1116077 |
| GCA 040011635 | | Klebsiella_395 | PDS000045400.2 | 2011 | Netherlands | PRJNA1116077 |
| GCA 040011755 | | Klebsiella_395 | PDS000045400.2 | 2011 | Netherlands | PRJNA1116077 |
| GCA 900508065 | | Klebsiella_395 | PDS000036210.15 | 2014 | Turkey | PRJEB10018 |
| GCA 001902355 | | Klebsiella_395 | PDS000036210.15 | 2015 | Germany | PRJNA329409 |
| GCA 902155915 | | Klebsiella_395 | PDS000036210.15 | 2014 | Turkey | PRJEB33308 |
| GCA 900507325 | | Klebsiella_395 | PDS000036210.15 | 2013 | Turkey | PRJEB10018 |
| GCA 926279955 | | Klebsiella_395 | PDS000036210.15 | 2016 | Russia | PRJEB49683 |
| GCA 947390255 | | Klebsiella_395 | PDS000036210.15 | 2017 | United Kingdom | PRJEB57368 |
| GCA 040981925 | | Klebsiella_395 | PDS000095854.2 | 2020 | China | PRJNA836393 |
| GCA 040981965 | | Klebsiella_395 | PDS000095854.2 | 2021 | China | PRJNA836393 |
| GCA 040982035 | | Klebsiella_395 | PDS000095854.2 | 2020 | China | PRJNA836393 |
| GCA 040981975 | | Klebsiella_395 | PDS000095854.2 | 2020 | China | PRJNA836393 |
| GCA 040981835 | | Klebsiella_395 | PDS000095854.2 | 2020 | China | PRJNA836393 |
| GCA 900508125 | | Klebsiella_395 | PDS000036210.15 | 2014 | Turkey | PRJEB10018 |
| GCA 900507975 | | Klebsiella_395 | PDS000036210.15 | 2014 | Turkey | PRJEB10018 |
| GCA 900507395 | | Klebsiella_395 | PDS000036210.15 | 2013 | Turkey | PRJEB10018 |
| GCA 900508105 | | Klebsiella_395 | PDS000036210.15 | 2014 | Turkey | PRJEB10018 |
| GCA 900508075 | | Klebsiella_395 | PDS000036210.15 | 2014 | Turkey | PRJEB10018 |
| GCA 900507365 | | Klebsiella_395 | PDS000036210.15 | 2014 | Turkey | PRJEB10018 |
| GCA 033901175 | | Klebsiella_395 | PDS000036210.15 | nan | Germany | PRJNA1036598 |
| GCA 022427005 | | Klebsiella_395 | PDS000036210.15 | 2015 | Turkey | PRJNA809152 |
| GCA 023022825 | | Klebsiella_395 | PDS000036210.15 | 2018 | Turkey | PRJNA789336 |
| GCA 956481615 | | Klebsiella_395 | PDS000036210.15 | 2018 | Turkey | PRJEB63349 |
| GCA 023021665 | | Klebsiella_395 | PDS000036210.15 | 2019 | Turkey | PRJNA789336 |
| GCA 023024455 | | Klebsiella_395 | PDS000036210.15 | 2019 | Turkey | PRJNA789336 |
| GCA 023025745 | | Klebsiella_395 | PDS000036210.15 | 2019 | Turkey | PRJNA789336 |
| GCA 956482755 | | Klebsiella_395 | PDS000036210.15 | 2018 | Turkey | PRJEB63349 |
| GCA 018447595 | | Klebsiella_395 | PDS000036210.15 | 2018 | Germany | PRJNA692829 |
| GCA 018444225 | | Klebsiella_395 | PDS000036210.15 | 2018 | Germany | PRJNA692829 |

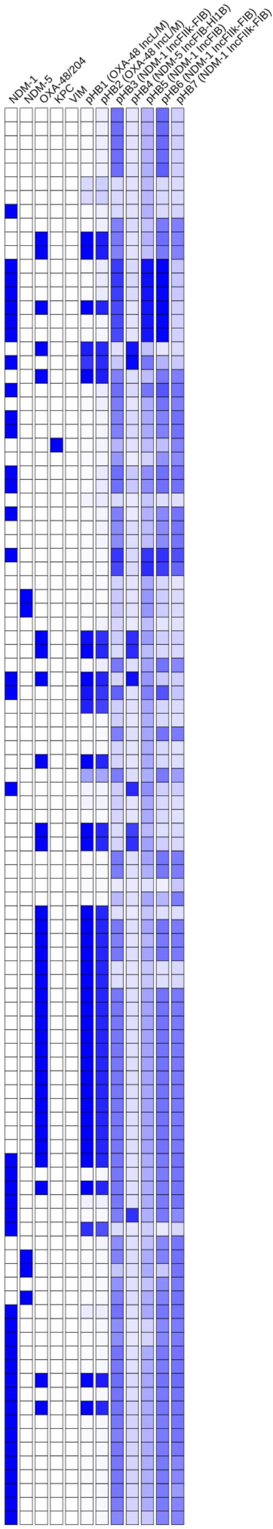

### Tree 17: GCA_043863205

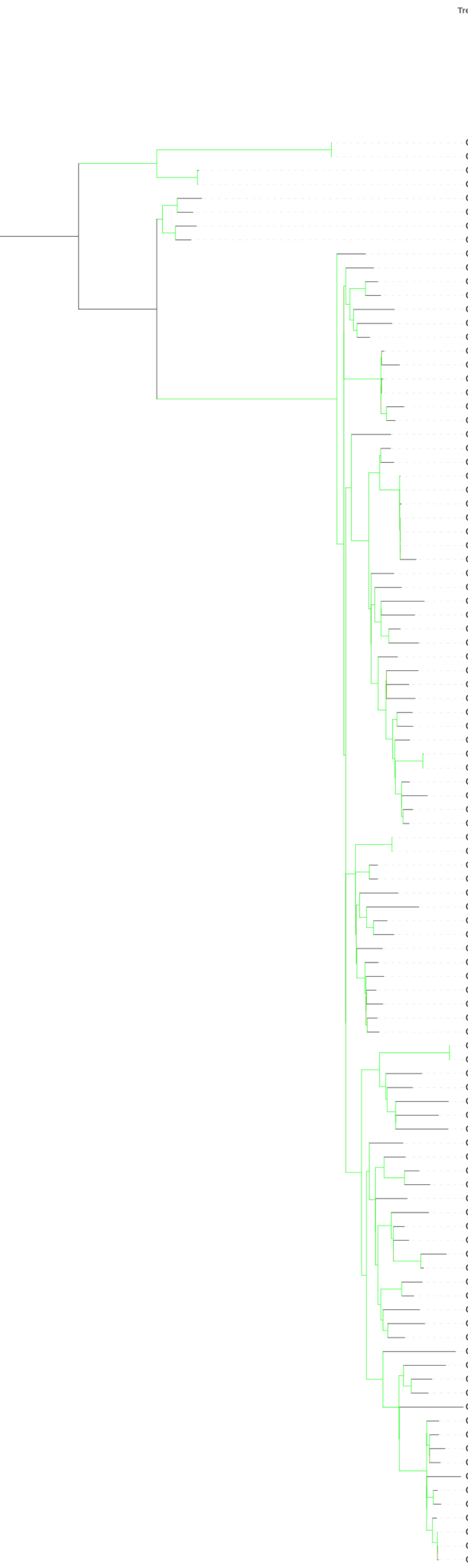


| Genome | Isolate | MLST | SNP Cluster | Collection Year | Country | BioProject |
|---|---|---|---|---|---|---|
| GCA 043861595 | **HBH_1697_10** | Klebsiella_218 | PDS000199420.1 | 2010 | Tunisia | PRJNA1160997 |
| GCA 043863205 | **HBH_1966_10** | Klebsiella_218 | PDS000199420.1 | 2010 | Tunisia | PRJNA1160997 |
| GCA 025095775 | PDT001411130.1 | Klebsiella_- | PDS000117560.1 | 2020 | Singapore | PRJNA577535 |
| GCA 025116725 | PDT001411119.1 | Klebsiella_- | PDS000117560.1 | 2020 | Singapore | PRJNA577535 |
| GCA 032132235 | PDT001905376.1 | Klebsiella_218 | | 2020 | Japan | PRJDB16654 |
| GCA 025754235 | PDT001462439.1 | Klebsiella_218 | | 2012 | China | PRJNA742521 |
| GCA 031064275 | PDT001703578.1 | Klebsiella_218 | | 2019 | Japan | PRJDB10842 |
| GCA 021871995 | PDT001198369.1 | Klebsiella_218 | | 2015 | Japan | PRJDB12075 |
| GCA 022270395 | PDT000433770.1 | Klebsiella_- | | 2015 | Singapore | PRJNA398288 |
| GCA 026192115 | PDT001488393.1 | Klebsiella_218 | | 2020 | India | PRJNA663753 |
| GCA 046764615 | PDT002587022.1 | Klebsiella_218 | | 2017 | China | PRJNA1210003 |
| GCA 037006065 | PDT002098866.1 | Klebsiella_218 | | 2020 | China | PRJNA957912 |
| GCA 030780955 | PDT001706344.1 | Klebsiella_218 | | 2020 | Japan | PRJDB10842 |
| GCA 004312025 | PDT000471471.1 | Klebsiella_218 | | 2014 | Japan | PRJDB7163 |
| GCA 046579675 | PDT002560807.1 | Klebsiella_218 | | 2022 | Germany | PRJNA1001661 |
| GCA 015209475 | PDT000878357.1 | Klebsiella_218 | PDS000074977.2 | 2013 | Russia | PRJNA269675 |
| GCA 015209425 | PDT000878354.1 | Klebsiella_218 | | 2013 | Russia | PRJNA269675 |
| GCA 015209155 | PDT000878341.1 | Klebsiella_218 | PDS000074977.2 | 2013 | Russia | PRJNA269675 |
| GCA 015645745 | PDT000897308.1 | Klebsiella_218 | PDS000074977.2 | 2013 | Russia | PRJNA269675 |
| GCA 015209245 | PDT000878346.1 | Klebsiella_218 | | 2013 | Russia | PRJNA269675 |
| GCA 015209345 | PDT000878350.1 | Klebsiella_218 | PDS000074977.2 | 2013 | Russia | PRJNA269675 |
| GCA 034407125 | PDT002029117.1 | Klebsiella_218 | | 2019 | India | PRJNA1045567 |
| GCA 025754415 | PDT001462448.1 | Klebsiella_218 | | 2010 | China | PRJNA742521 |
| GCA 038376345 | PDT002142121.1 | Klebsiella_218 | | 2021 | China | PRJNA1081623 |
| GCA 021878175 | PDT001197013.1 | Klebsiella_218 | PDS000100586.2 | 2019 | China | PRJNA733872 |
| GCA 021878215 | PDT001197012.1 | Klebsiella_218 | PDS000100586.2 | 2019 | China | PRJNA733872 |
| GCA 021878315 | PDT001197006.1 | Klebsiella_218 | PDS000100586.2 | 2019 | China | PRJNA733872 |
| GCA 030503775 | PDT001821944.1 | Klebsiella_218 | PDS000100586.2 | 2019 | China | PRJNA732824 |
| GCA 021878275 | PDT001197007.1 | Klebsiella_218 | PDS000100586.2 | 2019 | China | PRJNA733872 |
| GCA 021878235 | PDT001197009.1 | Klebsiella_218 | PDS000100586.2 | 2019 | China | PRJNA733872 |
| GCA 021878255 | PDT001197008.1 | Klebsiella_218 | PDS000100586.2 | 2019 | China | PRJNA733872 |
| GCA 036991285 | PDT002098604.1 | Klebsiella_218 | | 2015 | China | PRJNA1003408 |
| GCA 019165655 | PDT001085909.1 | Klebsiella_218 | | 2017 | China | PRJNA736556 |
| GCA 034356535 | PDT002026206.1 | Klebsiella_- | | 2011 | China | PRJNA1050414 |
| GCA 040688885 | PDT002250982.1 | Klebsiella_218 | | 2019 | China | PRJNA1131666 |
| GCA 029664705 | PDT001686926.1 | Klebsiella_218 | | 2022 | China | PRJNA937035 |
| GCA 029526275 | PDT001676541.1 | Klebsiella_218 | | 2019 | China | PRJNA810290 |
| GCA 037006045 | PDT002098865.1 | Klebsiella_218 | | 2021 | China | PRJNA957912 |
| GCA 038375335 | PDT002142037.1 | Klebsiella_218 | | 2022 | China | PRJNA1081623 |
| GCA 034740755 | PDT002042443.1 | Klebsiella_218 | | 2021 | China | PRJNA1050895 |
| GCA 037006025 | PDT002098864.1 | Klebsiella_218 | | 2020 | China | PRJNA957912 |
| GCA 025755205 | PDT001462487.1 | Klebsiella_218 | | 2015 | China | PRJNA742521 |
| GCA 044990845 | PDT002474858.1 | Klebsiella_218 | | 2020 | China | PRJNA1181777 |
| GCA 034735395 | PDT002042177.1 | Klebsiella_218 | | 2021 | China | PRJNA1050895 |
| GCA 040986405 | PDT002279631.1 | Klebsiella_218 | PDS000190124.1 | nan | China | PRJNA1021041 |
| GCA 040986275 | PDT002279625.1 | Klebsiella_218 | PDS000190124.1 | nan | China | PRJNA1021041 |
| GCA 030778505 | PDT001706386.1 | Klebsiella_218 | | 2020 | Japan | PRJDB10842 |
| GCA 030820685 | PDT001706345.1 | Klebsiella_218 | | 2020 | Japan | PRJDB10842 |
| GCA 030823355 | PDT001701964.1 | Klebsiella_- | | 2019 | Japan | PRJDB10842 |
| GCA 031064965 | PDT001703354.1 | Klebsiella_218 | | 2019 | Japan | PRJDB10842 |
| GCA 015207945 | PDT000878326.1 | Klebsiella_218 | PDS000076457.1 | 2014 | Russia | PRJNA269675 |
| GCA 015645905 | PDT000897316.1 | Klebsiella_218 | PDS000076457.1 | 2014 | Russia | PRJNA269675 |
| GCA 038376325 | PDT002142120.1 | Klebsiella_218 | | 2021 | China | PRJNA1081623 |
| GCA 025754995 | PDT001462477.1 | Klebsiella_218 | | 2015 | China | PRJNA742521 |
| GCA 022233145 | PDT000443001.1 | Klebsiella_218 | | 2011 | Vietnam | PRJEB26814 |
| GCA 033107325 | PDT001960865.1 | Klebsiella_218 | | 2018 | Vietnam | PRJEB64408 |
| GCA 037005845 | PDT002098856.1 | Klebsiella_218 | | 2022 | China | PRJNA957912 |
| GCA 038375985 | PDT002142103.1 | Klebsiella_218 | | 2021 | China | PRJNA1081623 |
| GCA 033122265 | PDT001958236.1 | Klebsiella_218 | | 2015 | Norway | PRJEB27256 |
| GCA 037005985 | PDT002098862.1 | Klebsiella_218 | | 2020 | China | PRJNA957912 |
| GCA 046467495 | PDT002556599.1 | Klebsiella_5943 | | nan | NA | PRJNA786399 |
| GCA 036994305 | PDT002098755.1 | Klebsiella_218 | | 2009 | China | PRJNA1003408 |
| GCA 025754215 | PDT001462438.1 | Klebsiella_218 | | 2012 | China | PRJNA742521 |
| GCA 004311185 | PDT000471429.1 | Klebsiella_218 | | 2014 | Japan | PRJDB7163 |
| GCA 024496205 | PDT001376407.1 | Klebsiella_218 | | 2022 | China | PRJNA861119 |
| GCA 039609835 | PDT002160632.1 | Klebsiella_218 | PDS000183511.1 | 2023 | NA | PRJNA1100710 |
| GCA 039846465 | PDT002178426.1 | Klebsiella_218 | PDS000183511.1 | 2022 | NA | PRJNA1113824 |
| GCA 037005925 | PDT002098859.1 | Klebsiella_218 | | 2022 | China | PRJNA957912 |
| GCA 034732955 | PDT002042073.1 | Klebsiella_218 | | 2020 | China | PRJNA1050895 |
| GCA 038375945 | PDT002142101.1 | Klebsiella_218 | | 2021 | China | PRJNA1081623 |
| GCA 025755105 | PDT001462482.1 | Klebsiella_218 | | 2015 | China | PRJNA742521 |
| GCA 025754615 | PDT001462459.1 | Klebsiella_218 | | 2016 | China | PRJNA742521 |
| GCA 034728955 | PDT002041846.1 | Klebsiella_218 | | 2021 | China | PRJNA1050895 |
| GCA 016916715 | PDT000970766.1 | Klebsiella_218 | | 2018 | Russia | PRJNA269675 |
| GCA 046449285 | PDT002554330.1 | Klebsiella_218 | | 2014 | Russia | PRJNA1196851 |
| GCA 046448125 | PDT002554272.1 | Klebsiella_218 | | 2011 | Russia | PRJNA1196851 |
| GCA 026334715 | PDT001494308.1 | Klebsiella_218 | | 2020 | Ghana | PRJEB56918 |
| GCA 025755235 | PDT001462489.1 | Klebsiella_218 | | 2015 | China | PRJNA742521 |
| GCA 029963605 | PDT001704089.1 | Klebsiella_218 | | 2019 | Japan | PRJDB10842 |
| GCA 030390035 | PDT001808880.1 | Klebsiella_218 | | nan | USA | PRJNA788509 |
| GCA 031066625 | PDT001702973.1 | Klebsiella_218 | | 2019 | Japan | PRJDB10842 |
| GCA 030874805 | PDT001701706.1 | Klebsiella_218 | | 2019 | Japan | PRJDB10842 |
| GCA 023742455 | PDT001336647.1 | Klebsiella_5939 | | 2020 | China | PRJNA833902 |
| GCA 037005965 | PDT002098861.1 | Klebsiella_5939 | | 2021 | China | PRJNA957912 |
| GCA 021461305 | PDT001223095.1 | Klebsiella_218 | PDS000079882.2 | 2019 | Ethiopia | PRJNA769286 |
| GCA 034740855 | PDT002042448.1 | Klebsiella_218 | | 2021 | China | PRJNA1050895 |
| GCA 034725715 | PDT002041744.1 | Klebsiella_218 | | 2021 | China | PRJNA1050895 |
| GCA 034737075 | PDT002042260.1 | Klebsiella_218 | | 2021 | China | PRJNA1050895 |
| GCA 037005905 | PDT002098858.1 | Klebsiella_218 | | 2022 | China | PRJNA957912 |
| GCA 046623285 | PDT002566380.1 | Klebsiella_218 | | 2017 | Taiwan | PRJNA1202880 |
| GCA 021166075 | PDT001202986.1 | Klebsiella_218 | | nan | Taiwan | PRJNA549322 |
| GCA 004312805 | PDT000471505.1 | Klebsiella_218 | | 2014 | Japan | PRJDB7163 |
| GCA 046065025 | PDT002536231.1 | Klebsiella_218 | | 2021 | Czechia | PRJNA1130981 |
| GCA 039611815 | PDT002160730.1 | Klebsiella_218 | PDS000183580.1 | 2023 | NA | PRJNA1100710 |
| GCA 034739235 | PDT002042367.1 | Klebsiella_218 | | 2021 | China | PRJNA1050895 |
| GCA 037005885 | PDT002098857.1 | Klebsiella_218 | | 2022 | China | PRJNA957912 |
| GCA 034736075 | PDT002042211.1 | Klebsiella_218 | | 2021 | China | PRJNA1050895 |
| GCA 034724275 | PDT002041695.1 | Klebsiella_218 | | 2021 | China | PRJNA1050895 |
| GCA 040986225 | PDT002279623.1 | Klebsiella_218 | | nan | China | PRJNA1021041 |
| GCA 046464435 | PDT002556447.1 | Klebsiella_218 | PDS000097290.2 | nan | NA | PRJNA786399 |
| GCA 022010155 | PDT001100901.1 | Klebsiella_218 | PDS000097290.2 | 2009 | China | PRJNA323522 |
| GCA 046465555 | PDT002556502.1 | Klebsiella_218 | PDS000097290.2 | nan | NA | PRJNA786399 |
| GCA 020479465 | PDT001151320.1 | Klebsiella_218 | PDS000097290.2 | 2014 | China | PRJNA612328 |

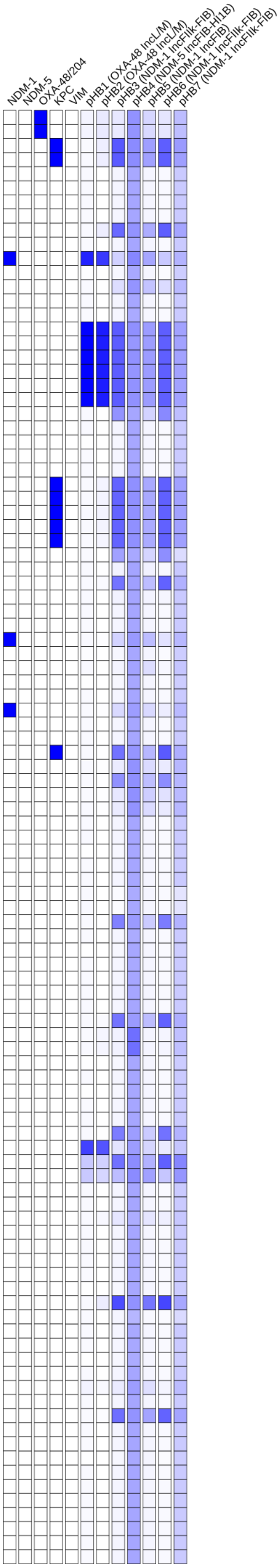

### Tree 18: GCA_043863245

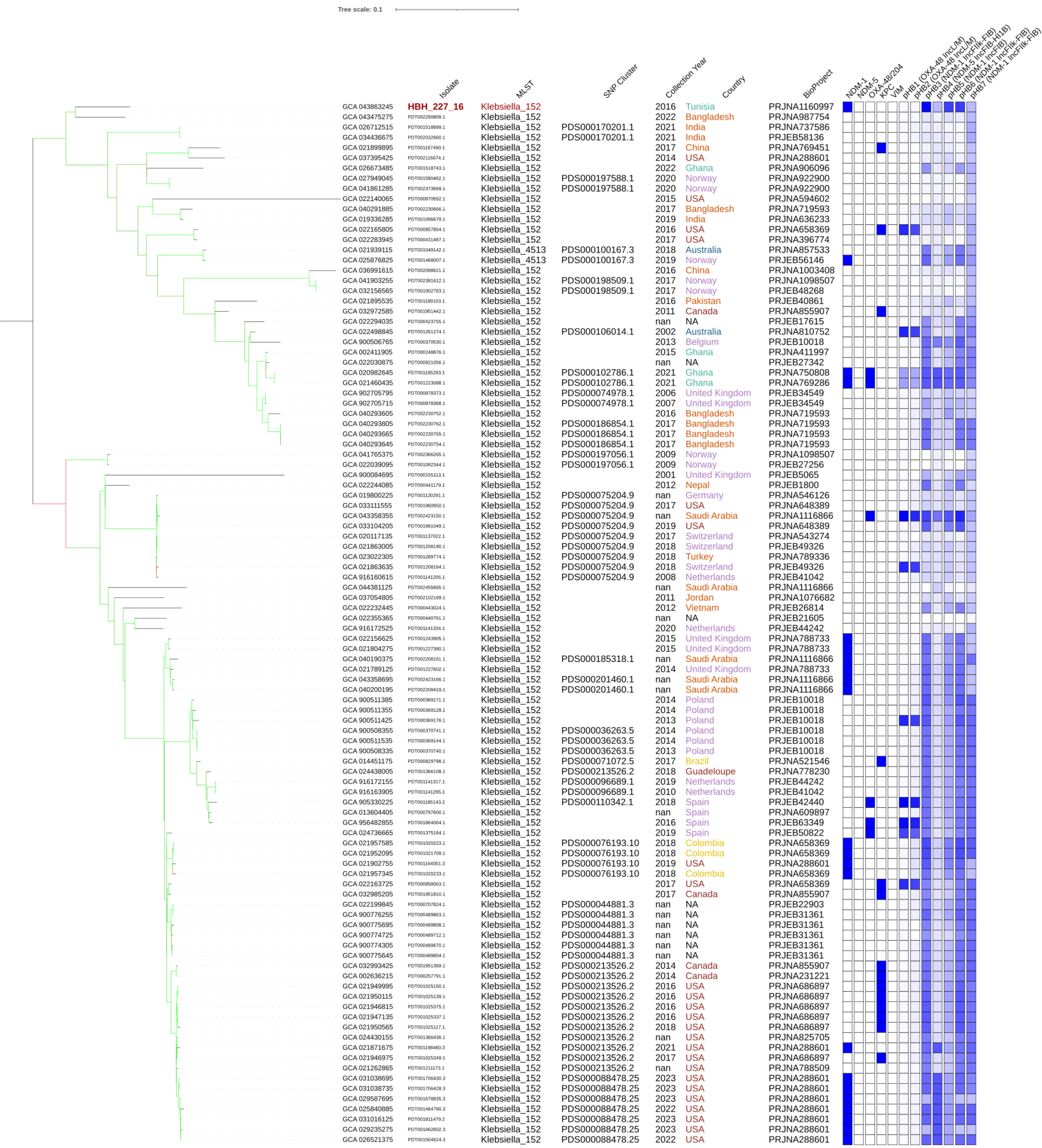

### Tree 19: GCA_043863305

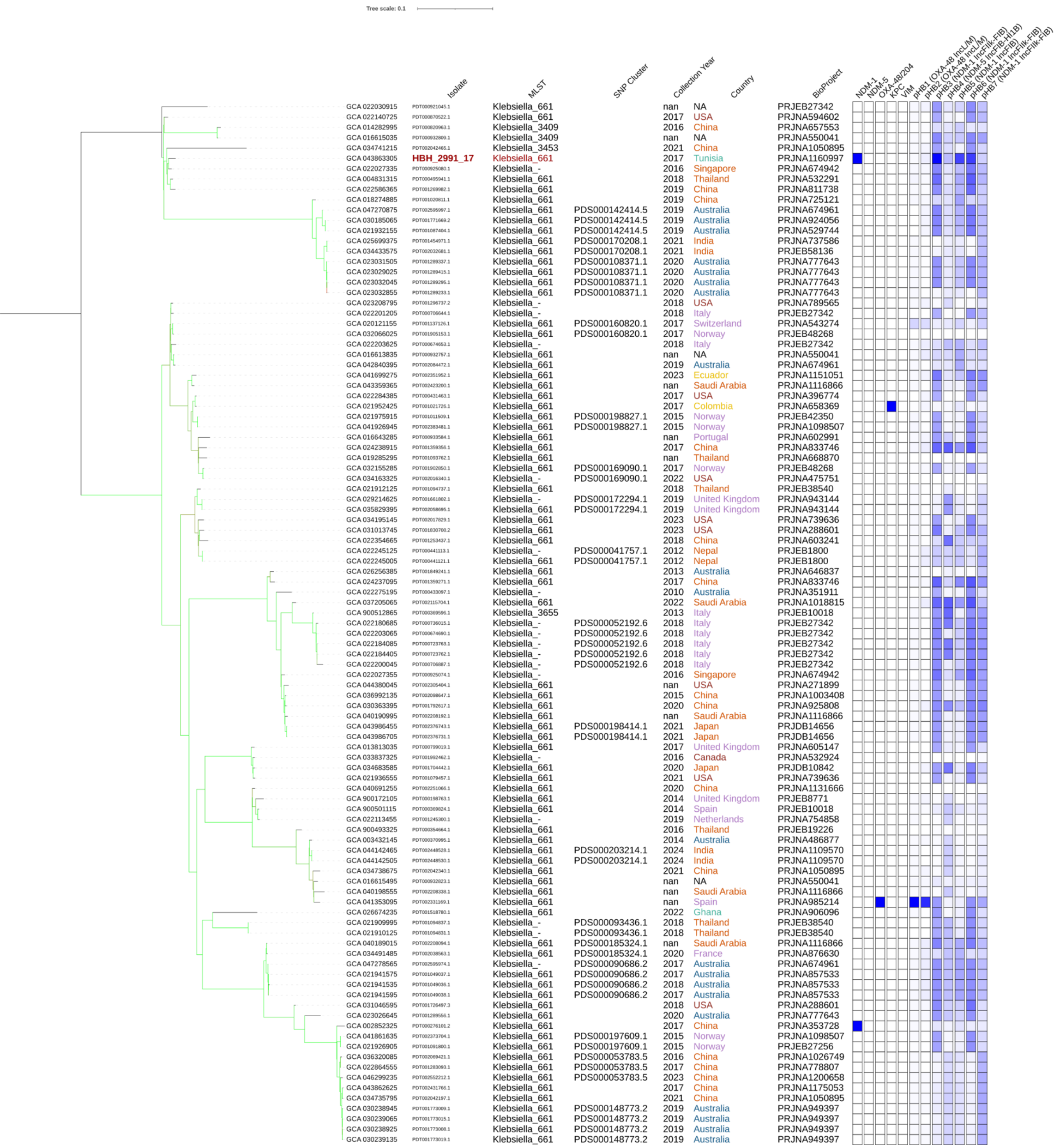

### Tree 20: GCA_043863325

Tree scale: 0.1

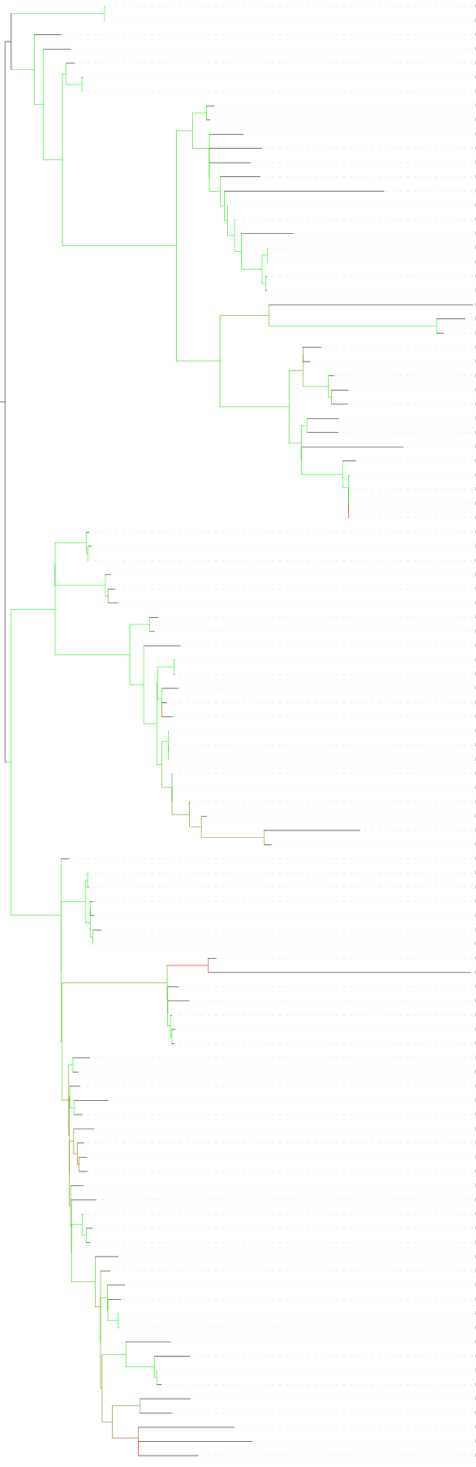

| | Isolate | MLST | SNP Cluster | Collection Year | Country | BioProject |
|---|---|---|---|---|---|---|
| GCA 022169805 | PDT000847255.1 | Klebsiella_15 | PDS000072336.1 | 2018 | Myanmar | PRJDB5126 |
| GCA 022169725 | PDT000847259.1 | Klebsiella_15 | PDS000072336.1 | 2018 | Myanmar | PRJDB5126 |
| GCA 041038315 | PDT002292032.1 | Klebsiella_15 | | nan | China | PRJNA1021041 |
| GCA 022211465 | PDT000631362.1 | Klebsiella_15 | PDS000062513.1 | 2014 | Canada | PRJNA564992 |
| GCA 037387465 | PDT002117100.1 | Klebsiella_15 | | 2014 | USA | PRJNA288601 |
| GCA 024603435 | PDT001379022.1 | Klebsiella_15 | PDS000115079.3 | 2018 | USA | PRJNA836696 |
| GCA 033104225 | PDT001961052.1 | Klebsiella_15 | PDS000115079.3 | 2019 | USA | PRJNA648389 |
| GCA 043863325 | **HBH_2648_17** | Klebsiella_15 | PDS000199428.1 | 2017 | Tunisia | PRJNA1160997 |
| GCA 043863125 | **HBH_1334_16** | Klebsiella_15 | PDS000199428.1 | 2016 | Tunisia | PRJNA1160997 |
| GCA 900513835 | PDT000369555.1 | Klebsiella_15 | PDS000036279.2 | 2014 | Montenegro | PRJEB10018 |
| GCA 012971665 | PDT000730634.1 | Klebsiella_15 | | 2017 | South Korea | PRJNA625837 |
| GCA 036236705 | PDT002066778.1 | Klebsiella_15 | | 2021 | Russia | PRJNA729628 |
| GCA 026197775 | PDT001488427.1 | Klebsiella_15 | | 2021 | China | PRJNA838135 |
| GCA 024582535 | PDT001381311.1 | Klebsiella_15 | | 2020 | China | PRJNA774095 |
| GCA 046465275 | PDT002556489.1 | Klebsiella_15 | PDS000214445.1 | nan | NA | PRJNA786399 |
| GCA 046466215 | PDT002556535.1 | Klebsiella_15 | PDS000214445.1 | nan | NA | PRJNA786399 |
| GCA 034735455 | PDT002042180.1 | Klebsiella_15 | | 2021 | China | PRJNA1050895 |
| GCA 039611895 | PDT002160734.1 | Klebsiella_15 | PDS000170844.3 | 2023 | NA | PRJNA1100710 |
| GCA 039844865 | PDT002178360.1 | Klebsiella_15 | PDS000170844.3 | 2022 | NA | PRJNA1113824 |
| GCA 034740115 | PDT002042411.1 | Klebsiella_15 | PDS000170844.3 | 2021 | China | PRJNA1050895 |
| GCA 034739755 | PDT002042393.1 | Klebsiella_15 | PDS000170844.3 | 2021 | China | PRJNA1050895 |
| GCA 030360345 | PDT001792583.1 | Klebsiella_15 | | 2016 | Mexico | PRJNA985271 |
| GCA 046531795 | PDT002559559.1 | Klebsiella_15 | | 2023 | China | PRJNA1095163 |
| GCA 025109045 | PDT001411010.1 | Klebsiella_15 | PDS000045329.17 | 2014 | Singapore | PRJNA577535 |
| GCA 900512275 | PDT000369264.1 | Klebsiella_15 | | 2014 | Germany | PRJEB10018 |
| GCA 900505905 | PDT000370325.1 | Klebsiella_15 | PDS000045329.17 | 2014 | Croatia | PRJEB10018 |
| GCA 022313055 | PDT000136320.2 | Klebsiella_15 | PDS000045329.17 | 2011 | United Kingdom | PRJNA306133 |
| GCA 022229825 | PDT000446046.1 | Klebsiella_15 | | 2015 | Germany | PRJEB18059 |
| GCA 022984665 | PDT001286497.1 | Klebsiella_15 | PDS000045329.17 | 2011 | Germany | PRJNA789142 |
| GCA 041039825 | PDT002292107.1 | Klebsiella_15 | | nan | China | PRJNA1021041 |
| GCA 022009575 | PDT000974742.1 | Klebsiella_15 | | 2018 | Australia | PRJNA857533 |
| GCA 900493705 | PDT000354582.1 | Klebsiella_15 | PDS000045331.1 | 2016 | Thailand | PRJEB19226 |
| GCA 043859485 | **HBH_545_12** | Klebsiella_15 | PDS000041726.21 | 2012 | Tunisia | PRJNA1160997 |
| GCA 022231935 | PDT000443857.1 | Klebsiella_15 | PDS000041726.21 | nan | NA | PRJEB8666 |
| GCA 022028755 | PDT000921131.1 | Klebsiella_15 | PDS000041726.21 | nan | NA | PRJEB27342 |
| GCA 022231425 | PDT000443996.1 | Klebsiella_15 | PDS000041726.21 | nan | NA | PRJEB8666 |
| GCA 022231985 | PDT000443854.1 | Klebsiella_15 | PDS000041726.21 | nan | NA | PRJEB8666 |
| GCA 916172225 | PDT001141321.1 | Klebsiella_15 | PDS000045329.17 | 2012 | Netherlands | PRJEB44242 |
| GCA 916163895 | PDT001141294.1 | Klebsiella_15 | PDS000045329.17 | 2013 | Netherlands | PRJEB41042 |
| GCA 916163945 | PDT001141299.1 | Klebsiella_15 | PDS000045329.17 | 2013 | Netherlands | PRJEB41042 |
| GCA 013620925 | PDT000797615.1 | Klebsiella_15 | PDS000066127.3 | 2012 | Portugal | PRJNA408270 |
| GCA 013521185 | PDT000797569.1 | Klebsiella_15 | PDS000066127.3 | 2012 | Portugal | PRJNA408270 |
| GCA 013620905 | PDT000797616.1 | Klebsiella_15 | PDS000066127.3 | 2013 | Portugal | PRJNA408270 |
| GCA 040493875 | PDT002239669.1 | Klebsiella_15 | PDS000124952.2 | 2021 | Brazil | PRJNA975155 |
| GCA 012241595 | PDT000721275.1 | Klebsiella_15 | PDS000124952.2 | 2019 | Brazil | PRJNA622302 |
| GCA 905231965 | PDT001185040.2 | Klebsiella_15 | PDS000045329.17 | 2018 | Spain | PRJEB42440 |
| GCA 047658805 | PDT002613000.1 | Klebsiella_15 | PDS000045329.17 | 2018 | Chile | PRJNA962904 |
| GCA 047569515 | PDT002613001.1 | Klebsiella_- | PDS000045329.17 | 2018 | Chile | PRJNA962904 |
| GCA 022171845 | PDT000785613.1 | Klebsiella_15 | | nan | Peru | PRJNA592157 |
| GCA 022181865 | PDT000730424.1 | Klebsiella_- | | 2017 | Canada | PRJNA564992 |
| GCA 047280045 | PDT002595881.1 | Klebsiella_- | | 2018 | Australia | PRJNA674961 |
| GCA 015643945 | PDT000896252.1 | Klebsiella_15 | PDS000045329.17 | 2015 | Spain | PRJNA641166 |
| GCA 022987385 | PDT001286532.1 | Klebsiella_15 | PDS000045329.17 | 2014 | Spain | PRJNA803387 |
| GCA 027866245 | PDT001562003.1 | Klebsiella_15 | PDS000045329.17 | 2018 | Spain | PRJNA803387 |
| GCA 015643785 | PDT000896244.1 | Klebsiella_15 | PDS000045329.17 | 2015 | Spain | PRJNA641166 |
| GCA 027866345 | PDT001561998.1 | Klebsiella_15 | PDS000045329.17 | 2018 | Spain | PRJNA803387 |
| GCA 022987425 | PDT001286523.1 | Klebsiella_15 | PDS000045329.17 | 2014 | Spain | PRJNA803387 |
| GCA 017595965 | PDT000992695.1 | Klebsiella_15 | PDS000045329.17 | 2017 | France | PRJNA675776 |
| GCA 047569615 | PDT002612972.1 | Klebsiella_15 | PDS000045329.17 | 2018 | Chile | PRJNA962904 |
| GCA 032635905 | PDT001923292.1 | Klebsiella_15 | PDS000045329.17 | 2017 | Peru | PRJNA990326 |
| GCA 022212805 | PDT000628216.1 | Klebsiella_15 | PDS000045329.17 | 2007 | Canada | PRJNA564992 |
| GCA 017597315 | PDT000992763.1 | Klebsiella_15 | PDS000083760.2 | 2013 | France | PRJNA675776 |
| GCA 017596835 | PDT000992739.1 | Klebsiella_15 | PDS000083760.2 | 2015 | France | PRJNA675776 |
| GCA 017596705 | PDT000992732.1 | Klebsiella_15 | PDS000083760.2 | 2015 | France | PRJNA675776 |
| GCA 017597125 | PDT000992753.1 | Klebsiella_15 | PDS000083760.2 | 2014 | France | PRJNA675776 |
| GCA 025051015 | PDT001384770.1 | Klebsiella_15 | PDS000083760.2 | 2014 | France | PRJEB50837 |
| GCA 017597025 | PDT000992748.1 | Klebsiella_15 | PDS000083760.2 | 2015 | France | PRJNA675776 |
| GCA 022297035 | PDT000423575.1 | Klebsiella_15 | | nan | NA | PRJEB17615 |
| GCA 014835065 | PDT000852579.1 | Klebsiella_15 | PDS000179049.7 | 2017 | USA | PRJNA633565 |
| GCA 900513305 | PDT000369344.1 | Klebsiella_15 | | 2013 | Italy | PRJEB10018 |
| GCA 900512715 | PDT000369312.2 | Klebsiella_15 | | 2014 | Germany | PRJEB10018 |
| GCA 900506155 | PDT000370361.1 | Klebsiella_15 | PDS000045329.17 | 2014 | Croatia | PRJEB10018 |
| GCA 022288065 | PDT000427635.1 | Klebsiella_15 | PDS000045329.17 | nan | NA | PRJEB10018 |
| GCA 022288045 | PDT000427641.1 | Klebsiella_15 | PDS000045329.17 | nan | NA | PRJEB10018 |
| GCA 022115555 | PDT001244484.1 | Klebsiella_15 | | 2014 | United Kingdom | PRJNA788733 |
| GCA 037051865 | PDT002102068.1 | Klebsiella_15 | PDS000045329.17 | 2015 | Jordan | PRJNA1076682 |
| GCA 022320665 | PDT000537649.1 | Klebsiella_15 | PDS000045329.17 | 2012 | Norway | PRJEB27256 |
| GCA 022189205 | PDT000715443.1 | Klebsiella_15 | | 2013 | United Kingdom | PRJNA564424 |
| GCA 021980855 | PDT001009146.1 | Klebsiella_15 | PDS000045329.17 | 2018 | Italy | PRJEB27342 |
| GCA 006130925 | PDT000518697.1 | Klebsiella_15 | | 2015 | China | PRJNA510171 |
| GCA 037395695 | PDT002116651.1 | Klebsiella_15 | PDS000179049.7 | 2015 | USA | PRJNA288601 |
| GCA 043301705 | PDT002265375.1 | Klebsiella_15 | PDS000179049.7 | 2018 | USA | PRJNA1106484 |
| GCA 040559185 | PDT002244896.1 | Klebsiella_15 | PDS000179049.7 | 2016 | USA | PRJNA1088550 |
| GCA 022244675 | PDT000441141.1 | Klebsiella_15 | PDS000046916.2 | 2012 | Nepal | PRJEB1800 |
| GCA 040290845 | PDT002230601.1 | Klebsiella_15 | | 2018 | Bangladesh | PRJNA719593 |
| GCA 000707985 | PDT000033733.1 | Klebsiella_15 | PDS000045329.17 | 2012 | NA | PRJNA202045 |
| GCA 014903955 | PDT000865022.1 | Klebsiella_15 | PDS000045329.17 | nan | USA | PRJNA271899 |
| GCA 014903895 | PDT000865021.1 | Klebsiella_15 | PDS000045329.17 | nan | USA | PRJNA271899 |
| GCA 012970325 | PDT000730590.1 | Klebsiella_15 | | 2016 | South Korea | PRJNA625837 |
| GCA 002866645 | PDT000277132.1 | Klebsiella_15 | PDS000045329.17 | 2013 | Lebanon | PRJNA388272 |
| GCA 022305415 | PDT000176177.2 | Klebsiella_15 | PDS000104501.4 | nan | NA | PRJEB5065 |
| GCA 022239445 | PDT000441644.1 | Klebsiella_15 | PDS000106761.1 | 2014 | Germany | PRJEB18621 |
| GCA 022248595 | PDT000440781.1 | Klebsiella_15 | PDS000075194.4 | nan | NA | PRJEB21605 |
| GCA 025881125 | PDT001467779.1 | Klebsiella_15 | PDS000075194.4 | 2015 | Norway | PRJEB56146 |
| GCA 041401685 | PDT002333248.1 | Klebsiella_15 | PDS000053782.4 | 2021 | Vietnam | PRJNA1043438 |
| GCA 040306085 | PDT002230833.1 | Klebsiella_15 | PDS000186880.1 | 2017 | Bangladesh | PRJNA719593 |
| GCA 040306005 | PDT002230829.1 | Klebsiella_15 | PDS000186880.1 | 2017 | Bangladesh | PRJNA719593 |
| GCA 040306905 | PDT002230864.1 | Klebsiella_15 | PDS000186880.1 | 2017 | Bangladesh | PRJNA719593 |
| GCA 043430835 | PDT002421569.1 | Klebsiella_15 | PDS000045329.17 | 2019 | USA | PRJNA288601 |
| GCA 034295475 | PDT002023820.1 | Klebsiella_15 | | 2023 | USA | PRJNA288601 |
| GCA 047277665 | PDT002596029.1 | Klebsiella_15 | | 2019 | Australia | PRJNA674961 |
| GCA 022182505 | PDT000729794.1 | Klebsiella_15 | | 2016 | Canada | PRJNA564992 |
| GCA 022247465 | PDT000440938.1 | Klebsiella_15 | | nan | NA | PRJEB12699 |

NDM-1, NDM-5, OXA-48/204, KPC, VIM, pHB1 (OXA-48 IncL/M), pHB2 (OXA-48 IncL/M), pHB3 (NDM-1 IncFIIk-FIB), pHB4 (NDM-5 IncFIB-HI1B), pHB5 (NDM-1 IncFIB), pHB6 (NDM-1 IncFIIk-FIB), pHB7 (NDM-1 IncFIIk-FIB)

### Tree 21: GCA_043863345

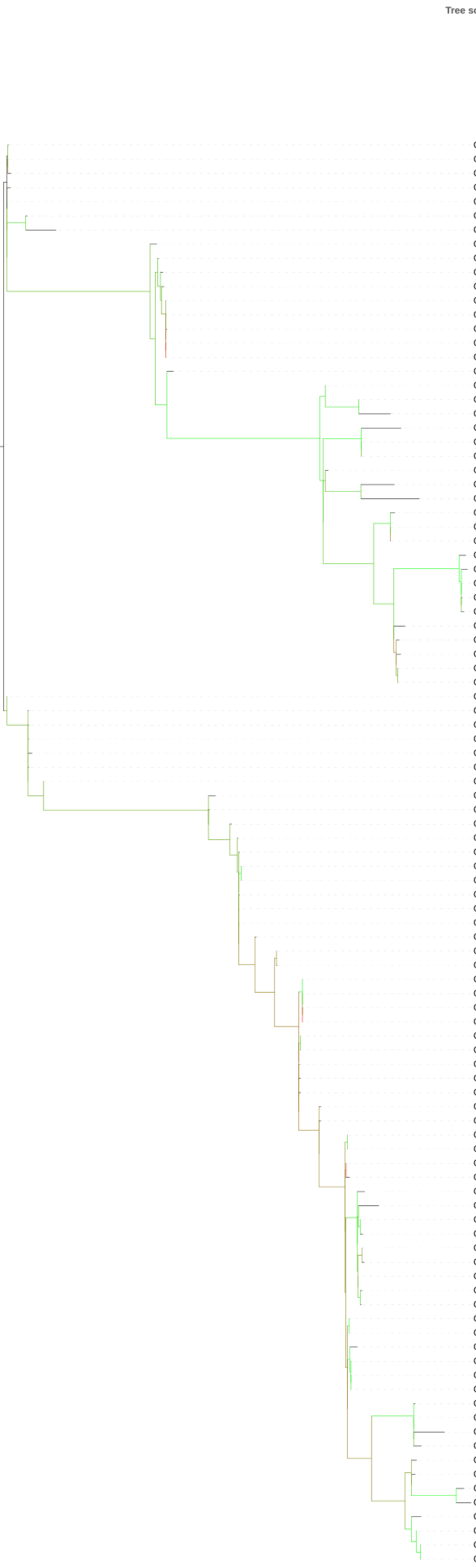


| Isolate (GCA) | Isolate | MLST | SNP Cluster | Collection Year | Country | BioProject |
|---|---|---|---|---|---|---|
| GCA 013134415 | PDT000744649.1 | Klebsiella_11 | PDS000036336.53 | 2014 | Poland | PRJNA610720 |
| GCA 013134625 | PDT000744658.1 | Klebsiella_11 | PDS000036336.53 | 2014 | Poland | PRJNA610720 |
| GCA 013134095 | PDT000744632.1 | Klebsiella_11 | PDS000036336.53 | 2014 | Poland | PRJNA610720 |
| GCA 013135335 | PDT000744694.1 | Klebsiella_11 | PDS000036336.53 | 2013 | Poland | PRJNA610720 |
| GCA 013134975 | PDT000744676.1 | Klebsiella_11 | PDS000036336.53 | 2014 | Poland | PRJNA610720 |
| GCA 013135085 | PDT000744682.1 | Klebsiella_11 | PDS000036336.53 | 2016 | Poland | PRJNA610720 |
| GCA 964215045 | PDT002343415.1 | Klebsiella_11 | PDS000113930.22 | 2023 | Slovakia | PRJEB72521 |
| GCA 013134425 | PDT000744648.1 | Klebsiella_11 | PDS000036336.53 | 2017 | Poland | PRJNA610720 |
| GCA 013135015 | PDT000744678.1 | Klebsiella_11 | PDS000036336.53 | 2016 | Poland | PRJNA610720 |
| GCA 030912725 | PDT001866003.1 | Klebsiella_11 | PDS000036336.53 | 2016 | Poland | PRJNA1005055 |
| GCA 013134715 | PDT000744663.1 | Klebsiella_11 | PDS000036336.53 | 2017 | Poland | PRJNA610720 |
| GCA 013134315 | PDT000744643.1 | Klebsiella_11 | PDS000036336.53 | 2017 | Poland | PRJNA610720 |
| GCA 013134195 | PDT000744637.1 | Klebsiella_11 | PDS000036336.53 | 2018 | Poland | PRJNA610720 |
| GCA 013134635 | PDT000744660.1 | Klebsiella_11 | | 2016 | Poland | PRJNA610720 |
| GCA 013134535 | PDT000744655.1 | Klebsiella_11 | PDS000036336.53 | 2017 | Poland | PRJNA610720 |
| GCA 013134515 | PDT000744653.1 | Klebsiella_11 | PDS000036336.53 | 2017 | Poland | PRJNA610720 |
| GCA 018107825 | PDT001010112.1 | Klebsiella_11 | PDS000072335.6 | 2015 | Malaysia | PRJNA541194 |
| GCA 002202255 | PDT000219971.1 | Klebsiella_11 | PDS000077811.10 | nan | NA | PRJNA292902 |
| GCA 021931595 | PDT001087435.1 | Klebsiella_11 | PDS000077811.10 | 2019 | Australia | PRJNA529744 |
| GCA 003115035 | PDT000313818.2 | Klebsiella_11 | PDS000077811.10 | 2016 | Vietnam | PRJDB6655 |
| GCA 003400795 | PDT000364535.1 | Klebsiella_11 | | 2015 | USA | PRJNA436373 |
| GCA 011044805 | PDT000701153.1 | Klebsiella_11 | PDS000077811.10 | 2019 | Russia | PRJNA606163 |
| GCA 011044715 | PDT000701149.1 | Klebsiella_11 | PDS000077811.10 | 2019 | Russia | PRJNA606163 |
| GCA 021932555 | PDT001087385.1 | Klebsiella_11 | PDS000077811.10 | 2019 | Australia | PRJNA529744 |
| GCA 016137005 | PDT000920683.1 | Klebsiella_11 | PDS000077811.10 | 2015 | Russia | PRJNA525009 |
| GCA 016136765 | PDT000920670.1 | Klebsiella_11 | PDS000077811.10 | 2015 | Russia | PRJNA525009 |
| GCA 021933295 | PDT001087345.1 | Klebsiella_11 | PDS000092775.2 | 2017 | Australia | PRJNA529744 |
| GCA 047270615 | PDT002596048.1 | Klebsiella_11 | PDS000092775.2 | 2017 | Australia | PRJNA674961 |
| GCA 021934475 | PDT001087281.1 | Klebsiella_11 | PDS000092775.2 | 2017 | Australia | PRJNA529744 |
| GCA 022251515 | PDT000440603.1 | Klebsiella_11 | PDS000045420.7 | 2012 | Spain | PRJEB1272 |
| GCA 021901255 | PDT001166807.1 | Klebsiella_11 | PDS000045421.52 | 2013 | Spain | PRJNA757198 |
| GCA 021900415 | PDT001166845.1 | Klebsiella_11 | PDS000045421.52 | 2014 | Spain | PRJNA757198 |
| GCA 905330575 | PDT001185164.2 | Klebsiella_11 | PDS000045421.52 | 2018 | Spain | PRJEB42440 |
| GCA 905218905 | PDT001184997.2 | Klebsiella_11 | PDS000045421.52 | 2018 | Spain | PRJEB42440 |
| GCA 022274375 | PDT000433150.1 | Klebsiella_11 | PDS000077811.10 | 2016 | Russia | PRJNA429779 |
| GCA 042910375 | PDT002091441.1 | Klebsiella_11 | PDS000077811.10 | 2014 | Vietnam | PRJDB6407 |
| GCA 003115015 | PDT000313817.2 | Klebsiella_11 | PDS000077811.10 | 2015 | Vietnam | PRJDB6655 |
| GCA 021905235 | PDT001141114.1 | Klebsiella_11 | PDS000077811.10 | 2017 | Vietnam | PRJNA641536 |
| GCA 021907035 | PDT001110127.1 | Klebsiella_11 | PDS000077811.10 | 2017 | Vietnam | PRJNA641536 |
| GCA 900510265 | PDT000369137.1 | Klebsiella_3666 | PDS000036336.53 | 2013 | Poland | PRJEB10018 |
| GCA 013134105 | PDT000744634.1 | Klebsiella_11 | PDS000036336.53 | 2016 | Poland | PRJNA610720 |
| GCA 013134935 | PDT000744674.1 | Klebsiella_11 | PDS000036336.53 | 2016 | Poland | PRJNA610720 |
| GCA 013135065 | PDT000744681.1 | Klebsiella_11 | PDS000036336.53 | 2016 | Poland | PRJNA610720 |
| GCA 013133885 | PDT000744624.1 | Klebsiella_11 | | 2016 | Poland | PRJNA610720 |
| GCA 013135225 | PDT000744689.1 | Klebsiella_11 | PDS000036336.53 | 2015 | Poland | PRJNA610720 |
| GCA 013134965 | PDT000744677.1 | Klebsiella_11 | PDS000036336.53 | 2016 | Poland | PRJNA610720 |
| GCA 956476295 | PDT001861531.1 | Klebsiella_11 | PDS000036336.53 | 2018 | Greece | PRJEB63349 |
| GCA 022116275 | PDT001244504.1 | Klebsiella_11 | | 2015 | United Kingdom | PRJNA788733 |
| GCA 900500715 | PDT000369925.1 | Klebsiella_11 | PDS000036336.53 | 2014 | Greece | PRJEB10018 |
| GCA 900501515 | PDT000369995.1 | Klebsiella_11 | PDS000036336.53 | 2014 | Greece | PRJEB10018 |
| GCA 900500745 | PDT000369911.1 | Klebsiella_11 | PDS000036336.53 | 2013 | Greece | PRJEB10018 |
| GCA 947662495 | PDT001594203.1 | Klebsiella_11 | PDS000036336.53 | 2015 | Greece | PRJEB58216 |
| GCA 028090835 | PDT001585546.1 | Klebsiella_11 | PDS000036336.53 | 2015 | Greece | PRJEB58216 |
| GCA 900502385 | PDT000369994.1 | Klebsiella_11 | PDS000036336.53 | 2014 | Greece | PRJEB10018 |
| GCA 900501265 | PDT000369891.1 | Klebsiella_11 | PDS000036336.53 | 2013 | Greece | PRJEB10018 |
| GCA 900500895 | PDT000369956.1 | Klebsiella_11 | PDS000036336.53 | 2014 | Greece | PRJEB10018 |
| GCA 956476705 | PDT001861619.1 | Klebsiella_11 | PDS000036336.53 | 2018 | Greece | PRJEB63349 |
| GCA 028090975 | PDT001585537.1 | Klebsiella_11 | PDS000036336.53 | 2014 | Greece | PRJEB58216 |
| GCA 947660525 | PDT001594156.1 | Klebsiella_11 | PDS000036336.53 | 2014 | Greece | PRJEB58216 |
| GCA 947662905 | PDT001592194.1 | Klebsiella_11 | PDS000036336.53 | 2016 | Greece | PRJEB58216 |
| GCA 947660195 | PDT001592186.1 | Klebsiella_11 | PDS000036336.53 | 2016 | Greece | PRJEB58216 |
| GCA 028090395 | PDT001585577.1 | Klebsiella_11 | PDS000036336.53 | 2016 | Greece | PRJEB58216 |
| GCA 028090015 | PDT001585568.1 | Klebsiella_11 | PDS000036336.53 | 2016 | Greece | PRJEB58216 |
| GCA 947660675 | PDT001592150.1 | Klebsiella_11 | PDS000036336.53 | 2015 | Greece | PRJEB58216 |
| GCA 028090695 | PDT001585552.1 | Klebsiella_11 | PDS000036336.53 | 2015 | Greece | PRJEB58216 |
| GCA 900501545 | PDT000369901.1 | Klebsiella_11 | PDS000036336.53 | 2013 | Greece | PRJEB10018 |
| GCA 900502325 | PDT000369993.1 | Klebsiella_11 | PDS000036336.53 | 2014 | Greece | PRJEB10018 |
| GCA 022378115 | PDT001243323.1 | Klebsiella_11 | PDS000036336.53 | 2015 | United Kingdom | PRJNA788733 |
| GCA 900500735 | PDT000369922.1 | Klebsiella_11 | PDS000036336.53 | 2014 | Greece | PRJEB10018 |
| GCA 956480625 | PDT001865593.1 | Klebsiella_11 | PDS000036336.53 | 2018 | Serbia | PRJEB63349 |
| GCA 947660585 | PDT001594174.1 | Klebsiella_11 | PDS000036336.53 | 2013 | Greece | PRJEB58216 |
| GCA 028091255 | PDT001585523.1 | Klebsiella_11 | PDS000036336.53 | 2013 | Greece | PRJEB58216 |
| GCA 902155945 | PDT000663177.1 | Klebsiella_3666 | PDS000036336.53 | 2013 | Poland | PRJEB33308 |
| GCA 013134365 | PDT000744646.1 | Klebsiella_11 | PDS000036336.53 | 2014 | Poland | PRJNA610720 |
| GCA 900516945 | PDT000369689.1 | Klebsiella_11 | PDS000036336.53 | 2014 | Bulgaria | PRJEB10018 |
| GCA 028089515 | PDT001585613.1 | Klebsiella_11 | PDS000036336.53 | 2018 | Greece | PRJEB58216 |
| GCA 028090035 | PDT001585602.1 | Klebsiella_11 | PDS000036336.53 | 2017 | Greece | PRJEB58216 |
| GCA 956476755 | PDT001861545.1 | Klebsiella_11 | PDS000036336.53 | 2017 | Greece | PRJEB63349 |
| GCA 947662925 | PDT001592218.1 | Klebsiella_11 | PDS000156641.12 | 2017 | Greece | PRJEB58216 |
| GCA 956472565 | PDT001861494.1 | Klebsiella_11 | PDS000156641.12 | 2018 | Greece | PRJEB63349 |
| GCA 028089615 | PDT001585606.1 | Klebsiella_11 | PDS000156641.12 | 2017 | Greece | PRJEB58216 |
| GCA 956476685 | PDT001861610.1 | Klebsiella_11 | PDS000156641.12 | 2018 | Greece | PRJEB63349 |
| GCA 956479255 | PDT001861577.1 | Klebsiella_11 | PDS000156641.12 | 2018 | Greece | PRJEB63349 |
| GCA 900504905 | PDT000370216.1 | Klebsiella_11 | PDS000065468.2 | 2014 | United Kingdom | PRJEB10018 |
| GCA 022159045 | PDT001243673.1 | Klebsiella_11 | PDS000065468.2 | 2014 | United Kingdom | PRJNA788733 |
| GCA 900513195 | PDT000369334.1 | Klebsiella_11 | | 2013 | Italy | PRJEB10018 |
| GCA 002180175 | PDT000217000.1 | Klebsiella_11 | PDS000005579.7 | nan | NA | PRJNA292902 |
| GCA 022316285 | PDT000111520.2 | Klebsiella_11 | PDS000005579.7 | 2014 | USA | PRJNA312972 |
| GCA 022316195 | PDT000111521.2 | Klebsiella_11 | PDS000005579.7 | 2013 | Germany | PRJNA312972 |
| GCA 002189415 | PDT000217894.1 | Klebsiella_11 | PDS000013916.2 | 2016 | Tunisia | PRJNA360371 |
| GCA 002189385 | PDT000217893.1 | Klebsiella_11 | PDS000013916.2 | 2016 | Tunisia | PRJNA360302 |
| GCA 021965615 | PDT001019819.1 | Klebsiella_11 | | 2018 | Lebanon | PRJNA658369 |
| GCA 043863345 | **HBH_658_17** | Klebsiella_11 | PDS000013916.2 | 2017 | Tunisia | PRJNA1160997 |
| GCA 900512615 | PDT000369296.1 | Klebsiella_11 | PDS000077811.10 | 2013 | Germany | PRJEB10018 |
| GCA 004141975 | PDT000460519.1 | Klebsiella_11 | | 2012 | Canada | PRJNA481972 |
| GCA 929618885 | PDT001250889.1 | Klebsiella_11 | PDS000084821.3 | 2019 | United Kingdom | PRJEB30134 |
| GCA 042695245 | PDT002404019.1 | Klebsiella_11 | PDS000140731.18 | 2024 | USA | PRJNA288601 |
| GCA 022186305 | PDT000716558.1 | Klebsiella_11 | | 2016 | Germany | PRJEB35890 |
| GCA 021787715 | PDT001227642.1 | Klebsiella_11 | PDS000103182.2 | 2014 | United Kingdom | PRJNA788733 |
| GCA 022139805 | PDT001244322.1 | Klebsiella_11 | PDS000103182.2 | 2015 | United Kingdom | PRJNA788733 |
| GCA 022106595 | PDT001244829.1 | Klebsiella_11 | PDS000103182.2 | 2015 | United Kingdom | PRJNA788733 |

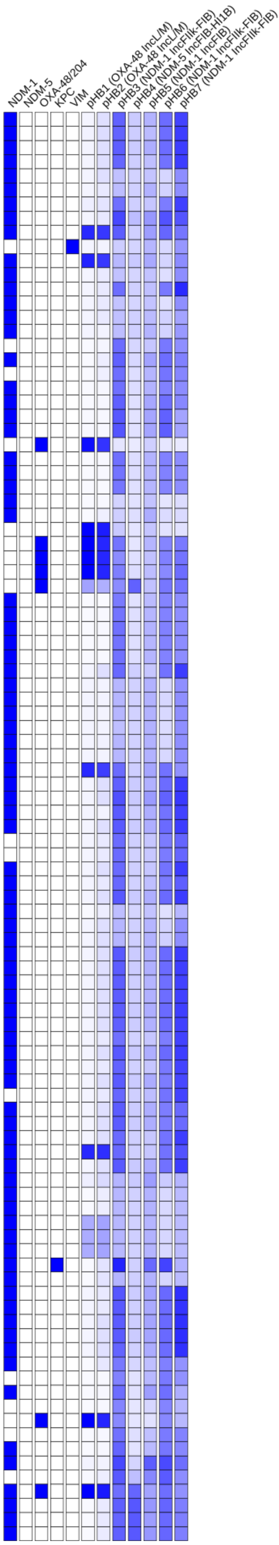

### Tree 22: GCA_043863385

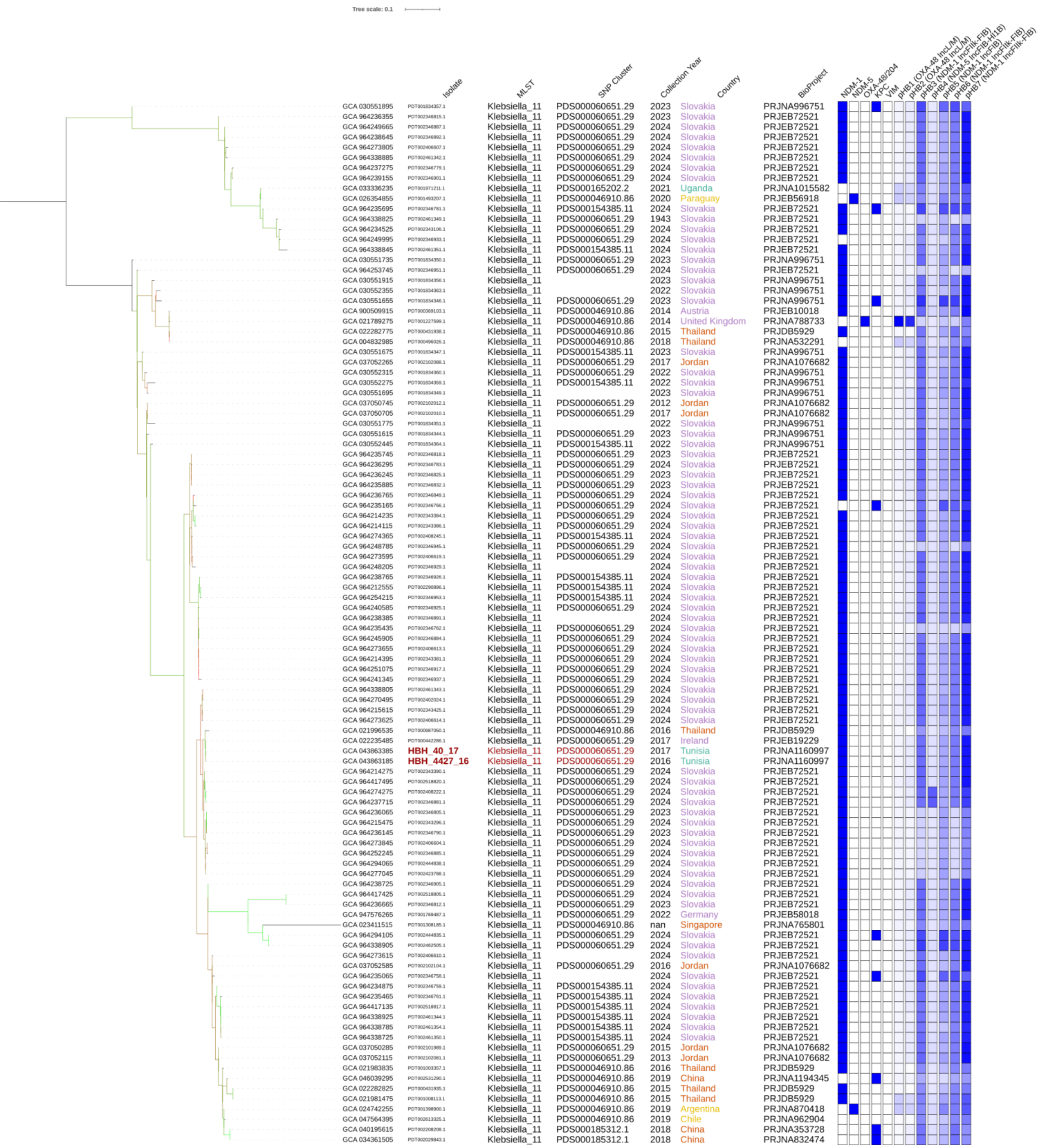

### Tree 23: GCA_043863405

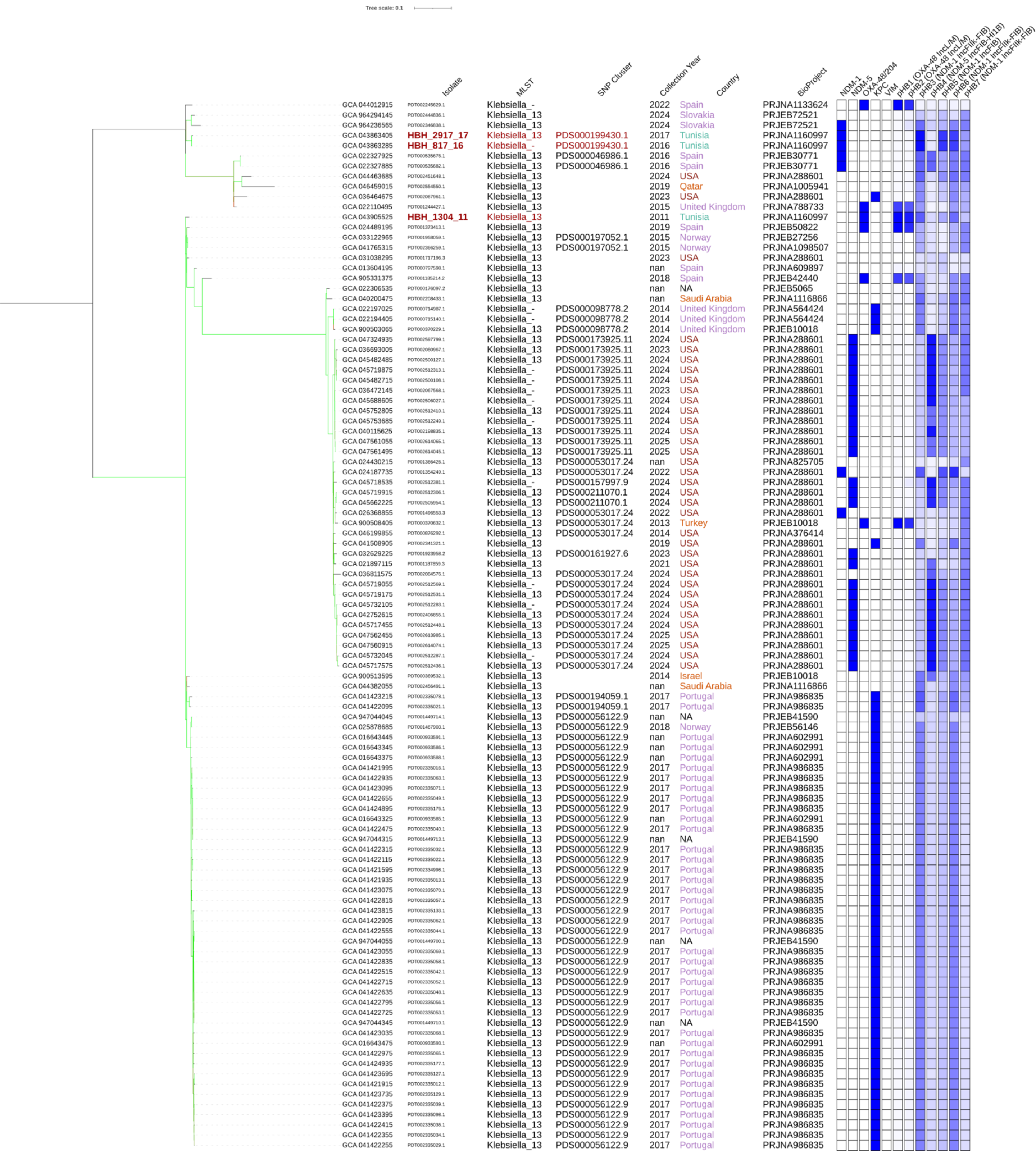

### Tree 24: GCA_043863445

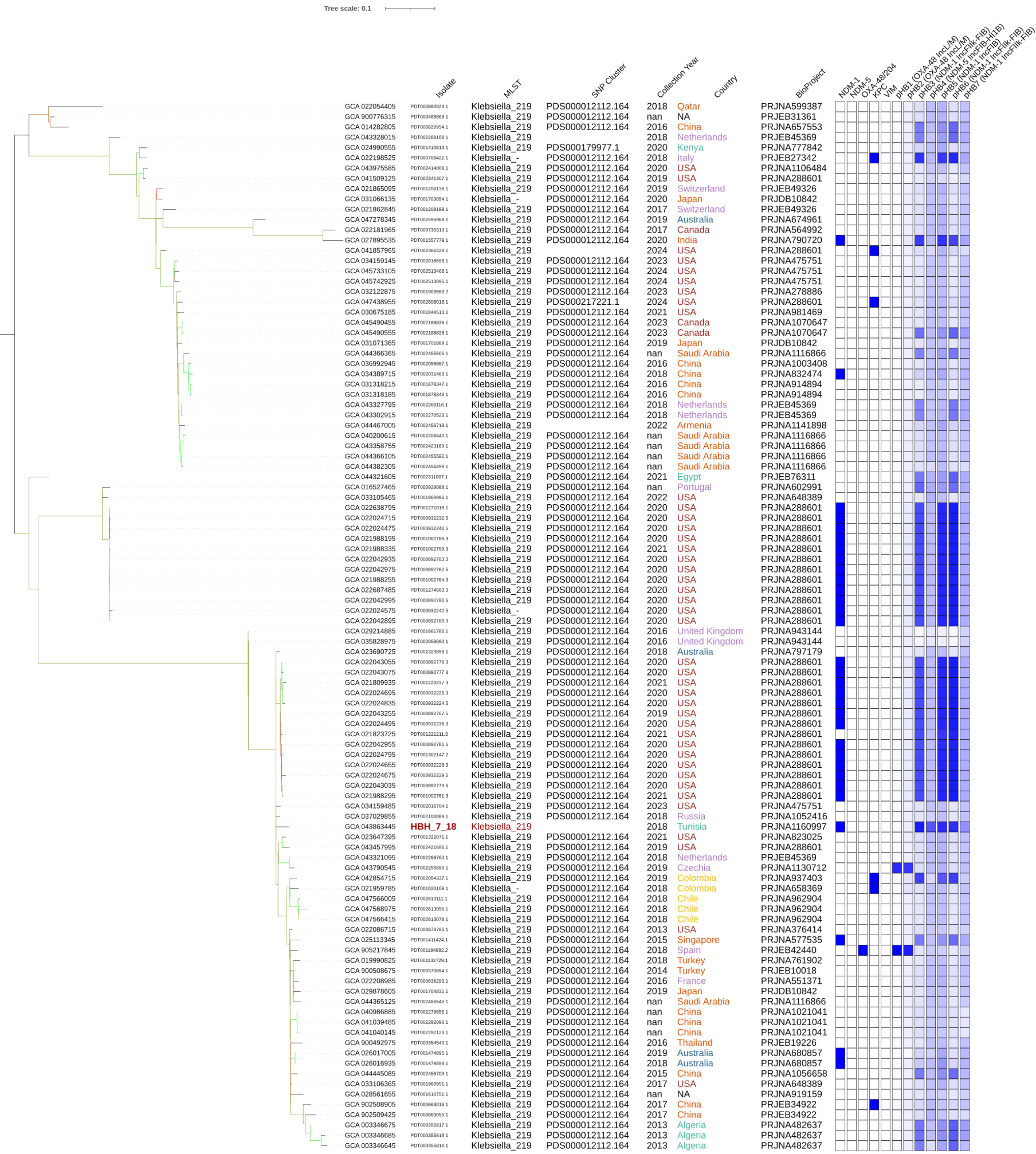

### Tree 25: GCA_043863535

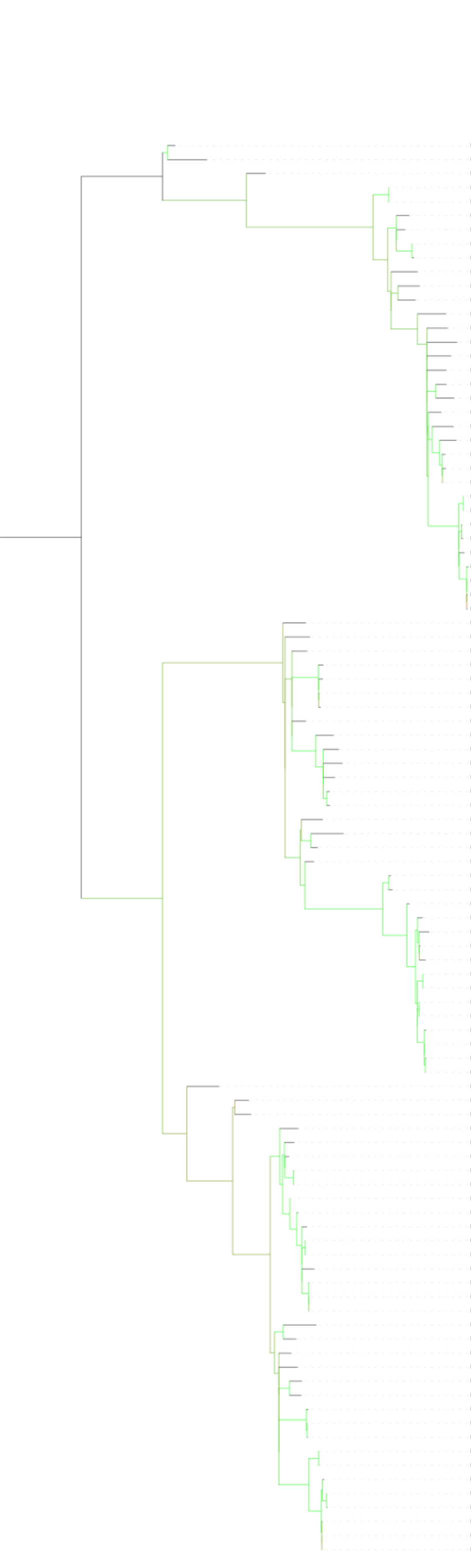


| | Isolate | MLST | SNP Cluster | Collection Year | Country | BioProject |
|---|---|---|---|---|---|---|
| GCA 016635355 | PDT000932944.1 | Klebsiella_147 | | 2015 | Australia | PRJNA480723 |
| GCA 025037385 | PDT001386114.1 | Klebsiella_147 | | 2019 | Canada | PRJNA855907 |
| GCA 031628955 | PDT001891056.1 | Klebsiella_147 | | 2021 | Thailand | PRJNA525849 |
| GCA 022212225 | PDT000630103.1 | Klebsiella_147 | PDS000053519.1 | 2010 | Canada | PRJNA564992 |
| GCA 022212665 | PDT000628739.1 | Klebsiella_147 | PDS000053519.1 | 2009 | Canada | PRJNA564992 |
| GCA 020120075 | PDT001137082.1 | Klebsiella_147 | PDS000075223.4 | 2014 | Switzerland | PRJNA543274 |
| GCA 022104055 | PDT001243382.1 | Klebsiella_147 | PDS000104526.1 | 2015 | United Kingdom | PRJNA788733 |
| GCA 902156305 | PDT000663206.1 | Klebsiella_147 | PDS000036186.4 | 2014 | Luxembourg | PRJEB33308 |
| GCA 900503645 | PDT000370117.1 | Klebsiella_147 | PDS000036186.4 | 2014 | Luxembourg | PRJEB10018 |
| GCA 900513535 | PDT000369525.1 | Klebsiella_147 | | 2013 | Israel | PRJEB10018 |
| GCA 022276345 | PDT000431990.1 | Klebsiella_147 | PDS000040359.4 | 2016 | Vietnam | PRJDB6407 |
| GCA 900502585 | PDT000370025.1 | Klebsiella_147 | PDS000156049.11 | 2014 | Denmark | PRJEB10018 |
| GCA 012971605 | PDT000730631.1 | Klebsiella_147 | | 2016 | South Korea | PRJNA625837 |
| GCA 040191035 | PDT002208194.1 | Klebsiella_147 | | nan | Saudi Arabia | PRJNA1116866 |
| GCA 029224285 | PDT001663092.1 | Klebsiella_147 | | 2019 | China | PRJNA838419 |
| GCA 022089675 | PDT000874576.1 | Klebsiella_147 | PDS000074889.1 | 2012 | USA | PRJNA376414 |
| GCA 032134505 | PDT001903152.1 | Klebsiella_147 | | 2018 | Norway | PRJEB48268 |
| GCA 022182885 | PDT000724039.1 | Klebsiella_147 | | 2018 | Italy | PRJEB27342 |
| GCA 916161875 | PDT001141251.1 | Klebsiella_147 | | 2013 | Netherlands | PRJEB41042 |
| GCA 022306355 | PDT000176139.2 | Klebsiella_147 | PDS000006642.4 | nan | NA | PRJEB5065 |
| GCA 031624155 | PDT001891027.1 | Klebsiella_147 | | 2020 | China | PRJNA845066 |
| GCA 020579485 | PDT001161797.1 | Klebsiella_147 | PDS000097792.5 | 2014 | Spain | PRJNA718833 |
| GCA 043625685 | PDT002332973.1 | Klebsiella_147 | PDS000099727.3 | nan | Spain | PRJEB55414 |
| GCA 900517095 | PDT000369887.1 | Klebsiella_147 | PDS000099727.3 | 2014 | Spain | PRJEB10018 |
| GCA 043625465 | PDT002332979.1 | Klebsiella_147 | PDS000099727.3 | nan | Spain | PRJEB55414 |
| GCA 022001215 | PDT000987012.1 | Klebsiella_147 | PDS000083378.2 | 2016 | Thailand | PRJDB5929 |
| GCA 022001185 | PDT000987015.1 | Klebsiella_147 | PDS000083378.2 | 2016 | Thailand | PRJDB5929 |
| GCA 022282305 | PDT000431960.1 | Klebsiella_147 | PDS000083378.2 | 2016 | Thailand | PRJDB5929 |
| GCA 021983395 | PDT001003375.1 | Klebsiella_147 | PDS000083378.2 | 2016 | Thailand | PRJDB5929 |
| GCA 022005405 | PDT000986943.1 | Klebsiella_147 | PDS000083378.2 | 2015 | Thailand | PRJDB5929 |
| GCA 021985635 | PDT001003212.1 | Klebsiella_147 | PDS000084451.3 | 2015 | Thailand | PRJDB5929 |
| GCA 021987055 | PDT001003115.1 | Klebsiella_147 | PDS000084451.3 | 2015 | Thailand | PRJDB5929 |
| GCA 021985595 | PDT001003215.1 | Klebsiella_147 | PDS000084451.3 | 2015 | Thailand | PRJDB5929 |
| GCA 021982475 | PDT001008034.1 | Klebsiella_147 | PDS000084451.3 | 2015 | Thailand | PRJDB5929 |
| GCA 033550765 | PDT001979154.1 | Klebsiella_147 | PDS000165990.1 | 2017 | Germany | PRJEB67440 |
| GCA 956464875 | PDT001820175.1 | Klebsiella_147 | | 2019 | Portugal | PRJEB62836 |
| GCA 900507055 | PDT000370555.1 | Klebsiella_147 | | 2014 | Belgium | PRJEB10018 |
| GCA 916161525 | PDT001141234.1 | Klebsiella_147 | PDS000036182.26 | 2013 | Netherlands | PRJEB41042 |
| GCA 024527215 | PDT001373245.1 | Klebsiella_147 | PDS000036182.26 | 2019 | Spain | PRJEB50822 |
| GCA 900516415 | PDT000369783.1 | Klebsiella_147 | PDS000036182.26 | 2013 | Spain | PRJEB10018 |
| GCA 900501685 | PDT000369810.1 | Klebsiella_147 | PDS000036182.26 | 2014 | Spain | PRJEB10018 |
| GCA 032042535 | PDT001903163.1 | Klebsiella_147 | | 2017 | Norway | PRJEB48268 |
| GCA 022240035 | PDT000441629.1 | Klebsiella_147 | | 2011 | Germany | PRJEB18621 |
| GCA 043863535 | **HBH_2232_18** | Klebsiella_147 | | 2018 | Tunisia | PRJNA1160997 |
| GCA 034513615 | PDT002038633.1 | Klebsiella_147 | | 2019 | France | PRJNA876630 |
| GCA 022237725 | PDT000441691.1 | Klebsiella_147 | | 2012 | Germany | PRJEB18621 |
| GCA 022241025 | PDT000441623.1 | Klebsiella_147 | PDS000080300.2 | 2011 | Germany | PRJEB18621 |
| GCA 022987485 | PDT001286510.1 | Klebsiella_147 | PDS000080300.2 | 2011 | Germany | PRJNA789142 |
| GCA 022181605 | PDT000732046.1 | Klebsiella_147 | | 2017 | Canada | PRJNA564992 |
| GCA 900517655 | PDT000369912.1 | Klebsiella_147 | PDS000036295.1 | 2013 | Spain | PRJEB10018 |
| GCA 022246065 | PDT000441038.1 | Klebsiella_147 | | nan | NA | PRJEB12699 |
| GCA 900510435 | PDT000369189.1 | Klebsiella_147 | | 2014 | France | PRJEB10018 |
| GCA 027610945 | PDT001558347.1 | Klebsiella_147 | PDS000130855.1 | 2007 | Bangladesh | PRJNA845975 |
| GCA 027625855 | PDT001558113.1 | Klebsiella_147 | PDS000130855.1 | 2013 | Bangladesh | PRJNA845975 |
| GCA 027615665 | PDT001558328.1 | Klebsiella_147 | PDS000009779.14 | 2006 | Bangladesh | PRJNA845975 |
| GCA 900512445 | PDT000369279.1 | Klebsiella_147 | PDS000009779.14 | 2014 | Germany | PRJEB10018 |
| GCA 030912625 | PDT001865998.1 | Klebsiella_147 | PDS000157007.2 | 2019 | Poland | PRJNA1005055 |
| GCA 900511365 | PDT000369134.1 | Klebsiella_147 | PDS000009779.14 | 2014 | Poland | PRJEB10018 |
| GCA 900511505 | PDT000369153.1 | Klebsiella_147 | PDS000009779.14 | 2013 | Poland | PRJEB10018 |
| GCA 003856595 | PDT000414016.1 | Klebsiella_147 | PDS000045262.1 | 2009 | Sweden | PRJNA413669 |
| GCA 003181055 | PDT000323199.1 | Klebsiella_147 | PDS000045262.1 | nan | Sweden | PRJNA472712 |
| GCA 022320785 | PDT001251452.1 | Klebsiella_147 | PDS000009779.14 | nan | United Kingdom | PRJNA787440 |
| GCA 034651715 | PDT002040257.1 | Klebsiella_147 | PDS000009779.14 | nan | NA | PRJNA1017778 |
| GCA 022295635 | PDT000423657.1 | Klebsiella_147 | PDS000009779.14 | nan | NA | PRJEB17615 |
| GCA 022295855 | PDT000423646.1 | Klebsiella_147 | PDS000009779.14 | nan | NA | PRJEB17615 |
| GCA 022298095 | PDT000423521.1 | Klebsiella_147 | PDS000009779.14 | nan | NA | PRJEB17615 |
| GCA 022298135 | PDT000423515.1 | Klebsiella_147 | PDS000009779.14 | nan | NA | PRJEB17615 |
| GCA 022298835 | PDT000423475.1 | Klebsiella_147 | PDS000100389.1 | nan | NA | PRJEB17615 |
| GCA 900084235 | PDT000155118.1 | Klebsiella_147 | | 2006 | United Kingdom | PRJEB5065 |
| GCA 900517045 | PDT000369761.1 | Klebsiella_147 | | 2013 | Hungary | PRJEB10018 |
| GCA 022211825 | PDT000630648.1 | Klebsiella_147 | PDS000164093.1 | 2013 | Canada | PRJNA564992 |
| GCA 030821325 | PDT001702632.1 | Klebsiella_147 | | 2019 | Japan | PRJDB10842 |
| GCA 041003655 | PDT002281177.1 | Klebsiella_147 | | 2021 | China | PRJNA1135755 |
| GCA 956477775 | PDT001861559.1 | Klebsiella_147 | PDS000106769.6 | 2018 | Greece | PRJEB63349 |
| GCA 028089455 | PDT001585616.1 | Klebsiella_147 | PDS000106769.6 | 2018 | Greece | PRJEB58216 |
| GCA 021815755 | PDT001219543.1 | Klebsiella_147 | PDS000006578.119 | 2011 | Canada | PRJNA725227 |
| GCA 025116845 | PDT001411104.1 | Klebsiella_147 | PDS000006578.119 | 2011 | Singapore | PRJNA577535 |
| GCA 022230785 | PDT000445955.1 | Klebsiella_147 | PDS000006578.119 | 2013 | Canada | PRJNA390933 |
| GCA 046202785 | PDT000435261.1 | Klebsiella_147 | PDS000006578.119 | 2014 | Singapore | PRJNA342983 |
| GCA 029492505 | PDT001672901.1 | Klebsiella_147 | PDS000006578.119 | nan | Singapore | PRJNA765801 |
| GCA 002192375 | PDT000218724.1 | Klebsiella_147 | PDS000006578.119 | nan | NA | PRJNA292902 |
| GCA 022230605 | PDT000445964.1 | Klebsiella_147 | PDS000006578.119 | 2014 | Canada | PRJNA390933 |
| GCA 043977955 | PDT002412003.1 | Klebsiella_147 | PDS000006578.119 | 2014 | Canada | PRJNA855907 |
| GCA 022230585 | PDT000445967.1 | Klebsiella_147 | PDS000006578.119 | 2014 | Canada | PRJNA390933 |
| GCA 027827355 | PDT001558040.1 | Klebsiella_147 | PDS000100083.5 | 2020 | Bangladesh | PRJNA845975 |
| GCA 900086515 | PDT000131554.1 | Klebsiella_147 | | 2009 | United Kingdom | PRJEB5065 |
| GCA 022246125 | PDT000441036.1 | Klebsiella_- | | nan | NA | PRJEB12699 |
| GCA 022212825 | PDT000628215.1 | Klebsiella_147 | | 2008 | Canada | PRJNA564992 |
| GCA 022293415 | PDT000423788.1 | Klebsiella_147 | PDS000052092.1 | nan | NA | PRJEB17615 |
| GCA 022298575 | PDT000423490.1 | Klebsiella_147 | | nan | NA | PRJEB17615 |
| GCA 030372715 | PDT001792663.1 | Klebsiella_147 | PDS000065404.6 | 2018 | Peru | PRJNA865026 |
| GCA 032635685 | PDT001923307.1 | Klebsiella_147 | PDS000065404.6 | 2017 | Peru | PRJNA990326 |
| GCA 032636005 | PDT001923285.1 | Klebsiella_147 | PDS000065404.6 | 2017 | Peru | PRJNA990326 |
| GCA 041765295 | PDT002366257.1 | Klebsiella_147 | PDS000197072.1 | 2013 | Norway | PRJNA1098507 |
| GCA 021917705 | PDT001092378.1 | Klebsiella_147 | PDS000197072.1 | 2013 | Norway | PRJEB27256 |
| GCA 027904215 | PDT001570538.1 | Klebsiella_147 | PDS000132251.1 | nan | Spain | PRJNA918858 |
| GCA 027907115 | PDT001570542.1 | Klebsiella_147 | PDS000132251.1 | nan | Spain | PRJNA918858 |
| GCA 027907135 | PDT001570541.1 | Klebsiella_147 | PDS000132251.1 | nan | Spain | PRJNA918858 |
| GCA 027904195 | PDT001570539.1 | Klebsiella_147 | PDS000132251.1 | nan | Spain | PRJNA918858 |
| GCA 027904235 | PDT001570537.1 | Klebsiella_147 | PDS000132251.1 | nan | Spain | PRJNA918858 |
| GCA 027907155 | PDT001570540.1 | Klebsiella_147 | PDS000132251.1 | nan | Spain | PRJNA918858 |

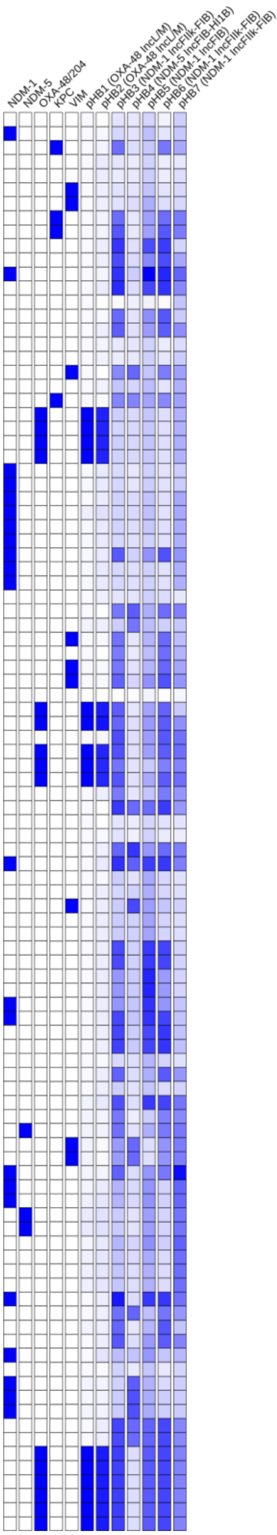

### Tree 26: GCA_043904805

Tree scale: 0.1

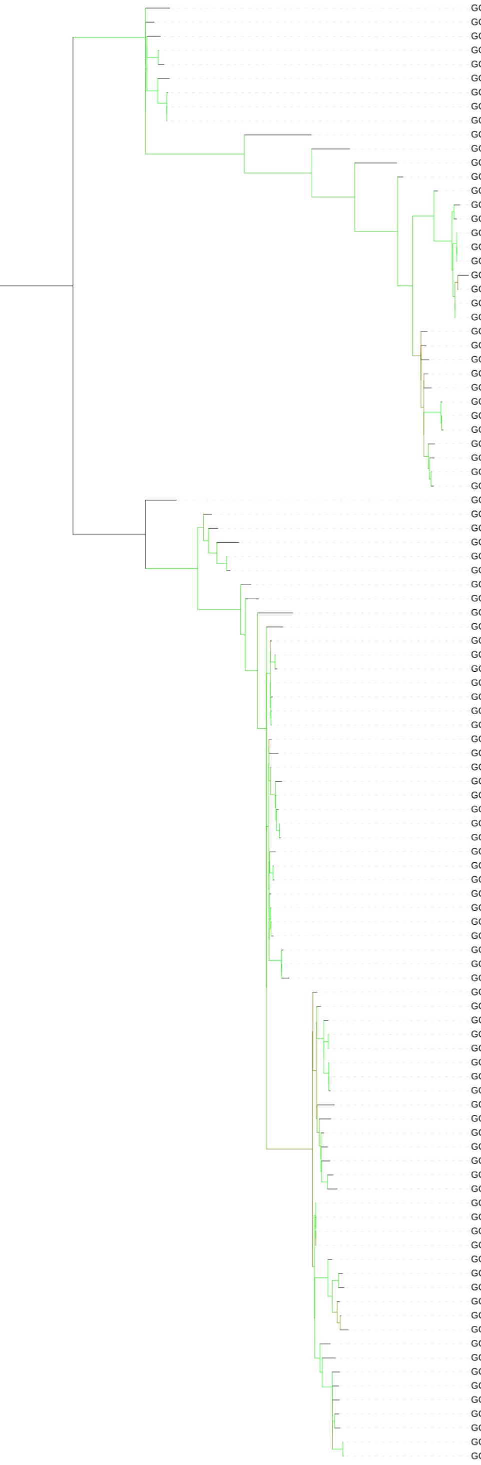

| | Isolate | MLST | SNP Cluster | Collection Year | Country | BioProject |
|---|---|---|---|---|---|---|
| GCA 900560105 | PDT000595231.1 | Klebsiella_15 | | nan | United Kingdom | PRJEB28457 |
| GCA 022247465 | PDT000440938.1 | Klebsiella_15 | | nan | NA | PRJEB12699 |
| GCA 022275625 | PDT000432631.1 | Klebsiella_15 | PDS000100772.1 | 2008 | Ireland | PRJNA396729 |
| GCA 010602475 | PDT000685977.1 | Klebsiella_15 | PDS000156654.1 | 2015 | USA | PRJNA553678 |
| GCA 010598085 | PDT000685960.1 | Klebsiella_15 | PDS000156654.1 | 2015 | USA | PRJNA553678 |
| GCA 947390215 | PDT001514280.1 | Klebsiella_15 | | 2017 | United Kingdom | PRJEB57368 |
| GCA 037107655 | PDT002109572.1 | Klebsiella_15 | PDS000184430.3 | 2024 | USA | PRJNA288601 |
| GCA 047123715 | PDT002590027.1 | Klebsiella_15 | PDS000184430.3 | 2023 | USA | PRJNA288601 |
| GCA 040107955 | PDT002192467.1 | Klebsiella_15 | PDS000184430.3 | 2024 | USA | PRJNA288601 |
| GCA 002248595 | PDT000233416.1 | Klebsiella_15 | | 2016 | Thailand | PRJNA389557 |
| GCA 022182505 | PDT000729794.1 | Klebsiella_15 | | 2016 | Canada | PRJNA564992 |
| GCA 047277665 | PDT002596029.1 | Klebsiella_15 | | 2019 | Australia | PRJNA674961 |
| GCA 002866645 | PDT000277132.1 | Klebsiella_15 | PDS000045329.17 | 2013 | Lebanon | PRJNA388272 |
| GCA 000707985 | PDT000033733.1 | Klebsiella_15 | PDS000045329.17 | 2012 | NA | PRJNA202045 |
| GCA 047280045 | PDT002595881.1 | Klebsiella_- | | 2018 | Australia | PRJNA674961 |
| GCA 022181865 | PDT000730424.1 | Klebsiella_- | | 2017 | Canada | PRJNA564992 |
| GCA 015643945 | PDT000896252.1 | Klebsiella_15 | PDS000045329.17 | 2015 | Spain | PRJNA641166 |
| GCA 022987385 | PDT001286532.1 | Klebsiella_15 | PDS000045329.17 | 2014 | Spain | PRJNA803387 |
| GCA 027866245 | PDT001562003.1 | Klebsiella_15 | PDS000045329.17 | 2018 | Spain | PRJNA803387 |
| GCA 047569615 | PDT002612972.1 | Klebsiella_15 | PDS000045329.17 | 2018 | Chile | PRJNA962904 |
| GCA 022987425 | PDT001286523.1 | Klebsiella_15 | PDS000045329.17 | 2014 | Spain | PRJNA803387 |
| GCA 015643785 | PDT000896244.1 | Klebsiella_15 | PDS000045329.17 | 2015 | Spain | PRJNA641166 |
| GCA 027866345 | PDT001561998.1 | Klebsiella_15 | PDS000045329.17 | 2018 | Spain | PRJNA803387 |
| GCA 021980855 | PDT001009146.1 | Klebsiella_15 | PDS000045329.17 | 2018 | Italy | PRJEB27342 |
| GCA 037051865 | PDT002102068.1 | Klebsiella_15 | PDS000045329.17 | 2015 | Jordan | PRJNA1076682 |
| GCA 037395695 | PDT002116651.1 | Klebsiella_15 | PDS000179049.7 | 2015 | USA | PRJNA288601 |
| GCA 022212805 | PDT000628216.1 | Klebsiella_15 | PDS000045329.17 | 2007 | Canada | PRJNA564992 |
| GCA 022244675 | PDT000441141.1 | Klebsiella_15 | PDS000046916.2 | 2012 | Nepal | PRJEB1800 |
| GCA 916172225 | PDT001141321.1 | Klebsiella_15 | PDS000045329.17 | 2012 | Netherlands | PRJEB44242 |
| GCA 916163945 | PDT001141299.1 | Klebsiella_15 | PDS000045329.17 | 2013 | Netherlands | PRJEB41042 |
| GCA 916163895 | PDT001141294.1 | Klebsiella_15 | PDS000045329.17 | 2013 | Netherlands | PRJEB41042 |
| GCA 022297035 | PDT000423575.1 | Klebsiella_15 | | nan | NA | PRJEB17615 |
| GCA 022313055 | PDT000136320.2 | Klebsiella_15 | PDS000045329.17 | 2011 | United Kingdom | PRJNA306133 |
| GCA 900506155 | PDT000370361.1 | Klebsiella_15 | PDS000045329.17 | 2014 | Croatia | PRJEB10018 |
| GCA 022288065 | PDT000427635.1 | Klebsiella_15 | PDS000045329.17 | nan | NA | PRJEB10018 |
| GCA 043859485 | **HBH_545_12** | Klebsiella_15 | PDS000041726.21 | 2012 | Tunisia | PRJNA1160997 |
| GCA 900505905 | PDT000370325.1 | Klebsiella_15 | PDS000045329.17 | 2014 | Croatia | PRJEB10018 |
| GCA 043863125 | **HBH_1334_16** | Klebsiella_15 | PDS000199428.1 | 2016 | Tunisia | PRJNA1160997 |
| GCA 036236705 | PDT002066778.1 | Klebsiella_15 | | 2021 | Russia | PRJNA729628 |
| GCA 046465275 | PDT002556489.1 | Klebsiella_15 | PDS000214445.1 | nan | NA | PRJNA786399 |
| GCA 046466215 | PDT002556535.1 | Klebsiella_15 | PDS000214445.1 | nan | NA | PRJNA786399 |
| GCA 037387465 | PDT002117100.1 | Klebsiella_15 | | 2014 | USA | PRJNA288601 |
| GCA 022258775 | PDT000437222.1 | Klebsiella_15 | | 2008 | Laos | PRJEB2111 |
| GCA 026369015 | PDT001496549.3 | Klebsiella_15 | | 2022 | USA | PRJNA288601 |
| GCA 022237205 | PDT000441700.1 | Klebsiella_15 | PDS000124938.1 | 2013 | Germany | PRJEB18621 |
| GCA 010179715 | PDT000679377.1 | Klebsiella_15 | PDS000041726.21 | 2016 | USA | PRJNA589891 |
| GCA 010179795 | PDT000679380.1 | Klebsiella_15 | PDS000041726.21 | 2015 | USA | PRJNA589891 |
| GCA 010179845 | PDT000679383.1 | Klebsiella_15 | PDS000041726.21 | 2015 | USA | PRJNA589891 |
| GCA 010179725 | PDT000679378.1 | Klebsiella_15 | PDS000041726.21 | 2014 | USA | PRJNA589891 |
| GCA 010179805 | PDT000679381.1 | Klebsiella_15 | PDS000041726.21 | 2015 | USA | PRJNA589891 |
| GCA 010179775 | PDT000679379.1 | Klebsiella_15 | PDS000041726.21 | 2016 | USA | PRJNA589891 |
| GCA 010179835 | PDT000679382.1 | Klebsiella_15 | PDS000041726.21 | 2016 | USA | PRJNA589891 |
| GCA 022076105 | PDT000875427.1 | Klebsiella_15 | PDS000041726.21 | 2012 | USA | PRJNA376414 |
| GCA 022067625 | PDT000875828.1 | Klebsiella_15 | PDS000041726.21 | 2014 | USA | PRJNA376414 |
| GCA 022089835 | PDT000874562.1 | Klebsiella_15 | PDS000041726.21 | 2013 | USA | PRJNA376414 |
| GCA 022063665 | PDT000876071.1 | Klebsiella_15 | PDS000041726.21 | 2013 | USA | PRJNA376414 |
| GCA 022071465 | PDT000875628.1 | Klebsiella_15 | PDS000041726.21 | 2013 | USA | PRJNA376414 |
| GCA 022081255 | PDT000875103.1 | Klebsiella_15 | PDS000041726.21 | 2014 | USA | PRJNA376414 |
| GCA 022067865 | PDT000875810.1 | Klebsiella_15 | PDS000041726.21 | 2014 | USA | PRJNA376414 |
| GCA 022068305 | PDT000875786.1 | Klebsiella_15 | PDS000041726.21 | 2014 | USA | PRJNA376414 |
| GCA 022064285 | PDT000876037.1 | Klebsiella_15 | PDS000041726.21 | 2013 | USA | PRJNA376414 |
| GCA 022070945 | PDT000875668.1 | Klebsiella_15 | PDS000041726.21 | 2013 | USA | PRJNA376414 |
| GCA 022070965 | PDT000875666.1 | Klebsiella_15 | PDS000041726.21 | 2013 | USA | PRJNA376414 |
| GCA 022070885 | PDT000875671.1 | Klebsiella_15 | PDS000041726.21 | 2013 | USA | PRJNA376414 |
| GCA 022072865 | PDT000875587.1 | Klebsiella_15 | PDS000041726.21 | 2013 | USA | PRJNA376414 |
| GCA 022071505 | PDT000875631.1 | Klebsiella_15 | PDS000041726.21 | 2013 | USA | PRJNA376414 |
| GCA 022072825 | PDT000875589.1 | Klebsiella_15 | PDS000041726.21 | 2013 | USA | PRJNA376414 |
| GCA 022060845 | PDT000876223.1 | Klebsiella_15 | PDS000041726.21 | 2013 | USA | PRJNA376414 |
| GCA 022061705 | PDT000876180.1 | Klebsiella_15 | PDS000041726.21 | 2015 | USA | PRJNA376414 |
| GCA 022060865 | PDT000876225.1 | Klebsiella_15 | PDS000041726.21 | 2013 | USA | PRJNA376414 |
| GCA 024918875 | PDT001409690.1 | Klebsiella_15 | PDS000041726.21 | 2016 | USA | PRJNA836696 |
| GCA 022060825 | PDT000876226.1 | Klebsiella_15 | PDS000041726.21 | 2015 | USA | PRJNA376414 |
| GCA 031038415 | PDT001716002.3 | Klebsiella_15 | | 2023 | USA | PRJNA288601 |
| GCA 021904015 | PDT001149771.1 | Klebsiella_15 | PDS000097194.4 | 2021 | USA | PRJNA288601 |
| GCA 021904075 | PDT001148880.3 | Klebsiella_15 | PDS000097194.4 | 2021 | USA | PRJNA288601 |
| GCA 040735645 | PDT002255402.1 | Klebsiella_15 | PDS000188634.2 | 2024 | USA | PRJNA288601 |
| GCA 039549035 | PDT002156713.1 | Klebsiella_15 | PDS000188634.2 | 2024 | USA | PRJNA288601 |
| GCA 041213875 | PDT002310441.1 | Klebsiella_15 | PDS000188634.2 | 2024 | USA | PRJNA288601 |
| GCA 022030895 | PDT000921052.1 | Klebsiella_15 | | nan | NA | PRJEB27342 |
| GCA 028883295 | PDT001647973.1 | Klebsiella_15 | | nan | NA | PRJNA915079 |
| GCA 043859735 | **HBH_600_13** | Klebsiella_15 | PDS000041726.21 | 2013 | Tunisia | PRJNA1160997 |
| GCA 022187525 | PDT000715769.1 | Klebsiella_15 | PDS000060613.1 | 2018 | Saudi Arabia | PRJEB36683 |
| GCA 022103415 | PDT001244308.1 | Klebsiella_15 | | 2014 | United Kingdom | PRJNA788733 |
| GCA 002189165 | PDT000217883.1 | Klebsiella_15 | | 2016 | Tunisia | PRJNA360233 |
| GCA 043904805 | **HBH_363_18** | Klebsiella_15 | | 2018 | Tunisia | PRJNA1160997 |
| GCA 022028755 | PDT000921131.1 | Klebsiella_15 | PDS000041726.21 | nan | NA | PRJEB27342 |
| GCA 022231935 | PDT000443857.1 | Klebsiella_15 | PDS000041726.21 | nan | NA | PRJEB8666 |
| GCA 022231425 | PDT000443996.1 | Klebsiella_15 | PDS000041726.21 | nan | NA | PRJEB8666 |
| GCA 022231985 | PDT000443854.1 | Klebsiella_15 | PDS000041726.21 | nan | NA | PRJEB8666 |
| GCA 002152405 | PDT000210736.1 | Klebsiella_2147 | PDS000041726.21 | 2015 | USA | PRJNA271899 |
| GCA 001033755 | PDT000068099.1 | Klebsiella_2147 | PDS000041726.21 | 2014 | NA | PRJNA271899 |
| GCA 001033885 | PDT000068105.1 | Klebsiella_15 | PDS000041726.21 | 2014 | NA | PRJNA271899 |
| GCA 014903595 | PDT000865015.1 | Klebsiella_15 | PDS000041726.21 | nan | USA | PRJNA271899 |
| GCA 001033455 | PDT000068086.1 | Klebsiella_- | PDS000041726.21 | 2014 | NA | PRJNA271899 |
| GCA 000688215 | PDT000030152.1 | Klebsiella_15 | PDS000041726.21 | 2013 | USA | PRJNA234115 |
| GCA 956481235 | PDT001864012.1 | Klebsiella_- | PDS000041726.21 | 2016 | Spain | PRJEB63349 |
| GCA 024488485 | PDT001373087.1 | Klebsiella_15 | | 2019 | Spain | PRJEB50822 |
| GCA 028561035 | PDT001610683.1 | Klebsiella_15 | PDS000106381.4 | 2020 | Spain | PRJEB57245 |
| GCA 040931645 | PDT002275246.1 | Klebsiella_15 | PDS000106381.4 | 2022 | Spain | PRJNA1132269 |
| GCA 905329805 | PDT001185122.2 | Klebsiella_15 | PDS000106381.4 | 2018 | Spain | PRJEB42440 |
| GCA 028562035 | PDT001610675.1 | Klebsiella_- | PDS000106381.4 | 2020 | Spain | PRJEB57245 |
| GCA 024490915 | PDT001373269.1 | Klebsiella_15 | PDS000106381.4 | 2019 | Spain | PRJEB50822 |
| GCA 028562135 | PDT001610669.1 | Klebsiella_15 | PDS000106381.4 | 2020 | Spain | PRJEB57245 |
| GCA 028562055 | PDT001610678.1 | Klebsiella_15 | PDS000106381.4 | 2020 | Spain | PRJEB57245 |

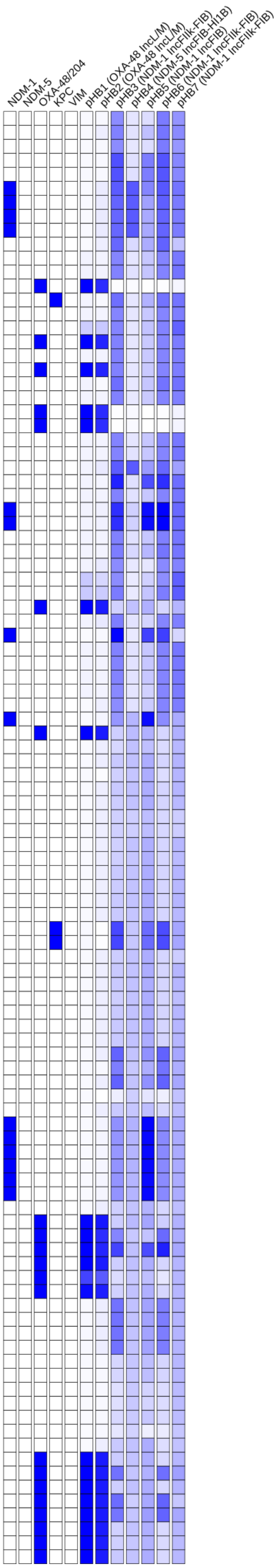

### Tree 27: GCA_043904845

Tree scale: 0.1

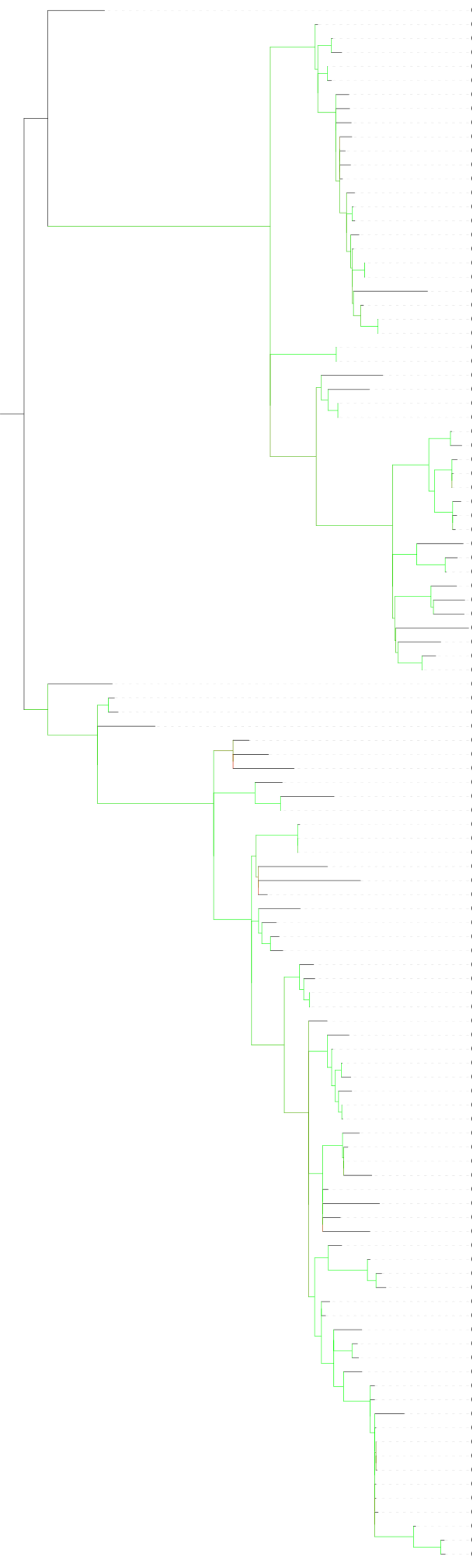

| | Isolate | MLST | SNP Cluster | Collection Year | Country | BioProject |
|---|---|---|---|---|---|---|
| GCA 037000505 | PDT002098839.1 | Klebsiella_383 | | 2015 | Egypt | PRJNA1079765 |
| GCA 017310305 | PDT000984751.1 | Klebsiella_383 | PDS000070357.5 | 2013 | China | PRJNA648010 |
| GCA 046466735 | PDT002556561.1 | Klebsiella_383 | PDS000070357.5 | nan | NA | PRJNA786399 |
| GCA 046466655 | PDT002556557.1 | Klebsiella_383 | PDS000070357.5 | nan | NA | PRJNA786399 |
| GCA 046466035 | PDT002556526.1 | Klebsiella_383 | PDS000070357.5 | nan | NA | PRJNA786399 |
| GCA 043109335 | PDT002420644.1 | Klebsiella_383 | PDS000070357.5 | 2014 | China | PRJNA1170063 |
| GCA 017311735 | PDT000984767.1 | Klebsiella_383 | PDS000070357.5 | 2013 | China | PRJNA648010 |
| GCA 017311645 | PDT000984763.1 | Klebsiella_383 | PDS000070357.5 | 2014 | China | PRJNA648010 |
| GCA 017311675 | PDT000984764.1 | Klebsiella_383 | PDS000070357.5 | 2017 | China | PRJNA648010 |
| GCA 014285595 | PDT000822624.1 | Klebsiella_383 | PDS000070357.5 | 2015 | China | PRJNA657553 |
| GCA 014289355 | PDT000822768.1 | Klebsiella_383 | PDS000070357.5 | 2013 | China | PRJNA657553 |
| GCA 017311635 | PDT000984762.1 | Klebsiella_383 | PDS000070357.5 | 2014 | China | PRJNA648010 |
| GCA 017311605 | PDT000984761.1 | Klebsiella_383 | PDS000070357.5 | 2013 | China | PRJNA648010 |
| GCA 014285645 | PDT000822627.1 | Klebsiella_383 | PDS000070357.5 | 2015 | China | PRJNA657553 |
| GCA 014285735 | PDT000822631.1 | Klebsiella_383 | PDS000070357.5 | 2014 | China | PRJNA657553 |
| GCA 046465235 | PDT002556486.1 | Klebsiella_383 | PDS000070357.5 | nan | NA | PRJNA786399 |
| GCA 014286025 | PDT000822646.1 | Klebsiella_383 | PDS000070357.5 | 2013 | China | PRJNA657553 |
| GCA 014285675 | PDT000822629.1 | Klebsiella_383 | PDS000070357.5 | 2013 | China | PRJNA657553 |
| GCA 014285835 | PDT000822636.1 | Klebsiella_383 | PDS000070357.5 | 2014 | China | PRJNA657553 |
| GCA 046465045 | PDT002556479.1 | Klebsiella_383 | PDS000070357.5 | nan | NA | PRJNA786399 |
| GCA 017311685 | PDT000984765.1 | Klebsiella_383 | PDS000070357.5 | 2017 | China | PRJNA648010 |
| GCA 014284855 | PDT000822587.1 | Klebsiella_383 | PDS000070357.5 | 2015 | China | PRJNA657553 |
| GCA 014285615 | PDT000822625.1 | Klebsiella_383 | PDS000070357.5 | 2015 | China | PRJNA657553 |
| GCA 014285545 | PDT000822621.1 | Klebsiella_383 | PDS000070357.5 | 2015 | China | PRJNA657553 |
| GCA 947662285 | PDT001590394.1 | Klebsiella_383 | PDS000133910.8 | 2007 | Greece | PRJEB58216 |
| GCA 028092175 | PDT001585477.1 | Klebsiella_383 | PDS000133910.8 | 2007 | Greece | PRJEB58216 |
| GCA 022104535 | PDT001243313.1 | Klebsiella_383 | PDS000106324.3 | 2016 | United Kingdom | PRJNA788733 |
| GCA 040201255 | PDT002208472.1 | Klebsiella_- | | nan | Saudi Arabia | PRJNA1116866 |
| GCA 022550215 | PDT001263624.1 | Klebsiella_383 | PDS000106324.3 | 2019 | Egypt | PRJNA720263 |
| GCA 022550175 | PDT001263622.1 | Klebsiella_383 | PDS000106324.3 | 2018 | Egypt | PRJNA720263 |
| GCA 012271775 | PDT000721361.1 | Klebsiella_383 | PDS000061038.3 | 2018 | Lebanon | PRJNA613441 |
| GCA 012271755 | PDT000721360.1 | Klebsiella_383 | PDS000061038.3 | 2018 | Lebanon | PRJNA613441 |
| GCA 021956345 | PDT001020277.1 | Klebsiella_383 | PDS000061038.3 | 2018 | Lebanon | PRJNA658369 |
| GCA 021965695 | PDT001019815.1 | Klebsiella_383 | PDS000061038.3 | 2018 | Lebanon | PRJNA658369 |
| GCA 021956085 | PDT001020290.1 | Klebsiella_383 | PDS000061038.3 | 2018 | Lebanon | PRJNA658369 |
| GCA 021962155 | PDT001020003.1 | Klebsiella_383 | PDS000061038.3 | 2018 | Lebanon | PRJNA658369 |
| GCA 021955425 | PDT001020322.1 | Klebsiella_383 | PDS000061038.3 | 2018 | Lebanon | PRJNA658369 |
| GCA 021968095 | PDT001019675.1 | Klebsiella_383 | PDS000061038.3 | 2018 | Lebanon | PRJNA658369 |
| GCA 043019365 | PDT002412609.1 | Klebsiella_383 | | 2024 | USA | PRJNA288601 |
| GCA 037493415 | PDT002120373.1 | Klebsiella_383 | PDS000179530.2 | 2024 | USA | PRJNA288601 |
| GCA 031043935 | PDT001729747.3 | Klebsiella_383 | PDS000179530.2 | 2023 | USA | PRJNA288601 |
| GCA 018441475 | PDT001046601.1 | Klebsiella_383 | | 2018 | Germany | PRJNA692829 |
| GCA 022017035 | PDT000948383.1 | Klebsiella_383 | | 2019 | Italy | PRJEB37610 |
| GCA 032721395 | PDT001927031.1 | Klebsiella_383 | | 2019 | Italy | PRJNA1026096 |
| GCA 021938955 | PDT001071296.1 | Klebsiella_383 | PDS000106730.2 | 2019 | Germany | PRJEB42370 |
| GCA 025037975 | PDT001386067.1 | Klebsiella_383 | | 2019 | Canada | PRJNA855907 |
| GCA 019781185 | PDT001118308.1 | Klebsiella_- | PDS000202475.1 | 2019 | Germany | PRJNA634442 |
| GCA 043832345 | PDT002430492.1 | Klebsiella_- | PDS000202475.1 | 2019 | Germany | PRJNA1070536 |
| GCA 022104735 | PDT001244769.1 | Klebsiella_383 | | 2016 | United Kingdom | PRJNA788733 |
| GCA 024087515 | PDT001350138.1 | Klebsiella_383 | PDS000108392.5 | 2016 | Qatar | PRJNA656934 |
| GCA 025144525 | PDT001416171.1 | Klebsiella_383 | PDS000108392.5 | 2016 | Qatar | PRJNA656934 |
| GCA 022550105 | PDT001263620.1 | Klebsiella_383 | | 2019 | Egypt | PRJNA720263 |
| GCA 023080795 | PDT001289734.1 | Klebsiella_383 | PDS000108392.5 | 2018 | Turkey | PRJNA789336 |
| GCA 016651765 | PDT000937804.1 | Klebsiella_383 | PDS000106324.3 | nan | Egypt | PRJNA689413 |
| GCA 016651915 | PDT000937812.1 | Klebsiella_383 | | nan | Egypt | PRJNA689413 |
| GCA 004803085 | PDT000495882.1 | Klebsiella_383 | | nan | Egypt | PRJNA508509 |
| GCA 022186005 | PDT000716575.1 | Klebsiella_383 | PDS000060640.1 | 2019 | Germany | PRJEB35890 |
| GCA 022186025 | PDT000716574.1 | Klebsiella_383 | PDS000060640.1 | 2019 | Germany | PRJEB35890 |
| GCA 037009425 | PDT002097979.1 | Klebsiella_6118 | PDS000175841.1 | 2024 | USA | PRJNA288601 |
| GCA 037009385 | PDT002097981.1 | Klebsiella_6118 | PDS000175841.1 | 2024 | USA | PRJNA288601 |
| GCA 037009405 | PDT002097980.1 | Klebsiella_6118 | PDS000175841.1 | 2024 | USA | PRJNA288601 |
| GCA 036455385 | PDT002074708.1 | Klebsiella_383 | | 2021 | Germany | PRJNA1060526 |
| GCA 046601475 | PDT002562417.1 | Klebsiella_383 | | 2021 | Singapore | PRJNA1206822 |
| GCA 922831335 | PDT001211077.1 | Klebsiella_383 | PDS000085592.11 | 2019 | United Kingdom | PRJEB48990 |
| GCA 042146905 | PDT002390135.1 | Klebsiella_383 | | 2024 | USA | PRJNA288601 |
| GCA 043000025 | PDT002129380.1 | Klebsiella_383 | | 2021 | Egypt | PRJNA1071125 |
| GCA 031047955 | PDT001725765.1 | Klebsiella_383 | PDS000180184.1 | 2020 | Egypt | PRJNA917066 |
| GCA 042999945 | PDT002129384.1 | Klebsiella_383 | PDS000180184.1 | 2021 | Egypt | PRJNA1071125 |
| GCA 032720515 | PDT001927068.1 | Klebsiella_383 | PDS000085592.11 | 2020 | Italy | PRJNA1026096 |
| GCA 947572755 | PDT001769582.1 | Klebsiella_383 | PDS000085592.11 | 2022 | Germany | PRJEB58018 |
| GCA 047426685 | PDT002605982.1 | Klebsiella_383 | PDS000085592.11 | 2019 | Italy | PRJNA1026096 |
| GCA 025889055 | PDT001469194.1 | Klebsiella_383 | PDS000085592.11 | 2019 | Italy | PRJNA787449 |
| GCA 021971055 | PDT001019316.3 | Klebsiella_383 | PDS000085592.11 | 2021 | USA | PRJNA288601 |
| GCA 027908495 | PDT001551547.1 | Klebsiella_383 | PDS000085592.11 | 2020 | Egypt | PRJNA825651 |
| GCA 043905145 | **HBH_3731_19** | Klebsiella_383 | PDS000085592.11 | 2019 | Tunisia | PRJNA1160997 |
| GCA 043905505 | **HBH_990_22** | Klebsiella_383 | PDS000085592.11 | 2022 | Tunisia | PRJNA1160997 |
| GCA 043905385 | **HBH_211_21** | Klebsiella_383 | PDS000085592.11 | 2021 | Tunisia | PRJNA1160997 |
| GCA 043905345 | **HBH_1362_21** | Klebsiella_383 | PDS000085592.11 | 2021 | Tunisia | PRJNA1160997 |
| GCA 043905355 | **HBH_3959_20** | Klebsiella_383 | PDS000085592.11 | 2020 | Tunisia | PRJNA1160997 |
| GCA 043905325 | **HBH_3788_20** | Klebsiella_383 | PDS000085592.11 | 2020 | Tunisia | PRJNA1160997 |
| GCA 043905185 | **HBH_1789_20** | Klebsiella_383 | PDS000098809.4 | 2020 | Tunisia | PRJNA1160997 |
| GCA 021898295 | PDT001172748.1 | Klebsiella_383 | PDS000098809.4 | 2020 | USA | PRJNA288601 |
| GCA 021898135 | PDT001174156.3 | Klebsiella_383 | PDS000098809.4 | 2020 | USA | PRJNA288601 |
| GCA 035082745 | PDT002045985.1 | Klebsiella_383 | | 2023 | USA | PRJNA288601 |
| GCA 043904905 | **HBH_487_19** | Klebsiella_383 | PDS000085592.11 | 2019 | Tunisia | PRJNA1160997 |
| GCA 043905405 | **HBH_3269_21** | Klebsiella_383 | | 2021 | Tunisia | PRJNA1160997 |
| GCA 043904845 | **HBH_1312_19** | Klebsiella_383 | PDS000085592.11 | 2019 | Tunisia | PRJNA1160997 |
| GCA 043905605 | **HBH_1879_22** | Klebsiella_383 | | 2022 | Tunisia | PRJNA1160997 |
| GCA 032721505 | PDT001927025.1 | Klebsiella_383 | PDS000217883.1 | 2019 | Italy | PRJNA1026096 |
| GCA 047501205 | PDT002615048.1 | Klebsiella_383 | PDS000217883.1 | 2021 | Tunisia | PRJNA1151248 |
| GCA 047501255 | PDT002615051.1 | Klebsiella_383 | PDS000217883.1 | 2023 | Tunisia | PRJNA1151248 |
| GCA 047501245 | PDT002615049.1 | Klebsiella_383 | PDS000217883.1 | 2023 | Tunisia | PRJNA1151248 |
| GCA 032721475 | PDT001927026.1 | Klebsiella_383 | PDS000085592.11 | 2019 | Italy | PRJNA1026096 |
| GCA 025892815 | PDT001469195.1 | Klebsiella_383 | PDS000085592.11 | 2019 | Italy | PRJNA787449 |
| GCA 035017125 | PDT002045096.1 | Klebsiella_383 | | 2022 | Libya | PRJNA998204 |
| GCA 035017085 | PDT002045094.1 | Klebsiella_383 | PDS000171211.1 | 2022 | Libya | PRJNA998204 |
| GCA 035016965 | PDT002045089.1 | Klebsiella_383 | PDS000171211.1 | 2022 | Libya | PRJNA998204 |
| GCA 039516715 | PDT002152585.1 | Klebsiella_383 | | 2024 | USA | PRJNA288601 |
| GCA 964277065 | PDT002423785.1 | Klebsiella_383 | PDS000192050.8 | 2024 | Slovakia | PRJEB72521 |
| GCA 964277015 | PDT002423787.1 | Klebsiella_383 | PDS000192050.8 | 2024 | Slovakia | PRJEB72521 |
| GCA 964339425 | PDT002468759.1 | Klebsiella_383 | PDS000192050.8 | 2024 | Slovakia | PRJEB72521 |
| GCA 964338835 | PDT002461347.1 | Klebsiella_383 | PDS000192050.8 | 2024 | Slovakia | PRJEB72521 |
| GCA 964338735 | PDT002462501.1 | Klebsiella_383 | PDS000192050.8 | 2024 | Slovakia | PRJEB72521 |
| GCA 964339405 | PDT002468761.1 | Klebsiella_383 | PDS000192050.8 | 2024 | Slovakia | PRJEB72521 |
| GCA 964339365 | PDT002468763.1 | Klebsiella_383 | PDS000192050.8 | 2024 | Slovakia | PRJEB72521 |
| GCA 964339305 | PDT002468760.1 | Klebsiella_383 | PDS000192050.8 | 2024 | Slovakia | PRJEB72521 |
| GCA 964417265 | PDT002518810.1 | Klebsiella_383 | PDS000192050.8 | 2024 | Slovakia | PRJEB72521 |
| GCA 964276885 | PDT002423793.1 | Klebsiella_383 | PDS000192050.8 | 2024 | Slovakia | PRJEB72521 |
| GCA 964339465 | PDT002468764.1 | Klebsiella_383 | PDS000192050.8 | 2024 | Slovakia | PRJEB72521 |
| GCA 964417235 | PDT002518801.1 | Klebsiella_383 | PDS000192050.8 | 2024 | Slovakia | PRJEB72521 |
| GCA 964417105 | PDT002518807.1 | Klebsiella_383 | PDS000192050.8 | 2024 | Slovakia | PRJEB72521 |

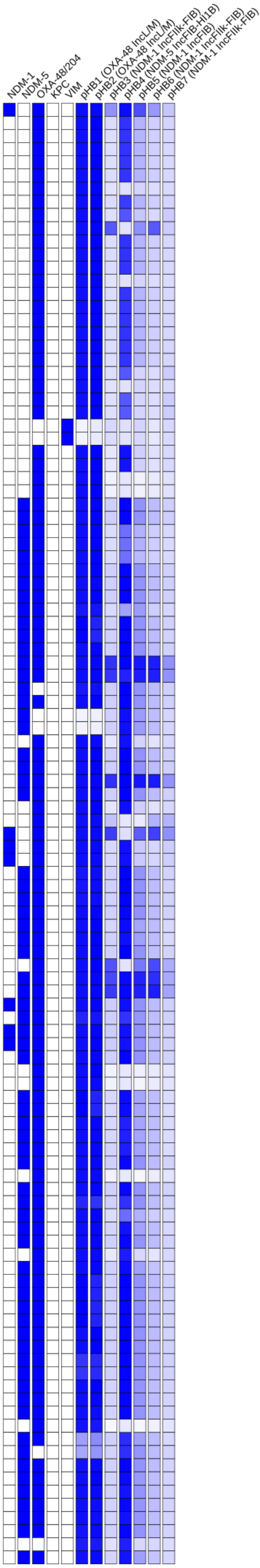

### Tree 28: GCA_043904865

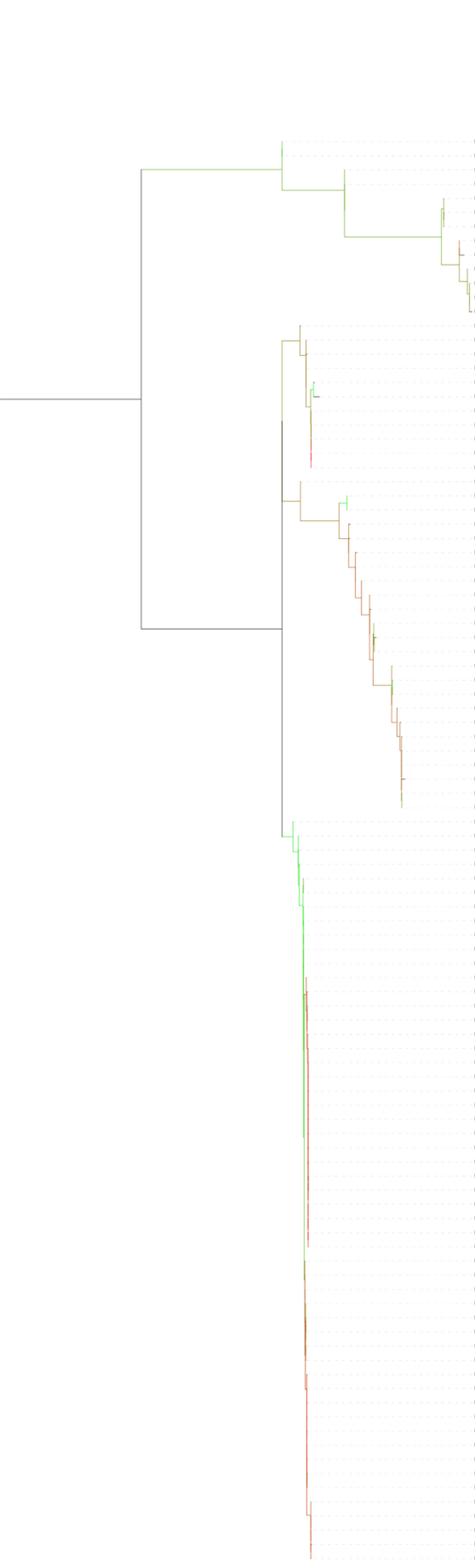


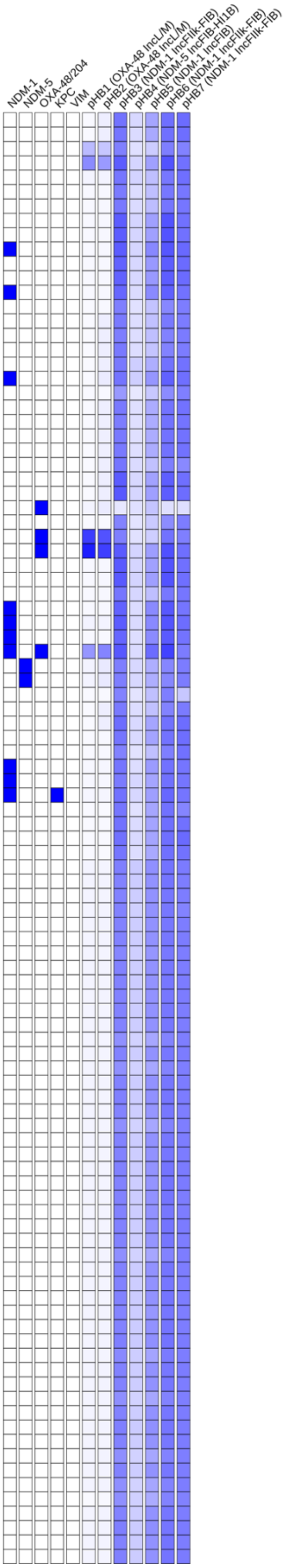


| | Isolate | MLST | SNP Cluster | Collection Year | Country | BioProject |
|---|---|---|---|---|---|---|
| GCA 027615765 | PDT001558318.1 | Klebsiella_340 | PDS000083373.14 | 2006 | Bangladesh | PRJNA845975 |
| GCA 027615465 | PDT001558345.1 | Klebsiella_340 | PDS000083373.14 | 2007 | Bangladesh | PRJNA845975 |
| GCA 900505565 | PDT000370452.1 | Klebsiella_340 | PDS000036321.13 | 2013 | Serbia | PRJEB10018 |
| GCA 900505585 | PDT000370454.1 | Klebsiella_340 | PDS000036321.13 | 2013 | Serbia | PRJEB10018 |
| GCA 900505405 | PDT000370438.1 | Klebsiella_340 | PDS000036321.13 | 2014 | Serbia | PRJEB10018 |
| GCA 900505465 | PDT000370444.1 | Klebsiella_340 | PDS000036321.13 | 2014 | Serbia | PRJEB10018 |
| GCA 900502515 | PDT000370030.1 | Klebsiella_340 | PDS000036321.13 | 2013 | Serbia | PRJEB10018 |
| GCA 900505455 | PDT000370442.1 | Klebsiella_340 | PDS000036321.13 | 2014 | Serbia | PRJEB10018 |
| GCA 900507085 | PDT000370461.1 | Klebsiella_340 | PDS000036321.13 | 2013 | Serbia | PRJEB10018 |
| GCA 900505115 | PDT000370394.1 | Klebsiella_340 | PDS000036321.13 | 2014 | Serbia | PRJEB10018 |
| GCA 900505105 | PDT000370395.1 | Klebsiella_340 | PDS000036321.13 | 2014 | Serbia | PRJEB10018 |
| GCA 900503495 | PDT000370420.1 | Klebsiella_340 | PDS000036321.13 | 2013 | Serbia | PRJEB10018 |
| GCA 902155965 | PDT000663175.1 | Klebsiella_340 | PDS000036321.13 | 2014 | Serbia | PRJEB33308 |
| GCA 022297255 | PDT000423562.1 | Klebsiella_340 | PDS000083373.14 | nan | NA | PRJEB17615 |
| GCA 022296975 | PDT000423578.1 | Klebsiella_340 | PDS000083373.14 | nan | NA | PRJEB17615 |
| GCA 022297095 | PDT000423571.1 | Klebsiella_340 | PDS000083373.14 | nan | NA | PRJEB17615 |
| GCA 022298895 | PDT000423473.1 | Klebsiella_340 | PDS000083373.14 | nan | NA | PRJEB17615 |
| GCA 022244755 | PDT000441138.1 | Klebsiella_340 | PDS000017086.10 | 2012 | Nepal | PRJEB1800 |
| GCA 027827735 | PDT001558013.1 | Klebsiella_- | PDS000083373.14 | 2010 | Bangladesh | PRJNA845975 |
| GCA 022275725 | PDT000432627.1 | Klebsiella_340 | PDS000083373.14 | 2008 | Ireland | PRJNA396729 |
| GCA 022275665 | PDT000432629.1 | Klebsiella_340 | PDS000083373.14 | 2008 | Ireland | PRJNA396729 |
| GCA 022275605 | PDT000432633.1 | Klebsiella_340 | PDS000083373.14 | 2008 | Ireland | PRJNA396729 |
| GCA 022275645 | PDT000432632.1 | Klebsiella_340 | PDS000083373.14 | 2008 | Ireland | PRJNA396729 |
| GCA 022275705 | PDT000432628.1 | Klebsiella_340 | PDS000083373.14 | 2008 | Ireland | PRJNA396729 |
| GCA 047058785 | PDT002566001.1 | Klebsiella_340 | | 2018 | Spain | PRJNA1150980 |
| GCA 021922125 | PDT001092057.1 | Klebsiella_340 | PDS000083373.14 | 2013 | Norway | PRJEB27256 |
| GCA 041860575 | PDT002369885.2 | Klebsiella_340 | PDS000083373.14 | 2013 | Norway | PRJNA1098507 |
| GCA 043904865 | **HBH_866_10** | Klebsiella_340 | | 2010 | Tunisia | PRJNA1160997 |
| GCA 032153575 | PDT001902908.1 | Klebsiella_340 | PDS000083373.14 | 2017 | Norway | PRJEB48268 |
| GCA 022287605 | PDT000427672.1 | Klebsiella_340 | PDS000036321.13 | nan | NA | PRJEB10018 |
| GCA 900505545 | PDT000370451.1 | Klebsiella_340 | PDS000036321.13 | 2013 | Serbia | PRJEB10018 |
| GCA 021921885 | PDT001092076.1 | Klebsiella_340 | PDS000036321.13 | 2014 | Norway | PRJEB27256 |
| GCA 956480655 | PDT001865600.1 | Klebsiella_340 | PDS000036321.13 | 2018 | Serbia | PRJEB63349 |
| GCA 956480695 | PDT001865601.1 | Klebsiella_340 | PDS000036321.13 | 2018 | Serbia | PRJEB63349 |
| GCA 022234485 | PDT000442756.1 | Klebsiella_340 | PDS000036321.13 | 2013 | Serbia | PRJNA449293 |
| GCA 900503575 | PDT000370443.1 | Klebsiella_340 | PDS000036321.13 | 2014 | Serbia | PRJEB10018 |
| GCA 041508645 | PDT002341334.1 | Klebsiella_340 | PDS000036321.13 | 2019 | USA | PRJNA288601 |
| GCA 022016735 | PDT000948692.1 | Klebsiella_340 | | 2019 | Qatar | PRJNA667192 |
| GCA 027616425 | PDT001558233.1 | Klebsiella_340 | PDS000130841.1 | 2021 | Bangladesh | PRJNA845975 |
| GCA 027617505 | PDT001558154.1 | Klebsiella_340 | PDS000130841.1 | 2021 | Bangladesh | PRJNA845975 |
| GCA 047276595 | PDT002596157.1 | Klebsiella_340 | PDS000083373.14 | 2018 | Australia | PRJNA674961 |
| GCA 022298635 | PDT000423487.1 | Klebsiella_340 | PDS000083373.14 | nan | NA | PRJEB17615 |
| GCA 022245285 | PDT000441101.1 | Klebsiella_340 | | 2012 | Nepal | PRJEB1800 |
| GCA 045010645 | PDT002049725.1 | Klebsiella_340 | PDS000083373.14 | 2019 | Bangladesh | PRJNA962904 |
| GCA 027615785 | PDT001558310.1 | Klebsiella_340 | PDS000083373.14 | 2007 | Bangladesh | PRJNA845975 |
| GCA 021982955 | PDT001003396.1 | Klebsiella_340 | PDS000083373.14 | 2016 | Thailand | PRJDB5929 |
| GCA 027884925 | PDT001557976.1 | Klebsiella_340 | PDS000130869.1 | 2013 | Bangladesh | PRJNA845975 |
| GCA 027828495 | PDT001557977.1 | Klebsiella_340 | PDS000130869.1 | 2015 | Bangladesh | PRJNA845975 |
| GCA 022302195 | PDT000193289.2 | Klebsiella_340 | PDS000083373.14 | 2008 | Thailand | PRJNA252957 |
| GCA 022302155 | PDT000193290.2 | Klebsiella_340 | PDS000083373.14 | 2008 | Thailand | PRJNA252957 |
| GCA 027828055 | PDT001557998.1 | Klebsiella_340 | PDS000083373.14 | 2007 | Bangladesh | PRJNA845975 |
| GCA 022302215 | PDT000193287.2 | Klebsiella_340 | PDS000083373.14 | 2008 | Thailand | PRJNA252957 |
| GCA 022008995 | PDT000986849.1 | Klebsiella_340 | PDS000083373.14 | 2015 | Thailand | PRJDB5929 |
| GCA 022027375 | PDT000925078.1 | Klebsiella_340 | PDS000083373.14 | 2016 | Singapore | PRJNA674942 |
| GCA 022002555 | PDT000986958.1 | Klebsiella_340 | PDS000083373.14 | 2015 | Thailand | PRJDB5929 |
| GCA 021981655 | PDT001008094.1 | Klebsiella_340 | PDS000083373.14 | 2017 | Thailand | PRJDB5929 |
| GCA 021982775 | PDT001007733.1 | Klebsiella_340 | PDS000083373.14 | 2017 | Thailand | PRJDB5929 |
| GCA 021986135 | PDT001003173.1 | Klebsiella_340 | PDS000083373.14 | 2017 | Thailand | PRJDB5929 |
| GCA 022005075 | PDT000986950.1 | Klebsiella_340 | PDS000083373.14 | 2015 | Thailand | PRJDB5929 |
| GCA 021988075 | PDT001003051.1 | Klebsiella_340 | PDS000083373.14 | 2016 | Thailand | PRJDB5929 |
| GCA 022005085 | PDT000986954.1 | Klebsiella_340 | PDS000083373.14 | 2015 | Thailand | PRJDB5929 |
| GCA 021983895 | PDT001003349.1 | Klebsiella_340 | PDS000083373.14 | 2017 | Thailand | PRJDB5929 |
| GCA 021985815 | PDT001003194.1 | Klebsiella_340 | PDS000083373.14 | 2017 | Thailand | PRJDB5929 |
| GCA 021987755 | PDT001003070.1 | Klebsiella_340 | PDS000083373.14 | 2017 | Thailand | PRJDB5929 |
| GCA 021981755 | PDT001008086.1 | Klebsiella_340 | PDS000083373.14 | 2016 | Thailand | PRJDB5929 |
| GCA 021984375 | PDT001003311.1 | Klebsiella_340 | PDS000083373.14 | 2017 | Thailand | PRJDB5929 |
| GCA 021984755 | PDT001003273.1 | Klebsiella_340 | PDS000083373.14 | 2016 | Thailand | PRJDB5929 |
| GCA 021984995 | PDT001003252.1 | Klebsiella_340 | PDS000083373.14 | 2016 | Thailand | PRJDB5929 |
| GCA 021988055 | PDT001003053.1 | Klebsiella_340 | PDS000083373.14 | 2016 | Thailand | PRJDB5929 |
| GCA 021984315 | PDT001003314.1 | Klebsiella_340 | PDS000083373.14 | 2017 | Thailand | PRJDB5929 |
| GCA 021984975 | PDT001003253.1 | Klebsiella_340 | PDS000083373.14 | 2016 | Thailand | PRJDB5929 |
| GCA 022002725 | PDT000986956.1 | Klebsiella_340 | PDS000083373.14 | 2015 | Thailand | PRJDB5929 |
| GCA 021982995 | PDT001003394.1 | Klebsiella_340 | PDS000083373.14 | 2016 | Thailand | PRJDB5929 |
| GCA 021987215 | PDT001003098.1 | Klebsiella_340 | PDS000083373.14 | 2016 | Thailand | PRJDB5929 |
| GCA 021986215 | PDT001003169.1 | Klebsiella_340 | PDS000083373.14 | 2017 | Thailand | PRJDB5929 |
| GCA 022005575 | PDT000986937.1 | Klebsiella_340 | PDS000083373.14 | 2015 | Thailand | PRJDB5929 |
| GCA 021987135 | PDT001003104.1 | Klebsiella_340 | PDS000083373.14 | 2017 | Thailand | PRJDB5929 |
| GCA 022005735 | PDT000986933.1 | Klebsiella_340 | PDS000083373.14 | 2015 | Thailand | PRJDB5929 |
| GCA 021982635 | PDT001008025.1 | Klebsiella_340 | PDS000083373.14 | 2016 | Thailand | PRJDB5929 |
| GCA 022001335 | PDT000987001.1 | Klebsiella_340 | PDS000083373.14 | 2016 | Thailand | PRJDB5929 |
| GCA 022005825 | PDT000986932.1 | Klebsiella_340 | PDS000083373.14 | 2015 | Thailand | PRJDB5929 |
| GCA 022005535 | PDT000986940.1 | Klebsiella_340 | PDS000083373.14 | 2015 | Thailand | PRJDB5929 |
| GCA 022005955 | PDT000986929.1 | Klebsiella_340 | PDS000083373.14 | 2015 | Thailand | PRJDB5929 |
| GCA 021984275 | PDT001003316.1 | Klebsiella_340 | PDS000083373.14 | 2017 | Thailand | PRJDB5929 |
| GCA 022001365 | PDT000987003.1 | Klebsiella_340 | PDS000083373.14 | 2016 | Thailand | PRJDB5929 |
| GCA 022005555 | PDT000986935.1 | Klebsiella_340 | PDS000083373.14 | 2015 | Thailand | PRJDB5929 |
| GCA 021986745 | PDT001003136.1 | Klebsiella_340 | PDS000083373.14 | 2016 | Thailand | PRJDB5929 |
| GCA 021981295 | PDT001008138.1 | Klebsiella_340 | PDS000083373.14 | 2017 | Thailand | PRJDB5929 |
| GCA 021985035 | PDT001003251.1 | Klebsiella_340 | PDS000083373.14 | 2016 | Thailand | PRJDB5929 |
| GCA 021982835 | PDT001003422.1 | Klebsiella_340 | PDS000083373.14 | 2017 | Thailand | PRJDB5929 |
| GCA 022000985 | PDT000987027.1 | Klebsiella_340 | PDS000083373.14 | 2016 | Thailand | PRJDB5929 |
| GCA 021984355 | PDT001003312.1 | Klebsiella_340 | PDS000083373.14 | 2017 | Thailand | PRJDB5929 |
| GCA 021987295 | PDT001003095.1 | Klebsiella_340 | PDS000083373.14 | 2016 | Thailand | PRJDB5929 |
| GCA 021986015 | PDT001003179.1 | Klebsiella_340 | PDS000083373.14 | 2017 | Thailand | PRJDB5929 |
| GCA 021984255 | PDT001003315.1 | Klebsiella_340 | PDS000083373.14 | 2017 | Thailand | PRJDB5929 |
| GCA 021987795 | PDT001003069.1 | Klebsiella_340 | PDS000083373.14 | 2017 | Thailand | PRJDB5929 |
| GCA 021981775 | PDT001008085.1 | Klebsiella_340 | PDS000083373.14 | 2016 | Thailand | PRJDB5929 |
| GCA 021987115 | PDT001003106.1 | Klebsiella_340 | PDS000083373.14 | 2017 | Thailand | PRJDB5929 |
| GCA 022005135 | PDT000986951.1 | Klebsiella_340 | PDS000083373.14 | 2015 | Thailand | PRJDB5929 |
| GCA 021985075 | PDT001003249.1 | Klebsiella_340 | PDS000083373.14 | 2016 | Thailand | PRJDB5929 |
| GCA 022000655 | PDT000987028.1 | Klebsiella_340 | PDS000083373.14 | 2016 | Thailand | PRJDB5929 |

### Tree 29: GCA_043904885

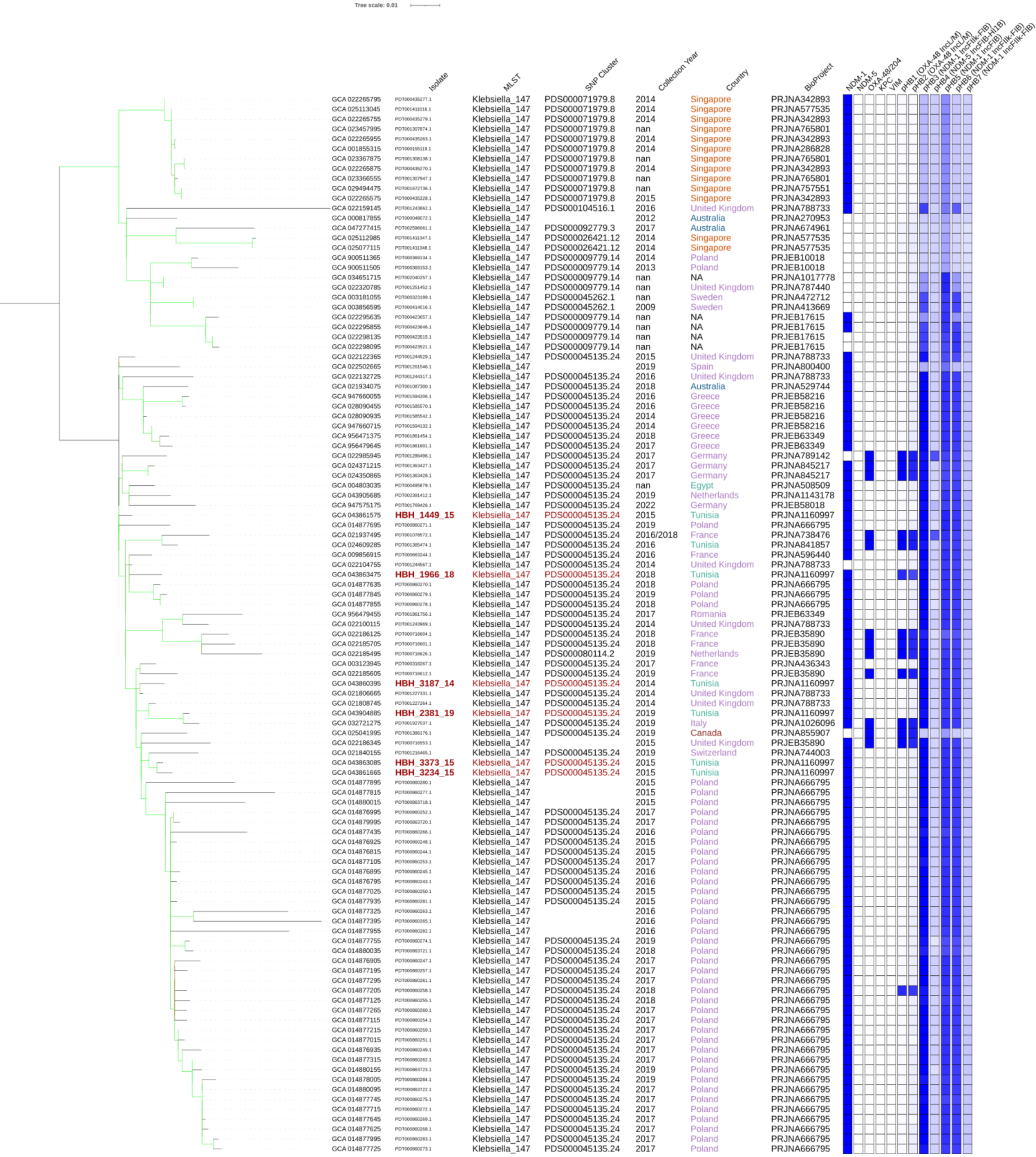

### Tree 30: GCA_043905265

Tree scale: 0.01

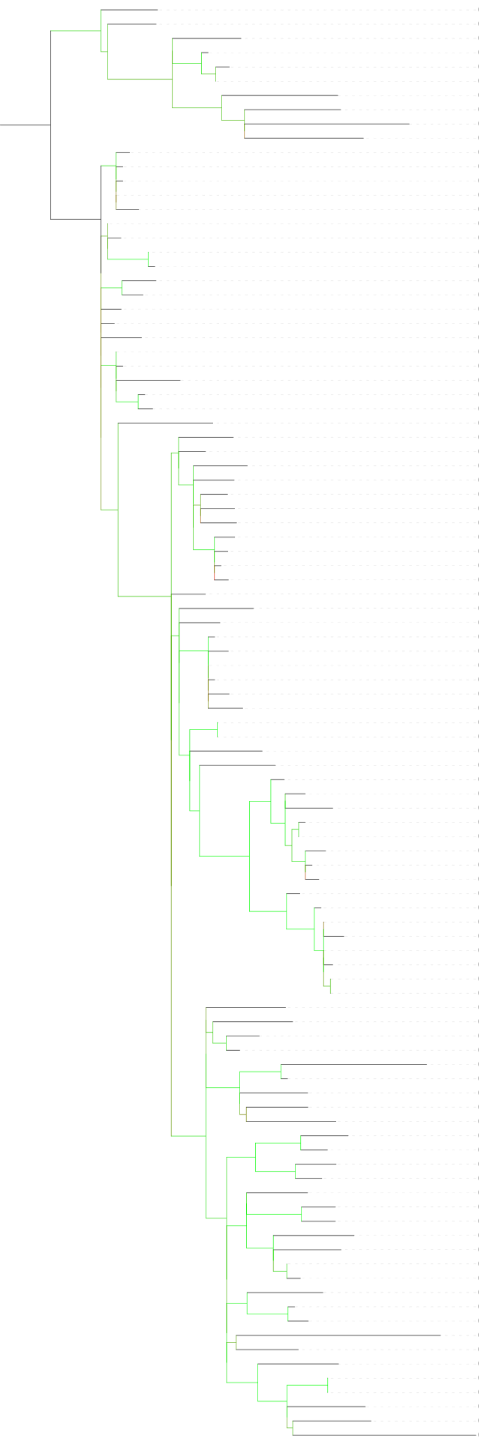

| | Isolate | MLST | SNP Cluster | Collection Year | Country | BioProject |
|---|---|---|---|---|---|---|
| GCA 023022385 | PDT001289768.1 | Klebsiella_- | PDS000060581.66 | 2018 | Turkey | PRJNA789336 |
| GCA 023021785 | PDT001289804.1 | Klebsiella_2096 | PDS000060581.66 | 2019 | Turkey | PRJNA789336 |
| GCA 023022865 | PDT001289743.1 | Klebsiella_2096 | PDS000060581.66 | 2019 | Turkey | PRJNA789336 |
| GCA 023022665 | PDT001289755.1 | Klebsiella_- | PDS000060581.66 | 2018 | Turkey | PRJNA789336 |
| GCA 023022785 | PDT001289751.1 | Klebsiella_- | PDS000060581.66 | 2018 | Turkey | PRJNA789336 |
| GCA 023022745 | PDT001289753.1 | Klebsiella_2096 | PDS000060581.66 | 2018 | Turkey | PRJNA789336 |
| GCA 040328715 | PDT002230899.1 | Klebsiella_2096 | PDS000060581.66 | 2018 | Saudi Arabia | PRJNA1107218 |
| GCA 021278205 | PDT001211937.1 | Klebsiella_2096 | PDS000060581.66 | 2019 | Pakistan | PRJNA704247 |
| GCA 017640385 | PDT000995792.1 | Klebsiella_2096 | PDS000060581.66 | 2018 | Germany | PRJNA644257 |
| GCA 022749215 | PDT001277597.1 | Klebsiella_2096 | PDS000060581.66 | 2019 | Turkey | PRJNA816050 |
| GCA 023024065 | PDT001289675.1 | Klebsiella_2096 | PDS000060581.66 | 2019 | Turkey | PRJNA789336 |
| GCA 023021825 | PDT001289799.1 | Klebsiella_2096 | PDS000060581.66 | 2019 | Turkey | PRJNA789336 |
| GCA 023026165 | PDT001289580.1 | Klebsiella_2096 | PDS000060581.66 | 2019 | Turkey | PRJNA789336 |
| GCA 023023225 | PDT001289731.1 | Klebsiella_2096 | PDS000060581.66 | 2018 | Turkey | PRJNA789336 |
| GCA 023021745 | PDT001289803.1 | Klebsiella_2096 | PDS000060581.66 | 2019 | Turkey | PRJNA789336 |
| GCA 023021865 | PDT001289796.1 | Klebsiella_2096 | PDS000060581.66 | 2018 | Turkey | PRJNA789336 |
| GCA 023026385 | PDT001289568.1 | Klebsiella_- | PDS000060581.66 | 2018 | Turkey | PRJNA789336 |
| GCA 023021725 | PDT001289805.1 | Klebsiella_- | PDS000060581.66 | 2019 | Turkey | PRJNA789336 |
| GCA 023025845 | PDT001289595.1 | Klebsiella_2096 | PDS000060581.66 | 2019 | Turkey | PRJNA789336 |
| GCA 023024205 | PDT001289668.1 | Klebsiella_- | PDS000060581.66 | 2019 | Turkey | PRJNA789336 |
| GCA 023024405 | PDT001289658.1 | Klebsiella_- | PDS000060581.66 | 2019 | Turkey | PRJNA789336 |
| GCA 023021605 | PDT001289815.1 | Klebsiella_- | PDS000060581.66 | 2018 | Turkey | PRJNA789336 |
| GCA 023024385 | PDT001289659.1 | Klebsiella_2096 | PDS000060581.66 | 2018 | Turkey | PRJNA789336 |
| GCA 023022505 | PDT001289762.1 | Klebsiella_- | PDS000060581.66 | 2018 | Turkey | PRJNA789336 |
| GCA 023026305 | PDT001289576.1 | Klebsiella_- | PDS000060581.66 | 2018 | Turkey | PRJNA789336 |
| GCA 023026365 | PDT001289571.1 | Klebsiella_2096 | PDS000060581.66 | 2018 | Turkey | PRJNA789336 |
| GCA 023465555 | PDT001289584.1 | Klebsiella_2096 | PDS000060581.66 | 2019 | Turkey | PRJNA789336 |
| GCA 023022585 | PDT001289759.1 | Klebsiella_- | PDS000060581.66 | 2018 | Turkey | PRJNA789336 |
| GCA 023026085 | PDT001289585.1 | Klebsiella_2096 | PDS000060581.66 | 2019 | Turkey | PRJNA789336 |
| GCA 023274385 | PDT001304199.1 | Klebsiella_2096 | PDS000060581.66 | 2021 | France | PRJNA832942 |
| GCA 023022145 | PDT001289779.1 | Klebsiella_- | PDS000060581.66 | 2018 | Turkey | PRJNA789336 |
| GCA 023080725 | PDT001289737.1 | Klebsiella_- | PDS000060581.66 | 2018 | Turkey | PRJNA789336 |
| GCA 956480265 | PDT001864140.1 | Klebsiella_2096 | PDS000060581.66 | 2017 | Turkey | PRJEB63349 |
| GCA 045319155 | PDT002203440.1 | Klebsiella_2096 | PDS000060581.66 | 2018 | Turkey | PRJNA1099381 |
| GCA 023022725 | PDT001289724.1 | Klebsiella_2096 | PDS000060581.66 | 2018 | Turkey | PRJNA789336 |
| GCA 023022805 | PDT001289748.1 | Klebsiella_2096 | PDS000060581.66 | 2018 | Turkey | PRJNA789336 |
| GCA 045318935 | PDT002203450.1 | Klebsiella_- | PDS000060581.66 | 2019 | Turkey | PRJNA1099381 |
| GCA 045318995 | PDT002203446.1 | Klebsiella_- | PDS000060581.66 | 2019 | Turkey | PRJNA1099381 |
| GCA 045319175 | PDT002203441.1 | Klebsiella_2096 | PDS000060581.66 | 2018 | Turkey | PRJNA1099381 |
| GCA 023025945 | PDT001289591.1 | Klebsiella_2096 | PDS000060581.66 | 2018 | Turkey | PRJNA789336 |
| GCA 045319195 | PDT002203436.1 | Klebsiella_2096 | PDS000060581.66 | 2018 | Turkey | PRJNA1099381 |
| GCA 016651605 | PDT000937796.1 | Klebsiella_2096 | PDS000060581.66 | nan | Egypt | PRJNA689413 |
| GCA 040328555 | PDT002230891.1 | Klebsiella_2096 | PDS000060581.66 | 2018 | Saudi Arabia | PRJNA1107218 |
| GCA 023022705 | PDT001289752.1 | Klebsiella_2096 | PDS000060581.66 | 2018 | Turkey | PRJNA789336 |
| GCA 023024345 | PDT001289660.1 | Klebsiella_- | PDS000060581.66 | 2019 | Turkey | PRJNA789336 |
| GCA 023024365 | PDT001289661.1 | Klebsiella_2096 | PDS000060581.66 | 2019 | Turkey | PRJNA789336 |
| GCA 023024125 | PDT001289671.1 | Klebsiella_2096 | PDS000060581.66 | 2019 | Turkey | PRJNA789336 |
| GCA 023024325 | PDT001289662.1 | Klebsiella_- | PDS000060581.66 | 2019 | Turkey | PRJNA789336 |
| GCA 023025725 | PDT001289603.1 | Klebsiella_2096 | PDS000060581.66 | 2019 | Turkey | PRJNA789336 |
| GCA 023025865 | PDT001289598.1 | Klebsiella_2096 | PDS000060581.66 | 2019 | Turkey | PRJNA789336 |
| GCA 023023445 | PDT001289701.1 | Klebsiella_2096 | PDS000060581.66 | 2018 | Turkey | PRJNA789336 |
| GCA 023026405 | PDT001289567.1 | Klebsiella_- | PDS000060581.66 | 2018 | Turkey | PRJNA789336 |
| GCA 023021905 | PDT001289798.1 | Klebsiella_2096 | PDS000060581.66 | 2019 | Turkey | PRJNA789336 |
| GCA 043905265 | **HBH_3672_20** | Klebsiella_2096 | PDS000060581.66 | 2020 | Tunisia | PRJNA1160997 |
| GCA 023022485 | PDT001289764.1 | Klebsiella_2096 | PDS000060581.66 | 2018 | Turkey | PRJNA789336 |
| GCA 023025605 | PDT001289606.1 | Klebsiella_2096 | PDS000060581.66 | 2019 | Turkey | PRJNA789336 |
| GCA 023021885 | PDT001289795.1 | Klebsiella_2096 | PDS000060581.66 | 2019 | Turkey | PRJNA789336 |
| GCA 023024245 | PDT001289666.1 | Klebsiella_- | PDS000060581.66 | 2019 | Turkey | PRJNA789336 |
| GCA 023024305 | PDT001289664.1 | Klebsiella_2096 | PDS000060581.66 | 2018 | Turkey | PRJNA789336 |
| GCA 023025685 | PDT001289604.1 | Klebsiella_2096 | PDS000060581.66 | 2019 | Turkey | PRJNA789336 |
| GCA 023021985 | PDT001289791.1 | Klebsiella_- | PDS000060581.66 | 2019 | Turkey | PRJNA789336 |
| GCA 023021965 | PDT001289792.1 | Klebsiella_2096 | PDS000060581.66 | 2019 | Turkey | PRJNA789336 |
| GCA 023275055 | PDT001304225.1 | Klebsiella_2096 | PDS000060581.66 | 2020 | France | PRJNA832942 |
| GCA 031039855 | PDT001232561.3 | Klebsiella_2096 | PDS000060581.66 | 2021 | USA | PRJNA288601 |
| GCA 029738715 | PDT001689449.3 | Klebsiella_2096 | PDS000060581.66 | 2022 | USA | PRJNA288601 |
| GCA 029749135 | PDT001689448.3 | Klebsiella_2096 | PDS000060581.66 | 2022 | USA | PRJNA288601 |
| GCA 032600685 | PDT001925789.2 | Klebsiella_2096 | PDS000060581.66 | 2023 | USA | PRJNA288601 |
| GCA 031038675 | PDT001707495.3 | Klebsiella_- | PDS000060581.66 | 2022 | USA | PRJNA288601 |
| GCA 029738935 | PDT001689450.3 | Klebsiella_- | PDS000060581.66 | 2022 | USA | PRJNA288601 |
| GCA 029749195 | PDT001689451.3 | Klebsiella_2096 | PDS000060581.66 | 2022 | USA | PRJNA288601 |
| GCA 040325645 | PDT002230872.1 | Klebsiella_2096 | PDS000060581.66 | 2018 | Saudi Arabia | PRJNA1107218 |
| GCA 023274075 | PDT001304186.1 | Klebsiella_2096 | PDS000060581.66 | 2019 | France | PRJNA832942 |
| GCA 040328695 | PDT002230898.1 | Klebsiella_2096 | PDS000060581.66 | 2018 | Saudi Arabia | PRJNA1107218 |
| GCA 040328475 | PDT002230887.1 | Klebsiella_2096 | PDS000060581.66 | 2018 | Saudi Arabia | PRJNA1107218 |
| GCA 024428875 | PDT001366404.1 | Klebsiella_2096 | PDS000060581.66 | nan | USA | PRJNA825705 |
| GCA 024430015 | PDT001366487.1 | Klebsiella_2096 | PDS000060581.66 | nan | USA | PRJNA825705 |
| GCA 044365005 | PDT002455539.1 | Klebsiella_2096 | PDS000060581.66 | nan | Saudi Arabia | PRJNA1116866 |
| GCA 037204925 | PDT002115697.1 | Klebsiella_2096 | PDS000060581.66 | 2022 | Saudi Arabia | PRJNA1018815 |
| GCA 023026425 | PDT001289569.1 | Klebsiella_2096 | PDS000060581.66 | 2018 | Turkey | PRJNA789336 |
| GCA 044379915 | PDT002455629.1 | Klebsiella_2096 | PDS000060581.66 | nan | Saudi Arabia | PRJNA1116866 |
| GCA 044366445 | PDT002455609.1 | Klebsiella_2096 | PDS000060581.66 | nan | Saudi Arabia | PRJNA1116866 |
| GCA 044366525 | PDT002455613.1 | Klebsiella_2096 | PDS000060581.66 | nan | Saudi Arabia | PRJNA1116866 |
| GCA 040328735 | PDT002230900.1 | Klebsiella_2096 | PDS000060581.66 | 2018 | Saudi Arabia | PRJNA1107218 |
| GCA 023275535 | PDT001304249.1 | Klebsiella_2096 | PDS000060581.66 | 2017 | France | PRJNA832942 |
| GCA 044366145 | PDT002455594.1 | Klebsiella_2096 | PDS000060581.66 | nan | Saudi Arabia | PRJNA1116866 |
| GCA 044365425 | PDT002455560.1 | Klebsiella_2096 | PDS000060581.66 | nan | Saudi Arabia | PRJNA1116866 |
| GCA 956481325 | PDT001864161.1 | Klebsiella_2096 | PDS000060581.66 | 2018 | Turkey | PRJEB63349 |
| GCA 956481575 | PDT001864166.1 | Klebsiella_2096 | PDS000060581.66 | 2018 | Turkey | PRJEB63349 |
| GCA 023275595 | PDT001304252.1 | Klebsiella_2096 | PDS000060581.66 | 2018 | France | PRJNA832942 |
| GCA 956481025 | PDT001864152.1 | Klebsiella_2096 | PDS000060581.66 | 2018 | Turkey | PRJEB63349 |
| GCA 037202245 | PDT002115593.1 | Klebsiella_2096 | PDS000060581.66 | 2022 | Saudi Arabia | PRJNA1018815 |
| GCA 043358915 | PDT002423177.1 | Klebsiella_2096 | PDS000060581.66 | nan | Saudi Arabia | PRJNA1116866 |
| GCA 044383125 | PDT002456521.1 | Klebsiella_2096 | PDS000060581.66 | nan | Saudi Arabia | PRJNA1116866 |
| GCA 029842045 | PDT001695770.1 | Klebsiella_2096 | PDS000060581.66 | 2018 | Pakistan | PRJNA868296 |
| GCA 025878705 | PDT001467902.1 | Klebsiella_2096 | PDS000060581.66 | 2018 | Norway | PRJEB56146 |
| GCA 044380805 | PDT002455655.1 | Klebsiella_2096 | PDS000060581.66 | nan | Saudi Arabia | PRJNA1116866 |
| GCA 021934935 | PDT001087257.1 | Klebsiella_2096 | PDS000060581.66 | 2017 | Australia | PRJNA529744 |
| GCA 047269925 | PDT002596114.1 | Klebsiella_2096 | PDS000060581.66 | 2017 | Australia | PRJNA674961 |
| GCA 022188325 | PDT000715645.1 | Klebsiella_2096 | PDS000060581.66 | 2018 | Saudi Arabia | PRJEB36683 |
| GCA 022188125 | PDT000715662.1 | Klebsiella_- | PDS000060581.66 | 2018 | Saudi Arabia | PRJEB36683 |
| GCA 022329185 | PDT000534832.1 | Klebsiella_- | PDS000060581.66 | 2015 | Oman | PRJNA544438 |

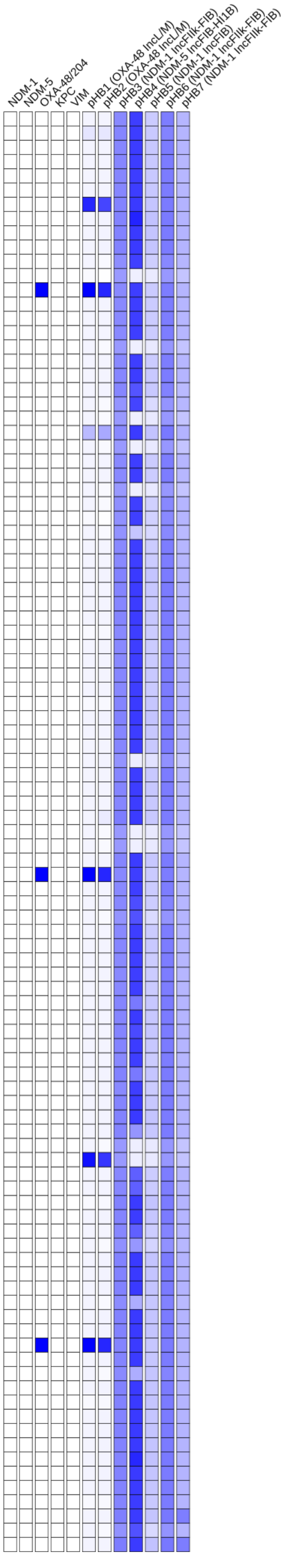

### Tree 31: GCA_043905305

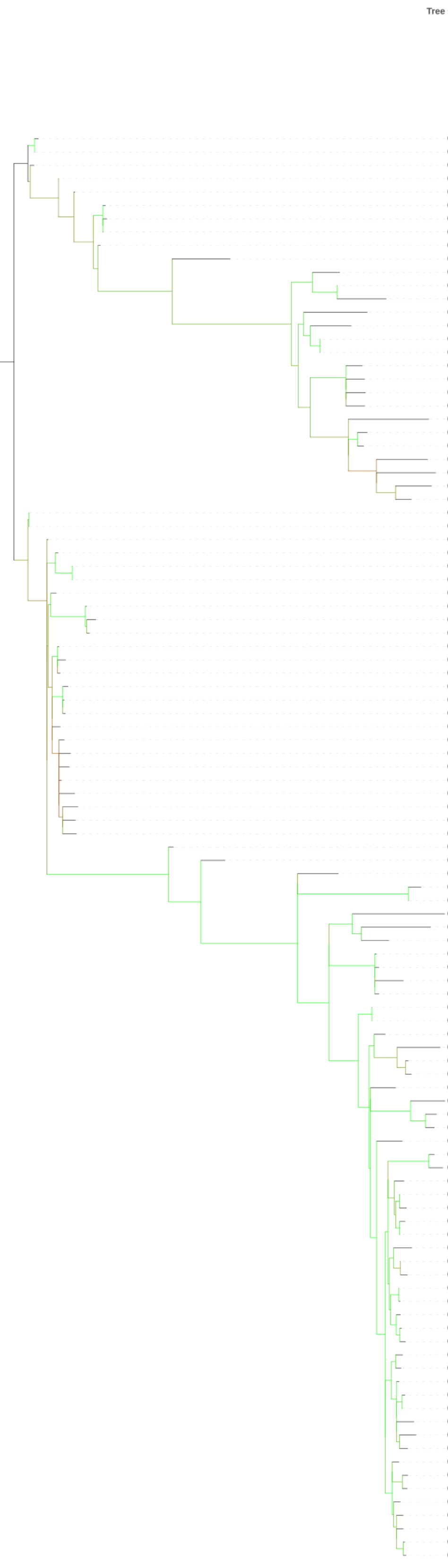


| | Isolate | MLST | SNP Cluster | Collection Year | Country | BioProject |
|---|---|---|---|---|---|---|
| GCA 014285715 | PDT000822630.1 | Klebsiella_383 | PDS000070357.5 | 2014 | China | PRJNA657553 |
| GCA 014285775 | PDT000822634.1 | Klebsiella_383 | PDS000070357.5 | 2014 | China | PRJNA657553 |
| GCA 046465065 | PDT002556478.1 | Klebsiella_383 | PDS000070357.5 | nan | NA | PRJNA786399 |
| GCA 046466655 | PDT002556557.1 | Klebsiella_383 | PDS000070357.5 | nan | NA | PRJNA786399 |
| GCA 046466735 | PDT002556561.1 | Klebsiella_383 | PDS000070357.5 | nan | NA | PRJNA786399 |
| GCA 046466415 | PDT002556545.1 | Klebsiella_383 | PDS000070357.5 | nan | NA | PRJNA786399 |
| GCA 043109335 | PDT002420644.1 | Klebsiella_383 | PDS000070357.5 | 2014 | China | PRJNA1170063 |
| GCA 046466035 | PDT002556526.1 | Klebsiella_383 | PDS000070357.5 | nan | NA | PRJNA786399 |
| GCA 017310305 | PDT000984751.1 | Klebsiella_383 | PDS000070357.5 | 2013 | China | PRJNA648010 |
| GCA 022104735 | PDT001244769.1 | Klebsiella_383 | | 2016 | United Kingdom | PRJNA788733 |
| GCA 004803085 | PDT000495882.1 | Klebsiella_383 | | nan | Egypt | PRJNA508509 |
| GCA 022186025 | PDT000716574.1 | Klebsiella_383 | PDS000060640.1 | 2019 | Germany | PRJEB35890 |
| GCA 022186005 | PDT000716575.1 | Klebsiella_383 | PDS000060640.1 | 2019 | Germany | PRJEB35890 |
| GCA 022104535 | PDT001243313.1 | Klebsiella_383 | PDS000106324.3 | 2016 | United Kingdom | PRJNA788733 |
| GCA 040201255 | PDT002208472.1 | Klebsiella_- | | nan | Saudi Arabia | PRJNA1116866 |
| GCA 022550215 | PDT001263624.1 | Klebsiella_383 | PDS000106324.3 | 2019 | Egypt | PRJNA720263 |
| GCA 022550175 | PDT001263622.1 | Klebsiella_383 | PDS000106324.3 | 2018 | Egypt | PRJNA720263 |
| GCA 043905145 | **HBH_3731_19** | Klebsiella_383 | PDS000085592.11 | 2019 | Tunisia | PRJNA1160997 |
| GCA 043904905 | **HBH_487_19** | Klebsiella_383 | PDS000085592.11 | 2019 | Tunisia | PRJNA1160997 |
| GCA 025892815 | PDT001469195.1 | Klebsiella_383 | PDS000085592.11 | 2019 | Italy | PRJNA787449 |
| GCA 021971055 | PDT001019316.3 | Klebsiella_383 | PDS000085592.11 | 2021 | USA | PRJNA288601 |
| GCA 037000505 | PDT002098839.1 | Klebsiella_383 | | 2015 | Egypt | PRJNA1079765 |
| GCA 025144525 | PDT001416171.1 | Klebsiella_383 | PDS000108392.5 | 2016 | Qatar | PRJNA656934 |
| GCA 024087515 | PDT001350138.1 | Klebsiella_383 | PDS000108392.5 | 2016 | Qatar | PRJNA656934 |
| GCA 025037975 | PDT001386067.1 | Klebsiella_383 | | 2019 | Canada | PRJNA855907 |
| GCA 922831335 | PDT001211077.1 | Klebsiella_383 | PDS000085592.11 | 2019 | United Kingdom | PRJEB48990 |
| GCA 016651765 | PDT000937804.1 | Klebsiella_383 | PDS000106324.3 | nan | Egypt | PRJNA689413 |
| GCA 023080795 | PDT001289734.1 | Klebsiella_383 | PDS000108392.5 | 2018 | Turkey | PRJNA789336 |
| GCA 014285835 | PDT000822636.1 | Klebsiella_383 | PDS000070357.5 | 2014 | China | PRJNA657553 |
| GCA 046465045 | PDT002556479.1 | Klebsiella_383 | PDS000070357.5 | nan | NA | PRJNA786399 |
| GCA 014285675 | PDT000822629.1 | Klebsiella_383 | PDS000070357.5 | 2013 | China | PRJNA657553 |
| GCA 014284855 | PDT000822587.1 | Klebsiella_383 | PDS000070357.5 | 2015 | China | PRJNA657553 |
| GCA 014285545 | PDT000822621.1 | Klebsiella_383 | PDS000070357.5 | 2015 | China | PRJNA657553 |
| GCA 014285615 | PDT000822625.1 | Klebsiella_383 | PDS000070357.5 | 2015 | China | PRJNA657553 |
| GCA 014286025 | PDT000822646.1 | Klebsiella_383 | PDS000070357.5 | 2013 | China | PRJNA657553 |
| GCA 037052525 | PDT002102101.1 | Klebsiella_383 | PDS000176495.1 | 2013 | Jordan | PRJNA1076682 |
| GCA 037052245 | PDT002102087.1 | Klebsiella_383 | PDS000176495.1 | 2015 | Jordan | PRJNA1076682 |
| GCA 037052285 | PDT002102089.1 | Klebsiella_383 | PDS000176495.1 | 2013 | Jordan | PRJNA1076682 |
| GCA 014285735 | PDT000822631.1 | Klebsiella_383 | PDS000070357.5 | 2014 | China | PRJNA657553 |
| GCA 014288455 | PDT000822723.1 | Klebsiella_383 | PDS000070357.5 | 2014 | China | PRJNA657553 |
| GCA 046465235 | PDT002556486.1 | Klebsiella_383 | PDS000070357.5 | nan | NA | PRJNA786399 |
| GCA 017311535 | PDT000984759.1 | Klebsiella_383 | PDS000070357.5 | 2015 | China | PRJNA648010 |
| GCA 017311595 | PDT000984760.1 | Klebsiella_383 | PDS000070357.5 | 2014 | China | PRJNA648010 |
| GCA 014285665 | PDT000822628.1 | Klebsiella_383 | PDS000070357.5 | 2014 | China | PRJNA657553 |
| GCA 014285645 | PDT000822627.1 | Klebsiella_383 | PDS000070357.5 | 2015 | China | PRJNA657553 |
| GCA 014289355 | PDT000822768.1 | Klebsiella_383 | PDS000070357.5 | 2013 | China | PRJNA657553 |
| GCA 014285595 | PDT000822624.1 | Klebsiella_383 | PDS000070357.5 | 2015 | China | PRJNA657553 |
| GCA 017311635 | PDT000984762.1 | Klebsiella_383 | PDS000070357.5 | 2014 | China | PRJNA648010 |
| GCA 017311605 | PDT000984761.1 | Klebsiella_383 | PDS000070357.5 | 2013 | China | PRJNA648010 |
| GCA 014288555 | PDT000822728.1 | Klebsiella_383 | PDS000070357.5 | 2014 | China | PRJNA657553 |
| GCA 017311675 | PDT000984764.1 | Klebsiella_383 | PDS000070357.5 | 2017 | China | PRJNA648010 |
| GCA 017311735 | PDT000984767.1 | Klebsiella_383 | PDS000070357.5 | 2013 | China | PRJNA648010 |
| GCA 017311645 | PDT000984763.1 | Klebsiella_383 | PDS000070357.5 | 2014 | China | PRJNA648010 |
| GCA 017311685 | PDT000984765.1 | Klebsiella_383 | PDS000070357.5 | 2017 | China | PRJNA648010 |
| GCA 043949915 | PDT002438925.1 | Klebsiella_383 | PDS000070357.5 | 2015 | China | PRJNA1173368 |
| GCA 021798805 | PDT001227454.1 | Klebsiella_383 | | 2014 | United Kingdom | PRJNA788733 |
| GCA 019781185 | PDT001118308.1 | Klebsiella_- | PDS000202475.1 | 2019 | Germany | PRJNA634442 |
| GCA 043832345 | PDT002430492.1 | Klebsiella_- | PDS000202475.1 | 2019 | Germany | PRJNA1070536 |
| GCA 900513485 | PDT000369524.1 | Klebsiella_383 | | 2013 | Israel | PRJEB10018 |
| GCA 916161735 | PDT001141241.1 | Klebsiella_383 | | 2015 | Netherlands | PRJEB41042 |
| GCA 900504425 | PDT000370281.1 | Klebsiella_383 | | 2014 | Romania | PRJEB10018 |
| GCA 043859635 | **HBH_233_09** | Klebsiella_383 | PDS000199419.2 | 2009 | Tunisia | PRJNA1160997 |
| GCA 043905305 | **HBH_416_10** | Klebsiella_383 | PDS000199419.2 | 2010 | Tunisia | PRJNA1160997 |
| GCA 043904745 | **HBH_30_10** | Klebsiella_383 | PDS000199419.2 | 2010 | Tunisia | PRJNA1160997 |
| GCA 043858235 | **HBH_116_09** | Klebsiella_383 | PDS000199419.2 | 2009 | Tunisia | PRJNA1160997 |
| GCA 028092175 | PDT001585477.1 | Klebsiella_383 | PDS000133910.8 | 2007 | Greece | PRJEB58216 |
| GCA 947662285 | PDT001590394.1 | Klebsiella_383 | PDS000133910.8 | 2007 | Greece | PRJEB58216 |
| GCA 022158485 | PDT001243735.1 | Klebsiella_383 | PDS000133910.8 | 2014 | United Kingdom | PRJNA788733 |
| GCA 001611055 | PDT000123366.1 | Klebsiella_383 | | 2011 | Greece | PRJNA295651 |
| GCA 022099655 | PDT001243972.1 | Klebsiella_383 | PDS000133910.8 | 2014 | United Kingdom | PRJNA788733 |
| GCA 916161915 | PDT001141253.1 | Klebsiella_383 | PDS000133910.8 | 2013 | Netherlands | PRJEB41042 |
| GCA 022268295 | PDT000434786.1 | Klebsiella_383 | | 2012 | Greece | PRJDB4948 |
| GCA 900500925 | PDT000369958.1 | Klebsiella_383 | PDS000007455.4 | 2014 | Greece | PRJEB10018 |
| GCA 001701935 | PDT000144375.1 | Klebsiella_383 | PDS000007455.4 | 2012 | Greece | PRJNA307517 |
| GCA 001701985 | PDT000144378.1 | Klebsiella_383 | PDS000007455.4 | 2013 | Greece | PRJNA307517 |
| GCA 916161705 | PDT001141240.1 | Klebsiella_383 | | 2012 | Netherlands | PRJEB41042 |
| GCA 028090635 | PDT001585554.1 | Klebsiella_383 | PDS000133910.8 | 2015 | Greece | PRJEB58216 |
| GCA 947660785 | PDT001592146.1 | Klebsiella_383 | PDS000133910.8 | 2015 | Greece | PRJEB58216 |
| GCA 916161845 | PDT001141248.1 | Klebsiella_383 | PDS000133910.8 | 2013 | Netherlands | PRJEB41042 |
| GCA 947660355 | PDT001594171.1 | Klebsiella_383 | PDS000133910.8 | 2012 | Greece | PRJEB58216 |
| GCA 028091395 | PDT001585517.1 | Klebsiella_383 | PDS000133910.8 | 2012 | Greece | PRJEB58216 |
| GCA 947660315 | PDT001594129.1 | Klebsiella_383 | PDS000133910.8 | 2012 | Greece | PRJEB58216 |
| GCA 028094055 | PDT001585432.1 | Klebsiella_383 | PDS000133910.8 | 2012 | Greece | PRJEB58216 |
| GCA 900502315 | PDT000369990.1 | Klebsiella_383 | PDS000133910.8 | 2014 | Greece | PRJEB10018 |
| GCA 900500905 | PDT000369957.1 | Klebsiella_383 | PDS000133910.8 | 2014 | Greece | PRJEB10018 |
| GCA 902156555 | PDT000663220.1 | Klebsiella_383 | PDS000133910.8 | 2014 | Greece | PRJEB33308 |
| GCA 028090855 | PDT001585545.1 | Klebsiella_383 | PDS000133910.8 | 2014 | Greece | PRJEB58216 |
| GCA 947663335 | PDT001592164.1 | Klebsiella_383 | PDS000133910.8 | 2014 | Greece | PRJEB58216 |
| GCA 033900045 | PDT002000540.1 | Klebsiella_383 | PDS000133910.8 | nan | Germany | PRJNA1036598 |
| GCA 947662625 | PDT001592161.1 | Klebsiella_383 | PDS000133910.8 | 2018 | Greece | PRJEB58216 |
| GCA 028089315 | PDT001585621.1 | Klebsiella_383 | PDS000133910.8 | 2018 | Greece | PRJEB58216 |
| GCA 028094275 | PDT001585420.1 | Klebsiella_383 | PDS000133910.8 | 2011 | Greece | PRJEB58216 |
| GCA 947660045 | PDT001592162.1 | Klebsiella_383 | PDS000133910.8 | 2011 | Greece | PRJEB58216 |
| GCA 916161185 | PDT001141222.1 | Klebsiella_383 | PDS000133910.8 | 2013 | Netherlands | PRJEB41042 |
| GCA 028091215 | PDT001585526.1 | Klebsiella_383 | PDS000133910.8 | 2013 | Greece | PRJEB58216 |
| GCA 028091235 | PDT001585524.1 | Klebsiella_383 | PDS000133910.8 | 2013 | Greece | PRJEB58216 |
| GCA 001611095 | PDT000123367.1 | Klebsiella_383 | PDS000133910.8 | 2013 | Greece | PRJNA295651 |
| GCA 947662095 | PDT001594164.1 | Klebsiella_383 | PDS000133910.8 | 2013 | Greece | PRJEB58216 |
| GCA 947662275 | PDT001594191.1 | Klebsiella_383 | PDS000133910.8 | 2013 | Greece | PRJEB58216 |
| GCA 028094175 | PDT001585425.1 | Klebsiella_383 | PDS000133910.8 | 2011 | Greece | PRJEB58216 |
| GCA 028091295 | PDT001585521.1 | Klebsiella_383 | PDS000133910.8 | 2013 | Greece | PRJEB58216 |
| GCA 947662185 | PDT001592188.1 | Klebsiella_383 | PDS000133910.8 | 2013 | Greece | PRJEB58216 |
| GCA 947660105 | PDT001592152.1 | Klebsiella_383 | PDS000133910.8 | 2011 | Greece | PRJEB58216 |
| GCA 947660445 | PDT001594195.1 | Klebsiella_383 | PDS000133910.8 | 2012 | Greece | PRJEB58216 |
| GCA 947662295 | PDT001592215.1 | Klebsiella_383 | PDS000133910.8 | 2012 | Greece | PRJEB58216 |
| GCA 028094135 | PDT001585429.1 | Klebsiella_383 | PDS000133910.8 | 2012 | Greece | PRJEB58216 |
| GCA 028094075 | PDT001585431.1 | Klebsiella_383 | PDS000133910.8 | 2012 | Greece | PRJEB58216 |

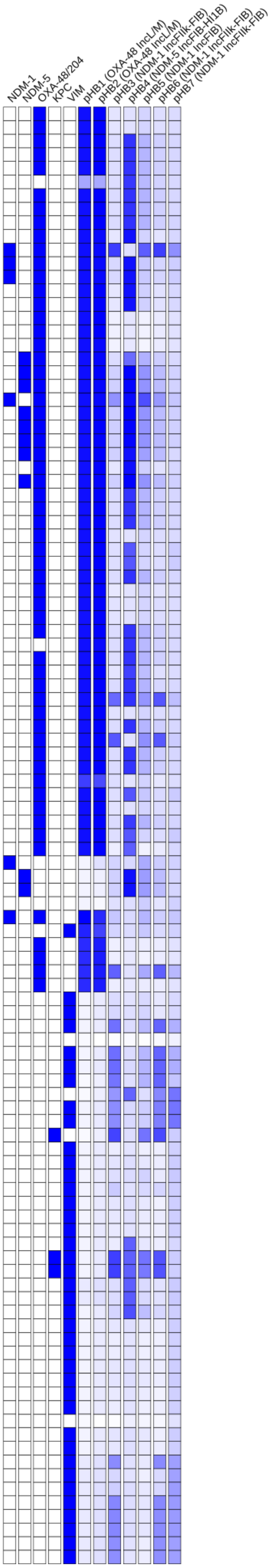

### Tree 32: GCA_043905425

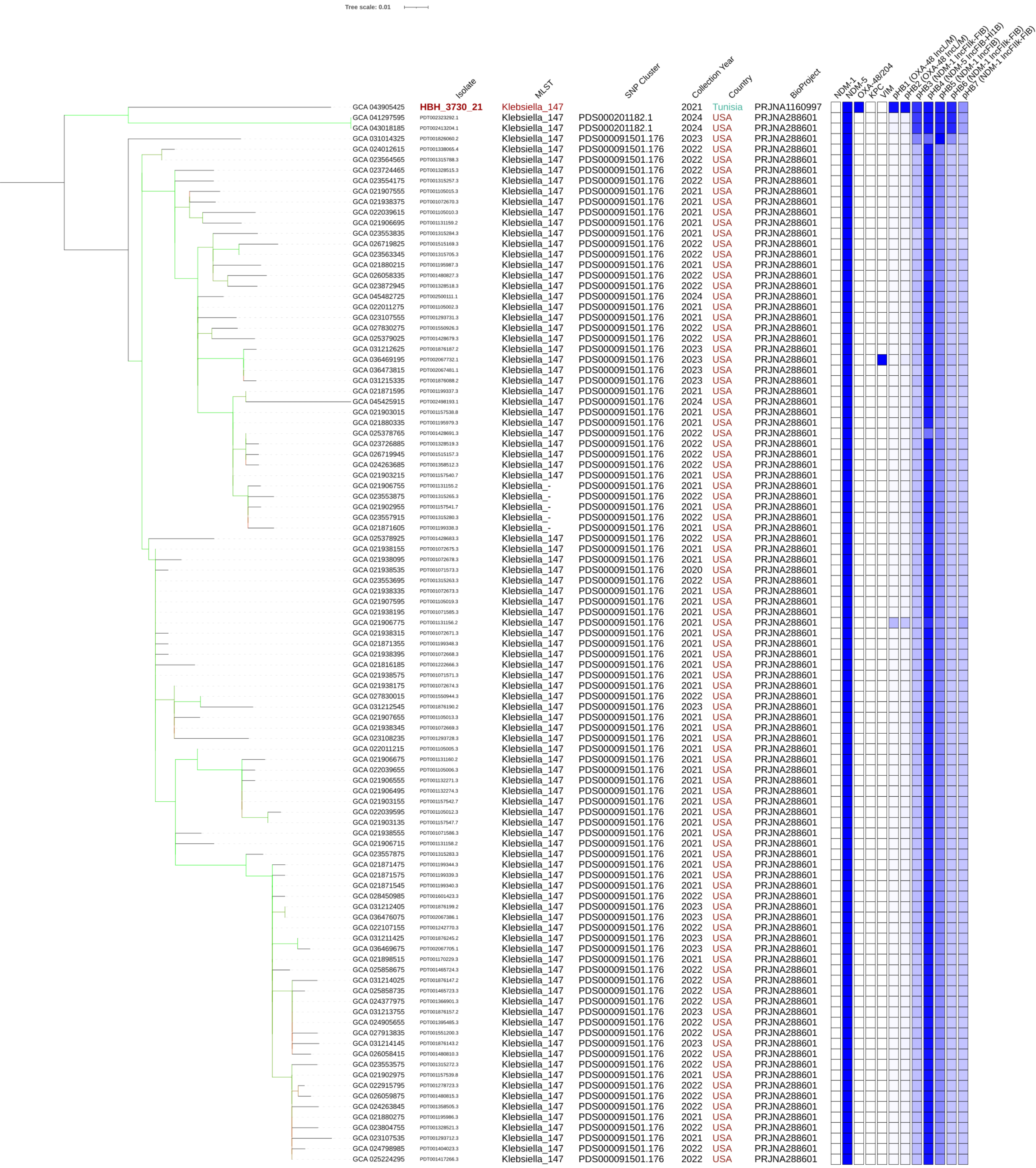

### Tree 33: GCA_043905485

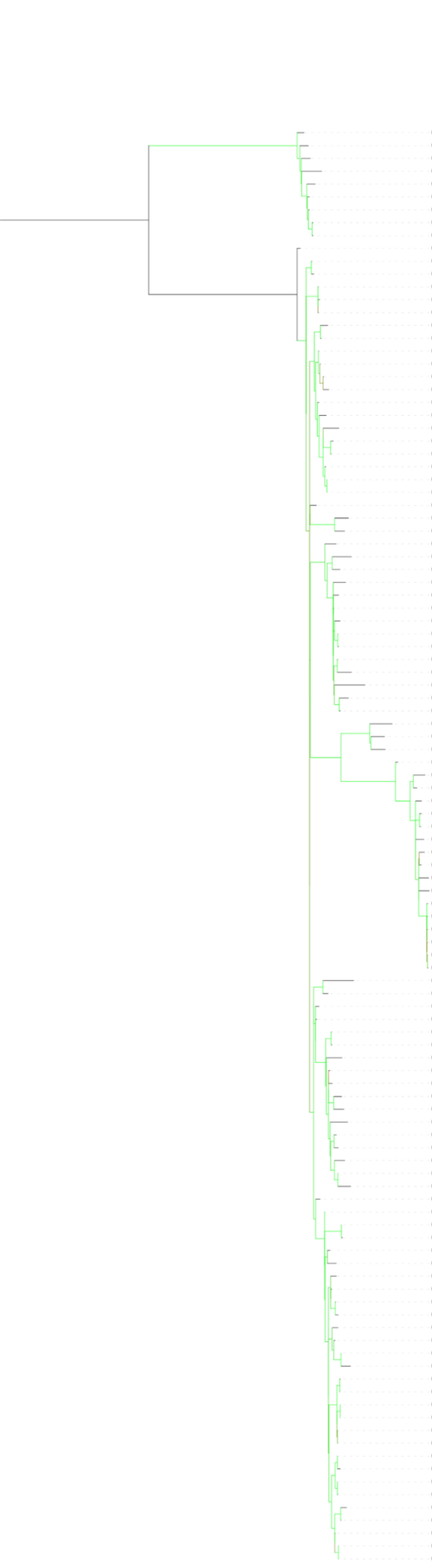


| | Isolate | MLST | SNP Cluster | Collection Year | Country | BioProject |
|---|---|---|---|---|---|---|
| GCA 034490305 | PDT002038556.1 | Klebsiella_101 | | 2019 | France | PRJNA876630 |
| GCA 035017385 | PDT002045108.1 | Klebsiella_101 | | 2022 | Libya | PRJNA998204 |
| GCA 021930655 | PDT001091033.3 | Klebsiella_101 | | 2021 | USA | PRJNA288601 |
| GCA 043904825 | **HBH_4008_18** | Klebsiella_101 | | 2018 | Tunisia | PRJNA1160997 |
| GCA 043905485 | **HBH_957_22** | Klebsiella_101 | PDS000080305.6 | 2022 | Tunisia | PRJNA1160997 |
| GCA 043832795 | PDT002430498.1 | Klebsiella_101 | PDS000080305.6 | 2019 | Germany | PRJNA1070536 |
| GCA 035017265 | PDT002045103.1 | Klebsiella_101 | PDS000080305.6 | 2022 | Libya | PRJNA998204 |
| GCA 016807625 | PDT000949476.1 | Klebsiella_101 | PDS000080305.6 | 2020 | Italy | PRJNA648931 |
| GCA 016807605 | PDT000949475.1 | Klebsiella_101 | PDS000080305.6 | 2020 | Italy | PRJNA648931 |
| GCA 032721215 | PDT001927038.1 | Klebsiella_101 | | 2019 | Italy | PRJNA1026096 |
| GCA 022296695 | PDT000423598.1 | Klebsiella_- | PDS000199425.1 | nan | NA | PRJEB17615 |
| GCA 022295035 | PDT000423690.1 | Klebsiella_- | PDS000199425.1 | nan | NA | PRJEB17615 |
| GCA 018350405 | PDT001033284.1 | Klebsiella_101 | PDS000088521.1 | 2014 | Italy | PRJNA685961 |
| GCA 018350335 | PDT001033281.1 | Klebsiella_101 | PDS000088521.1 | 2014 | Italy | PRJNA685961 |
| GCA 018350395 | PDT001033283.1 | Klebsiella_101 | PDS000088521.1 | 2014 | Italy | PRJNA685961 |
| GCA 002189305 | PDT000217889.1 | Klebsiella_101 | PDS000199425.1 | 2015 | Tunisia | PRJNA360369 |
| GCA 002189255 | PDT000217887.1 | Klebsiella_101 | PDS000199425.1 | 2015 | Tunisia | PRJNA360320 |
| GCA 022987525 | PDT001286506.1 | Klebsiella_101 | PDS000199425.1 | 2013 | Germany | PRJNA789142 |
| GCA 002189335 | PDT000217891.1 | Klebsiella_101 | PDS000199425.1 | 2014 | Tunisia | PRJNA360296 |
| GCA 002189265 | PDT000217888.1 | Klebsiella_101 | PDS000199425.1 | 2014 | Tunisia | PRJNA360291 |
| GCA 021806695 | PDT001227317.1 | Klebsiella_101 | PDS000199425.1 | 2015 | United Kingdom | PRJNA788733 |
| GCA 043860255 | **HBH_417_14** | Klebsiella_101 | PDS000199425.1 | 2014 | Tunisia | PRJNA1160997 |
| GCA 022160985 | PDT001243464.1 | Klebsiella_- | PDS000199425.1 | 2015 | United Kingdom | PRJNA788733 |
| GCA 046351875 | PDT002549677.1 | Klebsiella_101 | | 2024 | USA | PRJNA288601 |
| GCA 043863495 | **HBH_26_17** | Klebsiella_101 | PDS000199425.1 | 2017 | Tunisia | PRJNA1160997 |
| GCA 043863265 | **HBH_4479_16** | Klebsiella_101 | PDS000199425.1 | 2016 | Tunisia | PRJNA1160997 |
| GCA 956481515 | PDT001864169.1 | Klebsiella_101 | PDS000199425.1 | 2016 | Turkey | PRJEB63349 |
| GCA 021136355 | PDT001201698.1 | Klebsiella_101 | PDS000199425.1 | 2018 | Switzerland | PRJNA758223 |
| GCA 021135615 | PDT001201675.1 | Klebsiella_101 | PDS000199425.1 | 2018 | Switzerland | PRJNA759850 |
| GCA 900511565 | PDT000369164.1 | Klebsiella_101 | PDS000199425.1 | 2014 | France | PRJEB10018 |
| GCA 019149405 | PDT001085509.1 | Klebsiella_101 | | 2019 | India | PRJNA548120 |
| GCA 019149365 | PDT001085507.1 | Klebsiella_101 | | 2019 | India | PRJNA548120 |
| GCA 035017165 | PDT002045098.1 | Klebsiella_101 | PDS000041735.6 | 2022 | Libya | PRJNA998204 |
| GCA 043905285 | **HBH_3900_20** | Klebsiella_101 | | 2020 | Tunisia | PRJNA1160997 |
| GCA 900507575 | PDT000370572.1 | Klebsiella_101 | | 2014 | Turkey | PRJEB10018 |
| GCA 043859285 | **HBH_3025_12** | Klebsiella_101 | PDS000041735.6 | 2012 | Tunisia | PRJNA1160997 |
| GCA 043858445 | **HBH_95_11** | Klebsiella_101 | PDS000041735.6 | 2011 | Tunisia | PRJNA1160997 |
| GCA 022241145 | PDT000441621.1 | Klebsiella_101 | PDS000041735.6 | 2010 | Germany | PRJEB18621 |
| GCA 022108075 | PDT001244416.1 | Klebsiella_- | PDS000041735.6 | 2014 | United Kingdom | PRJNA788733 |
| GCA 001913175 | PDT000174387.1 | Klebsiella_101 | PDS000041735.6 | 2013 | Germany | PRJNA329409 |
| GCA 022237495 | PDT000441695.1 | Klebsiella_101 | PDS000041735.6 | 2013 | Germany | PRJEB18621 |
| GCA 043859275 | **HBH_722_12** | Klebsiella_101 | PDS000041735.6 | 2012 | Tunisia | PRJNA1160997 |
| GCA 022116135 | PDT001245102.1 | Klebsiella_101 | | nan | United Kingdom | PRJNA788733 |
| GCA 022237685 | PDT000441692.1 | Klebsiella_101 | | 2013 | Germany | PRJEB18621 |
| GCA 017311775 | PDT000984769.1 | Klebsiella_101 | | 2017 | China | PRJNA648010 |
| GCA 033447315 | PDT001974683.1 | Klebsiella_101 | PDS000041735.6 | 2012 | China | PRJNA1035291 |
| GCA 900506735 | PDT000370529.1 | Klebsiella_101 | | 2014 | Belgium | PRJEB10018 |
| GCA 032720335 | PDT001927077.1 | Klebsiella_101 | PDS000199425.1 | 2020 | Italy | PRJNA1026096 |
| GCA 956481655 | PDT001864141.1 | Klebsiella_101 | | 2017 | Turkey | PRJEB63349 |
| GCA 022240915 | PDT000441625.1 | Klebsiella_101 | PDS000041735.6 | 2011 | Germany | PRJEB18621 |
| GCA 902704805 | PDT000662758.1 | Klebsiella_101 | | 2016 | Italy | PRJEB22890 |
| GCA 900514115 | PDT000369585.1 | Klebsiella_101 | PDS000045320.1 | 2013 | Italy | PRJEB10018 |
| GCA 902703785 | PDT000662659.1 | Klebsiella_101 | PDS000045311.30 | 2016 | Italy | PRJEB22890 |
| GCA 018907275 | PDT001072374.1 | Klebsiella_101 | PDS000045311.30 | 2012 | Italy | PRJNA608247 |
| GCA 018907315 | PDT001072375.1 | Klebsiella_101 | PDS000045311.30 | 2013 | Italy | PRJNA608247 |
| GCA 003227515 | PDT000331482.1 | Klebsiella_101 | PDS000045312.1 | 2016 | Italy | PRJNA431724 |
| GCA 022292435 | PDT000427105.1 | Klebsiella_101 | PDS000045311.30 | nan | NA | PRJEB10018 |
| GCA 017974755 | PDT001007066.1 | Klebsiella_101 | PDS000045311.30 | 2017 | Italy | PRJNA687927 |
| GCA 902703695 | PDT000662653.1 | Klebsiella_101 | PDS000055633.53 | 2016 | Italy | PRJEB22890 |
| GCA 900514255 | PDT000369440.1 | Klebsiella_101 | PDS000045311.30 | 2013 | Italy | PRJEB10018 |
| GCA 003227495 | PDT000331481.1 | Klebsiella_2502 | PDS000045311.30 | 2016 | Italy | PRJNA431724 |
| GCA 003227475 | PDT000331480.1 | Klebsiella_2502 | PDS000045311.30 | 2016 | Italy | PRJNA431724 |
| GCA 003227455 | PDT000331479.1 | Klebsiella_2502 | PDS000045311.30 | 2016 | Italy | PRJNA431724 |
| GCA 003227175 | PDT000331467.1 | Klebsiella_2502 | PDS000045311.30 | 2016 | Italy | PRJNA431724 |
| GCA 003227155 | PDT000331465.1 | Klebsiella_2502 | PDS000045311.30 | 2016 | Italy | PRJNA431724 |
| GCA 003227105 | PDT000331462.1 | Klebsiella_2502 | PDS000045311.30 | 2016 | Italy | PRJNA431724 |
| GCA 022185645 | PDT000716609.1 | Klebsiella_101 | | 2018 | France | PRJEB35890 |
| GCA 022132325 | PDT001244810.1 | Klebsiella_101 | | 2014 | United Kingdom | PRJNA788733 |
| GCA 022228745 | PDT000446086.1 | Klebsiella_101 | PDS000199425.1 | 2015 | Germany | PRJEB18059 |
| GCA 043859655 | **HBH_2166_13** | Klebsiella_101 | PDS000199425.1 | 2013 | Tunisia | PRJNA1160997 |
| GCA 043863045 | **HBH_4442_15** | Klebsiella_101 | PDS000188993.3 | 2015 | Tunisia | PRJNA1160997 |
| GCA 043863225 | **HBH_275_16** | Klebsiella_101 | PDS000188993.3 | 2016 | Tunisia | PRJNA1160997 |
| GCA 922829475 | PDT001211067.1 | Klebsiella_101 | PDS000188993.3 | 2015 | United Kingdom | PRJEB48990 |
| GCA 046127015 | PDT002538802.1 | Klebsiella_101 | PDS000188993.3 | 2015 | New Zealand | PRJNA1102395 |
| GCA 022162785 | PDT001243259.1 | Klebsiella_101 | PDS000188993.3 | 2015 | United Kingdom | PRJNA788733 |
| GCA 956478775 | PDT001861761.1 | Klebsiella_3367 | | 2017 | Romania | PRJEB63349 |
| GCA 956476795 | PDT001861753.1 | Klebsiella_101 | | 2017 | Romania | PRJEB63349 |
| GCA 009710205 | PDT000637199.1 | Klebsiella_101 | | 2018/2019 | Romania | PRJNA579879 |
| GCA 021787895 | PDT001227768.1 | Klebsiella_101 | PDS000188993.3 | 2015 | United Kingdom | PRJNA788733 |
| GCA 040015615 | PDT002187526.1 | Klebsiella_101 | PDS000188993.3 | 2015 | Netherlands | PRJNA1116077 |
| GCA 956479425 | PDT001863967.2 | Klebsiella_101 | | 2017 | Romania | PRJEB63349 |
| GCA 009710605 | PDT000637218.1 | Klebsiella_101 | PDS000188993.3 | 2018/2019 | Romania | PRJNA579879 |
| GCA 009710685 | PDT000637223.1 | Klebsiella_101 | PDS000188993.3 | 2018/2019 | Romania | PRJNA579879 |
| GCA 900512775 | PDT000369255.1 | Klebsiella_101 | PDS000199425.1 | 2014 | Germany | PRJEB10018 |
| GCA 900502305 | PDT000369989.1 | Klebsiella_101 | PDS000199424.2 | 2014 | Greece | PRJEB10018 |
| GCA 028089835 | PDT001585588.1 | Klebsiella_101 | PDS000199424.2 | 2017 | Greece | PRJEB58216 |
| GCA 947662125 | PDT001592247.1 | Klebsiella_101 | PDS000199424.2 | 2017 | Greece | PRJEB58216 |
| GCA 003034545 | PDT000304395.1 | Klebsiella_101 | PDS000199424.2 | 2015 | Greece | PRJNA419295 |
| GCA 003034525 | PDT000304394.1 | Klebsiella_101 | PDS000199424.2 | 2015 | Greece | PRJNA419295 |
| GCA 003034395 | PDT000304387.1 | Klebsiella_101 | PDS000199424.2 | 2015 | Greece | PRJNA419295 |
| GCA 003034445 | PDT000304391.1 | Klebsiella_101 | PDS000199424.2 | 2015 | Greece | PRJNA419295 |
| GCA 003034425 | PDT000304389.1 | Klebsiella_101 | PDS000199424.2 | 2016 | Greece | PRJNA419295 |
| GCA 003034505 | PDT000304393.1 | Klebsiella_101 | PDS000199424.2 | 2016 | Greece | PRJNA419295 |
| GCA 043933365 | PDT002412058.1 | Klebsiella_101 | PDS000199424.2 | 2016 | Canada | PRJNA855907 |
| GCA 009826695 | PDT000663153.1 | Klebsiella_101 | PDS000199424.2 | 2016 | Greece | PRJNA597427 |
| GCA 043626345 | PDT002332935.1 | Klebsiella_101 | PDS000199424.2 | nan | Netherlands | PRJEB55414 |
| GCA 956478275 | PDT001861514.1 | Klebsiella_101 | PDS000199424.2 | 2018 | Greece | PRJEB63349 |
| GCA 028090535 | PDT001585562.1 | Klebsiella_101 | PDS000199424.2 | 2015 | Greece | PRJEB58216 |
| GCA 947659995 | PDT001594126.1 | Klebsiella_101 | PDS000199424.2 | 2015 | Greece | PRJEB58216 |
| GCA 028090175 | PDT001585563.1 | Klebsiella_101 | PDS000199424.2 | 2016 | Greece | PRJEB58216 |
| GCA 947662565 | PDT001594138.1 | Klebsiella_101 | PDS000199424.2 | 2016 | Greece | PRJEB58216 |
| GCA 028090115 | PDT001585559.1 | Klebsiella_101 | PDS000199424.2 | 2015 | Greece | PRJEB58216 |
| GCA 947660135 | PDT001594175.1 | Klebsiella_101 | PDS000199424.2 | 2015 | Greece | PRJEB58216 |
| GCA 900607635 | PDT000411391.1 | Klebsiella_101 | PDS000199424.2 | 2016 | Switzerland | PRJEB28660 |
| GCA 020119235 | PDT001137053.1 | Klebsiella_101 | PDS000199424.2 | 2016 | Switzerland | PRJNA543274 |
| GCA 028090375 | PDT001585579.1 | Klebsiella_101 | PDS000199424.2 | 2016 | Greece | PRJEB58216 |
| GCA 947662345 | PDT001592169.1 | Klebsiella_101 | PDS000199424.2 | 2016 | Greece | PRJEB58216 |
| GCA 044335685 | PDT002305468.1 | Klebsiella_101 | PDS000199424.2 | nan | USA | PRJNA271899 |
| GCA 034563395 | PDT002037997.1 | Klebsiella_101 | PDS000199424.2 | 2023 | USA | PRJNA288601 |
| GCA 009826665 | PDT000663152.1 | Klebsiella_101 | PDS000199424.2 | 2015 | Greece | PRJNA597427 |
| GCA 028089935 | PDT001585574.1 | Klebsiella_101 | PDS000199424.2 | 2016 | Greece | PRJEB58216 |
| GCA 947660705 | PDT001590391.1 | Klebsiella_101 | PDS000199424.2 | 2016 | Greece | PRJEB58216 |

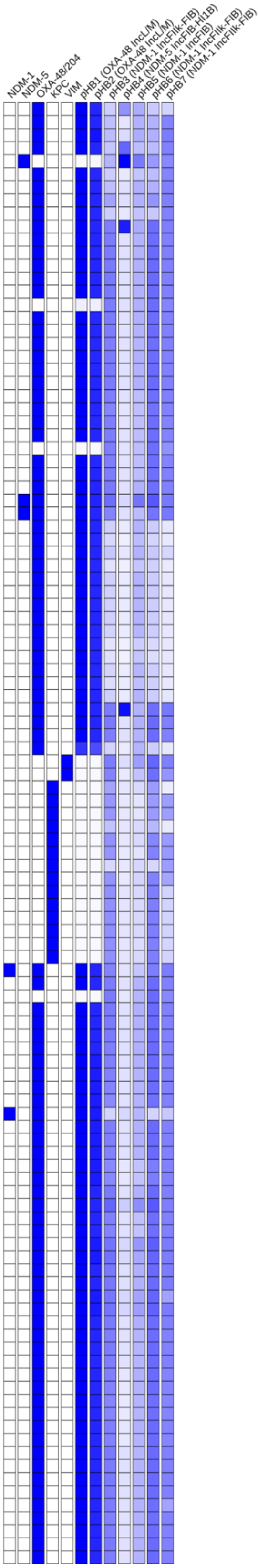

### Tree 34: GCA_043905525

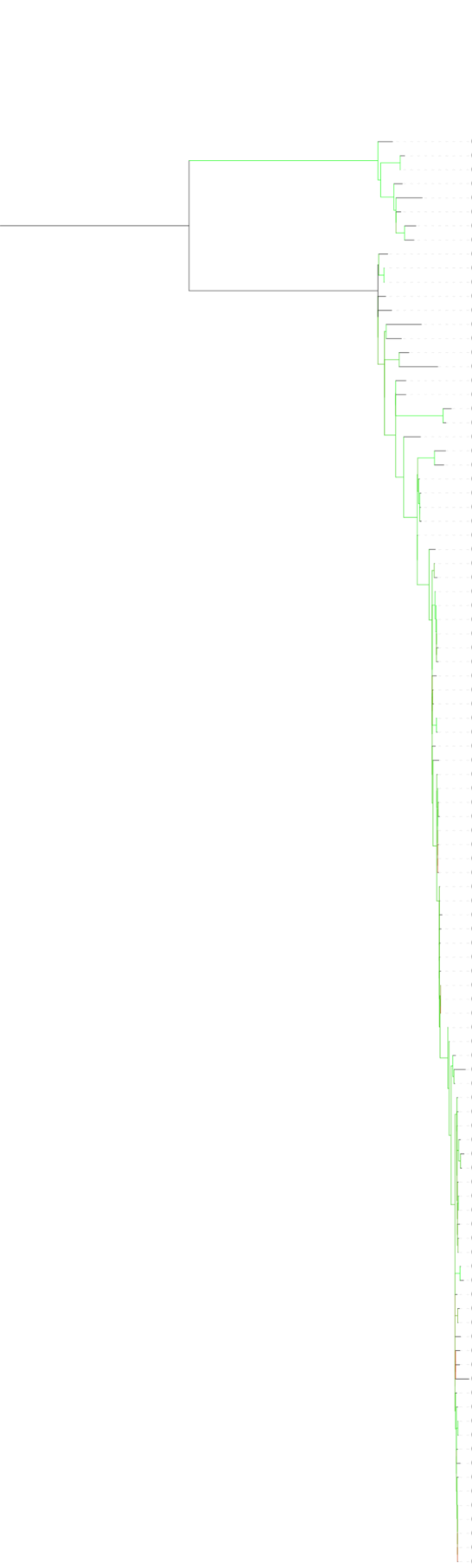


| Accession | Isolate | MLST | SNP Cluster | Collection Year | Country | BioProject |
|---|---|---|---|---|---|---|
| GCA 022306535 | PDT000176097.2 | Klebsiella_13 | | nan | NA | PRJEB5065 |
| GCA 900503065 | PDT000370229.1 | Klebsiella_13 | PDS000098778.2 | 2014 | United Kingdom | PRJEB10018 |
| GCA 022194405 | PDT000715140.1 | Klebsiella_- | PDS000098778.2 | 2014 | United Kingdom | PRJNA564424 |
| GCA 045752805 | PDT002512410.1 | Klebsiella_13 | PDS000173925.11 | 2024 | USA | PRJNA288601 |
| GCA 036811575 | PDT002084576.1 | Klebsiella_13 | PDS000053017.24 | 2024 | USA | PRJNA288601 |
| GCA 024430215 | PDT001366426.1 | Klebsiella_13 | PDS000053017.24 | nan | USA | PRJNA825705 |
| GCA 900508405 | PDT000370632.1 | Klebsiella_13 | PDS000053017.24 | 2013 | Turkey | PRJEB10018 |
| GCA 045662225 | PDT002505954.1 | Klebsiella_13 | PDS000211070.1 | 2024 | USA | PRJNA288601 |
| GCA 024489195 | PDT001373413.1 | Klebsiella_13 | | 2019 | Spain | PRJEB50822 |
| GCA 041765315 | PDT002366259.1 | Klebsiella_13 | PDS000197052.1 | 2015 | Norway | PRJNA1098507 |
| GCA 033122965 | PDT001958059.1 | Klebsiella_13 | PDS000197052.1 | 2015 | Norway | PRJEB27256 |
| GCA 031038295 | PDT001717196.3 | Klebsiella_13 | | 2023 | USA | PRJNA288601 |
| GCA 013604195 | PDT000797598.1 | Klebsiella_13 | | nan | Spain | PRJNA609897 |
| GCA 021816085 | PDT001219145.1 | Klebsiella_13 | | 2018 | Germany | PRJEB43549 |
| GCA 043905525 | **HBH_1304_11** | Klebsiella_13 | | 2011 | Tunisia | PRJNA1160997 |
| GCA 022313435 | PDT000136299.2 | Klebsiella_13 | | 2010 | United Kingdom | PRJNA306133 |
| GCA 905331375 | PDT001185214.2 | Klebsiella_13 | | 2018 | Spain | PRJEB42440 |
| GCA 044382055 | PDT002456491.1 | Klebsiella_13 | | nan | Saudi Arabia | PRJNA1116866 |
| GCA 900513595 | PDT000369532.1 | Klebsiella_13 | | 2014 | Israel | PRJEB10018 |
| GCA 041423215 | PDT002335078.1 | Klebsiella_13 | PDS000194059.1 | 2017 | Portugal | PRJNA986835 |
| GCA 041422095 | PDT002335021.1 | Klebsiella_13 | PDS000194059.1 | 2017 | Portugal | PRJNA986835 |
| GCA 008374255 | PDT000586403.1 | Klebsiella_13 | | 2018 | France | PRJNA563544 |
| GCA 008374425 | PDT000586411.1 | Klebsiella_13 | PDS000056122.9 | 2018 | France | PRJNA563544 |
| GCA 008374895 | PDT000586854.1 | Klebsiella_13 | PDS000056122.9 | 2018 | France | PRJNA563544 |
| GCA 041422315 | PDT002335032.1 | Klebsiella_13 | PDS000056122.9 | 2017 | Portugal | PRJNA986835 |
| GCA 041421545 | PDT002334995.1 | Klebsiella_13 | PDS000056122.9 | 2017 | Portugal | PRJNA986835 |
| GCA 041421595 | PDT002334998.1 | Klebsiella_13 | PDS000056122.9 | 2017 | Portugal | PRJNA986835 |
| GCA 041422115 | PDT002335022.1 | Klebsiella_13 | PDS000056122.9 | 2017 | Portugal | PRJNA986835 |
| GCA 041423635 | PDT002335122.1 | Klebsiella_13 | PDS000056122.9 | 2017 | Portugal | PRJNA986835 |
| GCA 008374475 | PDT000586413.1 | Klebsiella_13 | PDS000056122.9 | 2018 | France | PRJNA563544 |
| GCA 041423075 | PDT002335070.1 | Klebsiella_13 | PDS000056122.9 | 2017 | Portugal | PRJNA986835 |
| GCA 041422815 | PDT002335057.1 | Klebsiella_13 | PDS000056122.9 | 2017 | Portugal | PRJNA986835 |
| GCA 041424975 | PDT002335179.1 | Klebsiella_13 | PDS000056122.9 | 2017 | Portugal | PRJNA986835 |
| GCA 041424775 | PDT002335169.1 | Klebsiella_13 | PDS000056122.9 | 2017 | Portugal | PRJNA986835 |
| GCA 041421345 | PDT002334985.1 | Klebsiella_13 | PDS000056122.9 | 2017 | Portugal | PRJNA986835 |
| GCA 041424935 | PDT002335177.1 | Klebsiella_13 | PDS000056122.9 | 2017 | Portugal | PRJNA986835 |
| GCA 041422035 | PDT002335018.1 | Klebsiella_13 | PDS000056122.9 | 2017 | Portugal | PRJNA986835 |
| GCA 041421445 | PDT002334990.1 | Klebsiella_13 | PDS000056122.9 | 2017 | Portugal | PRJNA986835 |
| GCA 041421695 | PDT002335003.1 | Klebsiella_- | PDS000056122.9 | 2017 | Portugal | PRJNA986835 |
| GCA 016643445 | PDT000933591.1 | Klebsiella_13 | PDS000056122.9 | nan | Portugal | PRJNA602991 |
| GCA 016643345 | PDT000933586.1 | Klebsiella_13 | PDS000056122.9 | nan | Portugal | PRJNA602991 |
| GCA 025878685 | PDT001467903.1 | Klebsiella_13 | PDS000056122.9 | 2018 | Norway | PRJEB56146 |
| GCA 947044045 | PDT001449714.1 | Klebsiella_13 | PDS000056122.9 | nan | NA | PRJEB41590 |
| GCA 041421935 | PDT002335013.1 | Klebsiella_13 | PDS000056122.9 | 2017 | Portugal | PRJNA986835 |
| GCA 041423615 | PDT002335121.1 | Klebsiella_13 | PDS000056122.9 | 2017 | Portugal | PRJNA986835 |
| GCA 041421995 | PDT002335016.1 | Klebsiella_13 | PDS000056122.9 | 2017 | Portugal | PRJNA986835 |
| GCA 041422935 | PDT002335063.1 | Klebsiella_13 | PDS000056122.9 | 2017 | Portugal | PRJNA986835 |
| GCA 041422895 | PDT002335061.1 | Klebsiella_13 | PDS000056122.9 | 2017 | Portugal | PRJNA986835 |
| GCA 947044275 | PDT001449702.1 | Klebsiella_13 | PDS000056122.9 | nan | NA | PRJEB41590 |
| GCA 041422995 | PDT002335066.1 | Klebsiella_13 | PDS000056122.9 | 2017 | Portugal | PRJNA986835 |
| GCA 947043995 | PDT001449707.1 | Klebsiella_13 | PDS000056122.9 | nan | NA | PRJEB41590 |
| GCA 041422875 | PDT002335060.1 | Klebsiella_13 | PDS000056122.9 | 2017 | Portugal | PRJNA986835 |
| GCA 947044465 | PDT001449703.1 | Klebsiella_13 | PDS000056122.9 | nan | NA | PRJEB41590 |
| GCA 041422755 | PDT002335054.1 | Klebsiella_13 | PDS000056122.9 | 2017 | Portugal | PRJNA986835 |
| GCA 016643325 | PDT000933585.1 | Klebsiella_13 | PDS000056122.9 | nan | Portugal | PRJNA602991 |
| GCA 947044485 | PDT001449716.1 | Klebsiella_13 | PDS000056122.9 | nan | NA | PRJEB41590 |
| GCA 041422695 | PDT002335051.1 | Klebsiella_13 | PDS000056122.9 | 2017 | Portugal | PRJNA986835 |
| GCA 041422655 | PDT002335049.1 | Klebsiella_13 | PDS000056122.9 | 2017 | Portugal | PRJNA986835 |
| GCA 041424895 | PDT002335176.1 | Klebsiella_13 | PDS000056122.9 | 2017 | Portugal | PRJNA986835 |
| GCA 041423095 | PDT002335071.1 | Klebsiella_13 | PDS000056122.9 | 2017 | Portugal | PRJNA986835 |
| GCA 947044315 | PDT001449713.1 | Klebsiella_13 | PDS000056122.9 | nan | NA | PRJEB41590 |
| GCA 041422475 | PDT002335040.1 | Klebsiella_13 | PDS000056122.9 | 2017 | Portugal | PRJNA986835 |
| GCA 947044035 | PDT001449717.1 | Klebsiella_13 | PDS000056122.9 | nan | NA | PRJEB41590 |
| GCA 041422495 | PDT002335041.1 | Klebsiella_13 | PDS000056122.9 | 2017 | Portugal | PRJNA986835 |
| GCA 041423155 | PDT002335074.1 | Klebsiella_13 | PDS000056122.9 | 2017 | Portugal | PRJNA986835 |
| GCA 041422675 | PDT002335050.1 | Klebsiella_13 | PDS000056122.9 | 2017 | Portugal | PRJNA986835 |
| GCA 008374405 | PDT000586410.1 | Klebsiella_13 | PDS000056122.9 | 2018 | France | PRJNA563544 |
| GCA 008374205 | PDT000586400.1 | Klebsiella_13 | PDS000056122.9 | 2018 | France | PRJNA563544 |
| GCA 041423735 | PDT002335129.1 | Klebsiella_13 | PDS000056122.9 | 2017 | Portugal | PRJNA986835 |
| GCA 041423715 | PDT002335128.1 | Klebsiella_13 | PDS000056122.9 | 2017 | Portugal | PRJNA986835 |
| GCA 041423515 | PDT002335110.1 | Klebsiella_13 | PDS000056122.9 | 2017 | Portugal | PRJNA986835 |
| GCA 041423695 | PDT002335127.1 | Klebsiella_13 | PDS000056122.9 | 2017 | Portugal | PRJNA986835 |
| GCA 008374515 | PDT000586415.1 | Klebsiella_13 | PDS000056122.9 | 2018 | France | PRJNA563544 |
| GCA 041421915 | PDT002335012.1 | Klebsiella_13 | PDS000056122.9 | 2017 | Portugal | PRJNA986835 |
| GCA 041423495 | PDT002335108.1 | Klebsiella_13 | PDS000056122.9 | 2017 | Portugal | PRJNA986835 |
| GCA 041422375 | PDT002335039.1 | Klebsiella_13 | PDS000056122.9 | 2017 | Portugal | PRJNA986835 |
| GCA 041423395 | PDT002335098.1 | Klebsiella_13 | PDS000056122.9 | 2017 | Portugal | PRJNA986835 |
| GCA 041422415 | PDT002335036.1 | Klebsiella_13 | PDS000056122.9 | 2017 | Portugal | PRJNA986835 |
| GCA 041422255 | PDT002335029.1 | Klebsiella_13 | PDS000056122.9 | 2017 | Portugal | PRJNA986835 |
| GCA 041422355 | PDT002335034.1 | Klebsiella_13 | PDS000056122.9 | 2017 | Portugal | PRJNA986835 |
| GCA 041423035 | PDT002335068.1 | Klebsiella_13 | PDS000056122.9 | 2017 | Portugal | PRJNA986835 |
| GCA 947044455 | PDT001449709.1 | Klebsiella_13 | PDS000056122.9 | nan | NA | PRJEB41590 |
| GCA 016643475 | PDT000933593.1 | Klebsiella_13 | PDS000056122.9 | nan | Portugal | PRJNA602991 |
| GCA 041422975 | PDT002335065.1 | Klebsiella_13 | PDS000056122.9 | 2017 | Portugal | PRJNA986835 |
| GCA 041422775 | PDT002335055.1 | Klebsiella_13 | PDS000056122.9 | 2017 | Portugal | PRJNA986835 |
| GCA 041424655 | PDT002335163.1 | Klebsiella_13 | PDS000056122.9 | 2017 | Portugal | PRJNA986835 |
| GCA 041423815 | PDT002335133.1 | Klebsiella_13 | PDS000056122.9 | 2017 | Portugal | PRJNA986835 |
| GCA 016643375 | PDT000933588.1 | Klebsiella_13 | PDS000056122.9 | nan | Portugal | PRJNA602991 |
| GCA 041425055 | PDT002335183.1 | Klebsiella_13 | PDS000056122.9 | 2017 | Portugal | PRJNA986835 |
| GCA 041422905 | PDT002335062.1 | Klebsiella_13 | PDS000056122.9 | 2017 | Portugal | PRJNA986835 |
| GCA 041422955 | PDT002335064.1 | Klebsiella_13 | PDS000056122.9 | 2017 | Portugal | PRJNA986835 |
| GCA 947044055 | PDT001449700.1 | Klebsiella_13 | PDS000056122.9 | nan | NA | PRJEB41590 |
| GCA 041422555 | PDT002335044.1 | Klebsiella_13 | PDS000056122.9 | 2017 | Portugal | PRJNA986835 |
| GCA 041423055 | PDT002335069.1 | Klebsiella_13 | PDS000056122.9 | 2017 | Portugal | PRJNA986835 |
| GCA 947044015 | PDT001449706.1 | Klebsiella_13 | PDS000056122.9 | nan | NA | PRJEB41590 |
| GCA 041422515 | PDT002335042.1 | Klebsiella_13 | PDS000056122.9 | 2017 | Portugal | PRJNA986835 |
| GCA 041422835 | PDT002335058.1 | Klebsiella_13 | PDS000056122.9 | 2017 | Portugal | PRJNA986835 |
| GCA 041422795 | PDT002335056.1 | Klebsiella_13 | PDS000056122.9 | 2017 | Portugal | PRJNA986835 |
| GCA 041422635 | PDT002335048.1 | Klebsiella_13 | PDS000056122.9 | 2017 | Portugal | PRJNA986835 |
| GCA 041422725 | PDT002335053.1 | Klebsiella_13 | PDS000056122.9 | 2017 | Portugal | PRJNA986835 |
| GCA 041422715 | PDT002335052.1 | Klebsiella_13 | PDS000056122.9 | 2017 | Portugal | PRJNA986835 |
| GCA 947044345 | PDT001449710.1 | Klebsiella_13 | PDS000056122.9 | nan | NA | PRJEB41590 |

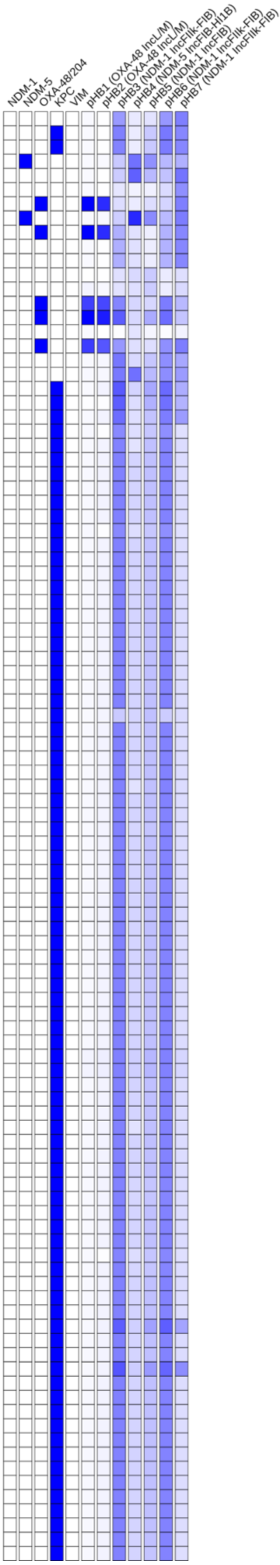

### Tree 35: GCA_043905545

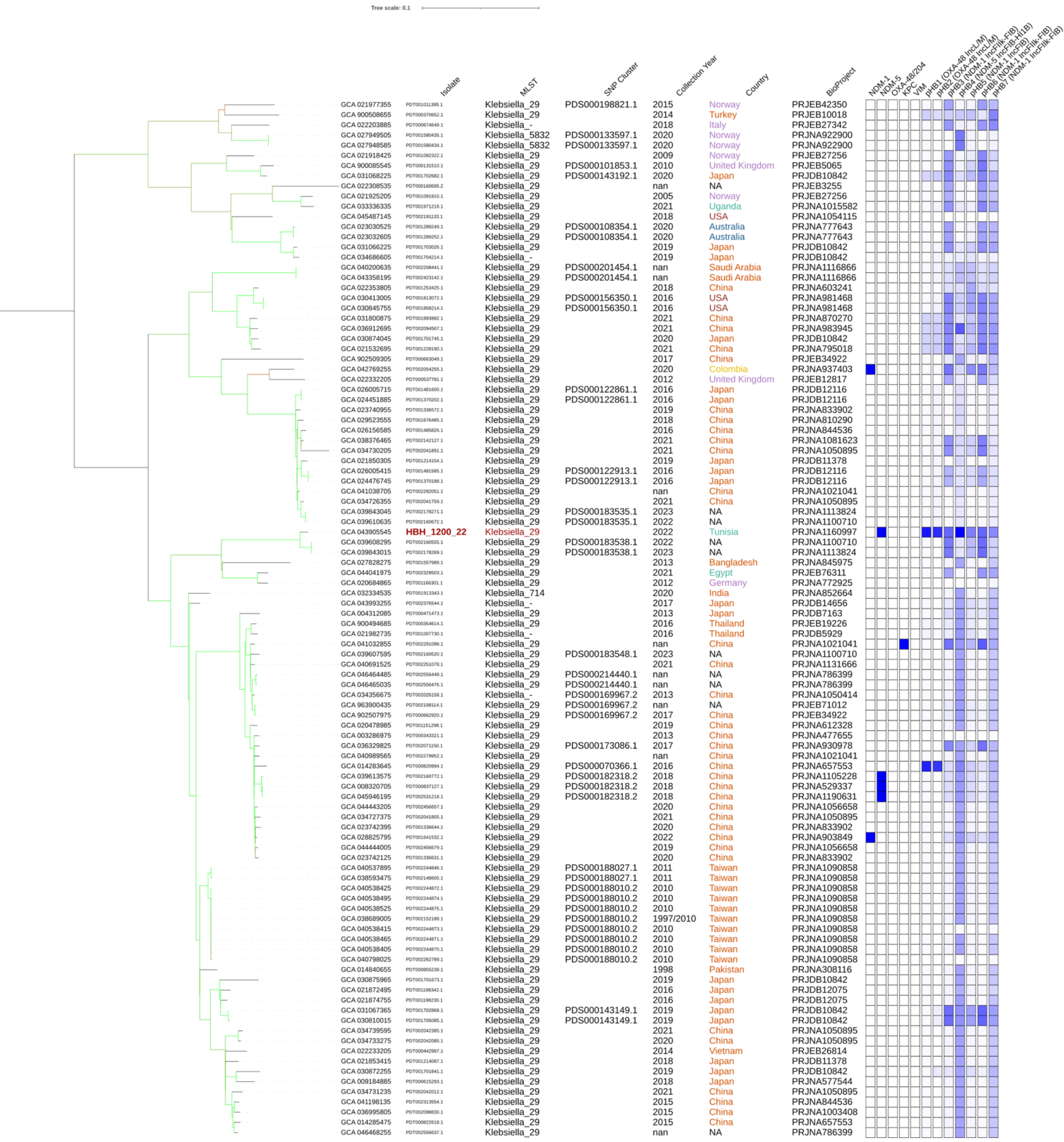

### Tree 36: GCA_043905565

Tree scale: 0.1

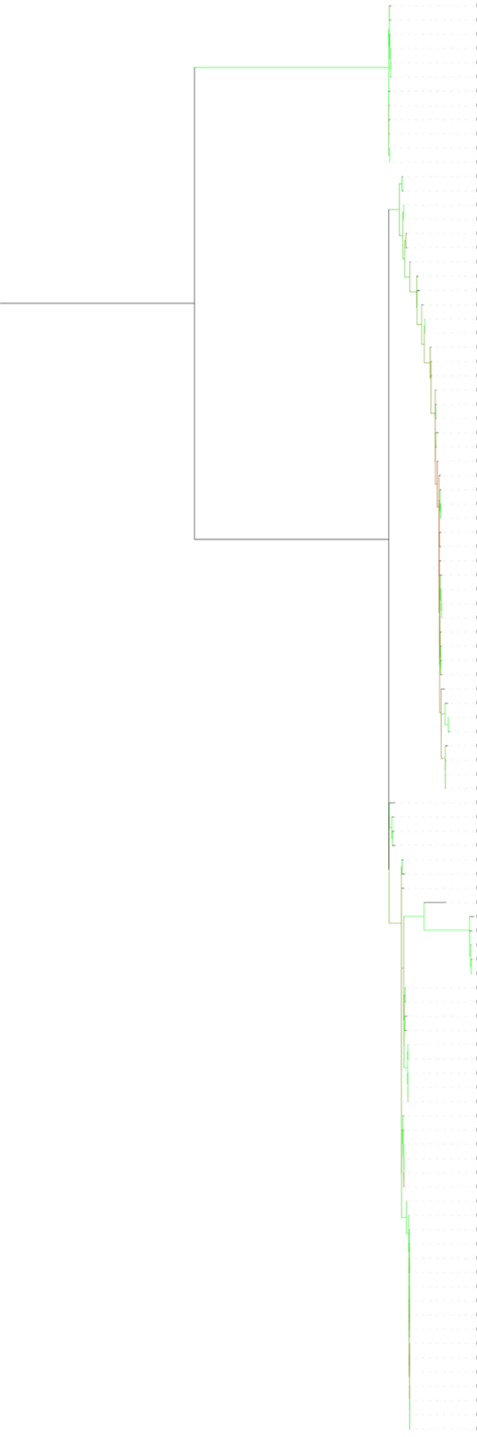

| | Isolate | MLST | SNP Cluster | Collection Year | Country | BioProject |
|---|---|---|---|---|---|---|
| GCA 002186835 | PDT000217809.1 | Klebsiella_1418 | | 2011 | Nigeria | PRJNA351846 |
| GCA 026672225 | PDT001518703.1 | Klebsiella_1418 | | 2022 | Ghana | PRJNA906096 |
| GCA 021136215 | PDT001201693.1 | Klebsiella_1418 | PDS000075208.5 | 2017 | Switzerland | PRJNA758223 |
| GCA 037051645 | PDT002102057.1 | Klebsiella_1418 | PDS000075208.5 | 2013 | Jordan | PRJNA1076682 |
| GCA 022055125 | PDT000879595.1 | Klebsiella_1418 | PDS000075208.5 | 2017 | Qatar | PRJNA656934 |
| GCA 042914675 | PDT002097740.1 | Klebsiella_1418 | PDS000075208.5 | 2022 | Netherlands | PRJNA1076808 |
| GCA 040430465 | PDT002237476.1 | Klebsiella_1418 | | 2021 | Ireland | PRJNA1126319 |
| GCA 043905565 | **HBH_61_21** | Klebsiella_1418 | | 2021 | Tunisia | PRJNA1160997 |
| GCA 900513125 | PDT000369333.1 | Klebsiella_1418 | | 2013 | Italy | PRJEB10018 |
| GCA 015731075 | PDT000903683.1 | Klebsiella_1418 | | 2019 | China | PRJNA665928 |
| GCA 033547775 | PDT001982204.1 | Klebsiella_1418 | PDS000208562.1 | 2022 | Saudi Arabia | PRJNA1018815 |
| GCA 045063005 | PDT002487922.1 | Klebsiella_1418 | PDS000208562.1 | 2022 | Saudi Arabia | PRJNA1018815 |
| GCA 021940255 | PDT001049085.1 | Klebsiella_3098 | PDS000074703.6 | 2018 | Australia | PRJNA857533 |
| GCA 026256345 | PDT001849263.1 | Klebsiella_3098 | PDS000074703.6 | 2013 | Australia | PRJNA646837 |
| GCA 041920645 | PDT002382459.2 | Klebsiella_45 | PDS000198705.1 | 2007 | Norway | PRJNA1098507 |
| GCA 043384065 | PDT002281230.1 | Klebsiella_45 | PDS000198705.1 | 2007 | Norway | PRJEB27256 |
| GCA 000826795 | PDT000049482.1 | Klebsiella_45 | | 2012 | USA | PRJNA261775 |
| GCA 004322955 | PDT000473852.1 | Klebsiella_45 | | 2014 | USA | PRJNA279669 |
| GCA 033103885 | PDT001961101.1 | Klebsiella_- | PDS000106393.3 | 2007 | Spain | PRJEB41835 |
| GCA 046126295 | PDT002538872.1 | Klebsiella_45 | PDS000212921.2 | 2018 | New Zealand | PRJNA1102395 |
| GCA 022248895 | PDT000440765.1 | Klebsiella_- | | nan | NA | PRJEB21605 |
| GCA 033104365 | PDT001961051.1 | Klebsiella_45 | | 2019 | USA | PRJNA648389 |
| GCA 046448265 | PDT002554279.1 | Klebsiella_45 | PDS000214277.1 | 2014 | Russia | PRJNA1196851 |
| GCA 046449325 | PDT002554332.1 | Klebsiella_45 | PDS000214277.1 | 2018 | Russia | PRJNA1196851 |
| GCA 036995725 | PDT002098826.1 | Klebsiella_45 | | 2016 | China | PRJNA1003408 |
| GCA 022184005 | PDT000723800.1 | Klebsiella_45 | | 2018 | Italy | PRJEB27342 |
| GCA 044322325 | PDT002306240.1 | Klebsiella_45 | | nan | USA | PRJNA271899 |
| GCA 964300705 | PDT002500299.1 | Klebsiella_45 | PDS000106675.2 | 2019 | Germany | PRJEB81699 |
| GCA 021975415 | PDT001011543.1 | Klebsiella_45 | | 2015 | Norway | PRJEB42350 |
| GCA 956480335 | PDT001864109.1 | Klebsiella_45 | | 2018 | Spain | PRJEB63349 |
| GCA 022098395 | PDT001096792.3 | Klebsiella_45 | | 2021 | USA | PRJNA288601 |
| GCA 023739135 | PDT001336493.1 | Klebsiella_45 | | 2021 | China | PRJNA833902 |
| GCA 022987545 | PDT001286508.1 | Klebsiella_45 | | 2012 | Germany | PRJNA789142 |
| GCA 032134355 | PDT001903162.1 | Klebsiella_45 | | 2017 | Norway | PRJEB48268 |
| GCA 043446965 | PDT002421688.1 | Klebsiella_45 | PDS000179055.2 | 2019 | USA | PRJNA288601 |
| GCA 022211745 | PDT000630652.1 | Klebsiella_45 | PDS000052944.18 | 2013 | Canada | PRJNA564992 |
| GCA 030019695 | PDT001737461.1 | Klebsiella_45 | PDS000052944.18 | 2013 | USA | PRJNA395086 |
| GCA 016903895 | PDT000967411.1 | Klebsiella_45 | | nan | USA | PRJNA231221 |
| GCA 022430345 | PDT001256202.1 | Klebsiella_45 | PDS000105716.1 | 2020 | USA | PRJNA325243 |
| GCA 022312935 | PDT000136333.2 | Klebsiella_45 | | 2011 | United Kingdom | PRJNA306133 |
| GCA 022285805 | PDT000431395.1 | Klebsiella_45 | | 2017 | USA | PRJNA396774 |
| GCA 043331735 | PDT002278350.1 | Klebsiella_45 | PDS000178660.3 | 2019 | USA | PRJNA1106484 |
| GCA 046015795 | PDT002530113.1 | Klebsiella_45 | PDS000178660.3 | 2018 | USA | PRJNA288601 |
| GCA 022033615 | PDT000901821.3 | Klebsiella_45 | PDS000178660.3 | 2018 | USA | PRJNA288601 |
| GCA 043569885 | PDT002347492.1 | Klebsiella_45 | PDS000100594.2 | 2009 | Norway | PRJEB27256 |
| GCA 900511045 | PDT000369071.1 | Klebsiella_45 | | 2013 | Czechia | PRJEB10018 |
| GCA 032046115 | PDT001903439.1 | Klebsiella_45 | | 2018 | Norway | PRJEB48268 |
| GCA 032042675 | PDT001903028.1 | Klebsiella_45 | | 2018 | Norway | PRJEB48268 |
| GCA 043709025 | PDT002376631.1 | Klebsiella_45 | | 2018 | Japan | PRJDB14656 |
| GCA 900509075 | PDT000370712.1 | Klebsiella_45 | | 2013 | Turkey | PRJEB10018 |
| GCA 916160865 | PDT001141212.1 | Klebsiella_45 | PDS000127598.3 | 2019 | Netherlands | PRJEB41042 |
| GCA 026888235 | PDT001525297.3 | Klebsiella_- | PDS000127598.3 | 2022 | USA | PRJNA288601 |
| GCA 022284885 | PDT000431439.1 | Klebsiella_45 | PDS000060037.37 | 2017 | USA | PRJNA396774 |
| GCA 022286675 | PDT000431353.1 | Klebsiella_45 | PDS000060037.37 | 2017 | USA | PRJNA396774 |
| GCA 022199725 | PDT000707850.1 | Klebsiella_- | PDS000060037.37 | nan | NA | PRJEB22903 |
| GCA 044380335 | PDT002305393.1 | Klebsiella_45 | PDS000060037.37 | nan | USA | PRJNA271899 |
| GCA 021922285 | PDT001092054.1 | Klebsiella_45 | | 2013 | Norway | PRJEB27256 |
| GCA 022141525 | PDT000870424.1 | Klebsiella_45 | | 2015 | Romania | PRJNA396838 |
| GCA 900515965 | PDT000369667.1 | Klebsiella_45 | | 2014 | Italy | PRJEB10018 |
| GCA 900085275 | PDT000131593.1 | Klebsiella_45 | | 2008 | United Kingdom | PRJEB5065 |
| GCA 019798025 | PDT001120287.1 | Klebsiella_2954 | | 2015 | Norway | PRJNA723772 |
| GCA 021918225 | PDT001092348.1 | Klebsiella_45 | PDS000093208.1 | 2009 | Norway | PRJEB27256 |
| GCA 018441205 | PDT001046596.1 | Klebsiella_45 | | 2019 | Germany | PRJNA692829 |
| GCA 002192315 | PDT000218722.1 | Klebsiella_45 | PDS000006905.10 | nan | NA | PRJNA292902 |
| GCA 026674905 | PDT001518814.1 | Klebsiella_45 | | 2022 | Ghana | PRJNA906096 |
| GCA 032076315 | PDT001904954.1 | Klebsiella_45 | | 2018 | Norway | PRJEB48268 |
| GCA 038593465 | PDT002148604.1 | Klebsiella_45 | PDS000213518.2 | 2020 | Malawi | PRJNA1102521 |
| GCA 964207685 | PDT002279439.1 | Klebsiella_45 | PDS000190165.1 | 2021 | Tanzania | PRJEB78367 |
| GCA 964207185 | PDT002279454.1 | Klebsiella_45 | PDS000190165.1 | 2021 | Tanzania | PRJEB78367 |
| GCA 026256265 | PDT001849267.1 | Klebsiella_45 | PDS000074716.5 | 2013 | Australia | PRJNA646837 |
| GCA 003432285 | PDT000371002.1 | Klebsiella_45 | PDS000074716.5 | 2013 | Australia | PRJNA486877 |
| GCA 029577515 | PDT001677826.1 | Klebsiella_45 | | 2019 | Italy | PRJNA949542 |
| GCA 043992935 | PDT002376563.1 | Klebsiella_45 | | 2016 | Japan | PRJDB14656 |
| GCA 022276145 | PDT000432070.1 | Klebsiella_45 | PDS000054827.4 | 2015 | Japan | PRJDB6565 |
| GCA 022207045 | PDT000651374.1 | Klebsiella_45 | PDS000054827.4 | 2015 | Japan | PRJDB6565 |
| GCA 022207445 | PDT000651336.1 | Klebsiella_45 | PDS000054827.4 | 2015 | Japan | PRJDB6565 |
| GCA 022207425 | PDT000651334.1 | Klebsiella_45 | PDS000054827.4 | 2015 | Japan | PRJDB6565 |
| GCA 022204645 | PDT000668791.1 | Klebsiella_45 | PDS000054827.4 | 2015 | Japan | PRJDB6565 |
| GCA 037052165 | PDT002102083.1 | Klebsiella_45 | | 2015 | Jordan | PRJNA1076682 |
| GCA 964303885 | PDT002500340.1 | Klebsiella_45 | | 2020 | Germany | PRJEB81699 |
| GCA 021865655 | PDT001207408.1 | Klebsiella_45 | PDS000101897.1 | 2019 | France | PRJEB34643 |
| GCA 021865675 | PDT001207409.1 | Klebsiella_45 | PDS000101897.1 | 2019 | France | PRJEB34643 |
| GCA 021866675 | PDT001207293.1 | Klebsiella_45 | PDS000101897.1 | 2019 | France | PRJEB34643 |
| GCA 021865695 | PDT001207404.1 | Klebsiella_45 | PDS000101897.1 | 2019 | France | PRJEB34643 |
| GCA 036476045 | PDT002067384.1 | Klebsiella_45 | PDS000060037.37 | 2023 | USA | PRJNA288601 |
| GCA 031212105 | PDT001876221.2 | Klebsiella_45 | PDS000060037.37 | 2023 | USA | PRJNA288601 |
| GCA 031211445 | PDT001876244.2 | Klebsiella_- | PDS000060037.37 | 2023 | USA | PRJNA288601 |
| GCA 045633225 | PDT002500126.1 | Klebsiella_- | PDS000060037.37 | 2024 | USA | PRJNA288601 |
| GCA 045720915 | PDT002512352.1 | Klebsiella_45 | PDS000060037.37 | 2024 | USA | PRJNA288601 |
| GCA 045430815 | PDT002498217.1 | Klebsiella_45 | PDS000060037.37 | 2023 | USA | PRJNA288601 |
| GCA 031209215 | PDT001876301.2 | Klebsiella_45 | PDS000060037.37 | 2023 | USA | PRJNA288601 |
| GCA 031214895 | PDT001876109.2 | Klebsiella_45 | PDS000060037.37 | 2023 | USA | PRJNA288601 |
| GCA 045731845 | PDT002512390.1 | Klebsiella_45 | PDS000060037.37 | 2024 | USA | PRJNA288601 |
| GCA 036474115 | PDT002067466.1 | Klebsiella_45 | PDS000060037.37 | 2023 | USA | PRJNA288601 |
| GCA 031208815 | PDT001876309.2 | Klebsiella_45 | PDS000060037.37 | 2023 | USA | PRJNA288601 |
| GCA 026057255 | PDT001480835.3 | Klebsiella_- | PDS000060037.37 | 2022 | USA | PRJNA288601 |
| GCA 024905435 | PDT001395499.3 | Klebsiella_- | PDS000060037.37 | 2022 | USA | PRJNA288601 |
| GCA 031208865 | PDT001876308.2 | Klebsiella_- | PDS000060037.37 | 2023 | USA | PRJNA288601 |
| GCA 031211925 | PDT001876219.2 | Klebsiella_45 | PDS000060037.37 | 2023 | USA | PRJNA288601 |
| GCA 031212885 | PDT001876173.2 | Klebsiella_45 | PDS000060037.37 | 2023 | USA | PRJNA288601 |
| GCA 036693205 | PDT002080954.1 | Klebsiella_45 | PDS000060037.37 | 2023 | USA | PRJNA288601 |

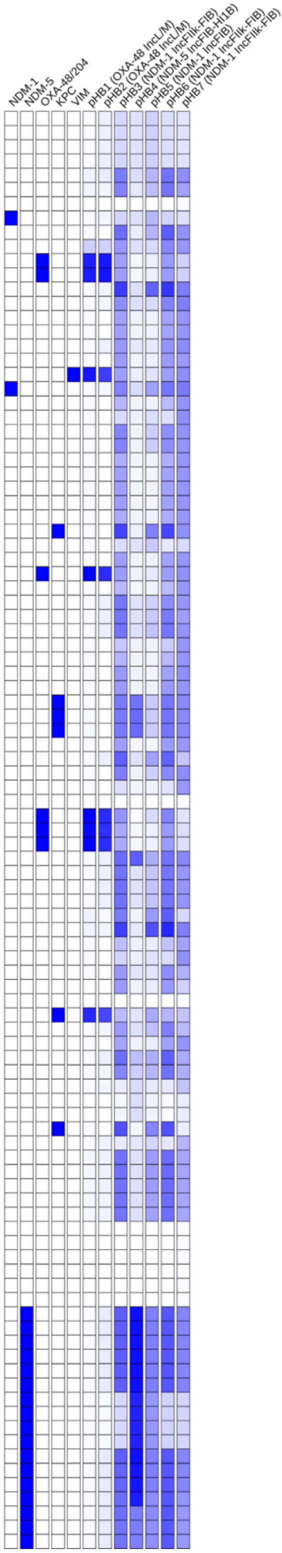

### Tree 37: GCA_043905585

Tree scale: 0.01

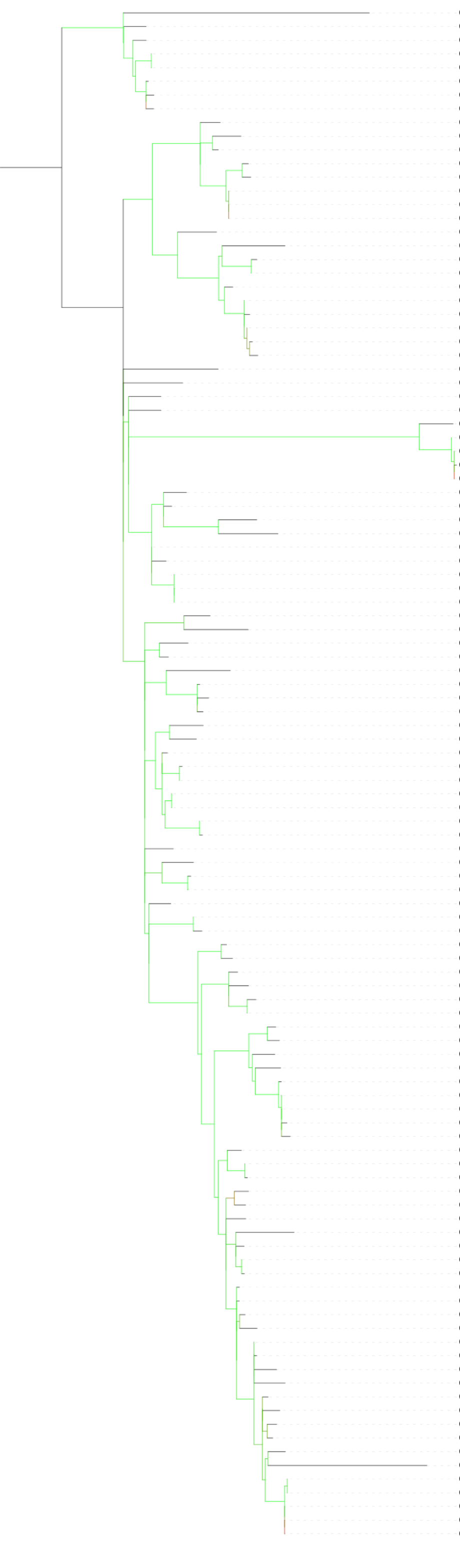

| | Isolate | MLST | SNP Cluster | Collection Year | Country | BioProject |
|---|---|---|---|---|---|---|
| GCA 043358855 | PDT002423174.1 | Klebsiella_101 | PDS000166495.5 | nan | Saudi Arabia | PRJNA1116866 |
| GCA 043358275 | PDT002423146.1 | Klebsiella_101 | PDS000166495.5 | nan | Saudi Arabia | PRJNA1116866 |
| GCA 022187905 | PDT000715723.1 | Klebsiella_101 | PDS000166495.5 | 2018 | Saudi Arabia | PRJEB36683 |
| GCA 040325825 | PDT002230876.1 | Klebsiella_- | PDS000166495.5 | 2018 | Saudi Arabia | PRJNA1107218 |
| GCA 040325855 | PDT002230877.1 | Klebsiella_- | PDS000166495.5 | 2018 | Saudi Arabia | PRJNA1107218 |
| GCA 033548075 | PDT001982219.1 | Klebsiella_- | PDS000166495.5 | 2022 | Saudi Arabia | PRJNA1018815 |
| GCA 033799885 | PDT001991616.1 | Klebsiella_- | PDS000166495.5 | 2021 | Saudi Arabia | PRJNA1018815 |
| GCA 033800185 | PDT001991631.1 | Klebsiella_- | PDS000166495.5 | 2021 | Saudi Arabia | PRJNA1018815 |
| GCA 022185685 | PDT000716605.1 | Klebsiella_101 | PDS000060649.2 | 2018 | France | PRJEB35890 |
| GCA 043863365 | **HBH_3230_17** | Klebsiella_101 | PDS000060649.2 | 2017 | Tunisia | PRJNA1160997 |
| GCA 022185745 | PDT000716602.1 | Klebsiella_101 | PDS000060649.2 | 2018 | France | PRJEB35890 |
| GCA 043905465 | **HBH_859_21** | Klebsiella_101 | PDS000060649.2 | 2021 | Tunisia | PRJNA1160997 |
| GCA 043905585 | **HBH_534_22** | Klebsiella_101 | PDS000060649.2 | 2022 | Tunisia | PRJNA1160997 |
| GCA 043863515 | **HBH_2052_18** | Klebsiella_101 | PDS000060649.2 | 2018 | Tunisia | PRJNA1160997 |
| GCA 043904785 | **HBH_2493_18** | Klebsiella_101 | PDS000060649.2 | 2018 | Tunisia | PRJNA1160997 |
| GCA 043863555 | **HBH_2343_18** | Klebsiella_101 | PDS000060649.2 | 2018 | Tunisia | PRJNA1160997 |
| GCA 956479465 | PDT001863965.2 | Klebsiella_101 | PDS000054005.14 | 2017 | Romania | PRJEB63349 |
| GCA 020576795 | PDT001161735.1 | Klebsiella_101 | PDS000054005.14 | 2018 | Spain | PRJNA718833 |
| GCA 009710345 | PDT000637205.1 | Klebsiella_101 | PDS000054005.14 | 2018/2019 | Romania | PRJNA579879 |
| GCA 009710085 | PDT000637192.1 | Klebsiella_101 | PDS000054005.14 | 2018/2019 | Romania | PRJNA579879 |
| GCA 028560855 | PDT001610689.1 | Klebsiella_101 | PDS000054005.14 | 2020 | Spain | PRJEB57245 |
| GCA 009710385 | PDT000637207.1 | Klebsiella_101 | PDS000054005.14 | 2018/2019 | Romania | PRJNA579879 |
| GCA 009710105 | PDT000637193.1 | Klebsiella_101 | PDS000054005.14 | 2018/2019 | Romania | PRJNA579879 |
| GCA 009710765 | PDT000637224.1 | Klebsiella_101 | PDS000054005.14 | 2018/2019 | Romania | PRJNA579879 |
| GCA 009646285 | PDT000626675.1 | Klebsiella_101 | PDS000054005.14 | 2018/2019 | Romania | PRJNA579879 |
| GCA 009710005 | PDT000637189.1 | Klebsiella_101 | PDS000054005.14 | 2018/2019 | Romania | PRJNA579879 |
| GCA 022122385 | PDT001244283.1 | Klebsiella_101 | | 2014 | United Kingdom | PRJNA788733 |
| GCA 043859795 | **HBH_3284_13** | Klebsiella_101 | | 2013 | Tunisia | PRJNA1160997 |
| GCA 022290985 | PDT000427324.1 | Klebsiella_2502 | PDS000045324.14 | nan | NA | PRJEB10018 |
| GCA 021819665 | PDT001219141.1 | Klebsiella_101 | PDS000045324.14 | 2018 | Germany | PRJEB43549 |
| GCA 018444275 | PDT001046631.1 | Klebsiella_101 | | 2018 | Germany | PRJNA692829 |
| GCA 043860495 | **HBH_170_15** | Klebsiella_101 | PDS000199426.1 | 2015 | Tunisia | PRJNA1160997 |
| GCA 043863105 | **HBH_2082_15** | Klebsiella_101 | PDS000199426.1 | 2015 | Tunisia | PRJNA1160997 |
| GCA 043860375 | **HBH_4306_14** | Klebsiella_101 | PDS000199426.1 | 2014 | Tunisia | PRJNA1160997 |
| GCA 043860455 | **HBH_84_15** | Klebsiella_101 | PDS000199426.1 | 2015 | Tunisia | PRJNA1160997 |
| GCA 900508885 | PDT000370690.1 | Klebsiella_101 | PDS000045324.14 | 2013 | Turkey | PRJEB10018 |
| GCA 900509165 | PDT000370720.1 | Klebsiella_101 | PDS000045324.14 | 2013 | Turkey | PRJEB10018 |
| GCA 023023525 | PDT001289697.1 | Klebsiella_- | PDS000045324.14 | 2018 | Turkey | PRJNA789336 |
| GCA 900508875 | PDT000370684.1 | Klebsiella_101 | PDS000045324.14 | 2014 | Turkey | PRJEB10018 |
| GCA 043858225 | **HBH_835_11** | Klebsiella_101 | PDS000045324.14 | 2011 | Tunisia | PRJNA1160997 |
| GCA 022239805 | PDT000441634.1 | Klebsiella_101 | PDS000045324.14 | 2012 | Germany | PRJEB18621 |
| GCA 022211485 | PDT000631361.1 | Klebsiella_101 | PDS000045324.14 | 2014 | Canada | PRJNA564992 |
| GCA 043976095 | PDT002412006.1 | Klebsiella_- | PDS000045324.14 | 2014 | Canada | PRJNA855907 |
| GCA 022230705 | PDT000445958.1 | Klebsiella_101 | PDS000045324.14 | 2014 | Canada | PRJNA390933 |
| GCA 900509195 | PDT000370721.1 | Klebsiella_101 | PDS000045328.1 | 2013 | Turkey | PRJEB10018 |
| GCA 900509145 | PDT000370717.1 | Klebsiella_101 | | 2014 | Turkey | PRJEB10018 |
| GCA 900517835 | PDT000369898.1 | Klebsiella_101 | PDS000045324.14 | 2013 | Greece | PRJEB10018 |
| GCA 022290735 | PDT000427340.1 | Klebsiella_- | PDS000045324.14 | nan | NA | PRJEB10018 |
| GCA 019433875 | PDT001101052.1 | Klebsiella_101 | | 2018 | Iran | PRJNA749021 |
| GCA 022426945 | PDT001256971.1 | Klebsiella_101 | PDS000080193.2 | 2015 | Turkey | PRJNA809139 |
| GCA 022426965 | PDT001256972.1 | Klebsiella_101 | PDS000080193.2 | 2015 | Turkey | PRJNA809150 |
| GCA 022426985 | PDT001256973.1 | Klebsiella_101 | PDS000080193.2 | 2015 | Turkey | PRJNA809151 |
| GCA 900507645 | PDT000370574.1 | Klebsiella_101 | PDS000045324.14 | 2014 | Turkey | PRJEB10018 |
| GCA 023023305 | PDT001289727.1 | Klebsiella_101 | PDS000045324.14 | 2019 | Turkey | PRJNA789336 |
| GCA 020120115 | PDT001137084.1 | Klebsiella_101 | PDS000045324.14 | 2013 | Switzerland | PRJNA543274 |
| GCA 022287345 | PDT000427831.1 | Klebsiella_- | PDS000045324.14 | nan | NA | PRJEB10018 |
| GCA 022287225 | PDT000427835.1 | Klebsiella_- | PDS000045324.14 | nan | NA | PRJEB10018 |
| GCA 900508185 | PDT000370728.1 | Klebsiella_101 | PDS000045324.14 | 2013 | Turkey | PRJEB10018 |
| GCA 022287305 | PDT000427833.1 | Klebsiella_101 | PDS000045324.14 | nan | NA | PRJEB10018 |
| GCA 022287265 | PDT000427836.1 | Klebsiella_- | PDS000045324.14 | nan | NA | PRJEB10018 |
| GCA 022287245 | PDT000427837.1 | Klebsiella_101 | PDS000045324.14 | nan | NA | PRJEB10018 |
| GCA 900508585 | PDT000370639.1 | Klebsiella_101 | PDS000045324.14 | 2014 | Turkey | PRJEB10018 |
| GCA 900508775 | PDT000370667.1 | Klebsiella_101 | PDS000045324.14 | 2014 | Turkey | PRJEB10018 |
| GCA 040270895 | PDT002227167.1 | Klebsiella_101 | PDS000045324.14 | 2015 | United Kingdom | PRJNA1123654 |
| GCA 022100675 | PDT001243870.1 | Klebsiella_101 | PDS000045324.14 | 2014 | United Kingdom | PRJNA788733 |
| GCA 022237435 | PDT000441697.1 | Klebsiella_101 | PDS000045324.14 | 2013 | Germany | PRJEB18621 |
| GCA 035017105 | PDT002045095.1 | Klebsiella_101 | PDS000045324.14 | 2022 | Libya | PRJNA998204 |
| GCA 035017425 | PDT002045110.1 | Klebsiella_101 | PDS000045324.14 | 2022 | Libya | PRJNA998204 |
| GCA 020116295 | PDT001137019.1 | Klebsiella_101 | PDS000104479.25 | 2013 | Switzerland | PRJNA543274 |
| GCA 020120475 | PDT001137102.1 | Klebsiella_101 | PDS000104479.25 | 2013 | Switzerland | PRJNA543274 |
| GCA 900505505 | PDT000370447.1 | Klebsiella_101 | PDS000104479.25 | 2014 | Serbia | PRJEB10018 |
| GCA 900505525 | PDT000370449.1 | Klebsiella_101 | PDS000104479.25 | 2014 | Serbia | PRJEB10018 |
| GCA 900505535 | PDT000370450.1 | Klebsiella_101 | PDS000104479.25 | 2014 | Serbia | PRJEB10018 |
| GCA 900505485 | PDT000370448.1 | Klebsiella_101 | PDS000104479.25 | 2014 | Serbia | PRJEB10018 |
| GCA 956476255 | PDT001863970.2 | Klebsiella_101 | PDS000104479.25 | 2016 | Serbia | PRJEB63349 |
| GCA 022234385 | PDT000442763.1 | Klebsiella_- | PDS000104479.25 | 2016 | Serbia | PRJNA449293 |
| GCA 022234645 | PDT000442748.1 | Klebsiella_101 | PDS000104479.25 | 2015 | Serbia | PRJNA449293 |
| GCA 033865285 | PDT001992447.1 | Klebsiella_101 | PDS000104479.25 | 2016 | Canada | PRJNA532924 |
| GCA 022235005 | PDT000442736.1 | Klebsiella_101 | PDS000104479.25 | 2016 | Serbia | PRJNA449293 |
| GCA 022234805 | PDT000442744.1 | Klebsiella_101 | PDS000104479.25 | 2015 | Serbia | PRJNA449293 |
| GCA 022234395 | PDT000442759.1 | Klebsiella_101 | PDS000104479.25 | 2015 | Serbia | PRJNA449293 |
| GCA 022234465 | PDT000442753.1 | Klebsiella_101 | PDS000104479.25 | 2016 | Serbia | PRJNA449293 |
| GCA 022234325 | PDT000442762.1 | Klebsiella_- | PDS000104479.25 | 2015 | Serbia | PRJNA449293 |
| GCA 900505315 | PDT000370421.1 | Klebsiella_101 | PDS000104479.25 | 2014 | Serbia | PRJEB10018 |
| GCA 900505595 | PDT000370458.1 | Klebsiella_101 | PDS000104479.25 | 2014 | Serbia | PRJEB10018 |
| GCA 900505555 | PDT000370456.1 | Klebsiella_101 | PDS000104479.25 | 2014 | Serbia | PRJEB10018 |
| GCA 900503565 | PDT000370439.1 | Klebsiella_101 | PDS000104479.25 | 2013 | Serbia | PRJEB10018 |
| GCA 900505275 | PDT000370412.1 | Klebsiella_101 | PDS000104479.25 | 2013 | Serbia | PRJEB10018 |
| GCA 900505255 | PDT000370411.1 | Klebsiella_101 | PDS000104479.25 | 2014 | Serbia | PRJEB10018 |
| GCA 900505445 | PDT000370441.1 | Klebsiella_101 | PDS000104479.25 | 2014 | Serbia | PRJEB10018 |
| GCA 900503505 | PDT000370423.1 | Klebsiella_101 | PDS000104479.25 | 2014 | Serbia | PRJEB10018 |
| GCA 900505145 | PDT000370398.1 | Klebsiella_101 | PDS000104479.25 | 2013 | Serbia | PRJEB10018 |
| GCA 900505195 | PDT000370399.1 | Klebsiella_101 | PDS000104479.25 | 2013 | Serbia | PRJEB10018 |
| GCA 900505065 | PDT000370390.1 | Klebsiella_101 | PDS000104479.25 | 2014 | Serbia | PRJEB10018 |
| GCA 902155925 | PDT000663180.1 | Klebsiella_101 | PDS000104479.25 | 2014 | Serbia | PRJEB33308 |
| GCA 020115935 | PDT001137010.1 | Klebsiella_101 | PDS000104479.25 | 2014 | Switzerland | PRJNA543274 |
| GCA 006459145 | PDT000532615.1 | Klebsiella_101 | PDS000104479.25 | nan | NA | PRJNA549322 |
| GCA 900507115 | PDT000370462.1 | Klebsiella_101 | PDS000104479.25 | 2014 | Serbia | PRJEB10018 |
| GCA 900505135 | PDT000370392.1 | Klebsiella_101 | PDS000104479.25 | 2014 | Serbia | PRJEB10018 |
| GCA 900505095 | PDT000370391.1 | Klebsiella_101 | PDS000104479.25 | 2014 | Serbia | PRJEB10018 |
| GCA 900505435 | PDT000370437.1 | Klebsiella_101 | PDS000104479.25 | 2014 | Serbia | PRJEB10018 |
| GCA 900505125 | PDT000370397.1 | Klebsiella_101 | PDS000104479.25 | 2014 | Serbia | PRJEB10018 |
| GCA 900507195 | PDT000370466.1 | Klebsiella_101 | PDS000104479.25 | 2014 | Serbia | PRJEB10018 |
| GCA 900502985 | PDT000370139.1 | Klebsiella_101 | PDS000104479.25 | 2013 | Slovenia | PRJEB10018 |
| GCA 900503095 | PDT000370151.1 | Klebsiella_101 | PDS000104479.25 | 2013 | Slovenia | PRJEB10018 |
| GCA 022234105 | PDT000442775.1 | Klebsiella_101 | PDS000104479.25 | 2017 | Serbia | PRJNA449293 |
| GCA 956478885 | PDT001863982.2 | Klebsiella_101 | PDS000104479.25 | 2017 | Serbia | PRJEB63349 |
| GCA 022759985 | PDT001277612.1 | Klebsiella_101 | PDS000104479.25 | 2013 | Switzerland | PRJNA769291 |
| GCA 022759945 | PDT001277610.1 | Klebsiella_101 | PDS000104479.25 | 2013 | Switzerland | PRJNA769291 |
| GCA 022759925 | PDT001277609.1 | Klebsiella_101 | PDS000104479.25 | 2013 | Switzerland | PRJNA769291 |
| GCA 022759965 | PDT001277611.1 | Klebsiella_101 | PDS000104479.25 | 2013 | Switzerland | PRJNA769291 |
| GCA 022760005 | PDT001277613.1 | Klebsiella_101 | PDS000104479.25 | 2013 | Switzerland | PRJNA769291 |

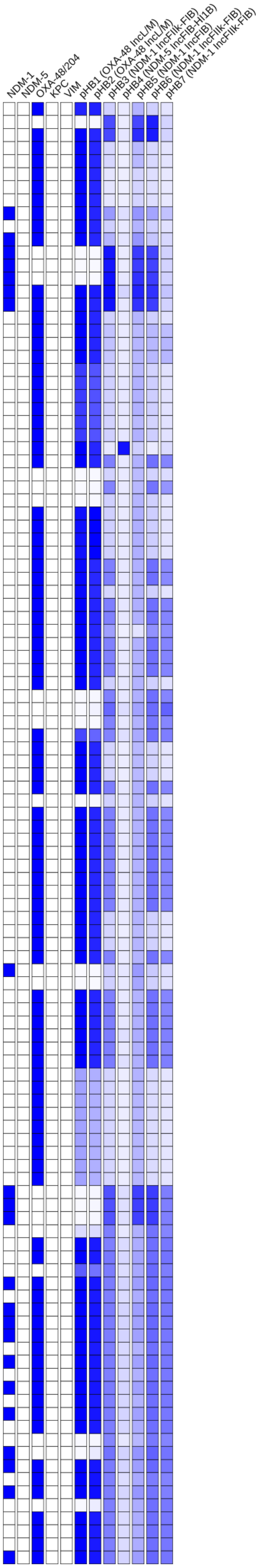

### Tree 38: GCA_043905605

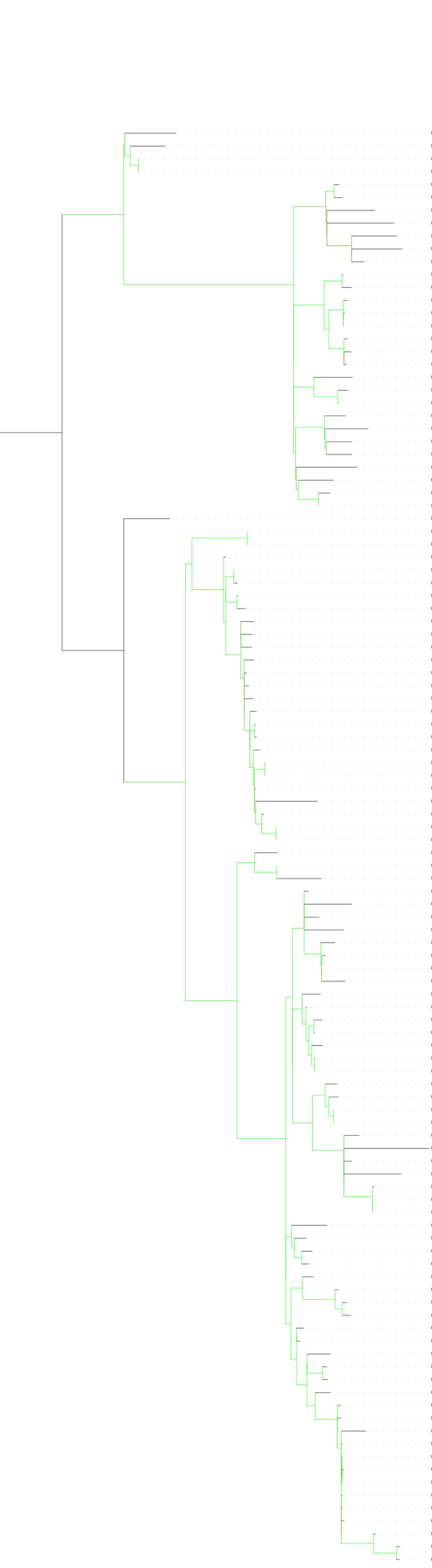


| | Isolate | MLST | SNP Cluster | Collection Year | Country | BioProject |
|---|---|---|---|---|---|---|
| GCA 022104535 | PDT001243313.1 | Klebsiella_383 | PDS000106324.3 | 2016 | United Kingdom | PRJNA788733 |
| GCA 040201255 | PDT002208472.1 | Klebsiella_- | | nan | Saudi Arabia | PRJNA1116866 |
| GCA 022550215 | PDT001263624.1 | Klebsiella_383 | PDS000106324.3 | 2019 | Egypt | PRJNA720263 |
| GCA 022550175 | PDT001263622.1 | Klebsiella_383 | PDS000106324.3 | 2018 | Egypt | PRJNA720263 |
| GCA 024087515 | PDT001350138.1 | Klebsiella_383 | PDS000108392.5 | 2016 | Qatar | PRJNA656934 |
| GCA 025144525 | PDT001416171.1 | Klebsiella_383 | PDS000108392.5 | 2016 | Qatar | PRJNA656934 |
| GCA 022550105 | PDT001263620.1 | Klebsiella_383 | | 2019 | Egypt | PRJNA720263 |
| GCA 037000505 | PDT002098839.1 | Klebsiella_383 | | 2015 | Egypt | PRJNA1079765 |
| GCA 016651765 | PDT000937804.1 | Klebsiella_383 | PDS000106324.3 | nan | Egypt | PRJNA689413 |
| GCA 016651915 | PDT000937812.1 | Klebsiella_383 | | nan | Egypt | PRJNA689413 |
| GCA 023080795 | PDT001289734.1 | Klebsiella_383 | PDS000108392.5 | 2018 | Turkey | PRJNA789336 |
| GCA 012271775 | PDT000721361.1 | Klebsiella_383 | PDS000061038.3 | 2018 | Lebanon | PRJNA613441 |
| GCA 012271755 | PDT000721360.1 | Klebsiella_383 | PDS000061038.3 | 2018 | Lebanon | PRJNA613441 |
| GCA 021956345 | PDT001020277.1 | Klebsiella_383 | PDS000061038.3 | 2018 | Lebanon | PRJNA658369 |
| GCA 021965695 | PDT001019815.1 | Klebsiella_383 | PDS000061038.3 | 2018 | Lebanon | PRJNA658369 |
| GCA 021956085 | PDT001020290.1 | Klebsiella_383 | PDS000061038.3 | 2018 | Lebanon | PRJNA658369 |
| GCA 021955425 | PDT001020322.1 | Klebsiella_383 | PDS000061038.3 | 2018 | Lebanon | PRJNA658369 |
| GCA 021962155 | PDT001020003.1 | Klebsiella_383 | PDS000061038.3 | 2018 | Lebanon | PRJNA658369 |
| GCA 021968095 | PDT001019675.1 | Klebsiella_383 | PDS000061038.3 | 2018 | Lebanon | PRJNA658369 |
| GCA 043019365 | PDT002412609.1 | Klebsiella_383 | | 2024 | USA | PRJNA288601 |
| GCA 037493415 | PDT002120373.1 | Klebsiella_383 | PDS000179530.2 | 2024 | USA | PRJNA288601 |
| GCA 031043935 | PDT001729747.3 | Klebsiella_383 | PDS000179530.2 | 2023 | USA | PRJNA288601 |
| GCA 018441475 | PDT001046601.1 | Klebsiella_383 | | 2018 | Germany | PRJNA692829 |
| GCA 044467905 | PDT002451619.1 | Klebsiella_383 | PDS000125001.14 | 2022 | USA | PRJNA1106484 |
| GCA 022017035 | PDT000948383.1 | Klebsiella_383 | | 2019 | Italy | PRJEB37610 |
| GCA 032721395 | PDT001927031.1 | Klebsiella_383 | | 2019 | Italy | PRJNA1026096 |
| GCA 021938955 | PDT001071296.1 | Klebsiella_383 | PDS000106730.2 | 2019 | Germany | PRJEB42370 |
| GCA 025037975 | PDT001386067.1 | Klebsiella_383 | | 2019 | Canada | PRJNA855907 |
| GCA 019781185 | PDT001118308.1 | Klebsiella_- | PDS000202475.1 | 2019 | Germany | PRJNA634442 |
| GCA 043832345 | PDT002430492.1 | Klebsiella_- | PDS000202475.1 | 2019 | Germany | PRJNA1070536 |
| GCA 022104735 | PDT001244769.1 | Klebsiella_383 | | 2016 | United Kingdom | PRJNA788733 |
| GCA 028092175 | PDT001585477.1 | Klebsiella_383 | PDS000133910.8 | 2007 | Greece | PRJEB58216 |
| GCA 947662285 | PDT001590394.1 | Klebsiella_383 | PDS000133910.8 | 2007 | Greece | PRJEB58216 |
| GCA 017310305 | PDT000984751.1 | Klebsiella_383 | PDS000070357.5 | 2013 | China | PRJNA648010 |
| GCA 046466035 | PDT002556526.1 | Klebsiella_383 | PDS000070357.5 | nan | NA | PRJNA786399 |
| GCA 043109335 | PDT002420644.1 | Klebsiella_383 | PDS000070357.5 | 2014 | China | PRJNA1170063 |
| GCA 046466735 | PDT002556561.1 | Klebsiella_383 | PDS000070357.5 | nan | NA | PRJNA786399 |
| GCA 046466655 | PDT002556557.1 | Klebsiella_383 | PDS000070357.5 | nan | NA | PRJNA786399 |
| GCA 017311675 | PDT000984764.1 | Klebsiella_383 | PDS000070357.5 | 2017 | China | PRJNA648010 |
| GCA 017311645 | PDT000984763.1 | Klebsiella_383 | PDS000070357.5 | 2014 | China | PRJNA648010 |
| GCA 017311735 | PDT000984767.1 | Klebsiella_383 | PDS000070357.5 | 2013 | China | PRJNA648010 |
| GCA 014285595 | PDT000822624.1 | Klebsiella_383 | PDS000070357.5 | 2015 | China | PRJNA657553 |
| GCA 017311605 | PDT000984761.1 | Klebsiella_383 | PDS000070357.5 | 2013 | China | PRJNA648010 |
| GCA 014289355 | PDT000822768.1 | Klebsiella_383 | PDS000070357.5 | 2013 | China | PRJNA657553 |
| GCA 017311635 | PDT000984762.1 | Klebsiella_383 | PDS000070357.5 | 2014 | China | PRJNA648010 |
| GCA 014285645 | PDT000822627.1 | Klebsiella_383 | PDS000070357.5 | 2015 | China | PRJNA657553 |
| GCA 014285735 | PDT000822631.1 | Klebsiella_383 | PDS000070357.5 | 2014 | China | PRJNA657553 |
| GCA 046465235 | PDT002556486.1 | Klebsiella_383 | PDS000070357.5 | nan | NA | PRJNA786399 |
| GCA 014286025 | PDT000822646.1 | Klebsiella_383 | PDS000070357.5 | 2013 | China | PRJNA657553 |
| GCA 014285835 | PDT000822636.1 | Klebsiella_383 | PDS000070357.5 | 2014 | China | PRJNA657553 |
| GCA 046465045 | PDT002556479.1 | Klebsiella_383 | PDS000070357.5 | nan | NA | PRJNA786399 |
| GCA 014285675 | PDT000822629.1 | Klebsiella_383 | PDS000070357.5 | 2013 | China | PRJNA657553 |
| GCA 017311685 | PDT000984765.1 | Klebsiella_383 | PDS000070357.5 | 2017 | China | PRJNA648010 |
| GCA 014284855 | PDT000822587.1 | Klebsiella_383 | PDS000070357.5 | 2015 | China | PRJNA657553 |
| GCA 014285615 | PDT000822625.1 | Klebsiella_383 | PDS000070357.5 | 2015 | China | PRJNA657553 |
| GCA 014285545 | PDT000822621.1 | Klebsiella_383 | PDS000070357.5 | 2015 | China | PRJNA657553 |
| GCA 004803085 | PDT000495882.1 | Klebsiella_383 | | nan | Egypt | PRJNA508509 |
| GCA 022186025 | PDT000716574.1 | Klebsiella_383 | PDS000060640.1 | 2019 | Germany | PRJEB35890 |
| GCA 022186005 | PDT000716575.1 | Klebsiella_383 | PDS000060640.1 | 2019 | Germany | PRJEB35890 |
| GCA 043904905 | **HBH_487_19** | Klebsiella_383 | PDS000085592.11 | 2019 | Tunisia | PRJNA1160997 |
| GCA 043905405 | **HBH_3269_21** | Klebsiella_383 | | 2021 | Tunisia | PRJNA1160997 |
| GCA 043904845 | **HBH_1312_19** | Klebsiella_383 | PDS000085592.11 | 2019 | Tunisia | PRJNA1160997 |
| GCA 043905605 | **HBH_1879_22** | Klebsiella_383 | | 2022 | Tunisia | PRJNA1160997 |
| GCA 043905185 | **HBH_1789_20** | Klebsiella_383 | PDS000098809.4 | 2020 | Tunisia | PRJNA1160997 |
| GCA 021898295 | PDT001172748.1 | Klebsiella_383 | PDS000098809.4 | 2020 | USA | PRJNA288601 |
| GCA 021898135 | PDT001174156.3 | Klebsiella_383 | PDS000098809.4 | 2020 | USA | PRJNA288601 |
| GCA 035082745 | PDT002045985.1 | Klebsiella_383 | | 2023 | USA | PRJNA288601 |
| GCA 027908495 | PDT001551547.1 | Klebsiella_383 | PDS000085592.11 | 2020 | Egypt | PRJNA825651 |
| GCA 043905145 | **HBH_3731_19** | Klebsiella_383 | PDS000085592.11 | 2019 | Tunisia | PRJNA1160997 |
| GCA 043905385 | **HBH_211_21** | Klebsiella_383 | PDS000085592.11 | 2021 | Tunisia | PRJNA1160997 |
| GCA 043905505 | **HBH_990_22** | Klebsiella_383 | PDS000085592.11 | 2022 | Tunisia | PRJNA1160997 |
| GCA 043905345 | **HBH_1362_21** | Klebsiella_383 | PDS000085592.11 | 2021 | Tunisia | PRJNA1160997 |
| GCA 043905325 | **HBH_3788_20** | Klebsiella_383 | PDS000085592.11 | 2020 | Tunisia | PRJNA1160997 |
| GCA 043905355 | **HBH_3959_20** | Klebsiella_383 | PDS000085592.11 | 2020 | Tunisia | PRJNA1160997 |
| GCA 032720515 | PDT001927068.1 | Klebsiella_383 | PDS000085592.11 | 2020 | Italy | PRJNA1026096 |
| GCA 947572755 | PDT001769582.1 | Klebsiella_383 | PDS000085592.11 | 2022 | Germany | PRJEB58018 |
| GCA 025889055 | PDT001469194.1 | Klebsiella_383 | PDS000085592.11 | 2019 | Italy | PRJNA787449 |
| GCA 047426685 | PDT002605982.1 | Klebsiella_383 | PDS000085592.11 | 2019 | Italy | PRJNA1026096 |
| GCA 021971055 | PDT001019316.3 | Klebsiella_383 | PDS000085592.11 | 2021 | USA | PRJNA288601 |
| GCA 046601475 | PDT002562417.1 | Klebsiella_383 | | 2021 | Singapore | PRJNA1206822 |
| GCA 922831335 | PDT001211077.1 | Klebsiella_383 | PDS000085592.11 | 2019 | United Kingdom | PRJEB48990 |
| GCA 036455385 | PDT002074708.1 | Klebsiella_383 | | 2021 | Germany | PRJNA1060526 |
| GCA 037009425 | PDT002097979.1 | Klebsiella_6118 | PDS000175841.1 | 2024 | USA | PRJNA288601 |
| GCA 037009385 | PDT002097981.1 | Klebsiella_6118 | PDS000175841.1 | 2024 | USA | PRJNA288601 |
| GCA 037009405 | PDT002097980.1 | Klebsiella_6118 | PDS000175841.1 | 2024 | USA | PRJNA288601 |
| GCA 042146905 | PDT002390135.1 | Klebsiella_383 | | 2024 | USA | PRJNA288601 |
| GCA 043000025 | PDT002129380.1 | Klebsiella_383 | | 2021 | Egypt | PRJNA1071125 |
| GCA 042999945 | PDT002129384.1 | Klebsiella_383 | PDS000180184.1 | 2021 | Egypt | PRJNA1071125 |
| GCA 031047955 | PDT001725765.1 | Klebsiella_383 | PDS000180184.1 | 2020 | Egypt | PRJNA917066 |
| GCA 032721505 | PDT001927025.1 | Klebsiella_383 | PDS000217883.1 | 2019 | Italy | PRJNA1026096 |
| GCA 047501205 | PDT002615048.1 | Klebsiella_383 | PDS000217883.1 | 2021 | Tunisia | PRJNA1151248 |
| GCA 047501255 | PDT002615051.1 | Klebsiella_383 | PDS000217883.1 | 2023 | Tunisia | PRJNA1151248 |
| GCA 047501245 | PDT002615049.1 | Klebsiella_383 | PDS000217883.1 | 2023 | Tunisia | PRJNA1151248 |
| GCA 032721475 | PDT001927026.1 | Klebsiella_383 | PDS000085592.11 | 2019 | Italy | PRJNA1026096 |
| GCA 025892815 | PDT001469195.1 | Klebsiella_383 | PDS000085592.11 | 2019 | Italy | PRJNA787449 |
| GCA 035017125 | PDT002045096.1 | Klebsiella_383 | | 2022 | Libya | PRJNA998204 |
| GCA 035017085 | PDT002045094.1 | Klebsiella_383 | PDS000171211.1 | 2022 | Libya | PRJNA998204 |
| GCA 035016965 | PDT002045089.1 | Klebsiella_383 | PDS000171211.1 | 2022 | Libya | PRJNA998204 |
| GCA 039516715 | PDT002152585.1 | Klebsiella_383 | | 2024 | USA | PRJNA288601 |
| GCA 964277015 | PDT002423787.1 | Klebsiella_383 | PDS000192050.8 | 2024 | Slovakia | PRJEB72521 |
| GCA 964277065 | PDT002423785.1 | Klebsiella_383 | PDS000192050.8 | 2024 | Slovakia | PRJEB72521 |
| GCA 964339425 | PDT002468759.1 | Klebsiella_383 | PDS000192050.8 | 2024 | Slovakia | PRJEB72521 |
| GCA 964338835 | PDT002461347.1 | Klebsiella_383 | PDS000192050.8 | 2024 | Slovakia | PRJEB72521 |
| GCA 964338735 | PDT002462501.1 | Klebsiella_383 | PDS000192050.8 | 2024 | Slovakia | PRJEB72521 |
| GCA 964339365 | PDT002468763.1 | Klebsiella_383 | PDS000192050.8 | 2024 | Slovakia | PRJEB72521 |
| GCA 964339405 | PDT002468761.1 | Klebsiella_383 | PDS000192050.8 | 2024 | Slovakia | PRJEB72521 |
| GCA 964417265 | PDT002518810.1 | Klebsiella_383 | PDS000192050.8 | 2024 | Slovakia | PRJEB72521 |
| GCA 964339305 | PDT002468760.1 | Klebsiella_383 | PDS000192050.8 | 2024 | Slovakia | PRJEB72521 |
| GCA 964276885 | PDT002423793.1 | Klebsiella_383 | PDS000192050.8 | 2024 | Slovakia | PRJEB72521 |
| GCA 964339465 | PDT002468764.1 | Klebsiella_383 | PDS000192050.8 | 2024 | Slovakia | PRJEB72521 |
| GCA 964417105 | PDT002518807.1 | Klebsiella_383 | PDS000192050.8 | 2024 | Slovakia | PRJEB72521 |
| GCA 964417235 | PDT002518801.1 | Klebsiella_383 | PDS000192050.8 | 2024 | Slovakia | PRJEB72521 |

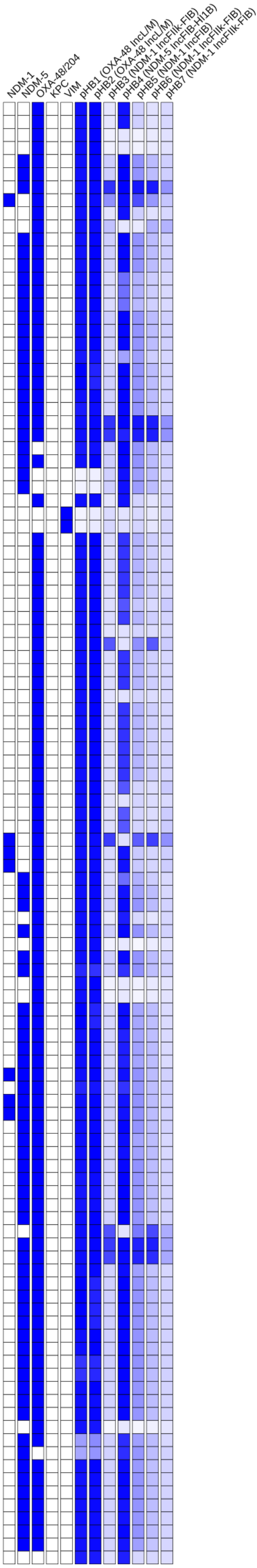

### Tree 39: GCA_043905625

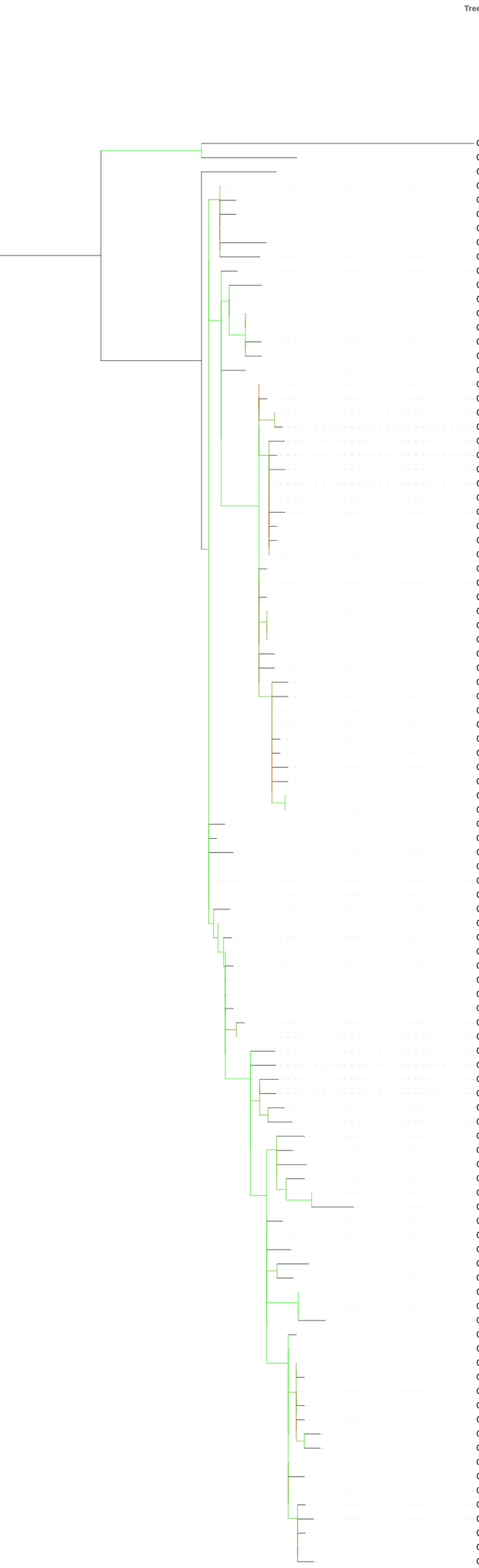


| | Isolate | MLST | SNP Cluster | Collection Year | Country | BioProject |
|---|---|---|---|---|---|---|
| GCA 045571255 | PDT002505357.1 | Klebsiella_147 | PDS000210545.1 | 2024 | USA | PRJNA288601 |
| GCA 043905625 | **HBH_20_22** | Klebsiella_147 | PDS000199417.1 | 2022 | Tunisia | PRJNA1160997 |
| GCA 031014325 | PDT001826060.2 | Klebsiella_147 | PDS000091501.176 | 2023 | USA | PRJNA288601 |
| GCA 021938175 | PDT001072674.3 | Klebsiella_147 | PDS000091501.176 | 2021 | USA | PRJNA288601 |
| GCA 027830015 | PDT001550944.3 | Klebsiella_147 | PDS000091501.176 | 2022 | USA | PRJNA288601 |
| GCA 021907655 | PDT001105013.3 | Klebsiella_147 | PDS000091501.176 | 2021 | USA | PRJNA288601 |
| GCA 021938345 | PDT001072669.3 | Klebsiella_147 | PDS000091501.176 | 2021 | USA | PRJNA288601 |
| GCA 023108235 | PDT001293728.3 | Klebsiella_147 | PDS000091501.176 | 2021 | USA | PRJNA288601 |
| GCA 021906855 | PDT001131150.2 | Klebsiella_147 | PDS000091501.176 | 2021 | USA | PRJNA288601 |
| GCA 021938555 | PDT001071586.3 | Klebsiella_147 | PDS000091501.176 | 2021 | USA | PRJNA288601 |
| GCA 026252075 | PDT001488210.3 | Klebsiella_147 | PDS000091501.176 | 2022 | USA | PRJNA288601 |
| GCA 022011215 | PDT001105005.3 | Klebsiella_147 | PDS000091501.176 | 2021 | USA | PRJNA288601 |
| GCA 021906495 | PDT001132274.3 | Klebsiella_147 | PDS000091501.176 | 2021 | USA | PRJNA288601 |
| GCA 021903155 | PDT001157542.7 | Klebsiella_147 | PDS000091501.176 | 2021 | USA | PRJNA288601 |
| GCA 021906635 | PDT001131219.2 | Klebsiella_147 | PDS000091501.176 | 2021 | USA | PRJNA288601 |
| GCA 022039595 | PDT001105012.3 | Klebsiella_147 | PDS000091501.176 | 2021 | USA | PRJNA288601 |
| GCA 021906715 | PDT001131158.2 | Klebsiella_147 | PDS000091501.176 | 2021 | USA | PRJNA288601 |
| GCA 021871545 | PDT001199340.3 | Klebsiella_147 | PDS000091501.176 | 2021 | USA | PRJNA288601 |
| GCA 021871475 | PDT001199344.3 | Klebsiella_147 | PDS000091501.176 | 2021 | USA | PRJNA288601 |
| GCA 023557875 | PDT001315283.3 | Klebsiella_147 | PDS000091501.176 | 2021 | USA | PRJNA288601 |
| GCA 022475195 | PDT001257970.3 | Klebsiella_147 | PDS000091501.176 | 2022 | USA | PRJNA288601 |
| GCA 023804755 | PDT001328521.3 | Klebsiella_147 | PDS000091501.176 | 2022 | USA | PRJNA288601 |
| GCA 022915795 | PDT001278723.3 | Klebsiella_147 | PDS000091501.176 | 2022 | USA | PRJNA288601 |
| GCA 024263845 | PDT001358505.3 | Klebsiella_147 | PDS000091501.176 | 2022 | USA | PRJNA288601 |
| GCA 021902975 | PDT001157539.8 | Klebsiella_147 | PDS000091501.176 | 2021 | USA | PRJNA288601 |
| GCA 021880275 | PDT001195986.3 | Klebsiella_147 | PDS000091501.176 | 2021 | USA | PRJNA288601 |
| GCA 023553575 | PDT001315272.3 | Klebsiella_147 | PDS000091501.176 | 2022 | USA | PRJNA288601 |
| GCA 026059875 | PDT001480815.3 | Klebsiella_147 | PDS000091501.176 | 2022 | USA | PRJNA288601 |
| GCA 024798985 | PDT001404023.3 | Klebsiella_147 | PDS000091501.176 | 2022 | USA | PRJNA288601 |
| GCA 025224295 | PDT001417266.3 | Klebsiella_147 | PDS000091501.176 | 2022 | USA | PRJNA288601 |
| GCA 021871575 | PDT001199339.3 | Klebsiella_147 | PDS000091501.176 | 2021 | USA | PRJNA288601 |
| GCA 021898515 | PDT001170229.3 | Klebsiella_147 | PDS000091501.176 | 2021 | USA | PRJNA288601 |
| GCA 028450985 | PDT001601423.3 | Klebsiella_147 | PDS000091501.176 | 2022 | USA | PRJNA288601 |
| GCA 028451065 | PDT001601421.3 | Klebsiella_147 | PDS000091501.176 | 2022 | USA | PRJNA288601 |
| GCA 031212405 | PDT001876199.2 | Klebsiella_147 | PDS000091501.176 | 2023 | USA | PRJNA288601 |
| GCA 036476075 | PDT002067386.1 | Klebsiella_147 | PDS000091501.176 | 2023 | USA | PRJNA288601 |
| GCA 031211425 | PDT001876245.2 | Klebsiella_147 | PDS000091501.176 | 2023 | USA | PRJNA288601 |
| GCA 025858675 | PDT001465724.3 | Klebsiella_147 | PDS000091501.176 | 2022 | USA | PRJNA288601 |
| GCA 031214145 | PDT001876143.2 | Klebsiella_147 | PDS000091501.176 | 2023 | USA | PRJNA288601 |
| GCA 028451045 | PDT001601416.3 | Klebsiella_147 | PDS000091501.176 | 2022 | USA | PRJNA288601 |
| GCA 024905655 | PDT001395485.3 | Klebsiella_147 | PDS000091501.176 | 2022 | USA | PRJNA288601 |
| GCA 031213755 | PDT001876157.2 | Klebsiella_147 | PDS000091501.176 | 2023 | USA | PRJNA288601 |
| GCA 045662295 | PDT002506028.1 | Klebsiella_147 | PDS000091501.176 | 2024 | USA | PRJNA288601 |
| GCA 025858735 | PDT001465723.3 | Klebsiella_147 | PDS000091501.176 | 2022 | USA | PRJNA288601 |
| GCA 027913835 | PDT001551200.3 | Klebsiella_147 | PDS000091501.176 | 2022 | USA | PRJNA288601 |
| GCA 031214025 | PDT001876147.2 | Klebsiella_147 | PDS000091501.176 | 2022 | USA | PRJNA288601 |
| GCA 026058415 | PDT001480810.3 | Klebsiella_147 | PDS000091501.176 | 2022 | USA | PRJNA288601 |
| GCA 025224175 | PDT001417274.3 | Klebsiella_147 | PDS000091501.176 | 2022 | USA | PRJNA288601 |
| GCA 021938095 | PDT001072678.3 | Klebsiella_147 | PDS000091501.176 | 2021 | USA | PRJNA288601 |
| GCA 021938535 | PDT001071573.3 | Klebsiella_147 | PDS000091501.176 | 2020 | USA | PRJNA288601 |
| GCA 021816185 | PDT001222666.3 | Klebsiella_147 | PDS000091501.176 | 2021 | USA | PRJNA288601 |
| GCA 021938155 | PDT001072675.3 | Klebsiella_147 | PDS000091501.176 | 2021 | USA | PRJNA288601 |
| GCA 023553695 | PDT001315263.3 | Klebsiella_147 | PDS000091501.176 | 2022 | USA | PRJNA288601 |
| GCA 021906775 | PDT001131156.2 | Klebsiella_147 | PDS000091501.176 | 2021 | USA | PRJNA288601 |
| GCA 025224095 | PDT001417278.3 | Klebsiella_147 | PDS000091501.176 | 2022 | USA | PRJNA288601 |
| GCA 021938575 | PDT001071571.3 | Klebsiella_147 | PDS000091501.176 | 2021 | USA | PRJNA288601 |
| GCA 021938395 | PDT001072668.3 | Klebsiella_147 | PDS000091501.176 | 2021 | USA | PRJNA288601 |
| GCA 021938195 | PDT001071585.3 | Klebsiella_147 | PDS000091501.176 | 2021 | USA | PRJNA288601 |
| GCA 021871355 | PDT001199348.3 | Klebsiella_147 | PDS000091501.176 | 2021 | USA | PRJNA288601 |
| GCA 021907595 | PDT001105019.3 | Klebsiella_147 | PDS000091501.176 | 2021 | USA | PRJNA288601 |
| GCA 021938335 | PDT001072673.3 | Klebsiella_147 | PDS000091501.176 | 2021 | USA | PRJNA288601 |
| GCA 021938315 | PDT001072671.3 | Klebsiella_147 | PDS000091501.176 | 2021 | USA | PRJNA288601 |
| GCA 024012615 | PDT001338065.4 | Klebsiella_147 | PDS000091501.176 | 2022 | USA | PRJNA288601 |
| GCA 023564565 | PDT001315788.3 | Klebsiella_147 | PDS000091501.176 | 2022 | USA | PRJNA288601 |
| GCA 023724465 | PDT001328515.3 | Klebsiella_147 | PDS000091501.176 | 2022 | USA | PRJNA288601 |
| GCA 023554175 | PDT001315257.3 | Klebsiella_147 | PDS000091501.176 | 2022 | USA | PRJNA288601 |
| GCA 021907555 | PDT001105015.3 | Klebsiella_147 | PDS000091501.176 | 2021 | USA | PRJNA288601 |
| GCA 021938375 | PDT001072670.3 | Klebsiella_147 | PDS000091501.176 | 2021 | USA | PRJNA288601 |
| GCA 021907615 | PDT001105020.3 | Klebsiella_147 | PDS000091501.176 | 2021 | USA | PRJNA288601 |
| GCA 021906695 | PDT001131159.2 | Klebsiella_147 | PDS000091501.176 | 2021 | USA | PRJNA288601 |
| GCA 023553835 | PDT001315284.3 | Klebsiella_147 | PDS000091501.176 | 2021 | USA | PRJNA288601 |
| GCA 023563345 | PDT001315705.3 | Klebsiella_147 | PDS000091501.176 | 2022 | USA | PRJNA288601 |
| GCA 021880215 | PDT001195987.3 | Klebsiella_147 | PDS000091501.176 | 2021 | USA | PRJNA288601 |
| GCA 023872945 | PDT001328518.3 | Klebsiella_147 | PDS000091501.176 | 2022 | USA | PRJNA288601 |
| GCA 026058335 | PDT001480827.3 | Klebsiella_147 | PDS000091501.176 | 2022 | USA | PRJNA288601 |
| GCA 031214825 | PDT001876112.2 | Klebsiella_147 | PDS000091501.176 | 2023 | USA | PRJNA288601 |
| GCA 045482725 | PDT002500111.1 | Klebsiella_147 | PDS000091501.176 | 2024 | USA | PRJNA288601 |
| GCA 022011275 | PDT001105002.3 | Klebsiella_147 | PDS000091501.176 | 2021 | USA | PRJNA288601 |
| GCA 023107555 | PDT001293731.3 | Klebsiella_147 | PDS000091501.176 | 2021 | USA | PRJNA288601 |
| GCA 025379025 | PDT001428679.3 | Klebsiella_147 | PDS000091501.176 | 2022 | USA | PRJNA288601 |
| GCA 027830275 | PDT001550926.3 | Klebsiella_147 | PDS000091501.176 | 2022 | USA | PRJNA288601 |
| GCA 036469195 | PDT002067732.1 | Klebsiella_147 | PDS000091501.176 | 2023 | USA | PRJNA288601 |
| GCA 036473815 | PDT002067481.1 | Klebsiella_147 | PDS000091501.176 | 2023 | USA | PRJNA288601 |
| GCA 045433955 | PDT002498179.1 | Klebsiella_147 | PDS000091501.176 | 2024 | USA | PRJNA288601 |
| GCA 021871595 | PDT001199337.3 | Klebsiella_147 | PDS000091501.176 | 2021 | USA | PRJNA288601 |
| GCA 021880335 | PDT001195979.3 | Klebsiella_147 | PDS000091501.176 | 2021 | USA | PRJNA288601 |
| GCA 025378765 | PDT001428691.3 | Klebsiella_147 | PDS000091501.176 | 2022 | USA | PRJNA288601 |
| GCA 023726885 | PDT001328519.3 | Klebsiella_147 | PDS000091501.176 | 2022 | USA | PRJNA288601 |
| GCA 023564765 | PDT001315777.3 | Klebsiella_147 | PDS000091501.176 | 2022 | USA | PRJNA288601 |
| GCA 024263685 | PDT001358512.3 | Klebsiella_147 | PDS000091501.176 | 2022 | USA | PRJNA288601 |
| GCA 026719945 | PDT001515157.3 | Klebsiella_147 | PDS000091501.176 | 2022 | USA | PRJNA288601 |
| GCA 036469015 | PDT002067746.1 | Klebsiella_147 | PDS000091501.176 | 2023 | USA | PRJNA288601 |
| GCA 045424695 | PDT002498201.1 | Klebsiella_147 | PDS000091501.176 | 2024 | USA | PRJNA288601 |
| GCA 021903015 | PDT001157538.8 | Klebsiella_147 | PDS000091501.176 | 2021 | USA | PRJNA288601 |
| GCA 031209245 | PDT001876299.2 | Klebsiella_- | PDS000091501.176 | 2023 | USA | PRJNA288601 |
| GCA 021903215 | PDT001157540.7 | Klebsiella_147 | PDS000091501.176 | 2021 | USA | PRJNA288601 |
| GCA 021906755 | PDT001131155.2 | Klebsiella_- | PDS000091501.176 | 2021 | USA | PRJNA288601 |
| GCA 023553875 | PDT001315265.3 | Klebsiella_- | PDS000091501.176 | 2022 | USA | PRJNA288601 |
| GCA 021902955 | PDT001157541.7 | Klebsiella_- | PDS000091501.176 | 2021 | USA | PRJNA288601 |
| GCA 023557915 | PDT001315280.3 | Klebsiella_- | PDS000091501.176 | 2022 | USA | PRJNA288601 |
| GCA 021871605 | PDT001199338.3 | Klebsiella_- | PDS000091501.176 | 2021 | USA | PRJNA288601 |

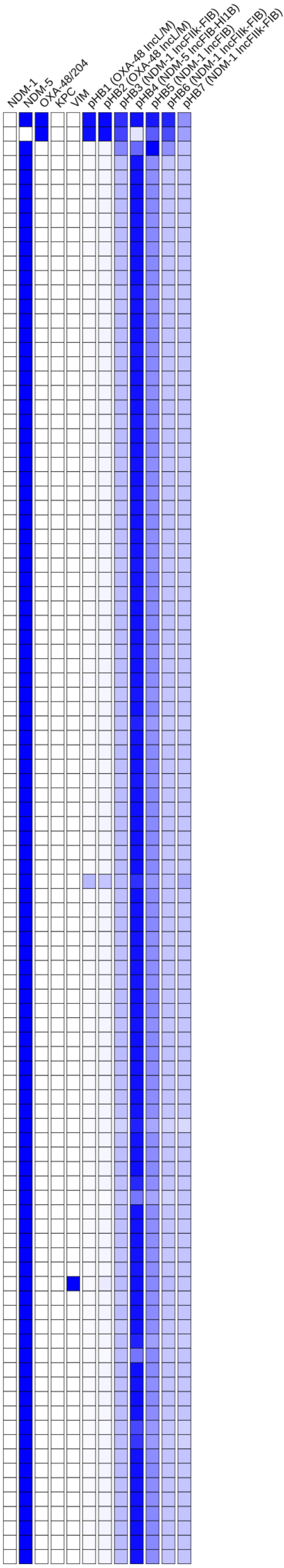

### Tree 40: GCA_043905645

Tree scale: 0.1

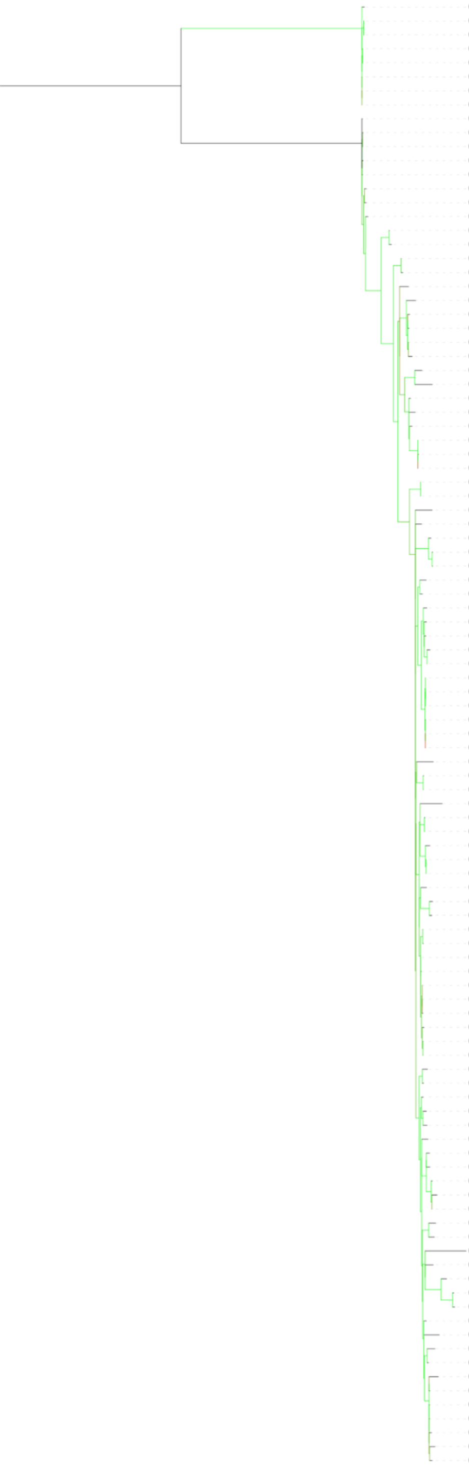

| | Isolate | MLST | SNP Cluster | Collection Year | Country | BioProject |
|---|---|---|---|---|---|---|
| GCA 022038275 | PDT000893789.3 | Klebsiella_4843 | PDS000052089.184 | 2019 | USA | PRJNA288601 |
| GCA 022034195 | PDT000899437.1 | Klebsiella_4843 | PDS000052089.184 | 2019 | USA | PRJNA288601 |
| GCA 041756215 | PDT002363661.1 | Klebsiella_4843 | PDS000052089.184 | 2019 | USA | PRJNA288601 |
| GCA 041756655 | PDT002363660.1 | Klebsiella_4843 | PDS000052089.184 | 2019 | USA | PRJNA288601 |
| GCA 022039435 | PDT000893652.1 | Klebsiella_4843 | PDS000052089.184 | 2019 | USA | PRJNA288601 |
| GCA 043301965 | PDT002265360.1 | Klebsiella_4843 | PDS000052089.184 | 2018 | USA | PRJNA1106484 |
| GCA 041767165 | PDT002364337.1 | Klebsiella_4843 | PDS000052089.184 | 2018 | USA | PRJNA288601 |
| GCA 022039255 | PDT000893665.3 | Klebsiella_4843 | PDS000052089.184 | 2019 | USA | PRJNA288601 |
| GCA 018068425 | PDT001009219.1 | Klebsiella_147 | PDS000052089.184 | 2019 | USA | PRJNA288601 |
| GCA 018068525 | PDT001009222.1 | Klebsiella_147 | PDS000052089.184 | 2019 | USA | PRJNA288601 |
| GCA 043448245 | PDT002421413.1 | Klebsiella_147 | PDS000052089.184 | 2019 | USA | PRJNA288601 |
| GCA 043458055 | PDT002421633.1 | Klebsiella_147 | PDS000052089.184 | 2019 | USA | PRJNA288601 |
| GCA 043442185 | PDT002421417.1 | Klebsiella_147 | PDS000052089.184 | 2019 | USA | PRJNA288601 |
| GCA 018068485 | PDT001009221.1 | Klebsiella_147 | PDS000052089.184 | 2019 | USA | PRJNA288601 |
| GCA 018068465 | PDT001009220.1 | Klebsiella_147 | PDS000052089.184 | 2019 | USA | PRJNA288601 |
| GCA 031036015 | PDT001726355.3 | Klebsiella_147 | PDS000052089.184 | 2018 | USA | PRJNA288601 |
| GCA 021943635 | PDT001042195.1 | Klebsiella_147 | PDS000052089.184 | 2016 | USA | PRJNA725484 |
| GCA 021943655 | PDT001042194.1 | Klebsiella_147 | PDS000052089.184 | 2016 | Germany | PRJNA725484 |
| GCA 021801655 | PDT001227339.1 | Klebsiella_147 | PDS000156049.11 | 2014 | United Kingdom | PRJNA788733 |
| GCA 021798605 | PDT001227461.1 | Klebsiella_147 | PDS000156049.11 | 2014 | United Kingdom | PRJNA788733 |
| GCA 022114895 | PDT001245144.1 | Klebsiella_147 | PDS000156049.11 | 2014 | United Kingdom | PRJNA788733 |
| GCA 947576575 | PDT001769496.1 | Klebsiella_147 | PDS000156049.11 | 2022 | Germany | PRJEB58018 |
| GCA 022169745 | PDT000847257.1 | Klebsiella_147 | PDS000156049.11 | 2018 | Myanmar | PRJDB5126 |
| GCA 022283325 | PDT000431873.1 | Klebsiella_147 | PDS000156049.11 | 2016 | Myanmar | PRJDB5126 |
| GCA 022276245 | PDT000432066.1 | Klebsiella_147 | PDS000156049.11 | 2015 | Myanmar | PRJDB5126 |
| GCA 022276265 | PDT000432048.1 | Klebsiella_147 | PDS000156049.11 | 2016 | Myanmar | PRJDB5126 |
| GCA 045027735 | PDT002474493.1 | Klebsiella_147 | PDS000156049.11 | 2017 | India | PRJNA939406 |
| GCA 024998405 | PDT001412313.1 | Klebsiella_147 | PDS000156049.11 | 2020 | India | PRJNA738274 |
| GCA 022282545 | PDT000431951.1 | Klebsiella_147 | PDS000156049.11 | 2016 | Thailand | PRJDB5929 |
| GCA 004832395 | PDT000495993.1 | Klebsiella_147 | PDS000156049.11 | 2018 | Thailand | PRJNA532291 |
| GCA 021980415 | PDT001008159.1 | Klebsiella_147 | PDS000156049.11 | 2016 | Thailand | PRJDB5929 |
| GCA 021980595 | PDT001008111.1 | Klebsiella_147 | PDS000156049.11 | 2017 | Thailand | PRJDB5929 |
| GCA 021984435 | PDT001003302.1 | Klebsiella_147 | PDS000156049.11 | 2017 | Thailand | PRJDB5929 |
| GCA 021982115 | PDT001008059.1 | Klebsiella_147 | PDS000156049.11 | 2016 | Thailand | PRJDB5929 |
| GCA 022283345 | PDT000431869.1 | Klebsiella_147 | PDS000156049.11 | 2016 | Myanmar | PRJDB5126 |
| GCA 022208685 | PDT000650074.1 | Klebsiella_147 | PDS000156049.11 | 2016 | Myanmar | PRJDB8975 |
| GCA 023925325 | PDT001345521.1 | Klebsiella_147 | PDS000106697.6 | 2019 | Israel | PRJEB53419 |
| GCA 021875215 | PDT001197766.1 | Klebsiella_147 | PDS000156049.11 | 2013 | Singapore | PRJNA577535 |
| GCA 021982215 | PDT001008055.1 | Klebsiella_147 | PDS000056180.7 | 2016 | Thailand | PRJDB5929 |
| GCA 021983255 | PDT001003383.1 | Klebsiella_147 | PDS000056180.7 | 2016 | Thailand | PRJDB5929 |
| GCA 021981075 | PDT001008160.1 | Klebsiella_147 | PDS000056180.7 | 2016 | Thailand | PRJDB5929 |
| GCA 004803225 | PDT000495886.1 | Klebsiella_147 | PDS000077015.34 | nan | Egypt | PRJNA508509 |
| GCA 004803235 | PDT000495887.1 | Klebsiella_147 | PDS000077015.34 | nan | Egypt | PRJNA508509 |
| GCA 022352825 | PDT001118267.2 | Klebsiella_147 | PDS000077015.34 | 2019 | Italy | PRJNA643814 |
| GCA 022353485 | PDT001118270.2 | Klebsiella_147 | PDS000077015.34 | 2019 | Italy | PRJNA643814 |
| GCA 021942915 | PDT001042232.1 | Klebsiella_147 | PDS000077015.34 | 2019 | Italy | PRJNA725484 |
| GCA 021943695 | PDT001042193.1 | Klebsiella_147 | PDS000077015.34 | 2019 | Italy | PRJNA725484 |
| GCA 023466435 | PDT000906262.2 | Klebsiella_147 | PDS000077015.34 | 2019 | Italy | PRJNA643814 |
| GCA 021942215 | PDT001042253.1 | Klebsiella_147 | PDS000077015.34 | 2019 | Italy | PRJNA725484 |
| GCA 021942155 | PDT001042257.1 | Klebsiella_147 | PDS000077015.34 | 2019 | Italy | PRJNA725484 |
| GCA 021942775 | PDT001042238.1 | Klebsiella_147 | PDS000077015.34 | 2019 | Italy | PRJNA725484 |
| GCA 021943555 | PDT001042198.1 | Klebsiella_147 | PDS000077015.34 | 2020 | Italy | PRJNA725484 |
| GCA 021944435 | PDT001042156.1 | Klebsiella_147 | PDS000077015.34 | 2019 | Italy | PRJNA725484 |
| GCA 021943755 | PDT001042189.1 | Klebsiella_147 | PDS000077015.34 | 2019 | Italy | PRJNA725484 |
| GCA 021983855 | PDT001003356.1 | Klebsiella_147 | PDS000104520.1 | 2016 | Thailand | PRJDB5929 |
| GCA 002202215 | PDT000219969.1 | Klebsiella_147 | PDS000156049.11 | nan | NA | PRJNA292902 |
| GCA 002202235 | PDT000219970.1 | Klebsiella_147 | PDS000156049.11 | nan | NA | PRJNA292902 |
| GCA 024273875 | PDT001345527.1 | Klebsiella_147 | PDS000156049.11 | 2018 | Israel | PRJEB53419 |
| GCA 021942955 | PDT001042230.1 | Klebsiella_147 | PDS000156049.11 | nan | USA | PRJNA725484 |
| GCA 037050025 | PDT002101976.1 | Klebsiella_147 | PDS000156049.11 | 2016 | Jordan | PRJNA1076682 |
| GCA 021942435 | PDT001042255.1 | Klebsiella_147 | PDS000156049.11 | nan | USA | PRJNA725484 |
| GCA 037052305 | PDT002102090.1 | Klebsiella_147 | PDS000156049.11 | 2013 | Jordan | PRJNA1076682 |
| GCA 021944875 | PDT001042137.1 | Klebsiella_147 | PDS000156049.11 | nan | USA | PRJNA725484 |
| GCA 022154425 | PDT000858721.1 | Klebsiella_147 | PDS000156049.11 | 2017 | USA | PRJNA658369 |
| GCA 021969795 | PDT001019592.1 | Klebsiella_147 | PDS000156049.11 | 2017 | USA | PRJNA658369 |
| GCA 021969955 | PDT001019579.1 | Klebsiella_147 | PDS000156049.11 | 2018 | USA | PRJNA658369 |
| GCA 037051225 | PDT002102036.1 | Klebsiella_147 | PDS000156049.11 | 2017 | Jordan | PRJNA1076682 |
| GCA 021943095 | PDT001042208.1 | Klebsiella_147 | PDS000156049.11 | nan | USA | PRJNA725484 |
| GCA 021943715 | PDT001042191.1 | Klebsiella_147 | PDS000156049.11 | nan | USA | PRJNA725484 |
| GCA 021944495 | PDT001042155.1 | Klebsiella_147 | PDS000156049.11 | nan | USA | PRJNA725484 |
| GCA 021943235 | PDT001042219.1 | Klebsiella_147 | PDS000156049.11 | nan | USA | PRJNA725484 |
| GCA 037051425 | PDT002102046.1 | Klebsiella_147 | PDS000156049.11 | 2015 | Jordan | PRJNA1076682 |
| GCA 021943995 | PDT001042177.1 | Klebsiella_147 | PDS000156049.11 | nan | USA | PRJNA725484 |
| GCA 021943595 | PDT001042197.1 | Klebsiella_147 | PDS000156049.11 | nan | USA | PRJNA725484 |
| GCA 021944135 | PDT001042166.1 | Klebsiella_147 | PDS000156049.11 | nan | USA | PRJNA725484 |
| GCA 037051365 | PDT002102043.1 | Klebsiella_147 | PDS000156049.11 | 2015 | Jordan | PRJNA1076682 |
| GCA 021775025 | PDT001227850.1 | Klebsiella_147 | PDS000156049.11 | 2016 | United Kingdom | PRJNA788733 |
| GCA 900502585 | PDT000370025.1 | Klebsiella_147 | PDS000156049.11 | 2014 | Denmark | PRJEB10018 |
| GCA 022160785 | PDT001243475.1 | Klebsiella_147 | PDS000156049.11 | 2014 | United Kingdom | PRJNA788733 |
| GCA 043829465 | PDT002430473.1 | Klebsiella_147 | PDS000103185.4 | 2020 | Germany | PRJNA1070536 |
| GCA 044381765 | PDT002456479.1 | Klebsiella_147 | PDS000201444.2 | nan | Saudi Arabia | PRJNA1116866 |
| GCA 016652065 | PDT000937820.1 | Klebsiella_147 | | nan | Egypt | PRJNA689413 |
| GCA 043905165 | **HBH_556_19** | Klebsiella_147 | PDS000090167.14 | 2019 | Tunisia | PRJNA1160997 |
| GCA 021837925 | PDT001216482.1 | Klebsiella_147 | PDS000090167.14 | 2019 | Switzerland | PRJNA744003 |
| GCA 035017145 | PDT002045097.1 | Klebsiella_147 | PDS000090167.14 | 2022 | Libya | PRJNA998204 |
| GCA 035017185 | PDT002045099.1 | Klebsiella_147 | PDS000090167.14 | 2022 | Libya | PRJNA998204 |
| GCA 035016945 | PDT002045088.1 | Klebsiella_147 | PDS000090167.14 | 2022 | Libya | PRJNA998204 |
| GCA 947574945 | PDT001769393.1 | Klebsiella_147 | PDS000199427.1 | 2022 | Germany | PRJEB58018 |
| GCA 043905225 | **HBH_3009_20** | Klebsiella_147 | PDS000199427.1 | 2020 | Tunisia | PRJNA1160997 |
| GCA 044343035 | PDT002455478.1 | Klebsiella_147 | PDS000205531.1 | 2022 | Senegal | PRJNA1073865 |
| GCA 043905645 | **HBH_3065_22** | Klebsiella_147 | | 2022 | Tunisia | PRJNA1160997 |
| GCA 043906065 | PDT002391396.1 | Klebsiella_147 | PDS000136743.8 | 2021 | Netherlands | PRJNA1143178 |
| GCA 043906525 | PDT002391374.1 | Klebsiella_147 | PDS000136743.8 | 2023 | Netherlands | PRJNA1143178 |
| GCA 043976415 | PDT002391348.1 | Klebsiella_147 | PDS000136743.8 | 2023 | Netherlands | PRJNA1143178 |
| GCA 043905445 | **HBH_1520_21** | Klebsiella_147 | PDS000090167.14 | 2021 | Tunisia | PRJNA1160997 |
| GCA 947577135 | PDT001769510.1 | Klebsiella_147 | PDS000129332.6 | 2022 | Germany | PRJEB58018 |
| GCA 032720655 | PDT001927061.1 | Klebsiella_147 | PDS000090167.14 | 2020 | Italy | PRJNA1026096 |
| GCA 043905245 | **HBH_110_20** | Klebsiella_147 | PDS000090167.14 | 2020 | Tunisia | PRJNA1160997 |
| GCA 018421315 | PDT001044123.1 | Klebsiella_147 | PDS000090167.14 | 2019 | Germany | PRJNA692829 |
| GCA 036455425 | PDT002074710.1 | Klebsiella_147 | PDS000090167.14 | 2020 | Germany | PRJNA1060526 |
| GCA 032721535 | PDT001927023.1 | Klebsiella_147 | PDS000090167.14 | 2019 | Italy | PRJNA1026096 |
| GCA 025889115 | PDT001469197.1 | Klebsiella_147 | PDS000090167.14 | 2019 | Italy | PRJNA787449 |
| GCA 018440925 | PDT001046592.1 | Klebsiella_147 | PDS000090167.14 | 2019 | Germany | PRJNA692829 |
| GCA 018422375 | PDT001044139.1 | Klebsiella_147 | PDS000090167.14 | 2019 | Germany | PRJNA692829 |
| GCA 043830075 | PDT002430479.1 | Klebsiella_147 | PDS000090167.14 | 2019 | Germany | PRJNA1070536 |

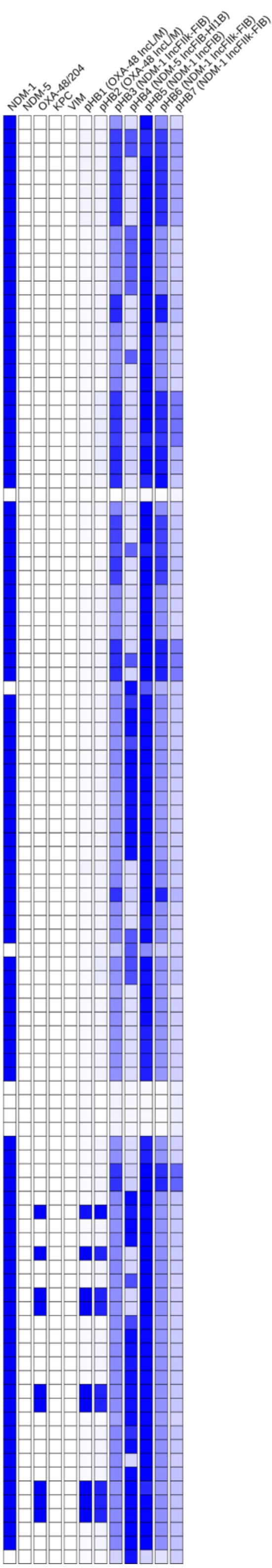

### Tree 41: GCA_043905665

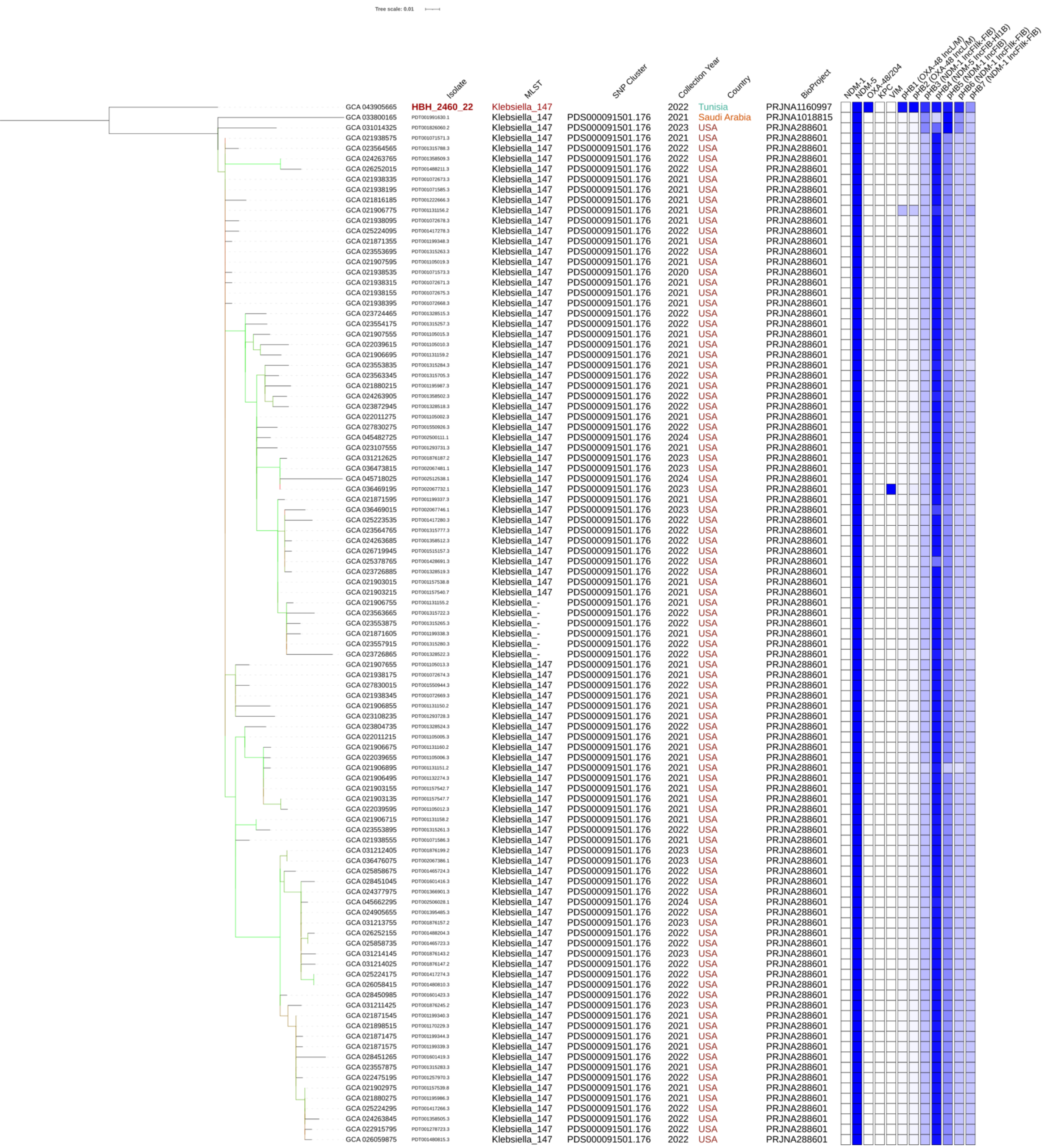

### Tree 42: MLST383_top400.raxml.support

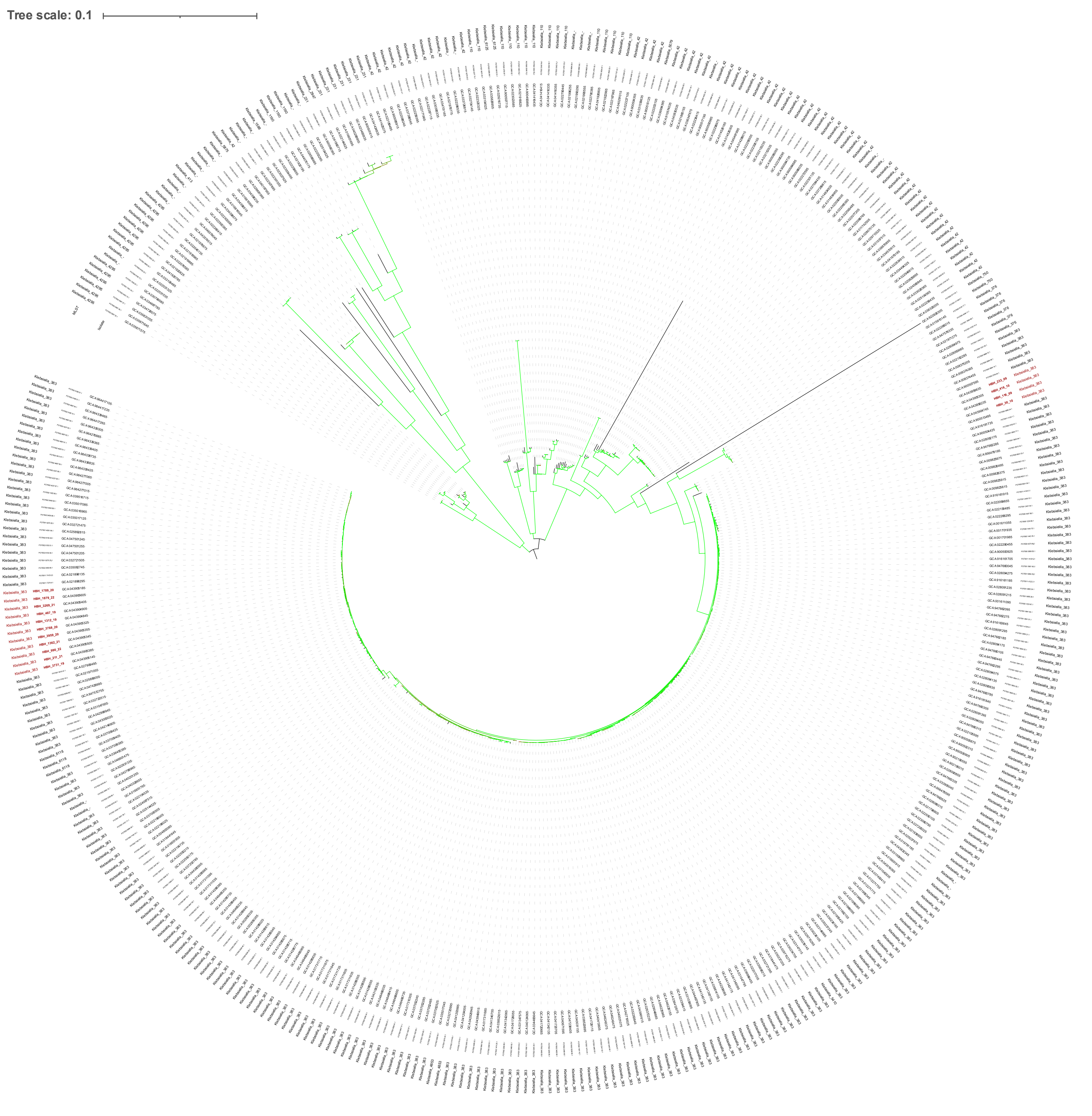

### Tree 43: ST383_ST376

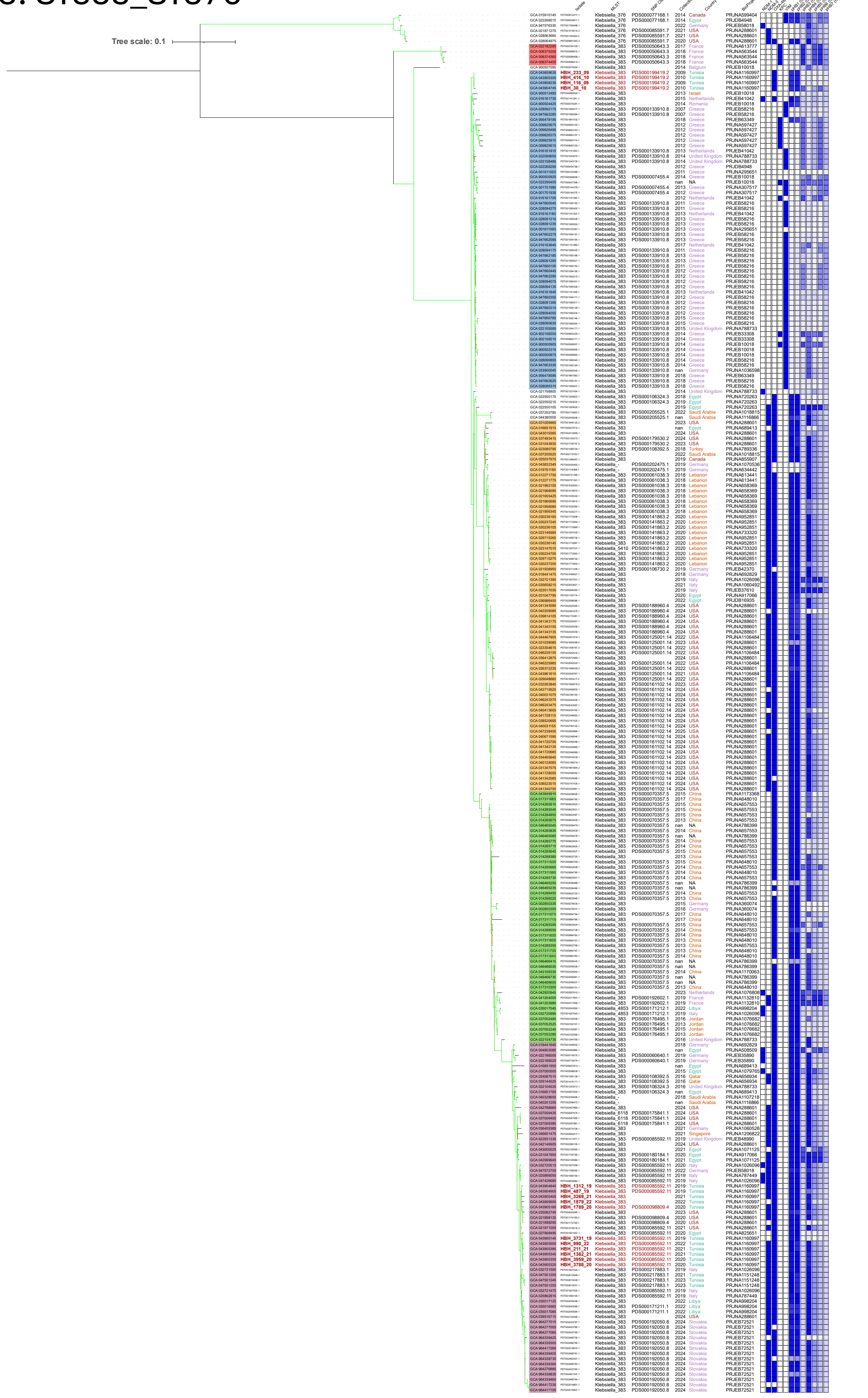
